# Comparative Cardiovascular Effectiveness of Glucagon-Like Peptide 1 Receptor Agonists and Sodium-Glucose Cotransporter-2 Inhibitors in Diabetes Mellitus

**DOI:** 10.64898/2026.02.23.26346890

**Authors:** Fan Bu, Ruopeng Wu, Anna Ostropolets, Arya Aminorroaya, Hsin Yi Chen, Yi Chai, Lovedeep S Dhingra, Thomas Falconer, Jason C. Hsu, Chungsoo Kim, Wallis CY Lau, Kenneth KC Man, Evan Minty, Daniel R Morales, Akihiko Nishimura, Phyllis Thangraraj, Mui Van Zandt, Can Yin, Rohan Khera, George Hripcsak, Marc A Suchard

## Abstract

**Background:** GLP-1 receptor agonists (GLP-1RAs) and SGLT2 inhibitors (SGLT2Is) have established cardiovascular benefits for patients with type 2 diabetes mellitus (T2DM), with similar class-level effectiveness found in previous studies. However, real-world comparative effectiveness assessments of individual agents remain limited.

**Objectives:** To compare the cardiovascular effectiveness of individual GLP-1RAs and SGLT2Is.

**Methods:** We conducted a multi-national, retrospective, new-user active-comparator cohort study using 10 US and non-US administrative claims and electronic health record databases. The study included 1,245,211 adults with T2DM receiving metformin who initiated second-line therapy with one of six GLP-1RAs (albiglutide, dulaglutide, exenatide, liraglutide, lixisenatide, semaglutide) or one of four SGLT2Is (canagliflozin, dapagliflozin, empagliflozin, ertugliflozin). Empagliflozin (393,499; 31.6%), semaglutide (235,585; 18.9%), dapagliflozin (208,666; 16.8%), and dulaglutide (207,348; 16.8%) were most commonly used. A secondary subgroup analysis included 316,242 patients with established cardiovascular diseases (CVD).

Primary outcomes were 3-point major adverse cardiovascular events (MACE: acute myocardial infarction, stroke, sudden cardiac death) and 4-point MACE (adding hospitalization/ER visit with heart failure). Secondary outcomes included the individual components. Hazard ratios (HRs) were estimated for pairwise agent comparisons while on-treatment (per-protocol) and over total follow-up using Cox proportional hazards models, with propensity score adjustments, negative control calibration, and pre-specified study diagnostics to guard against potential confounding. Random-effects meta-analysis produced summary HR estimates across data sources that passed diagnostics.

**Results:** Across the study cohort, individual GLP-1RAs and SGLT2Is demonstrated broadly similar cardiovascular effectiveness, both within and across drug classes. For example, semaglutide and empagliflozin showed comparable risks for 3-point MACE (meta-analytic HR 1.05; 95% CI 0.79–1.39) and 4-point MACE (meta-analytic HR 0.95; 95% CI 0.81–1.12), with consistent findings in the CVD subgroup. Study diagnostics confirmed adequate equipoise, covariate balance and statistical power to detect similarity in HRs between 0.8 and 1.2 for commonly used agents.

**Conclusions:** In this large-scale real-world study, individual GLP-1RAs and SGLT2Is exhibited largely comparable cardiovascular benefits, including in patients with established CVD. These findings align with network meta-analytic estimates from major cardiovascular outcome trials and broadly support current treatment guidelines. Clinical choices should be guided by relevant factors such as safety, adherence, tolerability, cost, and patient preference, where further work is needed.

## 1. Introduction

Type 2 diabetes mellitus (T2DM) is a major global health issue affecting more than 500 million people worldwide [1], with patients facing a higher risk of cardiovascular diseases compared to those without T2DM [2,3]. In recent years, treatment options for T2DM have expanded considerably with the development of newer second-line medications. Among these, two drug classes — glucagon-like peptide 1 receptor agonists (GLP-1RAs) and sodium-glucose cotransporter-2 inhibitors (SGLT2Is) — have shown cardioprotective benefits beyond glycemic control through placebo-controlled randomized clinical trials (RCTs) [4–10]. These benefits exceed those of other second-line medications such as dipeptidyl peptidase-4 inhibitors (DPP4I) or sulphonylureas, as demonstrated in active-comparator trials and meta-analyses of RCTs [11–18] and real-world observational studies [19–26].

The cardiovascular benefits of GLP-1RAs and SGLT2Is against major adverse cardiovascular events (MACE) are well-established [8–10,27–31]. However, limited evidence exists on the comparative effectiveness of individual agents within these drug classes. Current clinical guidelines recommend these drugs to T2DM patients with higher cardiovascular risks but do not provide specific guidance on the choice between individual GLP-1RAs and SGLT2Is [32]. RCTs providing head-to-head comparisons at the individual agent level remain scarce, and yet clinical decisions at the agent level are being made every day. Real-world evidence is therefore of great practical relevance, but most real-world studies have compared cardiovascular effectiveness at the drug class level [20,23–25,33–35]. Prior drug-level studies largely relied on a single data source [36] or focused on a specific agent [23], which limits the generalizability of the results. Furthermore, previous observational studies have conflicting results: GLP1-RAs were reported to provide more protection against ischemic stroke [25], and SGLT2Is were shown to be superior for heart failure hospitalizations [34], and yet a multi-national large-scale comparison identified no significant differences between these two classes across all MACE outcomes [24]. Given the limited and inconsistent evidence, cross- and within-class comparative effectiveness of individual GLP-1RAs and SGLT2Is remains unclear. However, such evidence can directly inform care to prescribe the more effective agent in clinical practice. Thus, a more comprehensive comparison across multiple cardiovascular outcomes and multiple data sources is needed.

To address these evidence gaps, we conducted a large-scale comparative effectiveness study of individual GLP-1RA and SGLT2I agents as part of Large-Scale Evidence Generation and Evaluation across a Network of Databases for Type 2 Diabetes Mellitus (LEGEND-T2DM) initiative. LEGEND-T2DM leverages a multinational observational data network covering more than 500 million patients [37], using administrative claims and electronic health records (EHRs) through the Observational Health Data Sciences and Informatics (OHDSI) global collaborative. [38]

In this study, we performed head-to-head pairwise comparisons across six GLP-1RAs and four SGLT2Is to evaluate the relative risk of six cardiovascular outcomes, in both general T2DM populations and subpopulations with established cardiovascular diseases (CVD). To ensure reliability and reproducibility, we used harmonized data sources and a standardized framework for cohort definition, data processing, statistical analysis and results interpretation, as pre-specified in our study protocol [39] We further made all analytic scripts and results publicly available to promote open science.

## 2. Materials and method

### 2.1 Data sources

Our study included five administrative claims and five EHR databases with coding systems mapped to OHDSI’s Observational Medical Outcomes Partnership (OMOP) common data model (CDM) [40]The US-based claims databases included Merative MarketScan Commercial Claims and Encounters (CCAE), Optum ClinFormatics Data Mart (Clinformatics), Merative MarketScan Medicare Supplemental Beneficiaries (MDCR), Merative MarketScan Multi-state Medicaid (MDCD) and IQVIA Open Claims (Open Claims). The US-based EHRs spanned Optum Electronic Health Records (OptumEHR), Columbia University Irving Medical Center database (CUIMC), and US Department of Veterans Affairs (VA); the VA data source includes EHRs from all 163 VA healthcare centers and over 900 community-based clinics, with date of death records Non-US EHRs were Germany Disease Analyzer (GermanyDA) and UK general practice non-identified medical records data (IMRD). See Supplementary Table 1 for description of these databases.

All data partners received institutional approval or exemption for their participation. For CCAE, MDCR, MDCD, Optum Cliniformatics, and OptumEHR, New England IRB determined the study as exempt from broad IRB approval. For Open Claims and GermanyDA, approval is provided for OHDSI network studies. For the VA database, use of VA-OMOP was reviewed by the VA Salt Lake City Health Care System Research and Development Committee and was determined to meet the criteria for exemption under Exemption Category 4 (3) and approved the request for Waiver of HIPAA Authorization (IRBnet #1875081-3).

### 2.2 Study design

We employed an active comparator, new-user cohort design. We considered a new-user if their first observed second-line treatment for T2DM is one of the six GLP-1RAs (albiglutide, dulaglutide, exenatide, liraglutide, lixisenatide, semaglutide) or one of the four SGLT2Is (canagliflozin, dapagliflozin, empagliflozin, ertugliflozin). We required each patient to have at least one year of observation time, a recorded T2DM diagnosis within one year and at least 3 months of metformin use before second-line treatment initiation, and no exposure to a comparator second-line drug or more than 30 days of insulin exposure prior to treatment initiation.

We studied six cardiovascular effectiveness outcomes (Supplementary Table 2). There were two primary effectiveness outcomes: (1) 3-point MACE, including acute myocardial infarction, stroke, and sudden cardiac death, and (2) 4-point MACE that additionally included hospitalization or ER visit with heart failure (HHF) [23,41]. We further considered the four individual MACE components as secondary outcomes. Detailed outcome cohort definitions including OMOP concept IDs are provided in the supplement (Section B) of the study protocol [39].

For each outcome, we excluded patients who had experienced the outcome prior to treatment initiation to improve balance in underlying risk profiles between two arms and avoid potential confounding of initiating a treatment due to a prior outcome. We employed an *on-treatment* (OT, also known as “per-protocol”) time-at-risk (TAR) definition that followed a patient until treatment discontinuation, and further considered an *intent-to-treat* (ITT) TAR that followed patients until the end of their observation periods as a secondary design, with time zero being the index of treatment initiation. Continuous drug exposure periods were defined as sequential prescription or dispensing records (dispensing records are available in all claims databases and OptumEHR) with <30-day time gaps.

To further investigate comparative effectiveness among patients with higher risks of cardiovascular diseases (CVD), we performed a secondary analysis focusing on subsets of patients with T2DM *and* established CVD with the same study designs described above. We identified established CVD by requiring at least one condition or procedure indicating established CVD prior to treatment initiation; see Supplementary Section 3.3 and Supplement A.4.8-A.4.10 of the study protocol [39] for more details.

### 2.3 Statistical analysis

To adjust for potential confounding, we used the large-scale propensity score (LSPS) method [42,43] to stratify or match patients within each pair of target and comparator exposure cohorts. This method fits a L-1 regularized logistic regression as the propensity score (PS) model using a large number (often over 10,000) of patient baseline characteristics as predictors; these included demographics, prior conditions, drug exposures, procedures, and lab measurements captured prior to treatment initiation (Supplementary Tables 3-39). L-1 regularization hyper-parameters were selected by 10-fold cross validation. We used 5 strata for PS stratification, and variable-ratio PS matching with a caliper of 0.2 and a maximum of 100 matched units for each target subject. Conditioned on the PS strata/matching-units, we then used Cox proportional hazards models to estimate population-level hazard ratios (HRs) between target and comparator treatments for the risk of each outcome in each data source under each TAR definition (OT and ITT). Across ten GLP-1RAs and SGLT2Is (45 pairwise comparisons) and six outcomes in ten databases using two TAR definitions and two PS adjustment approaches, we generated 45 × 6 × 10 × 2 × 2 = 10,800 HR estimates.

Residual bias from unmeasured and systematic sources often remains in observational studies even after controlling for measured confounding through PS-adjustment [44–46]. For each target-comparator comparison and outcome within each database, we conducted negative control outcome experiments where the null hypothesis of no effect is believed to be true, using a set of 100 control outcomes (Supplementary Table 40). We identified these controls through a data-rich algorithm [47] that identifies prevalent OMOP condition concept occurrences that lack evidence of association with exposures in published literature, drug-product labeling and spontaneous reports; these candidates were then adjudicated by clinical review (see Supplementary Section 10). Using the empirical null distributions learned from these negative controls, we calibrated each HR estimate, and its 95% CI [48]. We considered a calibrated HR to be significantly different from the null when its calibrated 95% CI does not include the null value. Given the large-scale evidence generation nature of the LEGEND framework, our analysis aims to produce comprehensive HR estimates of all pairwise comparisons and all outcomes whenever possible, rather than test specific hypotheses or compute individual p-values.

Within each data source, we executed all comparisons with at least 1,000 eligible subjects per arm. For each comparison within each data source, we performed an extensive array of previously-described study diagnostics [49] to assess the reliability and generalizability. We blinded the effect estimate and only inspected and reported the estimate if the study passed the diagnostics [46,50]. Briefly, these study diagnostics included: (1) preference score distributions between the target and comparator cohorts to evaluate empirical equipoise and generalizability; (2) differences between extensive patient characteristics in each arm to assess cohort balance before and after PS-adjustment [51,52]; (3) minimum detectable risk ratio (MDRR) as a proxy for statistical power [53,54]; (4) empirical calibration plots with negative controls outcomes to inspect residual bias [48]. We only reported HR estimates from analyses that passed all study diagnostics, specifically: ≥ 25% patients having preference scores between 0.3 and 0.7 in both cohorts (≥ 25% empirical equipoise), maximum absolute standardized mean difference (Max SMD) on patient covariates ≤ 0.15 after PS-adjustment, MDRR ≤ 4, and empirical average systematic error (EASE) evaluated from negative control outcome estimates ≤ 0.25. We aggregated HR estimates across data sources using random-effect meta-analysis [55]; only data sources that passed study diagnostics for each comparison contributed to the meta-analytic HR estimate.

### 2.4 Study execution and results dissemination

Pairwise effectiveness comparisons were executed across all databases using an open-source R package^1^ developed using the OHDSI HADES tools [56]. More study design details are available in the study protocol [39]. We have made all results available through a public, interactive R ShinyApp^2^. More results beyond those highlighted in the next section are available for exploration in the interactive R ShinyApp, including details on cohort attribution, PS distributions and fitted PS models, study diagnostics, HR estimates under alternative TAR definitions and alternative diagnostic thresholds.

## 3. Results

### 3.1 Characteristics of study cohorts

Our study identified a total of 1,245,211 second-line new-users of GLP-1RAs and SGLT2Is (**Table 1**). Among all drugs, empagliflozin was the most commonly used (393,499, 31.6%), followed by semaglutide (235,585, 18.9%), dapagliflozin (208,666, 16.8%) and dulaglutide (207,348, 16.8%), whereas ertugliflozin was the least used (13,676, 1.1%) and only found in the US Open Claims database. Across the six GLP-1RAs, only three (dulaglutide, exenatide, and semaglutide) had at least 1,000 new-users in at least one data source, and were thus included in subsequent analyses (i.e., comparative effectiveness involving albiglutide, liraglutide and lixisenatide was not reported). On-treatment time varies by drug as well. New-users of the SGLT2Is have relatively longer on-treatment times than the GLP-1 receptor agonists; within GLP-1RAs, new-users of exenatide and semaglutide had relatively shorter continuous exposure times than those of dulaglutide. The total follow-up times for patients who were initiated on canagliflozin and exenatide were the longest (median time > 2 yrs in most databases), possibly because these two drugs were the first approved within their respective classes. Across all drugs, the median follow-up time was at least one year in most databases.

**Table 1:** Population size and follow-up time for each drug ingredient within each data source. We report the total patient counts, and median and interquartile (IQR) times. When executing comparative studies, we have excluded within-database populations with < 1,000 new users. “On-treatment” =treatment initiation till discontinuation (also known as “per-protocol”). “Total follow-up”= treatment initiation till end of observation.

|  |  | On-treatment<br>time (in days) |  | Total follow-up<br>time (in days) |  |
| --- | --- | --- | --- | --- | --- |
|  | Patients | median | IQR | median | IQR |
| GLP1RA: |  |  |  |  |  |
| dulaglutide |  |  |  |  |  |
| CCAE | 15,697 | 167 | (65 - 569) | 475 | (205 - 1,255) |
| MDCR | 1,160 | 124 | (29 - 470) | 342 | (167 - 881) |
| MDCD | 2,632 | 122 | (51 - 405) | 300 | (128 - 974) |
| Clinformatics | 12,217 | 138 | (58 - 488) | 433 | (184 - 1,176) |
| Open Claims | 161,535 | 154 | (57 - 567) | 717 | (320 - 1,633) |
| OptumEHR | 14,107 | 101 | (83 - 314) | 617 | (306 - 1,412) |
| Total: | 207,348 |  |  |  |  |
| exenatide |  |  |  |  |  |
| CCAE | 2,792 | 93 | (29 - 397) | 816 | (370 - 1,833) |
| MDCR | 263 | 81 | (27 - 245) | 637 | (258 - 1,331) |
| MDCD | 1,177 | 95 | (29 - 355) | 863 | (436 - 1,782) |
| Clinformatics | 2,246 | 104 | (43 - 418) | 813 | (356 - 1,732) |
| Open Claims | 35,632 | 86 | (29 - 380) | 1,589 | (1,026 - 2,489) |
| OptumEHR | 3,367 | 84 | (83 - 241) | 1,118 | (570 - 2,037) |
| Total: | 45,477 |  |  |  |  |
| semaglutide |  |  |  |  |  |
| CCAE | 21,177 | 90 | (33 - 330) | 269 | (111 - 696) |
| MDCR | 1,290 | 83 | (29 - 261) | 197 | (96 - 464) |
| Clinformatics | 18,819 | 89 | (30 - 290) | 242 | (107 - 633) |
| Open Claims | 174,295 | 89 | (41 - 329) | 355 | (182 - 860) |
| CUIMC | 1,250 | 132 | (32 - 463) | 379 | (181 - 821) |
| OptumEHR | 14,064 | 61 | (60 - 187) | 344 | (179 - 791) |
| VA | 4,690 | 134 | (56 - 428) | 474 | (247 - 851) |
| Total: | 235,585 |  |  |  |  |
| SGLT2i: |  |  |  |  |  |
| canagliflozin |  |  |  |  |  |
| CCAE | 9,352 | 154 | (58 - 531) | 853 | (380 - 2,063) |
| MDCR | 954 | 144 | (50 - 547) | 622 | (286 - 1,613) |
| MDCD | 1,879 | 113 | (29 - 458) | 1,207 | (491 - 2,004) |
| Clinformatics | 7,414 | 132 | (56 - 514) | 927 | (416 - 2,045) |
| Open Claims | 111,119 | 128 | (29 - 564) | 2,046 | (1,430 - 2,596) |
| OptumEHR | 10,242 | 51 | (29 - 150) | 1,343 | (722 - 2,258) |
| Total: | 140,960 |  |  |  |  |
| dapagliflozin |  |  |  |  |  |
| CCAE | 17,305 | 160 | (63 - 566) | 524 | (223 - 1,458) |
| GermanyDA | 4,729 | 167 | (97 - 497) | 593 | (288 - 1,600) |
| MDCR | 1,084 | 120 | (58 - 403) | 263 | (137 - 957) |
| MDCD | 1,579 | 94 | (34 - 303) | 303 | (152 - 869) |
| Clinformatics | 7,517 | 105 | (52 - 320) | 269 | (128 - 629) |
| Open Claims | 163,072 | 137 | (59 - 516) | 811 | (337 - 2,145) |
| IMRD | 1,746 | 224 | (74 - 739) | 604 | (269 - 1,146) |
| OptumEHR | 11,634 | 30 | (27 - 118) | 740 | (347 - 1,792) |
| Total: | 208,666 |  |  |  |  |
| empagliflozin |  |  |  |  |  |
| CCAE | 24,019 | 157 | (66 - 528) | 455 | (189 - 1,138) |
| GermanyDA | 4,152 | 186 | (99 - 566) | 727 | (344 - 1,545) |
| MDCR | 3,399 | 98 | (29 - 370) | 276 | (138 - 710) |
| MDCD | 3,249 | 115 | (54 - 379) | 379 | (175 - 834) |
| Clinformatics | 30,420 | 145 | (64 - 470) | 420 | (176 - 1,067) |
| Open Claims | 302,829 | 148 | (66 - 523) | 664 | (313 - 1,496) |
| IMRD | 1,420 | 168 | (58 - 549) | 348 | (143 - 772) |
| OptumEHR | 24,011 | 89 | (89 - 285) | 577 | (279 - 1,302) |
| Total: | 393,499 |  |  |  |  |
| ertugliflozin |  |  |  |  |  |
| Open Claims | 13,676 | 122 | (57 - 414) | 700 | (363 - 1,153) |
| Total: | 13,676 |  |  |  |  |

We also reported drug uptake trends by year. **Figure 1** presents proportional incidence of initiation for a specific agent over all second-line drugs within each data source and by year. For brevity, we presented trends for two most commonly used drugs within each class: dulaglutide, semaglutide, empagliflozin and dapagliflozin (one agent per panel). Proportional incidence is the percentage of incidence of initiating a specific second-line drug over that of *all* second-line drugs in a year and within a data source, shown by a curve in each panel. All four agents had increasing uptake after 2015, with semaglutide and empagliflozin demonstrating the most rapid growth in recent years. Dulaglutide use rose earlier but subsequently plateaued, whereas dapagliflozin displayed greater variability across data sources (more among non-US data than US data). Overall, patterns were consistent between US claims and EHR data, with lower and more heterogeneous uptake in non-US databases. We refer interested readers to our previous class-level drug utilization and characterization study [37] for more analyses.

**Figure 1:**
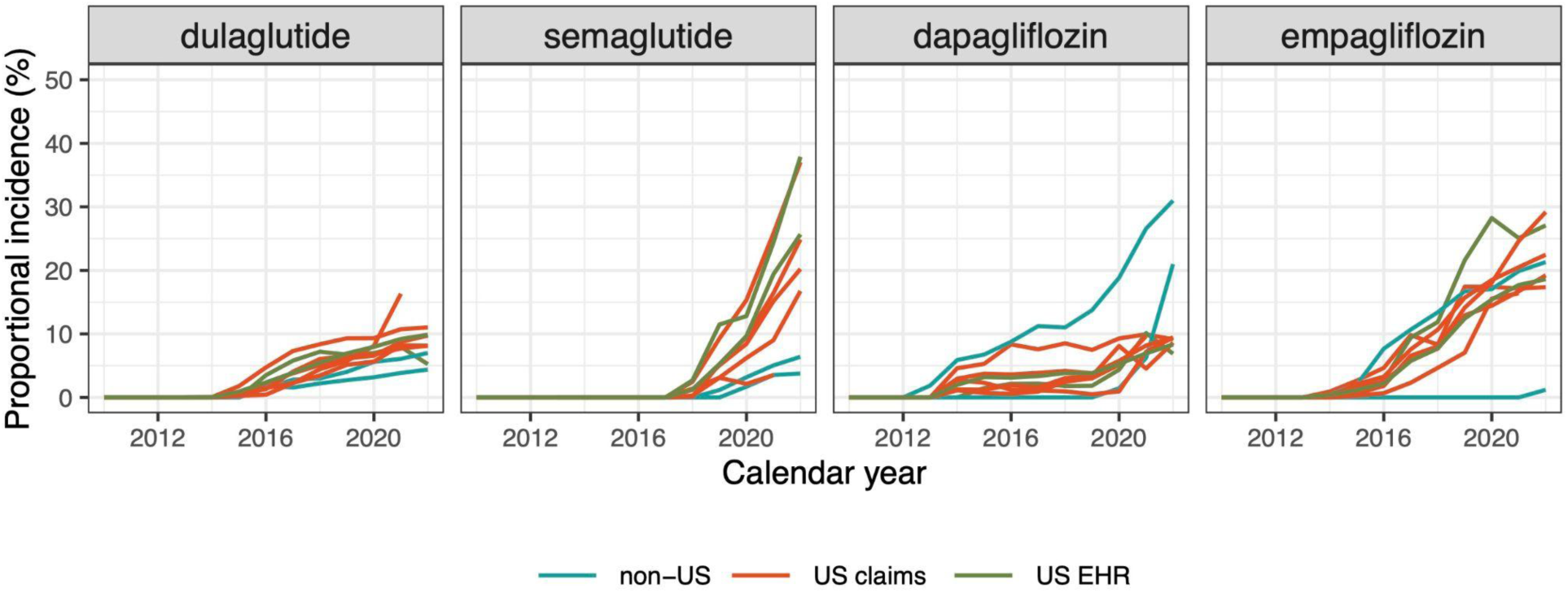
Yearly trends of proportional incidence of second-line drug initiation for a specific antihyperglycemic agent by year within each data source (one curve represents one data source). Here we present initiation trends for two most commonly used drugs within each class: dulaglutide, semaglutide, empagliflozin and dapagliflozin (one agent in each panel). Proportional incidence (in percentage) is calculated by dividing the yearly incidence of an agent by the total yearly incidence of all second-line drugs within a data source. We color data sources by US claims, US EHRs and non-US databases.

We examined patient baseline characteristics before and after PS adjustment (stratification and matching) across all comparisons and databases to ensure balance between target and comparator cohorts. Prior to PS adjustment, we observed differences in age, gender, history of cardiovascular diseases or obesity across all pairwise analyses except for dapagliflozin versus empagliflozin and dulaglutide versus semaglutide, where patients who were younger, female or obese would preferentially initiate on a GLP-1RA over a SGLT2I. In addition, compared with dulaglutide and semaglutide, dapagliflozin and empagliflozin new-users had a higher proportion of established history of cardiovascular diseases, with more noticeable differences in the Clinformatics and MDCD databases. After PS adjustment, the standardized differences (StdDiff) across all characteristics are greatly reduced, implying improved covariate balance (Supplementary Tables 41-75). As an example, **Table 2** presents patient characteristics before and after PS stratification for new-users of semaglutide and empagliflozin within the CCAE database. Notably, after PS stratification, the standardized differences (StdDiff) across all characteristics were greatly reduced, with none exceeding 0.1 in absolute values. We also reported characteristics with the largest absolute estimated coefficients in all fitted PS models (representing highest covariate importance) in Supplementary Tables 3-39.

**Table 2:** Baseline patient characteristics before and after propensity score stratification for semaglutide (T) vs empagliflozin (C) new users in the CCAE data source. For new users in each arm, we report on the mean and standard deviation (std) of age, and proportions satisfying categorical baseline characteristics. We also present the standardized difference (StdDiff) in each characteristic between the two arms. Less extreme StdDiffs suggest improved balance between patient cohorts through propensity score adjustment. Entries under the Medication use section refer to ATC drug classes.

| Characteristic | Before stratification |  |  | After stratification |  |  |
| --- | --- | --- | --- | --- | --- | --- |
|  | T | C | StdDiff | T | C | StdDiff |
| Age: mean | 50.9 | 53.2 | -0.26 | 51.9 | 52.2 | -0.03 |
| Age: std | 9.2 | 8.9 |  | 8.3 | 8.7 |  |
| Characteristic (in proportions) | T (%) | C (%) | StdDiff | T (%) | C (%) | StdDiff |
| Gender: female | 61.1 | 40.3 | 0.42 | 50.0 | 50.9 | -0.02 |
| **Medical history: General** |  |  |  |  |  |  |
| Chronic liver disease | 0.6 | 0.9 | -0.03 | 0.6 | 0.6 | 0.00 |
| Chronic obstructive lung disease | 2.2 | 2.6 | -0.03 | 2.4 | 2.7 | -0.02 |
| Dementia | 0.2 | 0.2 | 0.00 | 0.1 | 0.2 | -0.01 |
| Gastroesophageal reflux disease | 17.2 | 13.6 | 0.10 | 15.6 | 15.7 | 0.00 |
| Hyperlipidemia | 67.6 | 74.3 | -0.15 | 71.0 | 71.6 | -0.01 |
| Hypertensive disorder | 67.2 | 70.9 | -0.08 | 69.1 | 69.7 | -0.01 |
| Obesity | 48.9 | 30.2 | 0.38 | 40.0 | 41.2 | -0.02 |
| Renal impairment | 3.8 | 4.3 | -0.02 | 4.4 | 4.5 | 0.00 |
| Urinary tract infectious disease | 6.6 | 4.5 | 0.09 | 5.5 | 5.2 | 0.01 |
| Acute urinary tract infection | 0.3 | 0.1 | 0.05 | 0.3 | 0.2 | 0.02 |
| Closed fracture of lower limb | 1.1 | 0.9 | 0.02 | 1.1 | 1.2 | -0.01 |
| **Medical history: Cardiovascular disease** |  |  |  |  |  |  |
| Cerebrovascular disease | 1.8 | 2.5 | -0.04 | 2.0 | 2.2 | -0.01 |
| Coronary arteriosclerosis | 5.2 | 10.7 | -0.20 | 7.7 | 8.2 | -0.02 |
| Heart disease | 15.2 | 20.4 | -0.14 | 17.7 | 18.3 | -0.02 |
| Heart failure | 1.8 | 3.4 | -0.10 | 2.7 | 3.0 | -0.02 |
| Ischemic heart disease | 2.9 | 5.7 | -0.14 | 4.1 | 4.5 | -0.02 |
| Peripheral vascular disease | 4.0 | 4.5 | -0.02 | 4.6 | 4.3 | 0.01 |
| Pulmonary embolism | 0.7 | 0.4 | 0.04 | 0.6 | 0.4 | 0.04 |
| Venous thrombosis | 1.0 | 0.8 | 0.02 | 1.0 | 1.0 | 0.00 |
| **Medication use** |  |  |  |  |  |  |
| Agents acting on the renin-angiotensin system | 58.8 | 66.9 | -0.17 | 62.5 | 63.6 | -0.02 |
| Antibacterials for systemic use | 57.7 | 52.3 | 0.11 | 53.9 | 54.4 | -0.01 |
| Antithrombotic agents | 9.2 | 12.6 | -0.11 | 10.9 | 11.3 | -0.01 |
| Beta blocking agents | 23.2 | 26.9 | -0.08 | 24.8 | 25.7 | -0.02 |
| Calcium channel blockers | 20.4 | 21.8 | -0.04 | 21.2 | 21.5 | -0.01 |
| Diuretics | 39.5 | 35.8 | 0.08 | 37.2 | 38.3 | -0.02 |
| Immunosuppressants | 3.9 | 2.8 | 0.06 | 3.5 | 3.7 | -0.01 |
| Lipid modifying agents | 60.9 | 72.5 | -0.25 | 67.3 | 68.0 | -0.01 |

### 3.2 Meta-analytic HR estimates across cardiovascular effectiveness outcomes

We summarized the main results by presenting the meta-analytic HR estimates for the two primary effectiveness outcomes and four secondary outcomes comparing new-users of dulaglutide, semaglutide, dapagliflozin, and empagliflozin (**Figure 2**) whenever estimates are available. We select the two most commonly used drugs within each class as representatives and show calibrated HR estimates with 95% confidence intervals (CI) using an on-treatment (OT) TAR definition with PS stratification.

**Figure 2:**
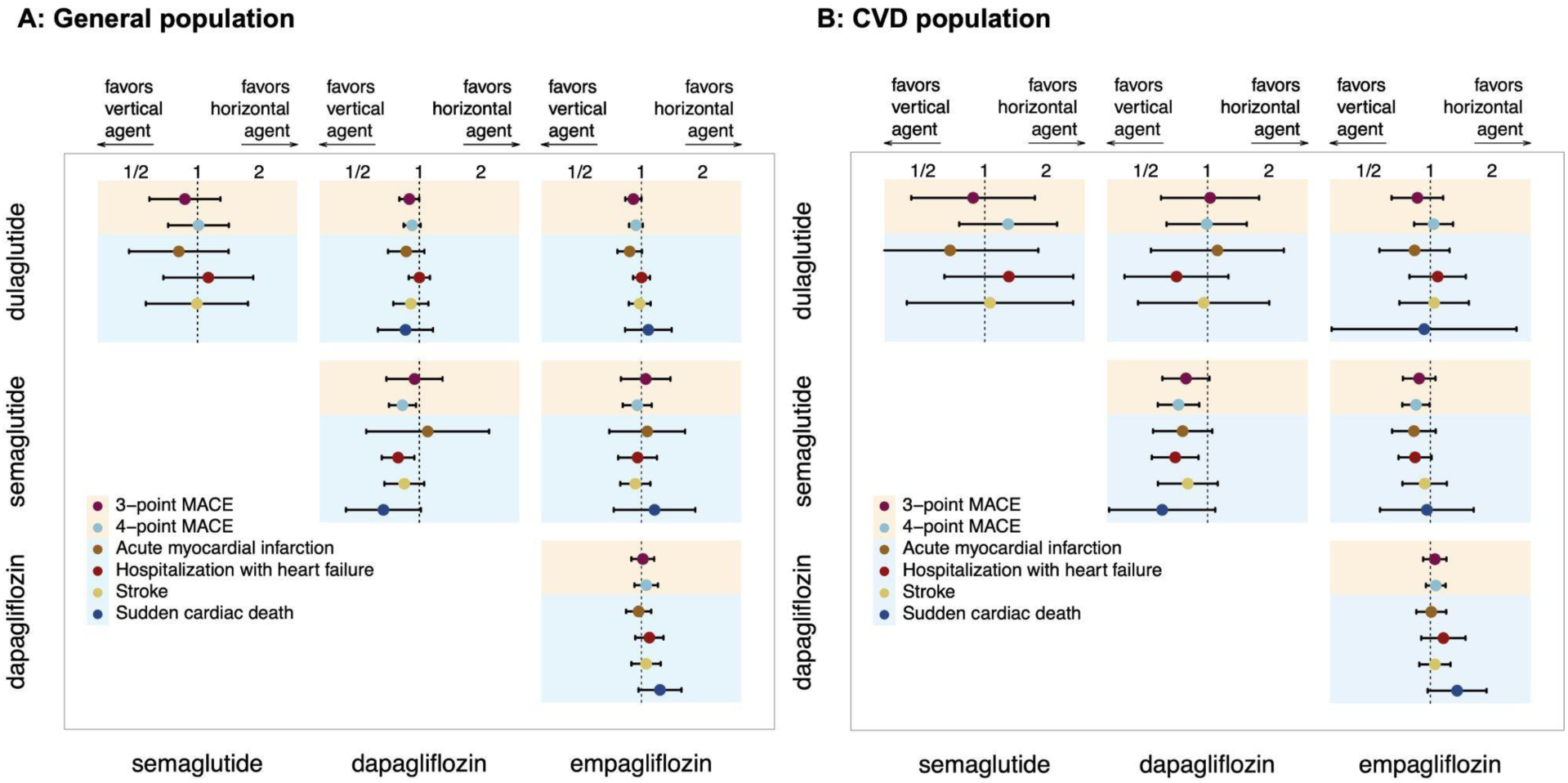
Meta-analytic hazard ratio (HR) estimates for cardiovascular effectiveness between GLP-1RA and SGLT2I drug new-users, for the general population (**A**) and subpopulation with CVD (**B**). Each cell within the matrix plot shows comparisons between the vertical drug as target and horizontal drug as comparator. Points indicate HR estimates and lines show their 95% CIs, both after negative control empirical calibration.

Our study found no significant within-class difference — HR estimates between the two GLP-1RAs, dulaglutide and semaglutide (top-left box), are close to 1 with 95% CIs covering the null (HR=1) across all outcomes, same as those between the two SGLT2Is, dapagliflozin and empagliflozin (bottom-right box). The cross-class comparisons yielded similar findings — the SGLT2Is, dapagliflozin and empagliflozin, show little difference from the GLP-1RAs, semaglutide and dulaglutide, across most outcomes. These results are consistent between the general (panel A) and CVD (panel B) populations, although there is higher uncertainty (wider 95% CIs) for the CVD subgroup analyses, given a population size of only 316,242 (25.4% of the general population; see Supplementary Table 78 for cohort sizes). Furthermore, pairwise comparisons regarding the other agents (GLP-1RA exenatide and SGLT2Is canagliflozin and ertugliflozin) found no significant difference in cardiovascular risks (Supplementary Figure 36). Analyses using PS matching (Supplementary Tables 85&87) yielded similar findings to the main results presented. In addition, results from the intent-to-treat (ITT) time-at-risk definition (Supplementary Tables 88-90), intended to reflect cardiovascular comparative effectiveness in longer-term follow-up, also largely agreed with those from the OT analyses.

We observed a potential risk difference of HHF between semaglutide and dapagliflozin in both the general population (HR = 0.79, 95% CI 0.65-0.94) and the CVD subgroup (HR = 0.70, 95% CI 0.54 - 0.90), and by extension small differences in 4-point MACE (general population HR = 0.83, 95% CI 0.71-0.96, CVD subgroup HR = 0.72, 95% CI 0.57-0.91); see Supplementary Tables 85-86,93-94. Although these comparisons passed study diagnostics, especially with balanced HF-related covariates (Supplementary Table 76), these estimates should be interpreted cautiously. This pattern is unexpected, as it contradicts existing RCT evidence [57–61] indicating stronger heart failure benefits for SGLT2Is such as dapagliflozin. The meta-analytic HR estimates were driven by two US claims databases (CCAE and Open Claims), and residual confounding or outcome misclassification cannot be ruled out, considering coding limitations in the databases. A similar but weaker trend was seen for semaglutide vs empagliflozin in the CVD subgroup (4-point MACE HR=0.85, 95% CI 0.73-0.99). Overall, this finding is exploratory and warrants further investigation.

### 3.3 Results variability across data sources

We further inspected the variability of HR estimates across data sources with the first primary outcome, 3-point MACE, as an example (**Figure 3**). Analyses that failed study diagnostics are grayed out (see an illustration of the pass/fail status of each diagnostic for this outcome in **Figure 4**), and we show meta-analytic estimates summarizing those that passed study diagnostics. Overall, HR estimates are directionally consistent across data sources that passed study diagnostics. At the database level, we still observed largely consistent cardiovascular effectiveness across individual GLP-1RAs and SGLT2Is, both within- and cross-classes. For comparisons where less than three databases passed study diagnostics (dulaglutide vs semaglutide, semaglutide vs dapagliflozin) the resulting meta-analytic HR estimates should be interpreted cautiously. We also provided additional database-level and meta-analytic HR estimates across all comparisons and outcomes using both TAR definitions (OT and ITT) and PS adjustment methods (stratification and matching) in Supplementary Figures 1-12.

**Figure 3:**
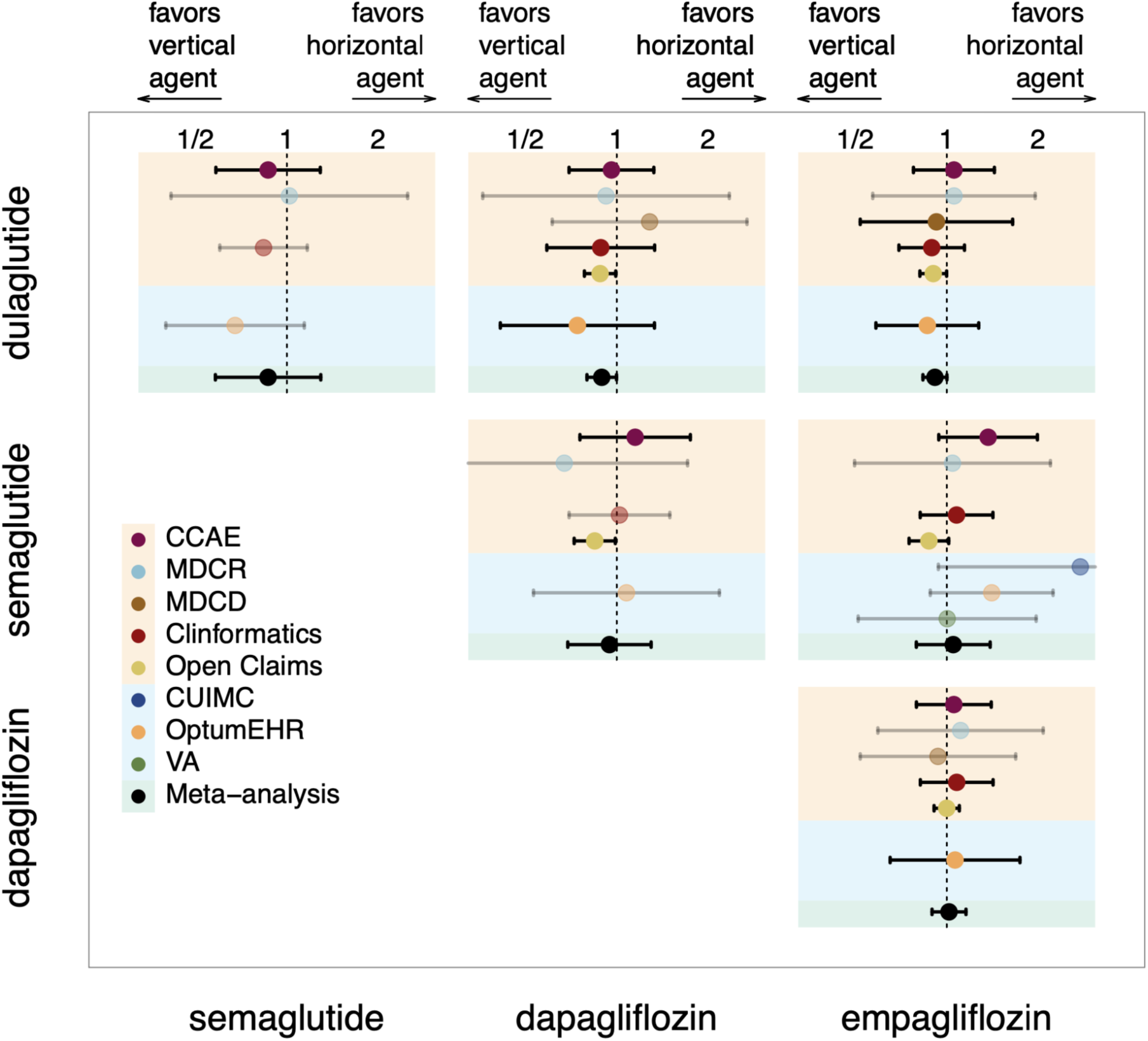
Calibrated HR estimates for 3-point MACE between across new-users of dulaglutide, semaglutide (GLP-1RAs), dapagliflozin and empagliflozin (SGLT2Is), from each data source whenever estimates are available. We also present meta-analytic HR estimates. Each cell within the matrix plot shows comparisons between the vertical drug as target and horizontal drug as comparator. Points indicate HR estimates and lines show their 95% CIs, both after negative control empirical calibration. A database has an empty entry in the plot if a pairwise analysis was not executed or HR estimate was not generated due to small cohort sizes. An entry is grayed out if that comparison failed study diagnostics and thus did not get included in the meta-analysis; the bottom entry in each cell (in black) shows the meta-analytic HR estimate by summarizing data-source level estimates that passed diagnostics.

**Figure 4:**
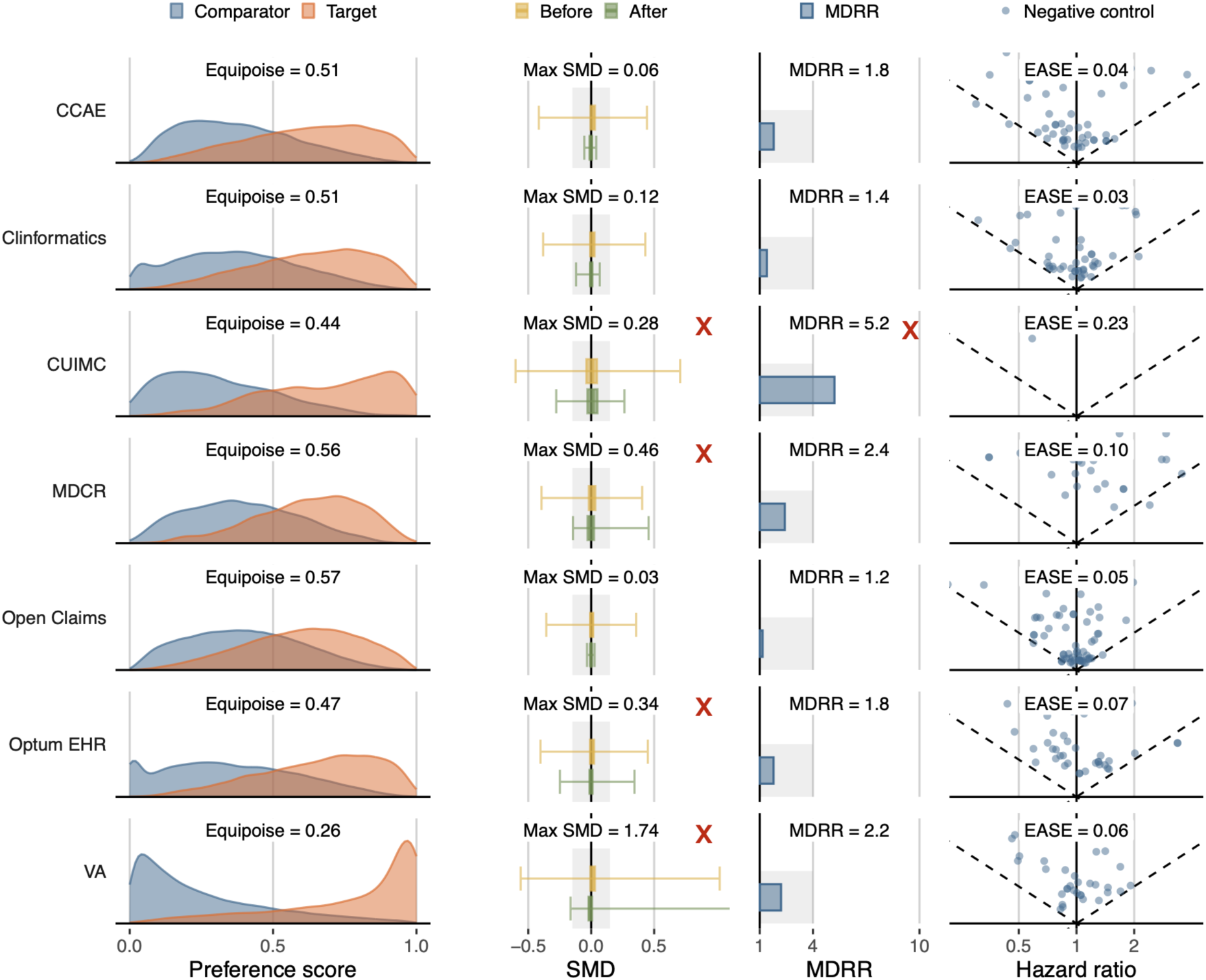
Study diagnostics for comparing new-users of semaglutide (target) and empagliflozin (comparator) on 3-pt MACE. We show diagnostics results on empirical equipoise, covariate balance (standardized mean differences, SMD), minimally detectable relative risk (MDRR), and empirical average systematic error (EASE) evaluated via negative control analysis, for each data source that produced an estimate. We use a red cross sign (**X**) to indicate that this comparison from a data source failed the diagnostic criterion in that column. From left to right, a data source level study would fail diagnostics if equipoise < 0.25, Max SMD > 0.15, MDRR > 4, or EASE > 0.25. For example, in this target-comparator-outcome combination, the study from CUIMC failed on two diagnostic metrics: Max SMD (covariate balance) and MDRR (power). For this comparison, analyses from MDCD and IMRD didn’t produce any estimates due to small cohort sizes and thus were not shown.

### 3.4 Study diagnostics: equipoise, balance, power and residual error

We performed comprehensive study diagnostics for all comparisons (Supplementary Figures 48-53) where more expansive diagnostic results are available on the public results RShinyApp. Overall we observed higher empirical equipoise and better covariate balance between drugs within the same class where new-users exhibited similar characteristics. Across the more commonly used drugs (dulaglutide, semaglutide, dapagliflozin, empagliflozin) and within the larger databases (e.g., CCAE, Clinformatics, OptumEHR, Open Claims) MDRRs were usually small, implying that meta-analytically we had sufficient statistical power to detect that two drugs were similar within a range of 0.8 to 1.2 in their hazard ratio. Here we present an example of study diagnostics for comparing semaglutide vs empagliflozin for 3-point MACE (**Figure 4**). In this example, analyses within CUIMC, MDCR, OptumEHR and VA data sources failed diagnostics mainly due to poor balance, and thus were excluded from subsequent meta-analysis.

## 4. Discussion

To our knowledge, this is the first large-scale study to compare the cardiovascular effectiveness of individual GLP-1RA and SGLT2I agents across multinational real-world health databases. Our study found that, among those drugs evaluated, individual GLP-1RAs and SGLT2Is show similar cardiovascular benefits, within and across drug classes.

While previous studies and meta-analyses have examined these drug classes as a whole [21,24,62], few have investigated at the individual drug level [23,36]. Notably, the 2025 ADA guidelines [32] continue to recommend SGLT2Iand GLP-1RAs largely interchangeably for treating hyperglycemia among T2DM patients with higher CVD risks, without guidance on specific agents within each class. This gap is in part due to the lack of clinical evidence, especially with RCTs, of head-to-head comparisons between individual drugs, both within and across classes. Our study helped fill this evidence gap by a large-scale, multi-site analysis of real-world health databases. Our findings suggest interchangeability in cardiovascular effectiveness among drugs within the same class, while largely supporting similar between-class cardiovascular benefits reported in network meta-analyses of major RCTs [63–65] and in non-interventional studies [24]; see **Figure 5** for a comparison between key results from our study and findings from previously reported results. It will be important to investigate agent-level comparative safety profiles on patient-centered outcomes, in particular on previously reported signals, such as higher rates of genitourinary infections and diabetic ketoacidosis (DKA) for SGLT2Is [66–68], and higher risks of gastrointestinal adverse events and gallbladder diseases for GLP-1RAs [69,70].

**Figure 5:**
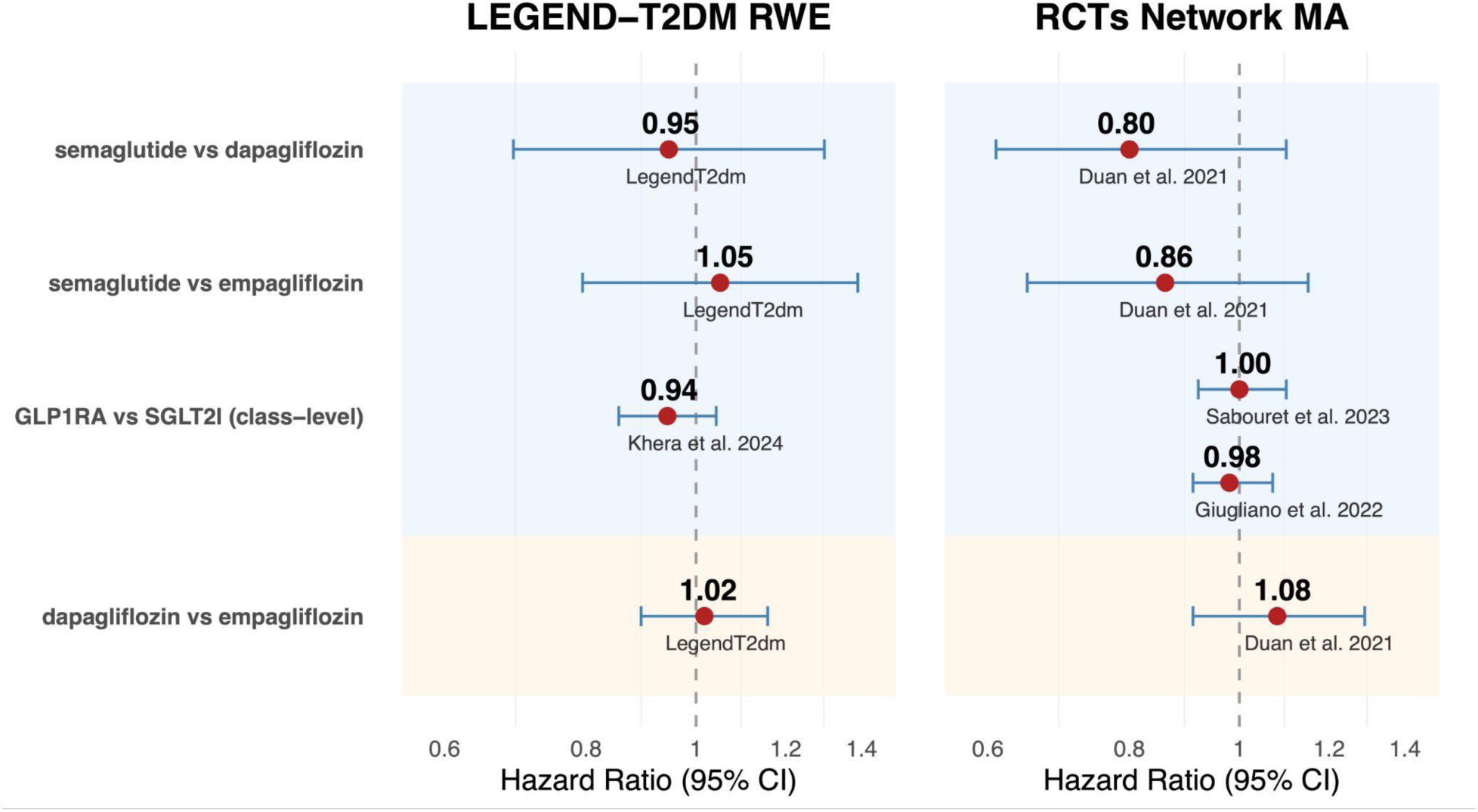
Contextualizing LegendT2dm drug-level comparison results. Comparing HR estimates for 3-point MACE for select drug-level comparisons with results from recent network meta-analyses (network MA) of major CVOTs. The top three rows (light-blue shaded) show cross-class comparisons between GLP-1RA and SGLT2I; the bottom row shows a comparison within the SGLT2I drug class. The left panel shows results from the LEGEND-T2DM study, specifically meta-analytic hazard ratio (HR) estimates using the on-treatment, PS stratified analysis; for completeness we also include the class-level HR estimate from Khera et al. (2024). The right panel shows results from two recent network MAs of major CVOTs: Duan et al. (2021) with ingredient-level HR estimates, and Giugliano et al. (2022) and Sabouret et al. (2023) with class-level HR estimates.

Our study used extensive PS adjustments, negative control empirical calibration, and pre-specified study diagnostics, but residual confounding from time-varying factors associated with discontinuation or switching cannot be excluded. We conducted sensitivity analyses to explore informative censoring on a subset of comparisons and data sources, where we implemented inverse probability censoring weighting (IPCW) with inverse probability treatment weighting (IPTW), and found that additionally using IPCW had limited impact on HR estimates compared with IPTW estimates (Supplementary Tables 99&100). Across ten international databases and more than 1.2 million total patients, our study was sufficiently powered to detect differences — and similarities — at the individual drug level among those most commonly used. Despite the large overall sample size, we did not produce estimates for some comparisons regarding less frequently used agents, due to poor balance or small sample sizes, especially from smaller databases or health systems with restrictive drug formularies (e.g., VA system where only semaglutide vs empagliflozin comparison was feasible).

The hazard difference we found between semaglutide and dapagliflozin associated with HHF, and by extension, 4-point MACE, should be interpreted cautiously as an exploratory finding. Our HHF outcome definition combined hospitalization and ER visits with HF as these two visit contexts often get mixed coding in observational data sources; this practical compromise could have introduced differential outcome misclassification error. As this signal contradicts existing literature supporting HF benefits of SGLT2Is [57–61], further exploration is needed. Future work may involve subgroup analyses on patients with or without history of heart failure, which could be an extension to our subgroup analysis on patients with higher CVD risks.

## Limitations and Future Work

This study carries limitations that could lead to multiple directions of future work. First, we evaluated individual GLP-1RAs and SGLT2Is through mutually exclusive exposure cohort definitions with data only extending through 2022, and therefore newer agents such as tirzepatide were not included in our study, and we could not assess the growing use of combination therapy or study sequential initiations and patient treatment pathways. A future study may evaluate additive or synergistic effects with expanded exposure cohort definitions.

Second, differential adherence and persistence due to differences in comparative safety or tolerability between individual drugs could introduce bias, especially with the ITT analyses where patient follow-up extends beyond discontinuation and with GLP-1RAs where moderately high discontinuation rates have been reported [71,72,73]. Additionally, secular trends — such as guideline shifts, trial spillover, and drug shortages — may also influence treatment initiation patterns that were not fully captured in our study. Given prior knowledge on these important temporal factors, a future study could incorporate them into a time-varying Cox model, although further work is needed to address scalability issues across a large data network. Further, the Cox PH model did not explicitly address competing risks with death or non-proportional hazards which could be addressed by extended survival models.

Our requirement of at least 1,000 patients per arm was intended to robustify study diagnostics as a study with small cohorts sizes could pass all diagnostics simply by chance but still show poor equipoise or balance; this, however, limited the evaluation of less commonly used agents in small databases, and we can explore lower cohort size thresholds in a future study. Similarly, future studies could consider more choices of study diagnostic thresholds as sensitivity analyses. Finally, partial patient overlap across large claims or EHR data sources is possible, as discussed in our study protocol [39]. Future work is also needed to examine comparative safety and patient adherence and persistence to provide strengthened evidence for clinical decisions.

Overall, our findings provide comparative evidence showing that individual GLP-1RAs and SGLT2Is offer broadly similar cardiovascular benefits, and reinforce the cross-drug flexibility in current clinical guidelines. Clinical decisions should be guided by other relevant factors as well, such as safety, drug availability, patient adherence and persistence.

## Data Availability

The study was executed in a distributed manner across the OHDSI data network. The data sources are proprietary, but all interested researchers are welcome to join OHDSI (https://www.ohdsi.org/).

## Acknowledgments

This study was partially funded through the National Institutes of Health grants K23 HL153775, R01 LM006910, R01 HG006139, R01HL69954, R01HL169171, R35GM160458, T15LM00707934 and an Intergovernmental Personnel Act agreement with the US Department of Veterans Affairs. The funders had no role in the design and conduct of the protocol; preparation, review, or approval of the manuscript; and decision to submit the manuscript for publication.

## Supplementary Material

Please refer to the attached supplementary material document.

## List of Abbreviations

CVD: cardiovascular diseases
EHR: electronic health record
GLP-1RA: glucagon-like peptide 1 receptor agonists
HHF: hospitalization or ER visit with heart failure
MACE: major adverse cardiovascular events
OMOP: Observational Medical Outcomes Partnership
PS: propensity score
SGLT2I: sodium-glucose cotransporter-2 inhibitors
T2DM: type-2 diabetes mellitus
TAR: time at risk

## Footnotes

1 The study package is available on Github at: https://github.com/ohdsi-studies/LegendT2dm

2 https://data.ohdsi.org/LegendT2dmDrugEvidenceExplorer/

