## supplement for "Comparative Cardiovascular Effectiveness of Glucagon-Like Peptide 1 Receptor Agonists and Sodium-Glucose Cotransporter-2 Inhibitors in Diabetes Mellitus"

### Supplementary Material

for

### Comparative Cardiovascular Effectiveness of Individual GLP-1RAs and SGLT2Is as Second-Line T2DM Treatments

**11 Additional Analyses with IPCW** **131**

**12 Bibliography** **132**

### 1 Description of data sources included in the study

**Supplementary Table 1:** LEGEND-T2DM data sources and the populations they cover. In all claims databases, dispensing records were available to identify drug exposure periods. In EHR databases, prescription records were used to infer drug exposure periods, except for OptumEHR where dispensing records were also available.

| Data source | Population | Patients | History | Data capture process and short description |
|---|---|---|---|---|
| **US administrative claims** | | | | |
| Merative MarketScan Commercial Claims and Encounters (CCAE) | Commercially insured, < 65 years | 142M | 2000 – | Adjudicated health insurance claims (e.g. inpatient, outpatient, and outpatient pharmacy) from large employers and health plans who provide private healthcare coverage to employees, their spouses and dependents. |
| Merative MarketScan Medicare Supplemental Database (MDCR) | Commercially insured, 65$+$ years | 10M | 2000 – | Adjudicated health insurance claims of retirees with primary or Medicare supplemental coverage through privately insured fee-for-service, point-of-service or capitated health plans. |
| Merative MarketScan Multi-State Medicaid Database (MDCD) | Medicaid enrollees, racially diverse | 26M | 2006 – | Adjudicated health insurance claims for Medicaid enrollees from multiple states and includes hospital discharge diagnoses, outpatient diagnoses and procedures, and outpatient pharmacy claims. |
| IQVIA Open Claims (Open Claims) | General | ~160M | 2010 – | Open, pre-adjudicated claims at anonymized patient level collected from office-based physicians and specialists via office management software and clearinghouse switch sources for the purpose of reimbursement. |
| Optum Clinformatics Data Mart (Clinformatics) | Commercially or Medicare insured | 85M | 2000 – | Inpatient and outpatient healthcare insurance claims. |
| **US electronic health records (EHRs)** | | | | |
| Optum Electronic Health Records (OptumEHR) | US, general | 93M | 2006 – | Clinical information, prescriptions, lab results, vital signs, body measurements, diagnoses and procedures derived from clinical notes using natural language processing. |
| Columbia University Irving Medical Center (CUIMC) | Academic medical center patients, racially diverse | 6M | 1989 – | General practice, specialists and inpatient hospital services from the New York-Presbyterian hospital and affiliated academic physician practices in New York. |
| Department of Veterans Affairs (VA) | Veterans, older, racially diverse | 12M | 2000 – | EHR entries from the national VA health care system, provided at 163 VA medical centers and 1,063 community-based outpatient clinics, with date of death records. |
| **non-US electronic health records (EHRs)** | | | | |
| IQVIA Germany Disease Analyzer (GermanyDA) | Germany, general | 39M | 1992 – | Collection from extracts of patient management software used by general practioners (GPs) and specialists practicing in ambulatory care settings to document patients' medical records within their office-based practice during a visit. |
| IQVIA Medical Research Data UK (IMRD) | UK, general | 6M | 2011 – | Non-identified patient data supplied from UK General Practices. |

**Notes on the VA data source**: The VA data source includes all EHR entries from care that our Veterans receive across all 163 VA medical and health care centers and over 900 community-based outpatient clinics and includes date of death records. The source covers almost 18 million individuals with at least one clinic visit in the VA and almost 9 million individuals with at least one laboratory measure and do not have a date of death recorded at time of study. The source does not link to additional external EHR information that may be available from the Department of Defense nor administrative claims records from Medicare benefits.

#### 2 Primary and secondary outcomes

**Supplementary Table 2:** Description of primary and secondary study outcomes.

| Outcome name | Brief description | Prior development |
|---|---|---|
| **Primary outcomes** | | |
| 3-point MACE | Condition record of acute myocardial infarction, hemorrhagic or ischemic stroke or sudden cardiac death during an inpatient or ER visit | [1–13] |
| 4-point MACE | 3-Point MACE + inpatient or ER visit (hospitalization) with heart failure condition record | [1–20] |
| **Secondary outcomes (individual MACE components)** | | |
| Acute myocardial infarction | Condition record of acute myocardial infarction during an inpatient or ER vist | [1–6] |
| Hospitalization with heart failure | Inpatient or ER visit with heart failure condition record | [14–20] |
| Stroke | Condition record of hemorrhagic or ischemic stroke during an inpatient or ER visit | [7–12] |
| Sudden cardiac death | Condition record of sudden cardiac death during an inpatient or ER visit | [4, 13] |

### 3 Fitted propensity score models and Kaplan-Meier plots between comparisons and data sources

#### 3.1 dulaglutide vs semaglutide

##### 3.1.1 CCAE

**Supplementary Table 3:** Coefficients of top-15 most influential covariates of the fitted propensity model. Positive coefficients indicate predictors of the target exposure.

| Coef | Covariate Name (total selected: 306) |
|---|---|
| 1.97 | index year: 2018 |
| 0.82 | index year: 2019 |
| -0.42 | index year: 2022 |
| 0.31 | index year: 2020 |
| 0.26 | index month: 2 |
| 0.24 | index month: 1 |
| 0.23 | measurement during day -30 through 0 days relative to index: Hemoglobin; glycosylated (A1C) |
| 0.22 | index month: 3 |
| -0.18 | index month: 11 |
| 0.17 | procedure_occurrence during day -365 through 0 days relative to index: Intravenous infusion, hydration; each additional hour (List separately in addition to code for primary procedure) |
| 0.17 | index month: 5 |
| 0.17 | condition_era group during day -30 through 0 days relative to index: Type 2 diabetes mellitus |
| 0.16 | condition_era group during day -30 through 0 days relative to index: Disorder due to type 2 diabetes mellitus |
| -0.16 | condition_era group during day -30 through 0 days relative to index: Obesity |
| -0.16 | drug_era group during day -365 through 0 days relative to index: icosapent ethyl |

**Supplementary Figure 1:** Kaplan Meier curve (top) and number of subjects at risk (bottom) for 3-pt MACE comparing dulaglutide vs semaglutide new-users in the CCAE data source.

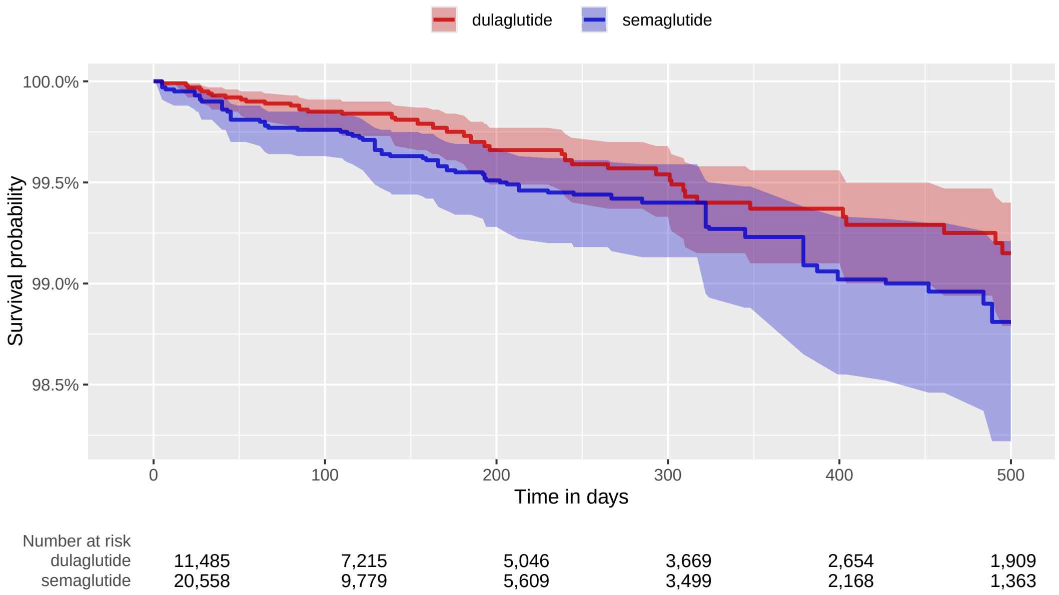

##### 3.1.2 OptumDOD

**Supplementary Table 4:** Coefficients of top-15 most influential covariates of the fitted propensity model. Positive coefficients indicate predictors of the target exposure.

| Coef | Covariate Name (total selected: 176) |
|---|---|
| 3.63 | index year: 2018 |
| 1.17 | index year: 2019 |
| 0.60 | index year: 2020 |
| 0.41 | procedure_occurrence during day -365 through 0 days relative to index: Nursing Facility |
| 0.28 | index month: 1 |
| 0.24 | index year: 2021 |
| 0.23 | measurement during day -30 through 0 days relative to index: Hemoglobin A1c measurement |
| 0.23 | index month: 2 |
| -0.21 | measurement during day -365 through 0 days relative to index: General health panel This panel must include the following: Comprehensive metabolic panel (80053) Blood count, complete (CBC), automated and automated differential WBC count (85025 or 85027 and 85004) OR Blood count, complete (CBC), automated (85027) and |
| 0.18 | index month: 3 |
| 0.18 | condition_era group during day -30 through 0 days relative to index: Disorder due to type 2 diabetes mellitus |
| -0.15 | measurement within normal range during day -365 through 0 days relative to index: Glucose [Mass/volume] in Serum or Plasma |
| -0.14 | measurement within normal range during day -365 through 0 days relative to index: Hemoglobin A1c/Hemoglobin.total in Blood |
| -0.14 | condition_era group during day -365 through 0 days relative to index: Obesity |
| 0.13 | condition_era group during day -30 through 0 days relative to index: Polyneuropathy |

**Supplementary Figure 2:** Kaplan Meier curve (top) and number of subjects at risk (bottom) for 3-pt MACE comparing dulaglutide vs semaglutide new-users in the OptumDOD data source.

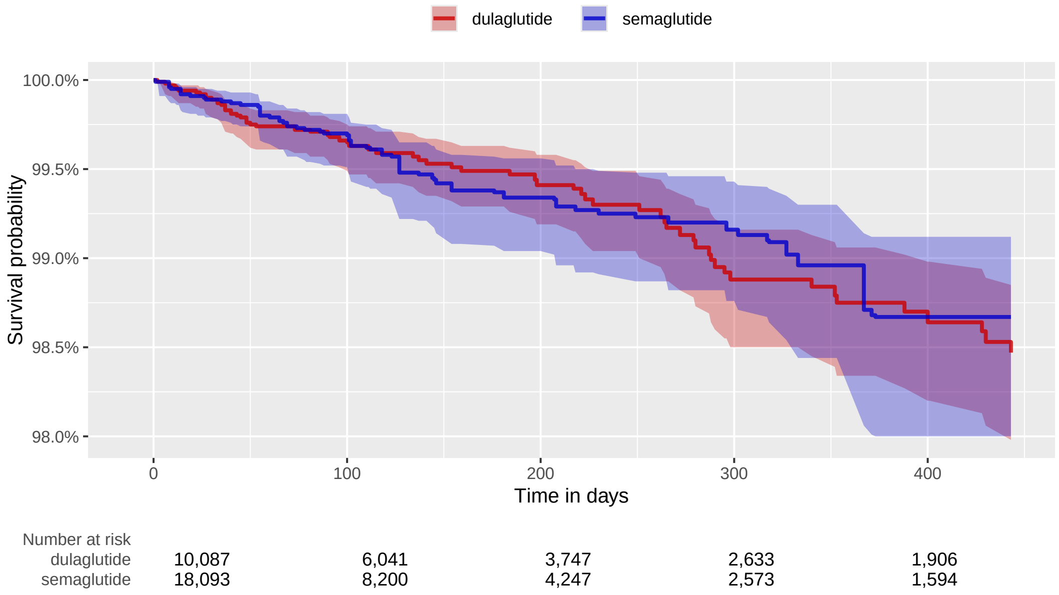

##### 3.1.3 MDCR

**Supplementary Table 5:** Coefficients of top-15 most influential covariates of the fitted propensity model. Positive coefficients indicate predictors of the target exposure.

| Coef | Covariate Name (total selected: 80) |
|---|---|
| 0.66 | index year: 2019 |
| -0.45 | drug_era group during day 0 through 0 days relative to index: CARDIOVASCULAR SYSTEM |
| 0.34 | index year: 2018 |
| 0.32 | device_exposure during day -365 through 0 days relative to index: Blood glucose test or reagent strips for home blood glucose monitor, per 50 strips |
| 0.29 | observation during day -365 through 0 days relative to index: Clinic - General Classification |
| 0.23 | index year: 2021 |
| -0.22 | measurement during day -365 through 0 days relative to index: General health panel This panel must include the following: Comprehensive metabolic panel (80053) Blood count, complete (CBC), automated and automated differential WBC count (85025 or 85027 and 85004) OR Blood count, complete (CBC), automated (85027) and |
| 0.20 | condition_era group during day -30 through 0 days relative to index: Type 2 diabetes mellitus |
| 0.20 | condition_era group during day -30 through 0 days relative to index: Measurement finding outside reference range |
| 0.19 | observation during day -365 through 0 days relative to index: Drugs Identification - Drugs Requiring Detailed Coding |
| -0.15 | condition_era group during day -365 through 0 days relative to index: Obesity |
| -0.13 | drug_era group during day 0 through 0 days relative to index: CALCIUM CHANNEL BLOCKERS AND DIURETICS |
| 0.13 | measurement during day -30 through 0 days relative to index: Hemoglobin A1c measurement |
| 0.12 | measurement within normal range during day -365 through 0 days relative to index: Sodium [Moles/volume] in Serum or Plasma |
| -0.12 | measurement during day -365 through 0 days relative to index: Vitamin D, 25-hydroxy measurement |

**Supplementary Figure 3:** Kaplan Meier curve (top) and number of subjects at risk (bottom) for 3-pt MACE comparing dulaglutide vs semaglutide new-users in the MDCR data source.

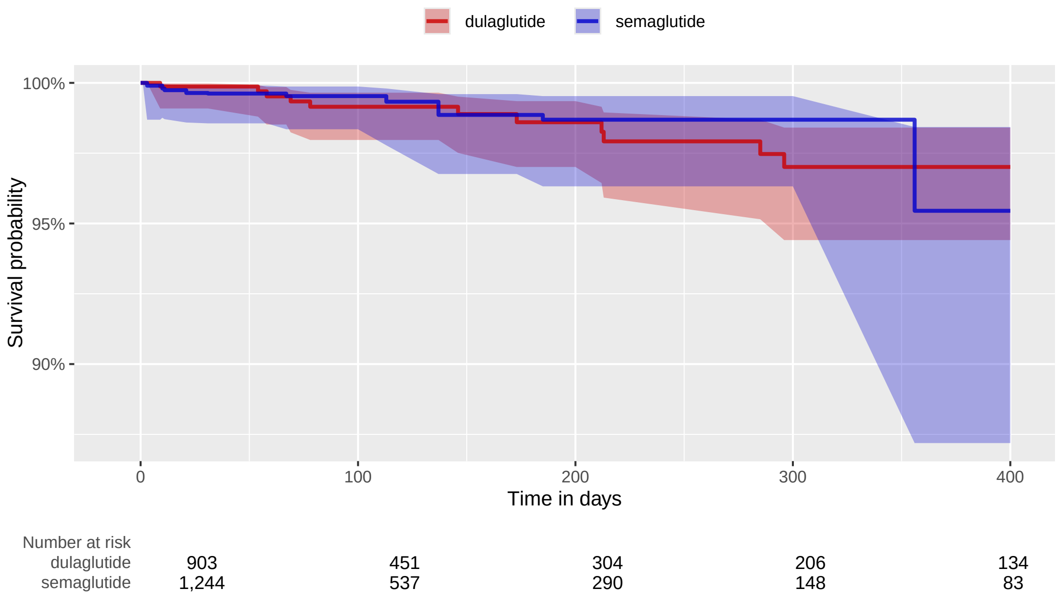

##### 3.1.4 Open Claims

**Supplementary Table 6:** Coefficients of top-15 most influential covariates of the fitted propensity model. Positive coefficients indicate predictors of the target exposure.

| Coef | Covariate Name (total selected: 0) |
|---|---|

##### 3.1.5 OptumEHR

**Supplementary Table 7:** Coefficients of top-15 most influential covariates of the fitted propensity model. Positive coefficients indicate predictors of the target exposure.

| Coef | Covariate Name (total selected: 506) |
|---|---|
| 4.98 | index year: 2017 |
| 2.08 | index year: 2018 |
| 0.92 | index year: 2016 |
| 0.88 | index year: 2019 |
| 0.37 | index year: 2020 |
| 0.35 | measurement during day -365 through 0 days relative to index: 2019-ncov coronavirus, sars-cov-2/2019-ncov (covid-19), any technique, multiple types or subtypes (includes all targets), non-cdc |
| 0.35 | measurement within normal range during day -365 through 0 days relative to index: Urea nitrogen/Creatinine [Mass Ratio] in Blood |
| -0.34 | measurement during day -365 through 0 days relative to index: Osmolality of Urine |
| -0.24 | measurement during day -365 through 0 days relative to index: Depth of Stone |
| 0.24 | measurement below normal range during day -365 through 0 days relative to index: Urea nitrogen/Creatinine [Mass Ratio] in Blood |
| -0.24 | index month: 11 |
| -0.23 | index year: 2022 |
| -0.23 | measurement during day -365 through 0 days relative to index: Globulin [Mass/volume] in Serum |
| -0.22 | observation during day -30 through 0 days relative to index: Dietary management surveillance |
| -0.22 | condition_era group during day -365 through 0 days relative to index: Metabolic syndrome X |

**Supplementary Figure 4:** Kaplan Meier curve (top) and number of subjects at risk (bottom) for 3-pt MACE comparing dulaglutide vs semaglutide new-users in the OptumEHR data source.

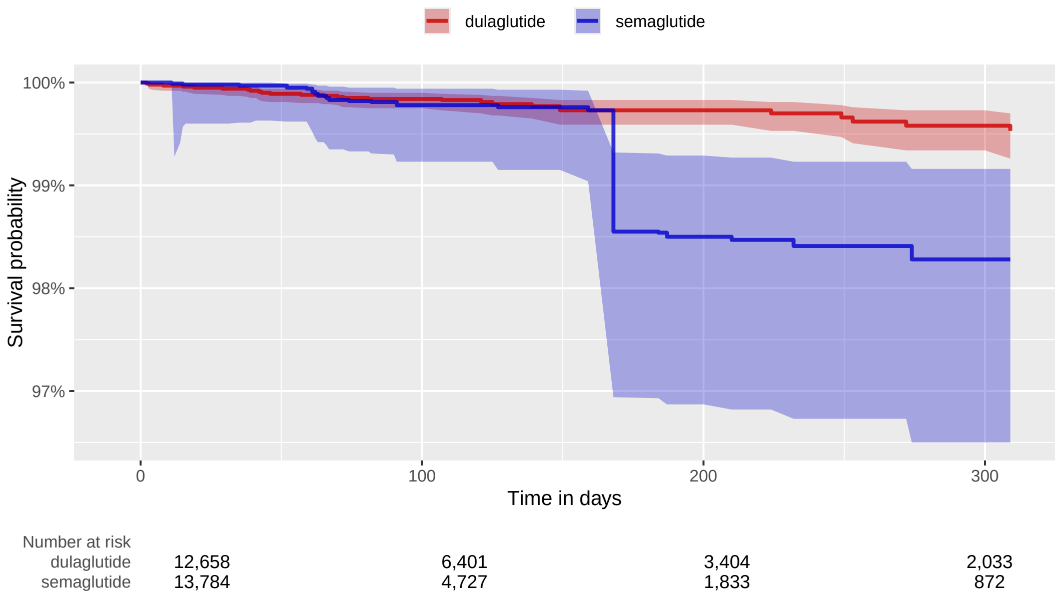

#### 3.2 dulaglutide vs dapagliflozin

##### 3.2.1 CCAE

**Supplementary Table 8:** Coefficients of top-15 most influential covariates of the fitted propensity model. Positive coefficients indicate predictors of the target exposure.

| Coef | Covariate Name (total selected: 601) |
|---|---|
| -2.03 | index year: 2014 |
| -0.95 | index year: 2015 |
| -0.54 | index year: 2016 |
| -0.53 | drug_era group during day -365 through 0 days relative to index: sacubitril |
| 0.46 | observation during day -30 through 0 days relative to index: Diabetes outpatient self-management training services, individual, per 30 minutes |
| -0.37 | condition_era group during day -30 through 0 days relative to index: Heart failure |
| -0.36 | observation during day -365 through 0 days relative to index: Body mass index 20-24 - normal |
| -0.33 | condition_era group during day -30 through 0 days relative to index: Cardiomyopathy |
| -0.32 | condition_era group during day -365 through 0 days relative to index: Chronic kidney disease stage 3A |
| 0.32 | gender = FEMALE |
| -0.31 | condition_era group during day -30 through 0 days relative to index: Myocardial disease |
| 0.29 | drug_era group during day -365 through 0 days relative to index: INSULINS AND ANALOGUES |
| -0.29 | drug_era group during day 0 through 0 days relative to index: ANTIMYCOTICS FOR SYSTEMIC USE |
| 0.25 | observation during day -365 through 0 days relative to index: Abnormal weight gain |
| 0.24 | condition_era group during day -30 through 0 days relative to index: Metabolic syndrome X |

**Supplementary Figure 5:** Kaplan Meier curve (top) and number of subjects at risk (bottom) for 3-pt MACE comparing dulaglutide vs dapagliflozin new-users in the CCAE data source.

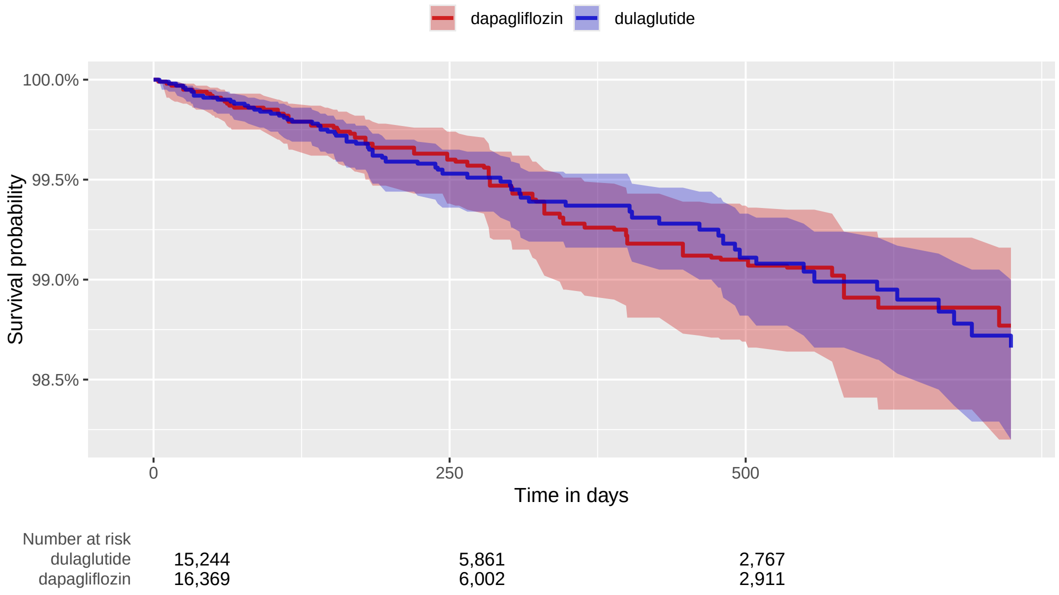

##### 3.2.2 OptumDOD

**Supplementary Table 9:** Coefficients of top-15 most influential covariates of the fitted propensity model. Positive coefficients indicate predictors of the target exposure.

| Coef | Covariate Name (total selected: 629) |
|---|---|
| 2.22 | index year: 2019 |
| 1.79 | index year: 2020 |
| -1.77 | index year: 2015 |
| -1.68 | index year: 2014 |
| 1.28 | index year: 2018 |
| -1.23 | drug_era group during day -365 through 0 days relative to index: sacubitril |
| 0.74 | index year: 2017 |
| 0.54 | drug_era group during day -365 through 0 days relative to index: Other viral vaccines |
| -0.48 | condition_era group during day -365 through 0 days relative to index: Acute heart failure |
| -0.48 | condition_era group during day -365 through 0 days relative to index: Chronic kidney disease stage 3B |
| 0.47 | procedure_occurrence during day -365 through 0 days relative to index: Radiologic examination, chest; single view, frontal (Deprecated) |
| -0.47 | age group: 80 - 84 |
| -0.39 | condition_era group during day -30 through 0 days relative to index: Abnormal cardiovascular function |
| -0.37 | procedure_occurrence during day -365 through 0 days relative to index: Advance care planning including the explanation and discussion of advance directives such as standard forms (with completion of such forms, when performed), by the physician or other qualified health care professional; first 30 minutes, face-to-face with |
| 0.36 | observation during day -365 through 0 days relative to index: Diabetes outpatient self-management training services, individual, per 30 minutes |

**Supplementary Figure 6:** Kaplan Meier curve (top) and number of subjects at risk (bottom) for 3-pt MACE comparing dulaglutide vs dapagliflozin new-users in the OptumDOD data source.

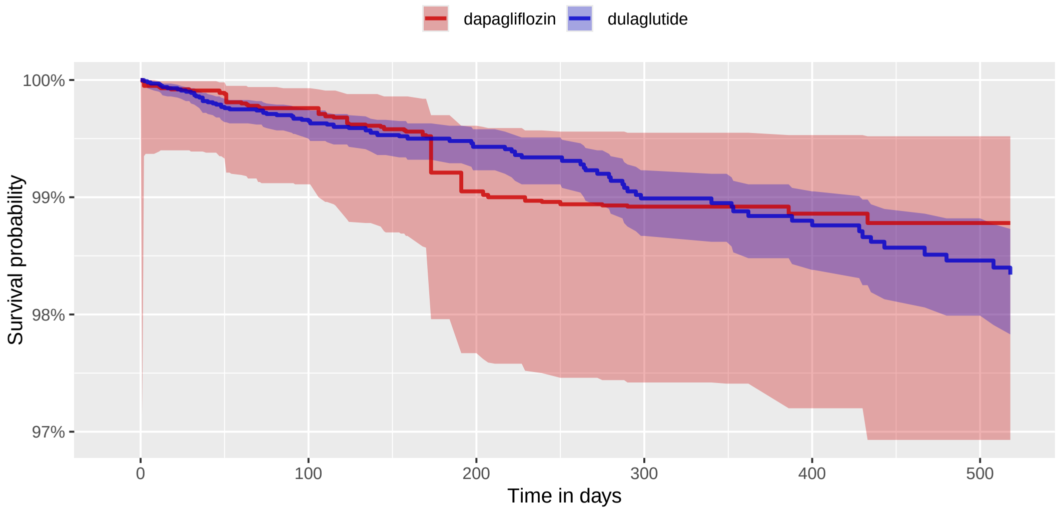

| Number at risk | | | | | | |
|---|---|---|---|---|---|---|
| dulaglutide | 11,770 | 7,085 | 4,480 | 3,165 | 2,309 | 1,690 |
| dapagliflozin | 6,664 | 3,410 | 1,840 | 1,093 | 636 | 354 |

##### 3.2.3 MDCR

**Supplementary Table 10:** Coefficients of top-15 most influential covariates of the fitted propensity model. Positive coefficients indicate predictors of the target exposure.

| Coef | Covariate Name (total selected: 139) |
|---|---|
| 0.55 | condition_era group during day -365 through 0 days relative to index: Obesity |
| 0.48 | measurement within normal range during day -365 through 0 days relative to index: Creatinine [Mass/volume] in Serum or Plasma |
| 0.40 | measurement within normal range during day -365 through 0 days relative to index: Potassium [Moles/volume] in Serum or Plasma |
| -0.39 | condition_era group during day -365 through 0 days relative to index: Chronic kidney disease stage 3B |
| -0.32 | condition_era group during day -365 through 0 days relative to index: Heart failure |
| 0.31 | index year: 2021 |
| -0.30 | procedure_occurrence during day -365 through 0 days relative to index: Brief emotional/behavioral assessment (eg, depression inventory, attention-deficit/hyperactivity disorder [ADHD] scale), with scoring and documentation, per standardized instrument |
| -0.28 | drug_era group during day 0 through 0 days relative to index: HMG CoA reductase inhibitors, other combinations |
| -0.28 | measurement above normal range during day -365 through 0 days relative to index: Urea nitrogen [Mass/volume] in Serum or Plasma |
| -0.24 | drug_era group during day 0 through 0 days relative to index: metformin |
| 0.24 | condition_era group during day -365 through 0 days relative to index: Pain |
| 0.23 | measurement within normal range during day -365 through 0 days relative to index: Sodium [Moles/volume] in Serum or Plasma |
| -0.23 | condition_era group during day -365 through 0 days relative to index: Ankylosis of spine |
| 0.23 | index year: 2019 |
| -0.22 | measurement during day -365 through 0 days relative to index: Urinalysis, by dip stick or tablet reagent for bilirubin, glucose, hemoglobin, ketones, leukocytes, nitrite, pH, protein, specific gravity, urobilinogen, any number of these constituents; automated, with microscopy |

**Supplementary Figure 7:** Kaplan Meier curve (top) and number of subjects at risk (bottom) for 3-pt MACE comparing dulaglutide vs dapagliflozin new-users in the MDCR data source.

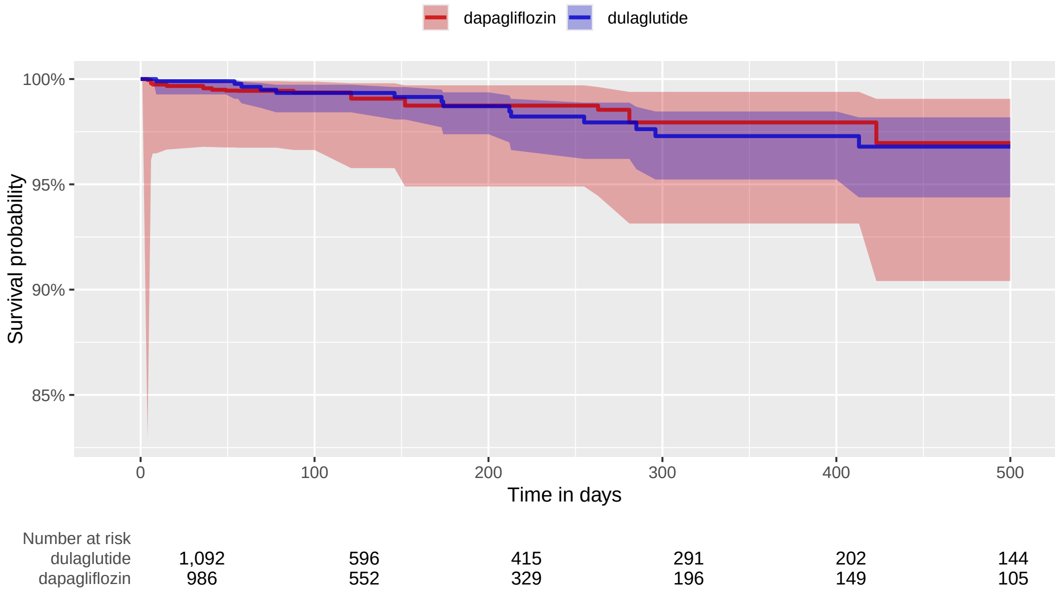

##### 3.2.4 MDCD

**Supplementary Table 11:** Coefficients of top-15 most influential covariates of the fitted propensity model. Positive coefficients indicate predictors of the target exposure.

| Coef | Covariate Name (total selected: 287) |
|---|---|
| -0.98 | condition_era group during day -30 through 0 days relative to index: Congestive heart failure |
| -0.73 | observation during day -365 through 0 days relative to index: Free-Standing Clinic - Other Freestanding Clinic |
| -0.72 | observation during day -30 through 0 days relative to index: Free-Standing Clinic - Rural Health-Clinic |
| -0.63 | observation during day -365 through 0 days relative to index: Non-emergency transportation: mini-bus, mountain area transports, or other transportation systems |
| -0.51 | device_exposure during day -365 through 0 days relative to index: ACCU-CHEK SOFTCLIX LANCETS |
| -0.45 | procedure_occurrence during day -365 through 0 days relative to index: Drug test(s), definitive, utilizing (1) drug identification methods able to identify individual drugs and distinguish between structural isomers (but not necessarily stereoisomers), including, but not limited to gc/ms (any type, single or tandem) and l... |
| -0.43 | device_exposure during day -365 through 0 days relative to index: ACCU-CHEK AVIVA PLUS TEST STRP |
| 0.37 | drug_era group during day -365 through 0 days relative to index: isopropyl alcohol |
| 0.36 | observation during day -365 through 0 days relative to index: Body mass index 40+ - severely obese |
| 0.35 | device_exposure during day -365 through 0 days relative to index: Lens, polycarbonate or equal, any index, per lens |
| -0.34 | device_exposure during day -365 through 0 days relative to index: Blood glucose test or reagent strips for home blood glucose monitor, per 50 strips |
| -0.33 | gender = MALE |
| -0.32 | measurement during day -365 through 0 days relative to index: Lipid panel This panel must include the following: Cholesterol, serum, total (82465) Lipoprotein, direct measurement, high density cholesterol (HDL cholesterol) (83718) Triglycerides (84478) |
| 0.31 | condition_era group during day -365 through 0 days relative to index: Sleep apnea |
| -0.31 | condition_era group during day -365 through 0 days relative to index: Psychotic disorder |

**Supplementary Figure 8:** Kaplan Meier curve (top) and number of subjects at risk (bottom) for 3-pt MACE comparing dulaglutide vs dapagliflozin new-users in the MDCD data source.

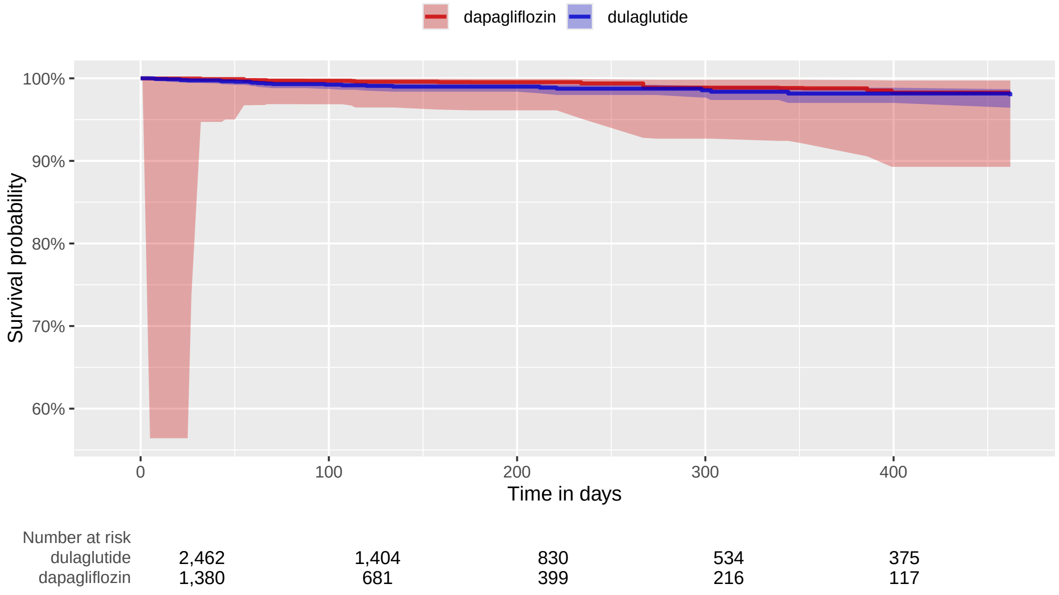

##### 3.2.5 Open Claims

**Supplementary Table 12:** Coefficients of top-15 most influential covariates of the fitted propensity model. Positive coefficients indicate predictors of the target exposure.

| Coef | Covariate Name (total selected: 1777) |
|---|---|
| -3.50 | index year: 2014 |
| -1.60 | index year: 2015 |
| -1.54 | drug_era group (DrugGroupEraLongTerm) during day -365 through 0 days relative to index: sacubitril |
| -1.12 | condition_era group (ConditionGroupEraLongTerm) during day -365 through 0 days relative to index: Chronic kidney disease stage 3B |
| -0.74 | condition_era group (ConditionGroupEraLongTerm) during day -365 through 0 days relative to index: Chronic kidney disease stage 3A |
| -0.70 | index year: 2016 |
| -0.60 | condition_era group (ConditionGroupEraLongTerm) during day -365 through 0 days relative to index: Pancreatitis |
| 0.54 | observation during day -30 through 0 days relative to index: Diabetes outpatient self-management training services, individual, per 30 minutes |
| -0.54 | age group: 85 - 89 |
| -0.50 | age group: 80 - 84 |
| -0.42 | observation during day -365 through 0 days relative to index: Body mass index 20-24 - normal |
| -0.40 | drug_era group (DrugGroupEraLongTerm) during day -365 through 0 days relative to index: icosapent ethyl |
| 0.33 | observation during day -365 through 0 days relative to index: Long-term current use of insulin |
| -0.33 | age group: 75 - 79 |
| -0.33 | condition_era group (ConditionGroupEraShortTerm) during day -30 through 0 days relative to index: Systolic heart failure |

**Supplementary Figure 9:** Kaplan Meier curve (top) and number of subjects at risk (bottom) for 3-pt MACE comparing dulaglutide vs dapagliflozin new-users in the Open Claims data source.

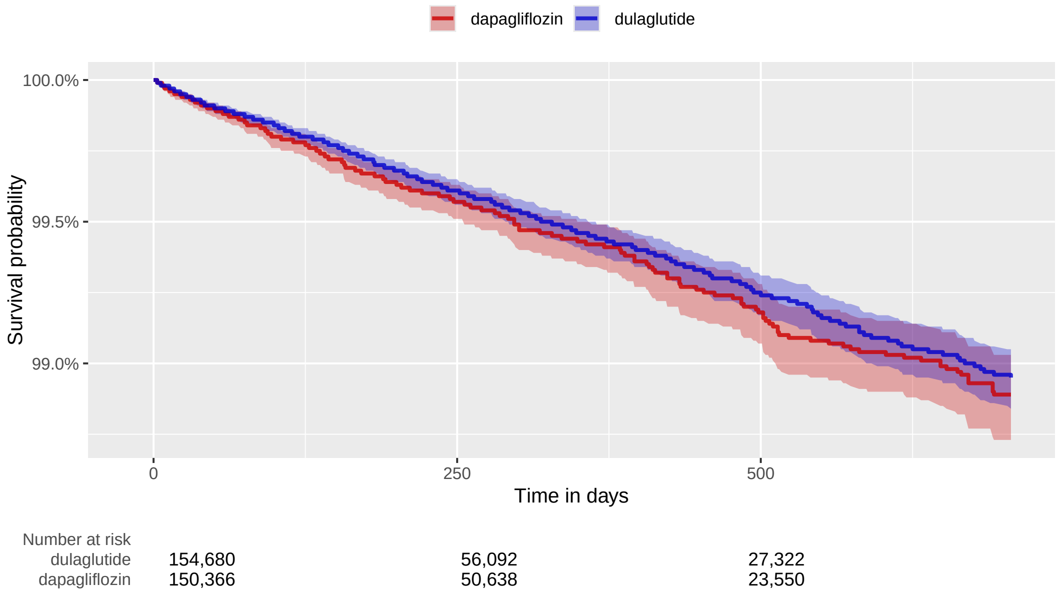

##### 3.2.6 OptumEHR

**Supplementary Table 13:** Coefficients of top-15 most influential covariates of the fitted propensity model. Positive coefficients indicate predictors of the target exposure.

| Coef | Covariate Name (total selected: 739) |
|---|---|
| -1.76 | index year: 2014 |
| -1.52 | index year: 2015 |
| -1.00 | drug_era group during day -365 through 0 days relative to index: sacubitril |
| 0.84 | observation during day -365 through 0 days relative to index: Intensive Care - Psychiatric |
| -0.62 | index year: 2016 |
| -0.40 | gender = MALE |
| 0.39 | procedure_occurrence during day -30 through 0 days relative to index: Office or other outpatient visit for the evaluation and management of a new patient, which requires these 3 key components: A comprehensive history; A comprehensive examination; Medical decision making of high complexity. Counseling and/or co-ordination of |
| -0.37 | observation during day -365 through 0 days relative to index: Medication list documented in medical record (COA) |
| -0.36 | condition_era group during day -30 through 0 days relative to index: Heart failure |
| 0.36 | measurement during day -365 through 0 days relative to index: Bicarbonate [Moles/volume] in Water |
| -0.34 | measurement during day -365 through 0 days relative to index: Alanine aminotransferase [Enzymatic activity/volume] in Serum or Plasma |
| -0.33 | condition_era group during day -30 through 0 days relative to index: Systolic dysfunction |
| -0.32 | observation during day -365 through 0 days relative to index: Body mass index 25-29 - overweight |
| -0.30 | procedure_occurrence during day -30 through 0 days relative to index: Electrocardiogram, routine ECG with at least 12 leads; with interpretation and report |
| -0.28 | age group: 80 - 84 |

**Supplementary Figure 10:** Kaplan Meier curve (top) and number of subjects at risk (bottom) for 3-pt MACE comparing dulaglutide vs dapagliflozin new-users in the OptumEHR data source.

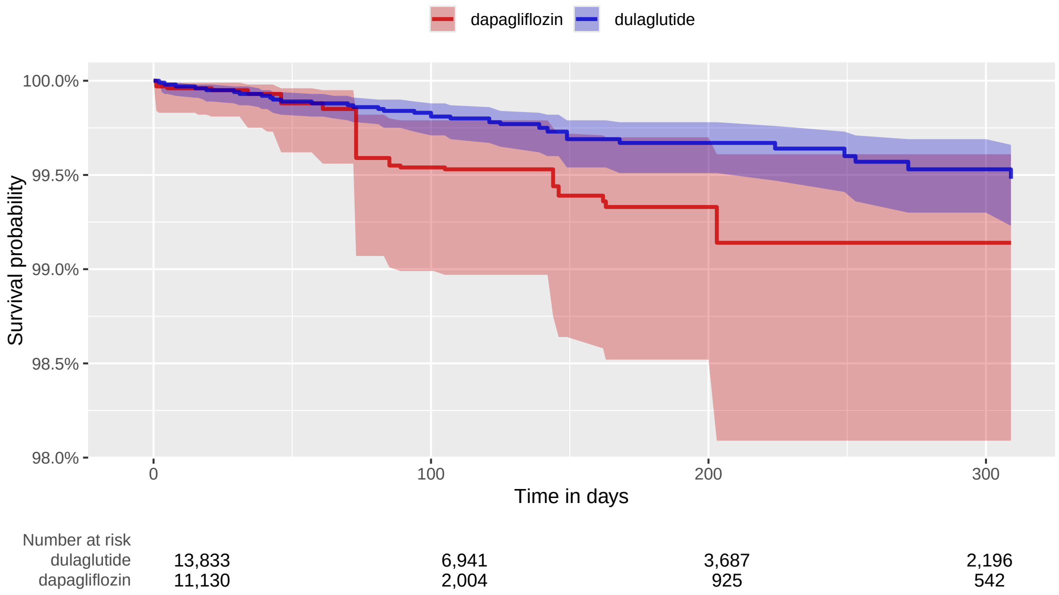

#### 3.3 dulaglutide vs empagliflozin

##### 3.3.1 CCAE

**Supplementary Table 14:** Coefficients of top-15 most influential covariates of the fitted propensity model. Positive coefficients indicate predictors of the target exposure.

| Coef | Covariate Name (total selected: 506) |
|---|---|
| 0.41 | index year: 2018 |
| -0.41 | index year: 2014 |
| 0.38 | gender = FEMALE |
| 0.35 | index year: 2016 |
| -0.34 | condition_era group during day -30 through 0 days relative to index: Myocardial disease |
| 0.34 | observation during day -365 through 0 days relative to index: Abnormal weight gain |
| 0.30 | observation during day -30 through 0 days relative to index: Diabetes outpatient self-management training services, individual, per 30 minutes |
| 0.25 | drug_era group during day -365 through 0 days relative to index: INSULINS AND ANALOGUES |
| 0.22 | condition_era group during day -30 through 0 days relative to index: Obesity |
| 0.22 | measurement during day -365 through 0 days relative to index: Insulin; total |
| -0.21 | observation during day -365 through 0 days relative to index: Body mass index 25-29 - overweight |
| -0.21 | measurement during day -30 through 0 days relative to index: Basic metabolic panel (Calcium, total) This panel must include the following: Calcium, total (82310) Carbon dioxide (bicarbonate) (82374) Chloride (82435) Creatinine (82565) Glucose (82947) Potassium (84132) Sodium (84295) Urea nitrogen (BUN) (84520) |
| 0.20 | condition_era group during day -365 through 0 days relative to index: Obesity |
| 0.20 | age group: 35 - 39 |
| -0.20 | condition_era group during day -30 through 0 days relative to index: Heart failure |

**Supplementary Figure 11:** Kaplan Meier curve (top) and number of subjects at risk (bottom) for 3-pt MACE comparing dulaglutide vs empagliflozin new-users in the CCAE data source.

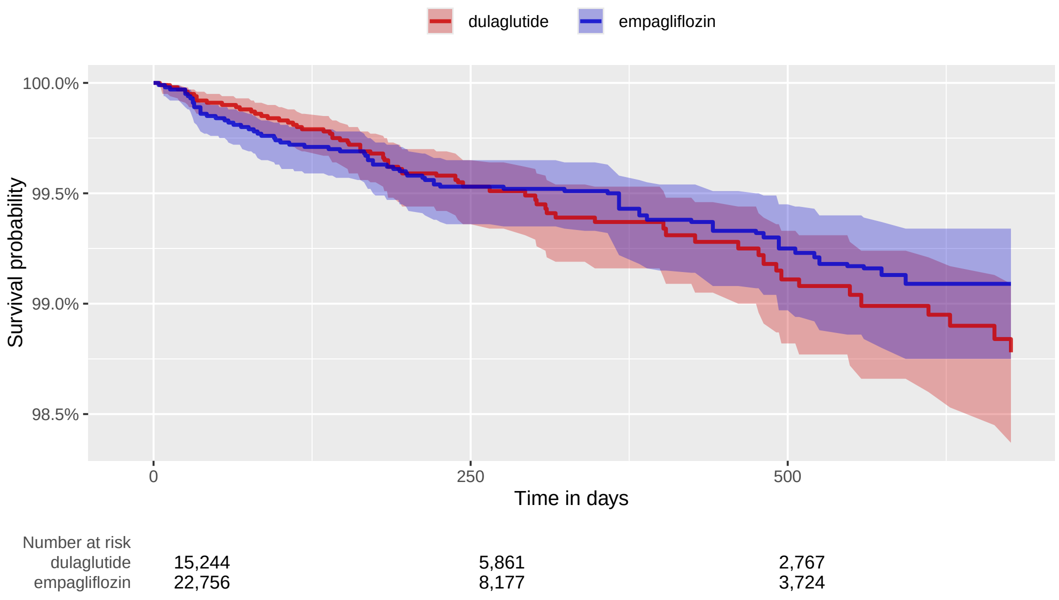

##### 3.3.2 OptumDOD

**Supplementary Table 15:** Coefficients of top-15 most influential covariates of the fitted propensity model. Positive coefficients indicate predictors of the target exposure.

| Coef | Covariate Name (total selected: 580) |
|---|---|
| -0.46 | drug_era group during day -365 through 0 days relative to index: sacubitril |
| 0.41 | observation during day -30 through 0 days relative to index: Diabetes outpatient self-management training services, individual, per 30 minutes |
| -0.33 | condition_era group during day -30 through 0 days relative to index: Congestive heart failure |
| -0.32 | age group: 80 - 84 |
| -0.31 | index year: 2015 |
| 0.29 | gender = FEMALE |
| 0.28 | condition_era group during day -365 through 0 days relative to index: Metabolic syndrome X |
| 0.27 | observation during day -365 through 0 days relative to index: Body mass index 40+ - severely obese |
| 0.26 | observation during day -365 through 0 days relative to index: Set-up portable x-ray equipment |
| 0.26 | index year: 2018 |
| 0.23 | procedure_occurrence during day -365 through 0 days relative to index: Nursing Facility |
| 0.23 | condition_era group during day -30 through 0 days relative to index: Obesity |
| -0.22 | observation during day -365 through 0 days relative to index: Body mass index 20-24 - normal |
| 0.21 | observation during day -365 through 0 days relative to index: Long-term current use of insulin |
| -0.21 | observation during day -365 through 0 days relative to index: Intensive Care - Post ICU |

**Supplementary Figure 12:** Kaplan Meier curve (top) and number of subjects at risk (bottom) for 3-pt MACE comparing dulaglutide vs empagliflozin new-users in the OptumDOD data source.

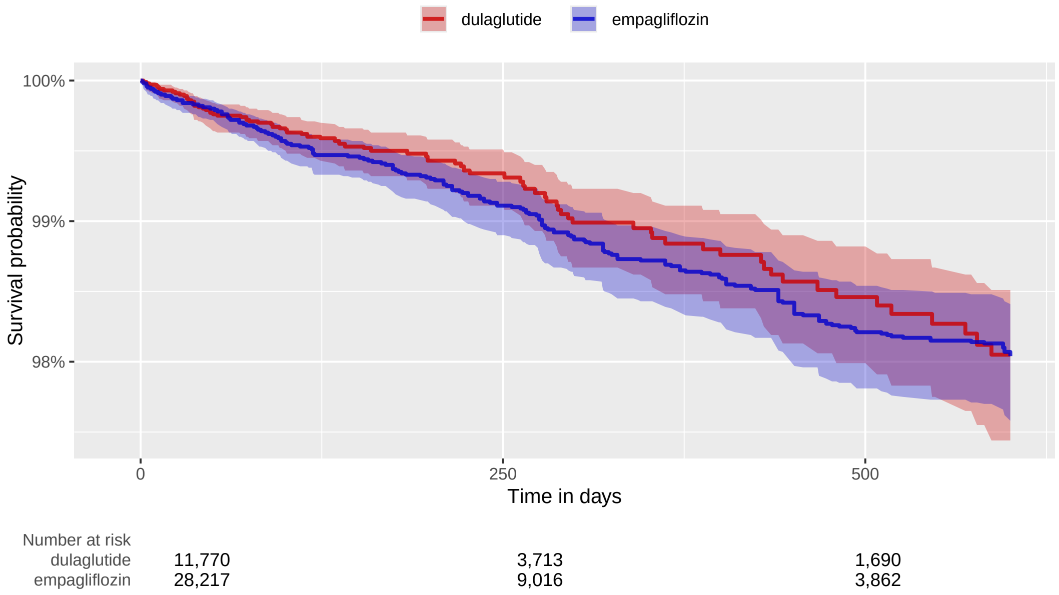

##### 3.3.3 MDCR

**Supplementary Table 16:** Coefficients of top-15 most influential covariates of the fitted propensity model. Positive coefficients indicate predictors of the target exposure.

| Coef | Covariate Name (total selected: 87) |
|---|---|
| -0.40 | condition_era group during day -365 through 0 days relative to index: Heart failure |
| 0.26 | condition_era group during day -365 through 0 days relative to index: Obesity |
| -0.25 | condition_era group during day -30 through 0 days relative to index: Vascular disorder |
| 0.24 | condition_era group during day -30 through 0 days relative to index: Obesity |
| 0.24 | condition_era group during day -365 through 0 days relative to index: Breathing-related sleep disorder |
| -0.24 | condition_era group during day -30 through 0 days relative to index: Heart disease |
| 0.23 | gender = FEMALE |
| 0.23 | index year: 2016 |
| 0.20 | drug_era group during day -365 through 0 days relative to index: Opioids in combination with antispasmodics |
| -0.17 | observation during day -365 through 0 days relative to index: Body mass index 25-29 - overweight |
| 0.16 | drug_era group during day -365 through 0 days relative to index: PSYCHOANALEPTICS |
| 0.14 | observation during day -365 through 0 days relative to index: Free-Standing Clinic - Rural Health-Clinic |
| 0.14 | age group: 65 - 69 |
| -0.13 | condition_era group during day -365 through 0 days relative to index: Myocardial disease |
| 0.12 | drug_era group during day -365 through 0 days relative to index: INTESTINAL ANTIINFECTIVES |

**Supplementary Figure 13:** Kaplan Meier curve (top) and number of subjects at risk (bottom) for 3-pt MACE comparing dulaglutide vs empagliflozin new-users in the MDCR data source.

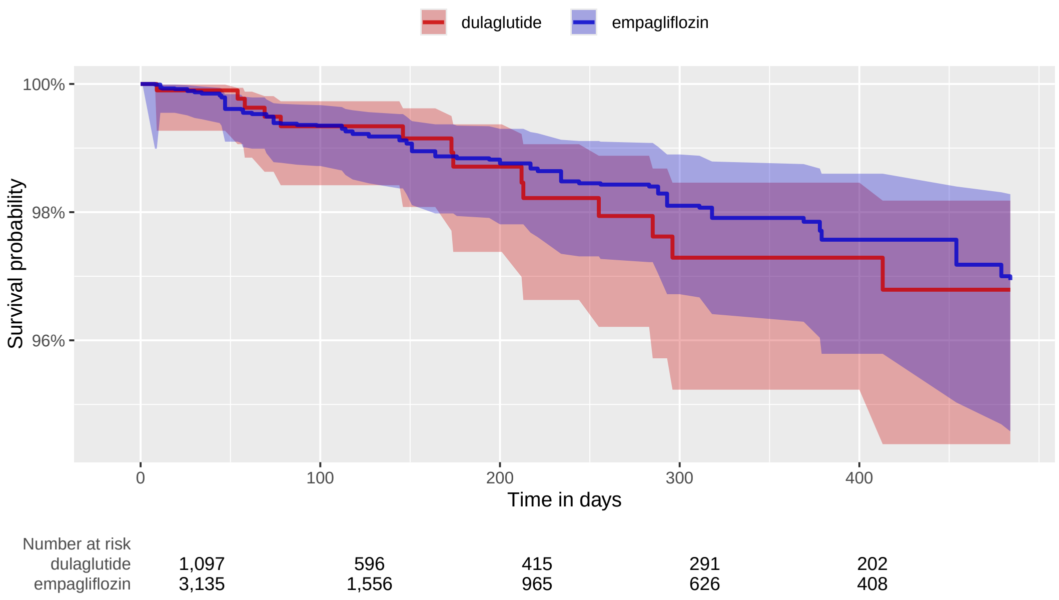

##### 3.3.4 MDCD

**Supplementary Table 17:** Coefficients of top-15 most influential covariates of the fitted propensity model. Positive coefficients indicate predictors of the target exposure.

| Coef | Covariate Name (total selected: 245) |
|---|---|
| -0.65 | observation during day -365 through 0 days relative to index: Non-emergency transportation: mini-bus, mountain area transports, or other transportation systems |
| -0.61 | index year: 2020 |
| -0.46 | device_exposure during day -365 through 0 days relative to index: ACCU-CHEK SOFTCLIX LANCETS |
| 0.40 | device_exposure during day -365 through 0 days relative to index: FREESTYLE LITE TEST STRIP |
| -0.33 | gender = MALE |
| 0.30 | condition_era group during day -365 through 0 days relative to index: Obesity |
| 0.29 | device_exposure during day -30 through 0 days relative to index: FREESTYLE 28G LANCETS |
| -0.29 | observation during day -365 through 0 days relative to index: Ambulatory Surgical Care - General Classification |
| -0.29 | condition_era group during day -365 through 0 days relative to index: Myocardial disease |
| -0.28 | condition_era group during day -365 through 0 days relative to index: Congestive heart failure |
| -0.27 | age group: 60 - 64 |
| 0.27 | observation during day -365 through 0 days relative to index: Body mass index 40+ - severely obese |
| -0.23 | observation during day -365 through 0 days relative to index: Review of all medications by a prescribing practitioner or clinical pharmacist (such as, prescriptions, OTCs, herbal therapies and supplements) documented in the medical record (COA) |
| 0.23 | condition_era group during day -365 through 0 days relative to index: Sleep apnea |
| -0.22 | observation during day -365 through 0 days relative to index: Body mass index 25-29 - overweight |

**Supplementary Figure 14:** Kaplan Meier curve (top) and number of subjects at risk (bottom) for 3-pt MACE comparing dulaglutide vs empagliflozin new-users in the MDCD data source.

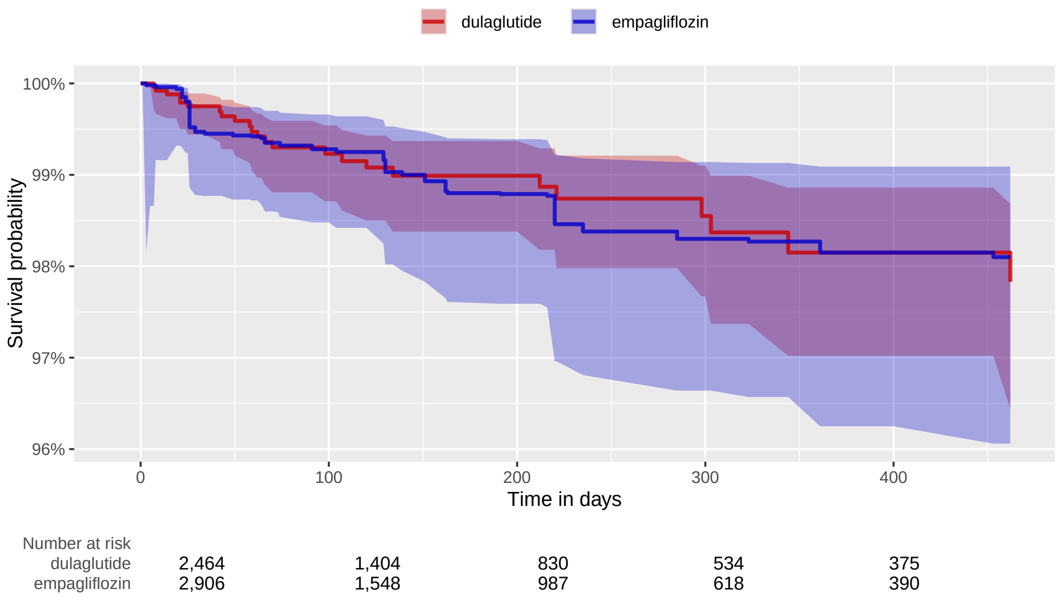

##### 3.3.5 Open Claims

**Supplementary Table 18:** Coefficients of top-15 most influential covariates of the fitted propensity model. Positive coefficients indicate predictors of the target exposure.

| Coef | Covariate Name (total selected: 1461) |
|---|---|
| -1.47 | index year: 2014 |
| -0.94 | drug_era group (DrugGroupEraLongTerm) during day -365 through 0 days relative to index: sacubitril |
| -0.66 | condition_era group (ConditionGroupEraLongTerm) during day -365 through 0 days relative to index: Pancreatitis |
| -0.60 | age group: 80 - 84 |
| -0.53 | age group: 75 - 79 |
| -0.46 | age group: 85 - 89 |
| -0.45 | observation during day -365 through 0 days relative to index: Body mass index 20-24 - normal |
| 0.40 | age group: 20 - 24 |
| 0.40 | index year: 2016 |
| 0.40 | observation during day -30 through 0 days relative to index: Diabetes outpatient self-management training services, individual, per 30 minutes |
| -0.39 | age group: 70 - 74 |
| 0.36 | age group: 25 - 29 |
| -0.35 | condition_era group (ConditionGroupEraLongTerm) during day -365 through 0 days relative to index: Chronic kidney disease stage 3A |
| -0.34 | condition_era group (ConditionGroupEraLongTerm) during day -365 through 0 days relative to index: Chronic kidney disease stage 3B |
| 0.32 | age group: 30 - 34 |

**Supplementary Figure 15:** Kaplan Meier curve (top) and number of subjects at risk (bottom) for 3-pt MACE comparing dulaglutide vs empagliflozin new-users in the Open Claims data source.

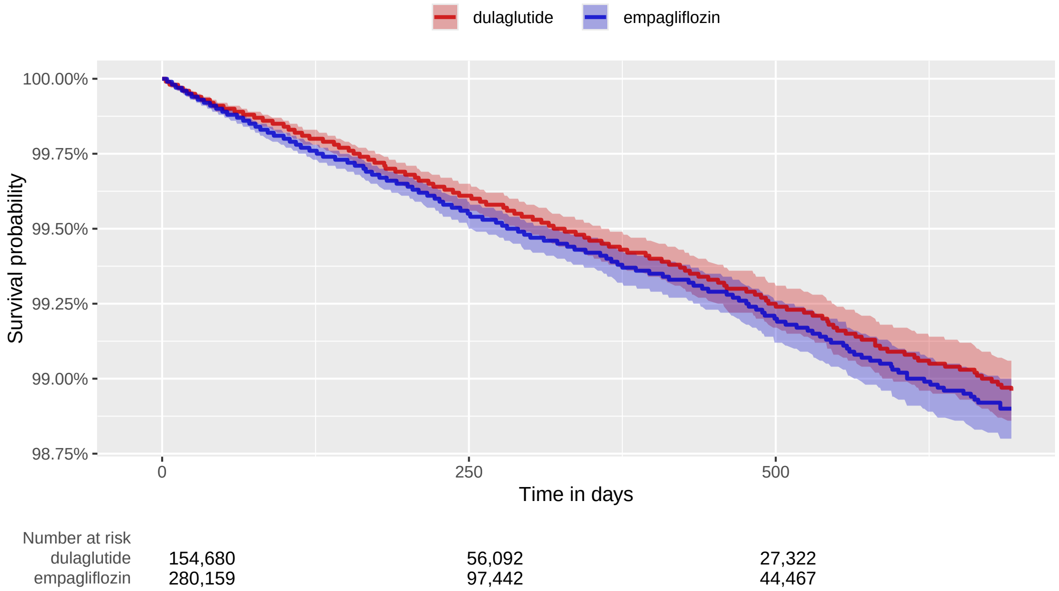

##### 3.3.6 OptumEHR

**Supplementary Table 19:** Coefficients of top-15 most influential covariates of the fitted propensity model. Positive coefficients indicate predictors of the target exposure.

| Coef | Covariate Name (total selected: 809) |
|---|---|
| 1.00 | observation during day -365 through 0 days relative to index: Intensive Care - Psychiatric |
| -0.58 | age group: 80 - 84 |
| 0.53 | index year: 2016 |
| -0.48 | age group: 75 - 79 |
| -0.46 | drug_era group during day -365 through 0 days relative to index: sacubitril |
| 0.41 | gender = FEMALE |
| 0.36 | observation during day -365 through 0 days relative to index: Abnormal weight gain |
| -0.35 | observation during day -365 through 0 days relative to index: Screening for depression is documented as negative, a follow-up plan is not required |
| -0.34 | observation during day -365 through 0 days relative to index: Body mass index 20-24 - normal |
| -0.31 | race = Asian |
| 0.29 | age group: 30 - 34 |
| -0.29 | age group: 70 - 74 |
| 0.28 | measurement during day -30 through 0 days relative to index: Creatine measurement, urine |
| 0.28 | age group: 25 - 29 |
| -0.27 | drug_era group during day -365 through 0 days relative to index: metoclopramide |

**Supplementary Figure 16:** Kaplan Meier curve (top) and number of subjects at risk (bottom) for 3-pt MACE comparing dulaglutide vs empagliflozin new-users in the OptumEHR data source.

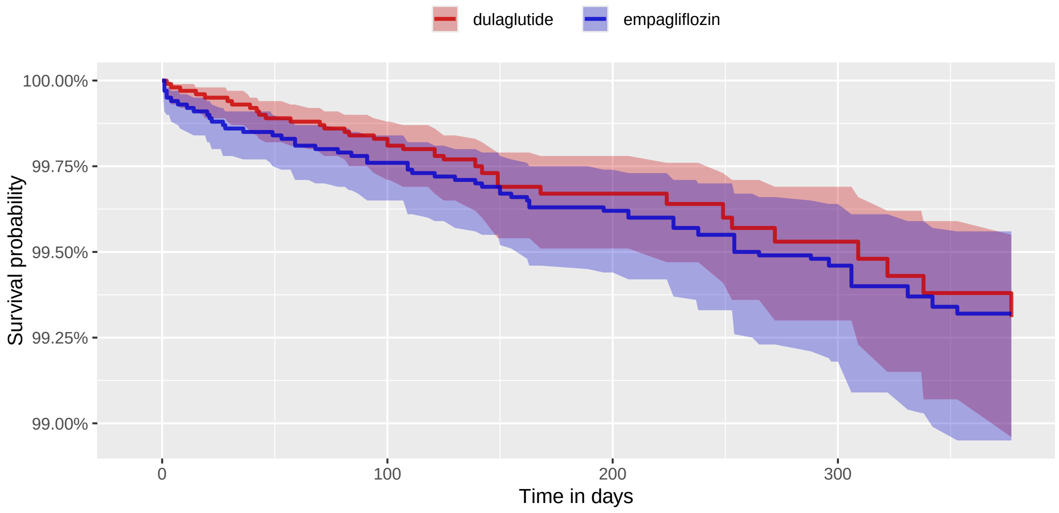

| Number at risk | | | | |
|---|---|---|---|---|
| dulaglutide | 13,833 | 6,941 | 3,687 | 2,196 |
| empagliflozin | 23,022 | 10,136 | 5,452 | 3,157 |

#### 3.4 semaglutide vs dapagliflozin

##### 3.4.1 CCAE

**Supplementary Table 20:** Coefficients of top-15 most influential covariates of the fitted propensity model. Positive coefficients indicate predictors of the target exposure.

| Coef | Covariate Name (total selected: 501) |
|---|---|
| -2.09 | index year: 2018 |
| -0.69 | index year: 2019 |
| -0.68 | condition_era group during day -30 through 0 days relative to index: Heart failure |
| -0.58 | condition_era group during day -365 through 0 days relative to index: Systolic dysfunction |
| -0.55 | drug_era group during day -30 through 0 days relative to index: sacubitril |
| -0.51 | condition_era group during day -365 through 0 days relative to index: Chronic kidney disease stage 3A |
| 0.47 | index year: 2022 |
| 0.42 | observation during day -365 through 0 days relative to index: Abnormal weight gain |
| -0.39 | gender = MALE |
| 0.35 | condition_era group during day -30 through 0 days relative to index: Obesity |
| -0.35 | index year: 2020 |
| -0.32 | measurement during day -30 through 0 days relative to index: Hemoglobin; glycosylated (A1C) |
| -0.31 | condition_era group during day -30 through 0 days relative to index: Myocardial disease |
| 0.29 | condition_era group during day -365 through 0 days relative to index: Prediabetes |
| 0.27 | condition_era group during day -30 through 0 days relative to index: Metabolic syndrome X |

**Supplementary Figure 17:** Kaplan Meier curve (top) and number of subjects at risk (bottom) for 3-pt MACE comparing semaglutide vs dapagliflozin new-users in the CCAE data source.

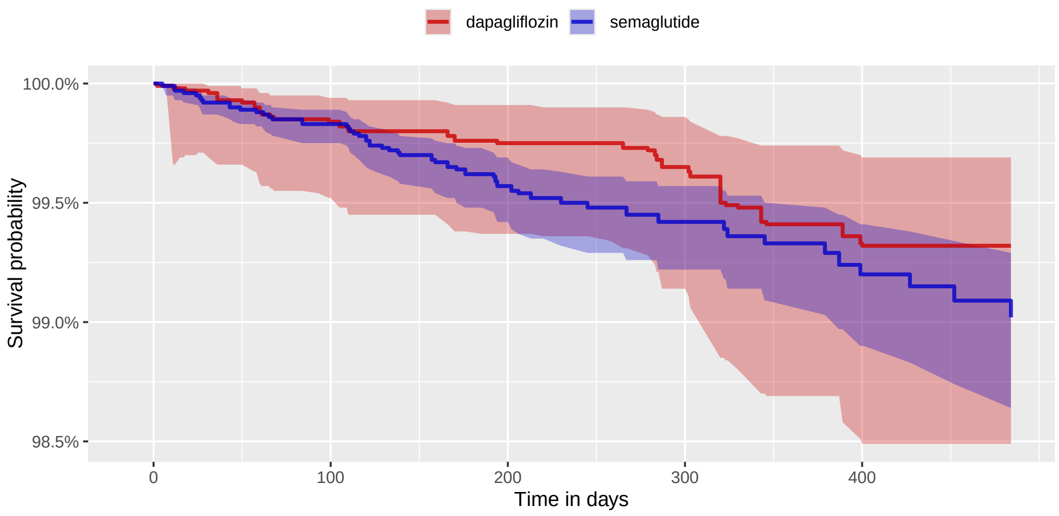

| Number at risk | | | | | |
|---|---|---|---|---|---|
| semaglutide | 20,559 | 9,779 | 5,609 | 3,499 | 2,168 |
| dapagliflozin | 10,356 | 6,177 | 4,274 | 2,956 | 2,120 |

##### 3.4.2 OptumDOD

**Supplementary Table 21:** Coefficients of top-15 most influential covariates of the fitted propensity model. Positive coefficients indicate predictors of the target exposure.

| Coef | Covariate Name (total selected: 477) |
|---|---|
| -3.01 | index year: 2018 |
| 1.02 | index year: 2020 |
| -0.89 | drug_era group during day -365 through 0 days relative to index: sacubitril |
| 0.84 | age group: 45 - 49 |
| -0.64 | CHADS2VASc |
| 0.64 | measurement during day -365 through 0 days relative to index: General health panel This panel must include the following: Comprehensive metabolic panel (80053) Blood count, complete (CBC), automated and automated differential WBC count (85025 or 85027 and 85004) OR Blood count, complete (CBC), automated (85027) and |
| 0.61 | age group: 50 - 54 |
| 0.60 | index year: 2019 |
| 0.53 | age group: 40 - 44 |
| -0.50 | age group: 80 - 84 |
| 0.47 | age group: 55 - 59 |
| 0.47 | drug_era group during day -365 through 0 days relative to index: Other viral vaccines |
| -0.45 | condition_era group during day -365 through 0 days relative to index: Chronic systolic heart failure |
| -0.44 | condition_era group during day -365 through 0 days relative to index: Chronic kidney disease stage 3B |
| -0.42 | age group: 85 - 89 |

**Supplementary Figure 18:** Kaplan Meier curve (top) and number of subjects at risk (bottom) for 3-pt MACE comparing semaglutide vs dapagliflozin new-users in the OptumDOD data source.

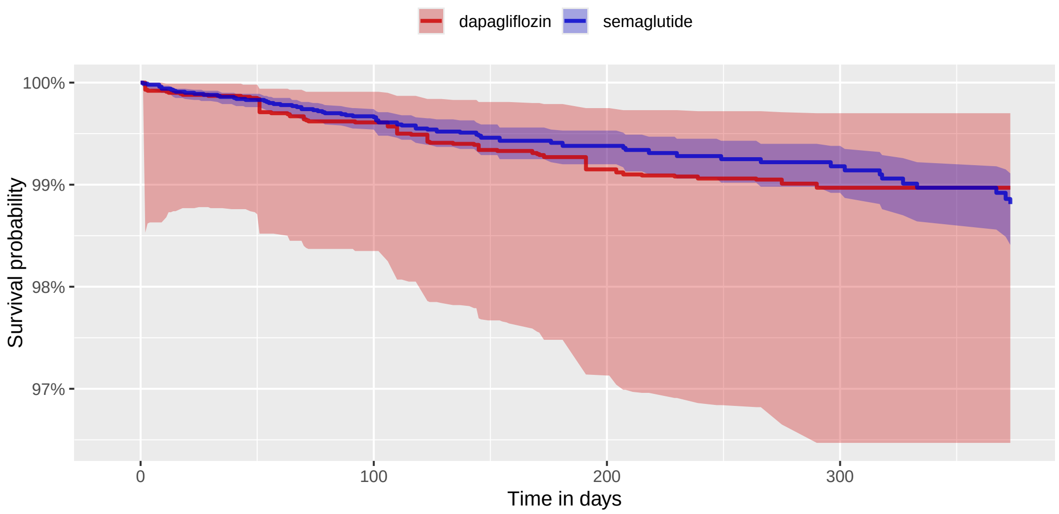

| Number at risk | | | | |
|---|---|---|---|---|
| semaglutide | 18,093 | 8,200 | 4,247 | 2,573 |
| dapagliflozin | 5,664 | 2,901 | 1,520 | 882 |

##### 3.4.3 MDCR

**Supplementary Table 22:** Coefficients of top-15 most influential covariates of the fitted propensity model. Positive coefficients indicate predictors of the target exposure.

| Coef | Covariate Name (total selected: 139) |
|---|---|
| -0.84 | condition_era group during day -30 through 0 days relative to index: Heart failure |
| 0.69 | condition_era group during day -365 through 0 days relative to index: Obesity |
| -0.63 | index year: 2020 |
| 0.51 | measurement within normal range during day -365 through 0 days relative to index: Creatinine [Mass/volume] in Serum or Plasma |
| -0.44 | index year: 2019 |
| -0.32 | gender = MALE |
| 0.31 | condition_era group during day -365 through 0 days relative to index: Major depressive disorder |
| -0.30 | drug_era group during day -365 through 0 days relative to index: valsartan |
| -0.28 | measurement during day -365 through 0 days relative to index: Urinalysis, by dip stick or tablet reagent for bilirubin, glucose, hemoglobin, ketones, leukocytes, nitrite, pH, protein, specific gravity, urobilinogen, any number of these constituents; automated, with microscopy |
| 0.25 | procedure_occurrence during day -365 through 0 days relative to index: Screening mammography, bilateral (2-view study of each breast), including computer-aided detection (CAD) when performed |
| -0.22 | condition_era group during day -30 through 0 days relative to index: Chronic kidney disease stage 3 |
| -0.22 | age group: 75 - 79 |
| -0.22 | condition_era group during day -30 through 0 days relative to index: Kidney disease |
| 0.20 | condition_era group during day -365 through 0 days relative to index: Pain |
| 0.20 | measurement within normal range during day -365 through 0 days relative to index: Carbon dioxide, total [Moles/volume] in Serum or Plasma |

**Supplementary Figure 19:** Kaplan Meier curve (top) and number of subjects at risk (bottom) for 3-pt MACE comparing semaglutide vs dapagliflozin new-users in the MDCR data source.

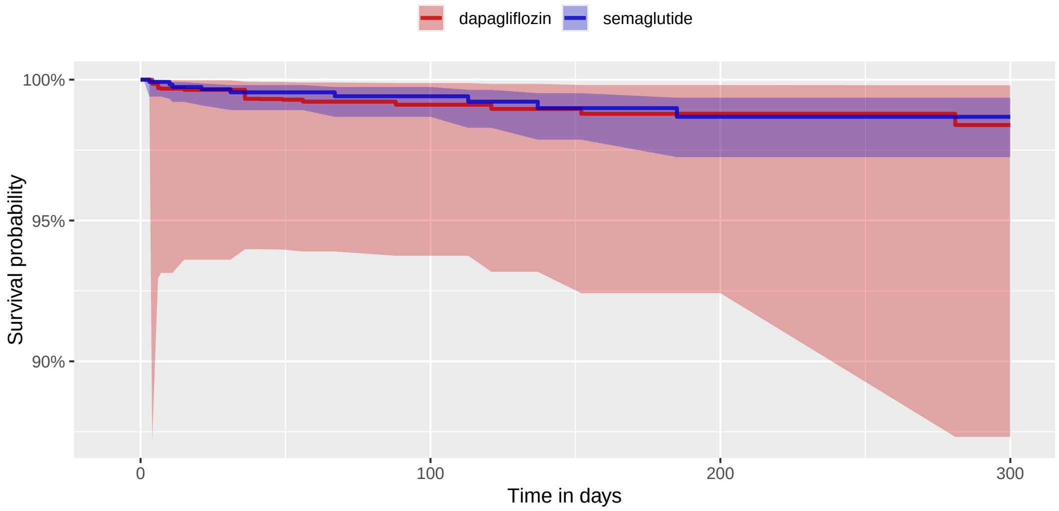

| Number at risk | | | | |
|---|---|---|---|---|
| semaglutide | 1,235 | 537 | 290 | 148 |
| dapagliflozin | 753 | 398 | 220 | 121 |

##### 3.4.4 Open Claims

**Supplementary Table 23:** Coefficients of top-15 most influential covariates of the fitted propensity model. Positive coefficients indicate predictors of the target exposure.

| Coef | Covariate Name (total selected: 1850) |
|---|---|
| -1.99 | index year: 2018 |
| -1.38 | drug_era group (DrugGroupEraLongTerm) during day -365 through 0 days relative to index: sacubitril |
| -0.96 | age group: 80 - 84 |
| -0.94 | age group: 85 - 89 |
| -0.68 | condition_era group (ConditionGroupEraLongTerm) during day -365 through 0 days relative to index: Chronic kidney disease stage 3B |
| -0.62 | age group: 75 - 79 |
| -0.52 | condition_era group (ConditionGroupEraShortTerm) during day -30 through 0 days relative to index: Cardiomyopathy |
| -0.49 | condition_era group (ConditionGroupEraShortTerm) during day -30 through 0 days relative to index: Chronic kidney disease stage 3B |
| -0.48 | condition_era group (ConditionGroupEraShortTerm) during day -30 through 0 days relative to index: Heart failure |
| -0.47 | index year: 2019 |
| -0.46 | observation during day -365 through 0 days relative to index: Body mass index 20-24 - normal |
| -0.44 | condition_era group (ConditionGroupEraLongTerm) during day -365 through 0 days relative to index: Pancreatitis |
| -0.43 | condition_era group (ConditionGroupEraLongTerm) during day -365 through 0 days relative to index: Persistent proteinuria |
| 0.41 | gender = FEMALE |
| -0.40 | age group: 70 - 74 |

**Supplementary Figure 20:** Kaplan Meier curve (top) and number of subjects at risk (bottom) for 3-pt MACE comparing semaglutide vs dapagliflozin new-users in the Open Claims data source.

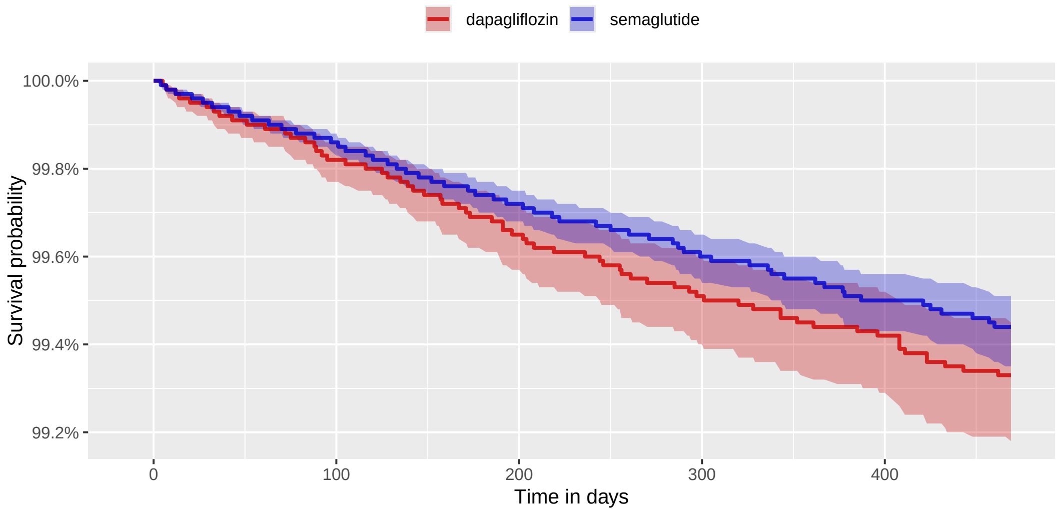

| Number at risk | | | | | |
|---|---|---|---|---|---|
| semaglutide | 167,135 | 79,493 | 46,639 | 28,691 | 18,004 |
| dapagliflozin | 105,079 | 56,924 | 38,232 | 26,102 | 18,122 |

##### 3.4.5 OptumEHR

**Supplementary Table 24:** Coefficients of top-15 most influential covariates of the fitted propensity model. Positive coefficients indicate predictors of the target exposure.

| Coef | Covariate Name (total selected: 737) |
|---|---|
| -5.64 | index year: 2017 |
| -2.13 | index year: 2018 |
| -1.99 | index year: 2016 |
| -1.59 | drug_era group during day -365 through 0 days relative to index: sacubitril |
| -0.78 | age group: 80 - 84 |
| -0.66 | index year: 2019 |
| 0.43 | measurement within normal range during day -365 through 0 days relative to index: Bile.microscopic observation [Identifier] in Tissue by Fouchet stain |
| 0.42 | condition_era group during day -365 through 0 days relative to index: Metabolic syndrome X |
| -0.42 | gender = MALE |
| 0.38 | observation during day -365 through 0 days relative to index: Abnormal weight gain |
| -0.38 | condition_era group during day -365 through 0 days relative to index: Heart failure |
| 0.36 | procedure_occurrence during day -30 through 0 days relative to index: Office or other outpatient visit for the evaluation and management of a new patient, which requires these 3 key components: A comprehensive history; A comprehensive examination; Medical decision making of high complexity. Counseling and/or coordination of |
| -0.35 | age group: 75 - 79 |
| 0.35 | condition_era group during day -30 through 0 days relative to index: Obesity |
| -0.33 | condition_era group during day -30 through 0 days relative to index: Systolic dysfunction |

**Supplementary Figure 21:** Kaplan Meier curve (top) and number of subjects at risk (bottom) for 3-pt MACE comparing semaglutide vs dapagliflozin new-users in the OptumEHR data source.

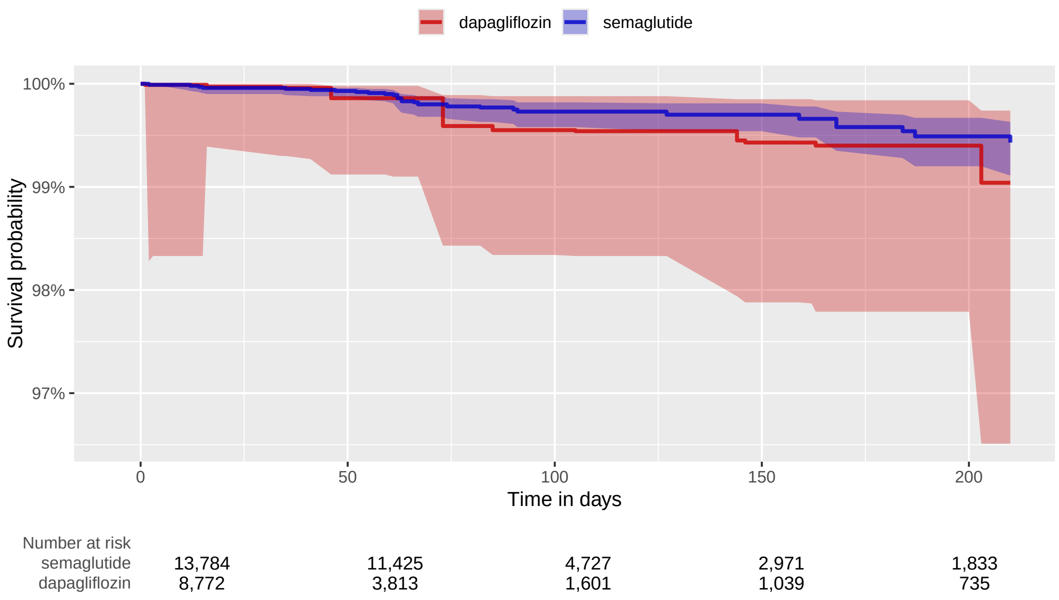

#### 3.5 semaglutide vs empagliflozin

##### 3.5.1 CCAE

**Supplementary Table 25:** Coefficients of top-15 most influential covariates of the fitted propensity model. Positive coefficients indicate predictors of the target exposure.

| Coef | Covariate Name (total selected: 627) |
|---|---|
| -1.57 | index year: 2018 |
| -1.03 | index year: 2019 |
| -0.53 | condition_era group during day -30 through 0 days relative to index: Heart failure |
| -0.47 | index year: 2020 |
| -0.47 | gender = MALE |
| -0.46 | drug_era group during day -30 through 0 days relative to index: sacubitril |
| 0.45 | index year: 2022 |
| 0.37 | condition_era group during day -30 through 0 days relative to index: Obesity |
| 0.37 | observation during day -365 through 0 days relative to index: Abnormal weight gain |
| -0.35 | condition_era group during day -365 through 0 days relative to index: Systolic dysfunction |
| 0.34 | observation during day -365 through 0 days relative to index: Body mass index 40+ - severely obese |
| -0.32 | index month: 1 |
| 0.31 | measurement during day -365 through 0 days relative to index: Unlisted chemistry procedure |
| 0.30 | age group: 30 - 34 |
| 0.29 | condition_era group during day -30 through 0 days relative to index: Metabolic syndrome X |

**Supplementary Figure 22:** Kaplan Meier curve (top) and number of subjects at risk (bottom) for 3-pt MACE comparing semaglutide vs empagliflozin new-users in the CCAE data source.

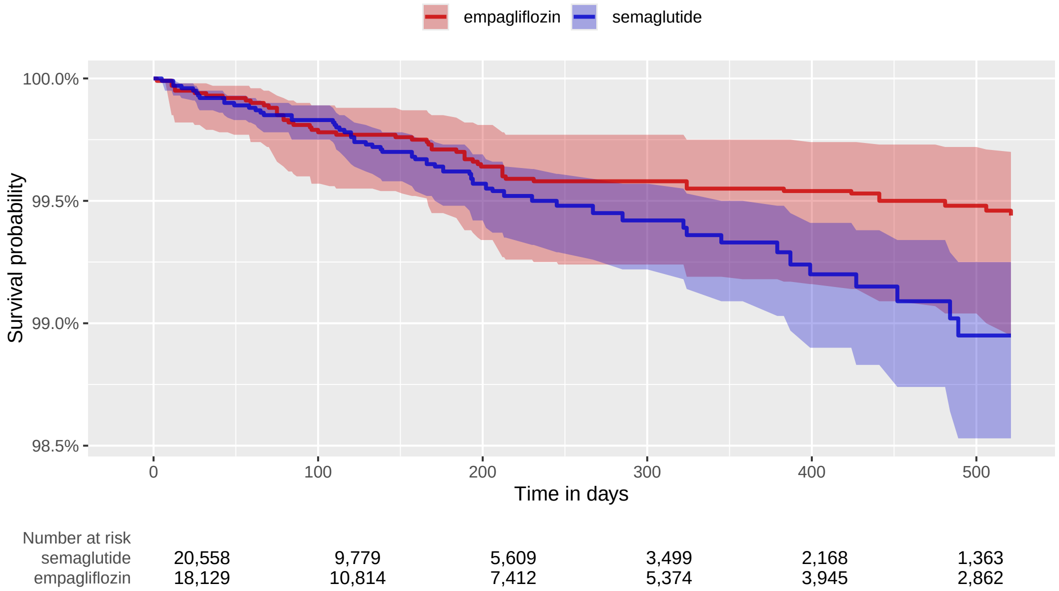

##### 3.5.2 OptumDOD

**Supplementary Table 26:** Coefficients of top-15 most influential covariates of the fitted propensity model. Positive coefficients indicate predictors of the target exposure.

| Coef | Covariate Name (total selected: 659) |
|---|---|
| -3.69 | index year: 2018 |
| -1.35 | index year: 2019 |
| -0.82 | index year: 2020 |
| -0.61 | age group: 85 - 89 |
| -0.52 | age group: 80 - 84 |
| 0.43 | measurement during day -365 through 0 days relative to index: Insulin, total measurement |
| 0.41 | gender = FEMALE |
| 0.39 | condition_era group during day -365 through 0 days relative to index: Metabolic syndrome X |
| -0.38 | age group: 75 - 79 |
| 0.38 | observation during day -365 through 0 days relative to index: Body mass index 40+ - severely obese |
| 0.37 | measurement within normal range during day -365 through 0 days relative to index: Hemoglobin A1c/Hemoglobin.total in Blood |
| -0.35 | index year: 2021 |
| 0.33 | condition_era group during day -30 through 0 days relative to index: Obesity |
| -0.33 | condition_era group during day -30 through 0 days relative to index: Heart failure |
| -0.32 | index month: 1 |

**Supplementary Figure 23:** Kaplan Meier curve (top) and number of subjects at risk (bottom) for 3-pt MACE comparing semaglutide vs empagliflozin new-users in the OptumDOD data source.

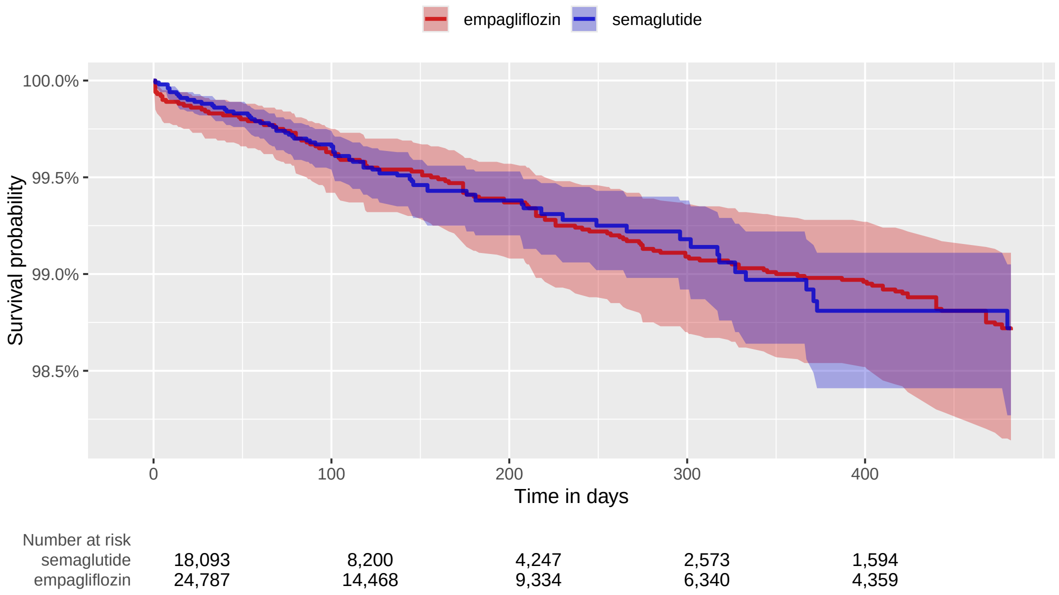

##### 3.5.3 MDCR

**Supplementary Table 27:** Coefficients of top-15 most influential covariates of the fitted propensity model. Positive coefficients indicate predictors of the target exposure.

| Coef | Covariate Name (total selected: 180) |
|---|---|
| -0.65 | index year: 2019 |
| 0.48 | condition_era group during day -365 through 0 days relative to index: Obesity |
| 0.44 | condition_era group during day -365 through 0 days relative to index: Metabolic syndrome X |
| -0.36 | device_exposure during day -365 through 0 days relative to index: Blood glucose test or reagent strips for home blood glucose monitor, per 50 strips |
| 0.36 | gender = FEMALE |
| -0.30 | index year: 2020 |
| -0.29 | condition_era group during day -30 through 0 days relative to index: Heart disease |
| -0.29 | condition_era group during day -30 through 0 days relative to index: Heart failure |
| -0.29 | condition_era group during day -365 through 0 days relative to index: Heart failure |
| 0.27 | drug_era group during day 0 through 0 days relative to index: CARDIOVASCULAR SYSTEM |
| -0.26 | condition_era group during day -365 through 0 days relative to index: Pure hypercholesterolemia |
| 0.26 | condition_era group during day -30 through 0 days relative to index: Obesity |
| -0.26 | index year: 2021 |
| -0.25 | measurement during day -30 through 0 days relative to index: Hemoglobin A1c measurement |
| -0.25 | age group: 75 - 79 |

**Supplementary Figure 24:** Kaplan Meier curve (top) and number of subjects at risk (bottom) for 3-pt MACE comparing semaglutide vs empagliflozin new-users in the MDCR data source.

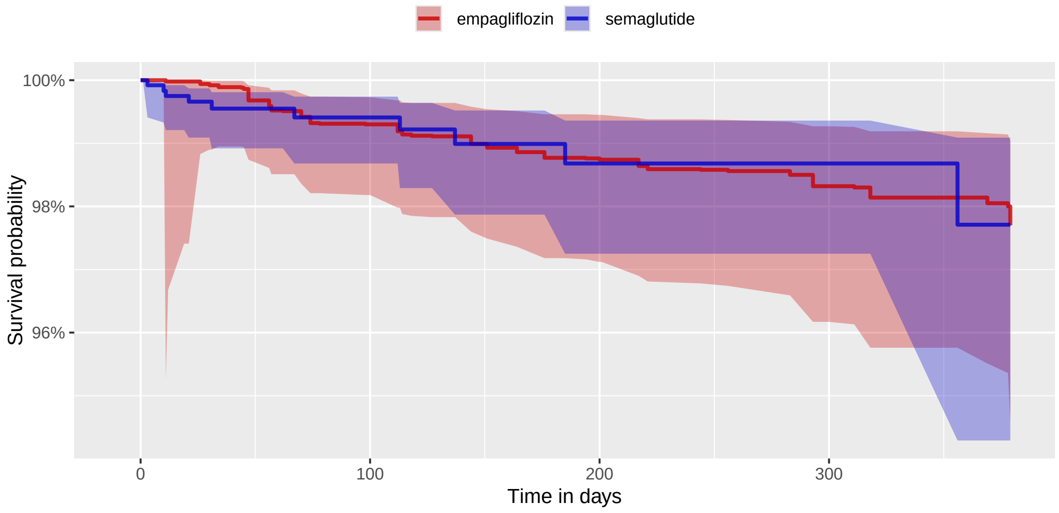

| Number at risk | | | | |
|---|---|---|---|---|
| semaglutide | 1,247 | 537 | 290 | 148 |
| empagliflozin | 2,741 | 1,321 | 790 | 499 |

##### 3.5.4 Open Claims

**Supplementary Table 28:** Coefficients of top-15 most influential covariates of the fitted propensity model. Positive coefficients indicate predictors of the target exposure.

| Coef | Covariate Name (total selected: 1588) |
|---|---|
| -1.83 | index year: 2018 |
| -1.02 | age group: 80 - 84 |
| -0.90 | index year: 2019 |
| -0.90 | age group: 85 - 89 |
| -0.76 | drug_era group (DrugGroupEraLongTerm) during day -365 through 0 days relative to index: sacubitril |
| -0.73 | age group: 75 - 79 |
| -0.57 | observation during day -30 through 0 days relative to index: Clinic visit/encounter, all-inclusive |
| -0.54 | age group: 70 - 74 |
| -0.51 | condition_era group (ConditionGroupEraLongTerm) during day -365 through 0 days relative to index: Pancreatitis |
| 0.50 | condition_era group (ConditionGroupEraLongTerm) during day -365 through 0 days relative to index: Metabolic syndrome X |
| -0.49 | observation during day -365 through 0 days relative to index: Clinic visit/encounter, all-inclusive |
| 0.45 | drug_era group (DrugGroupEraLongTerm) during day -365 through 0 days relative to index: phentermine |
| -0.44 | gender = MALE |
| -0.44 | index year: 2020 |
| 0.41 | drug_era group (DrugGroupEraLongTerm) during day -365 through 0 days relative to index: ANTIOBESITY PREPARATIONS, EXCL. DIET PRODUCTS |

**Supplementary Figure 25:** Kaplan Meier curve (top) and number of subjects at risk (bottom) for 3-pt MACE comparing semaglutide vs empagliflozin new-users in the Open Claims data source.

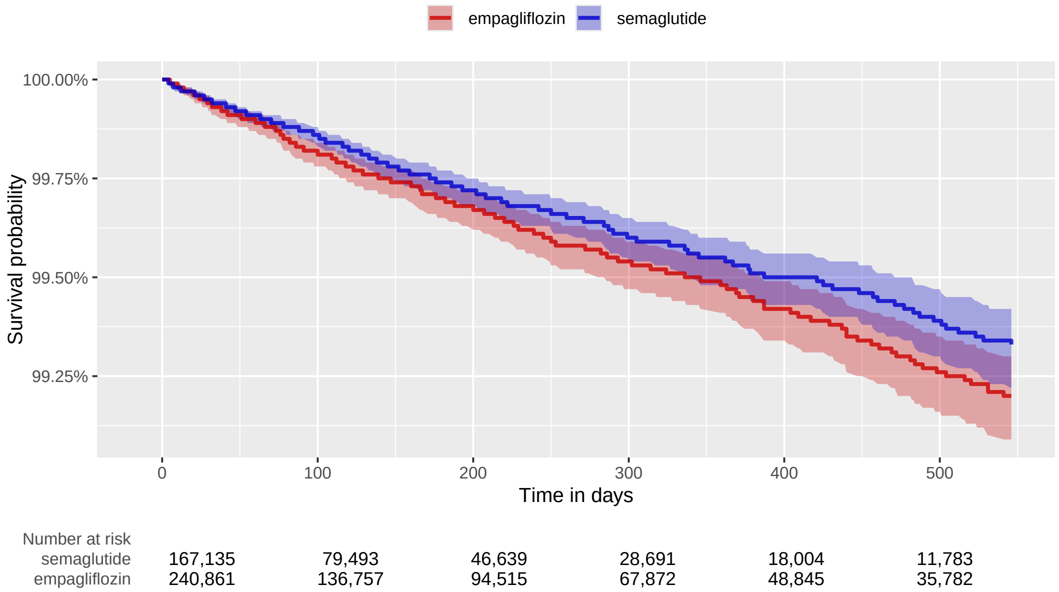

##### 3.5.5 CUIMC

**Supplementary Table 29:** Coefficients of top-15 most influential covariates of the fitted propensity model. Positive coefficients indicate predictors of the target exposure.

| Coef | Covariate Name (total selected: 210) |
|---|---|
| 0.98 | condition_era group during day -30 through 0 days relative to index: Obesity |
| -0.97 | index year: 2020 |
| -0.75 | index year: 2019 |
| -0.52 | condition_era group during day -30 through 0 days relative to index: Congestive heart failure |
| 0.48 | condition_era group during day -365 through 0 days relative to index: Morbid obesity |
| 0.47 | condition_era group during day -365 through 0 days relative to index: Snoring |
| -0.44 | condition_era group during day -30 through 0 days relative to index: Myocardial disease |
| -0.44 | condition_era group during day -365 through 0 days relative to index: Atherosclerosis of artery |
| -0.38 | age group: 80 - 84 |
| -0.37 | condition_era group during day -365 through 0 days relative to index: Congestive heart failure |
| -0.37 | age group: 75 - 79 |
| -0.37 | gender = MALE |
| 0.37 | measurement within normal range during day -365 through 0 days relative to index: Glucose [Mass/volume] in Serum or Plasma |
| -0.35 | measurement above normal range during day -365 through 0 days relative to index: Glucose [Mass/volume] in Serum or Plasma |
| -0.35 | measurement during day -30 through 0 days relative to index: Hemoglobin A1c/Hemoglobin.total in Blood |

**Supplementary Figure 26:** Kaplan Meier curve (top) and number of subjects at risk (bottom) for 3-pt MACE comparing semaglutide vs empagliflozin new-users in the CUIMC data source.

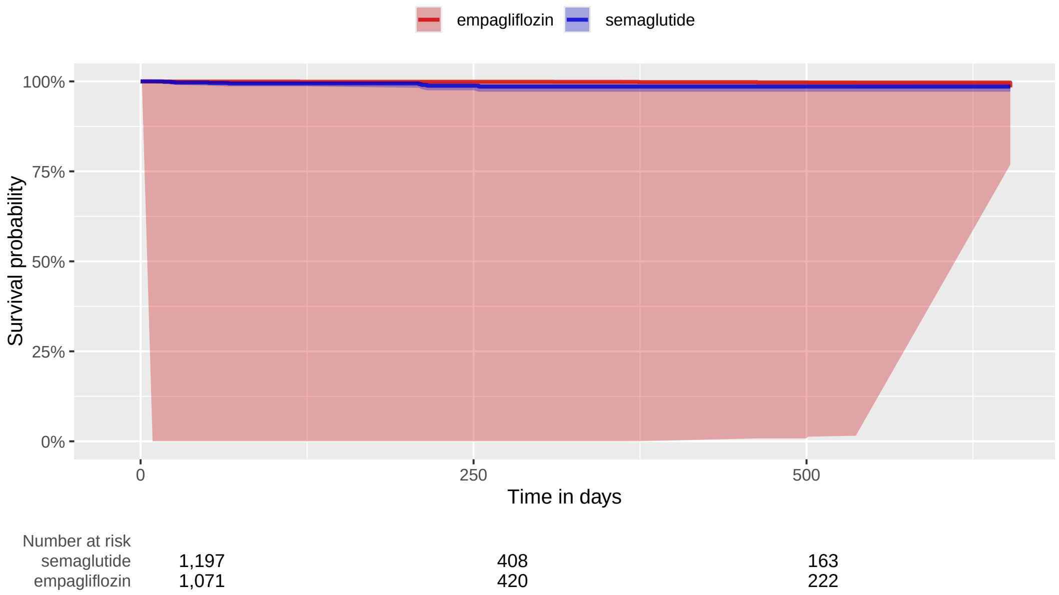

##### 3.5.6 OptumEHR

**Supplementary Table 30:** Coefficients of top-15 most influential covariates of the fitted propensity model. Positive coefficients indicate predictors of the target exposure.

| Coef | Covariate Name (total selected: 939) |
|---|---|
| -5.16 | index year: 2017 |
| -1.86 | index year: 2018 |
| -1.00 | drug_era group during day -365 through 0 days relative to index: sacubitril |
| -0.94 | index year: 2016 |
| -0.91 | age group: 80 - 84 |
| -0.86 | index year: 2019 |
| -0.58 | age group: 75 - 79 |
| -0.56 | age group: 85 - 89 |
| 0.51 | observation during day -365 through 0 days relative to index: Abnormal weight gain |
| 0.48 | gender = FEMALE |
| 0.48 | condition_era group during day -365 through 0 days relative to index: Metabolic syndrome X |
| -0.48 | condition_era group during day -30 through 0 days relative to index: Cardiomyopathy |
| -0.45 | index year: 2020 |
| 0.44 | age group: 25 - 29 |
| 0.37 | age group: 35 - 39 |

**Supplementary Figure 27:** Kaplan Meier curve (top) and number of subjects at risk (bottom) for 3-pt MACE comparing semaglutide vs empagliflozin new-users in the OptumEHR data source.

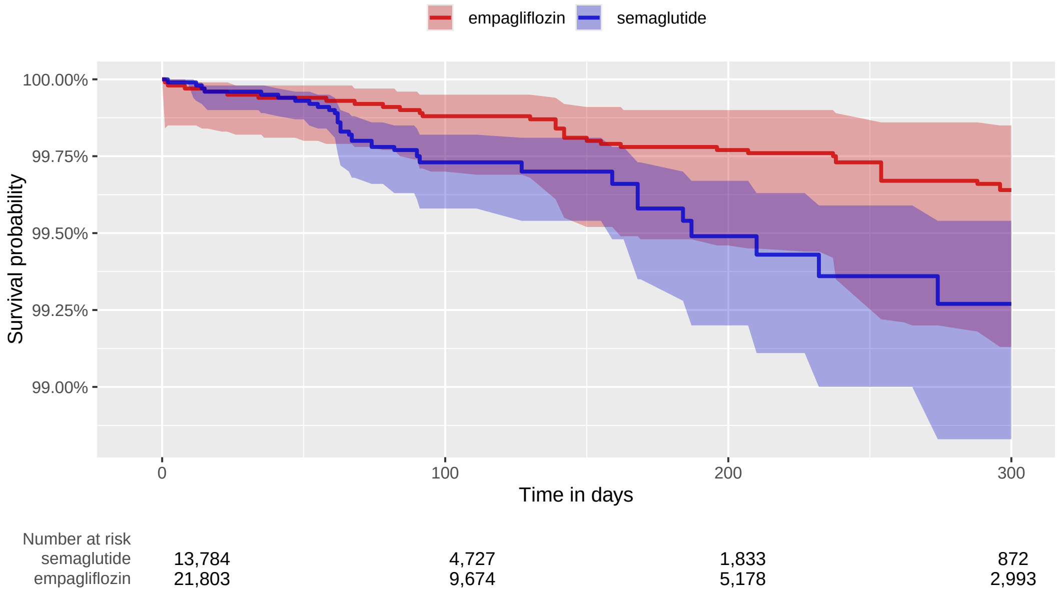

### 3.5.7 VA

**Supplementary Table 31:** Coefficients of top-15 most influential covariates of the fitted propensity model. Positive coefficients indicate predictors of the target exposure.

| Coef | Covariate Name (total selected: 1017) |
|---|---|
| -2.07 | device_exposure during day -30 through 0 days relative to index: TABLET CUTTER |
| 1.73 | device_exposure during day -365 through 0 days relative to index: NEEDLE,PEN 31G,8MM |
| 1.42 | device_exposure during day -365 through 0 days relative to index: NEEDLE,PEN 32G,4MM |
| 1.13 | condition_era group during day -30 through 0 days relative to index: Obesity |
| 1.06 | observation during day -30 through 0 days relative to index: Qualified nonphysician health care professional online digital assessment and management, for an established patient, for up to 7 days, cumulative time during the 7 days; 21 or more minutes |
| -0.94 | drug_era group during day -365 through 0 days relative to index: sacubitril |
| 0.93 | procedure_occurrence during day -30 through 0 days relative to index: Medication therapy management service(s) provided by a pharmacist, individual, face-to-face with patient, with assessment and intervention if provided; initial 15 minutes, new patient |
| 0.88 | device_exposure during day -365 through 0 days relative to index: NEEDLE,PEN 31G,5MM |
| 0.85 | device_exposure during day -365 through 0 days relative to index: SHARPS DISPOSAL CONTAINER 1 QUART SIZE |
| 0.82 | observation during day -30 through 0 days relative to index: Qualified nonphysician health care professional online digital assessment and management, for an established patient, for up to 7 days, cumulative time during the 7 days; 5-10 minutes |
| -0.78 | condition_era group during day -30 through 0 days relative to index: Heart failure |
| -0.71 | measurement during day -365 through 0 days relative to index: Platelet count, blood, automated |
| 0.69 | device_exposure during day -30 through 0 days relative to index: SHARPS DISPOSAL CONTAINER 1 GALLON SIZE |
| 0.64 | measurement within normal range during day -365 through 0 days relative to index: Basophils/100 leukocytes in Blood by Manual count |
| 0.62 | observation during day -365 through 0 days relative to index: Long-term current use of insulin |

#### 3.6 dapagliflozin vs empagliflozin

##### 3.6.1 CCAE

**Supplementary Table 32:** Coefficients of top-15 most influential covariates of the fitted propensity model. Positive coefficients indicate predictors of the target exposure.

| Coef | Covariate Name (total selected: 366) |
|---|---|
| 1.11 | index year: 2014 |
| 0.92 | index year: 2016 |
| 0.90 | index year: 2015 |
| 0.44 | index year: 2018 |
| -0.37 | index year: 2019 |
| 0.32 | condition_era group during day -365 through 0 days relative to index: Chronic kidney disease stage 3A |
| 0.18 | condition_era group during day -365 through 0 days relative to index: Chronic disease of genitourinary system |
| 0.17 | condition_era group during day -365 through 0 days relative to index: Hypertensive heart disease |
| -0.15 | measurement during day -365 through 0 days relative to index: Electrolyte panel This panel must include the following: Carbon dioxide (bicarbonate) (82374) Chloride (82435) Potassium (84132) Sodium (84295) |
| -0.15 | measurement during day -365 through 0 days relative to index: Albumin; urine (eg, microalbumin), quantitative |
| 0.13 | observation during day -365 through 0 days relative to index: Days per week of moderate to vigorous physical activity |
| -0.13 | index year: 2020 |
| 0.13 | drug_era group during day -365 through 0 days relative to index: sacubitril |
| 0.12 | condition_era group during day -30 through 0 days relative to index: Cardiomyopathy |
| 0.12 | index year: 2017 |

**Supplementary Figure 28:** Kaplan Meier curve (top) and number of subjects at risk (bottom) for 3-pt MACE comparing dapagliflozin vs empagliflozin new-users in the CCAE data source.

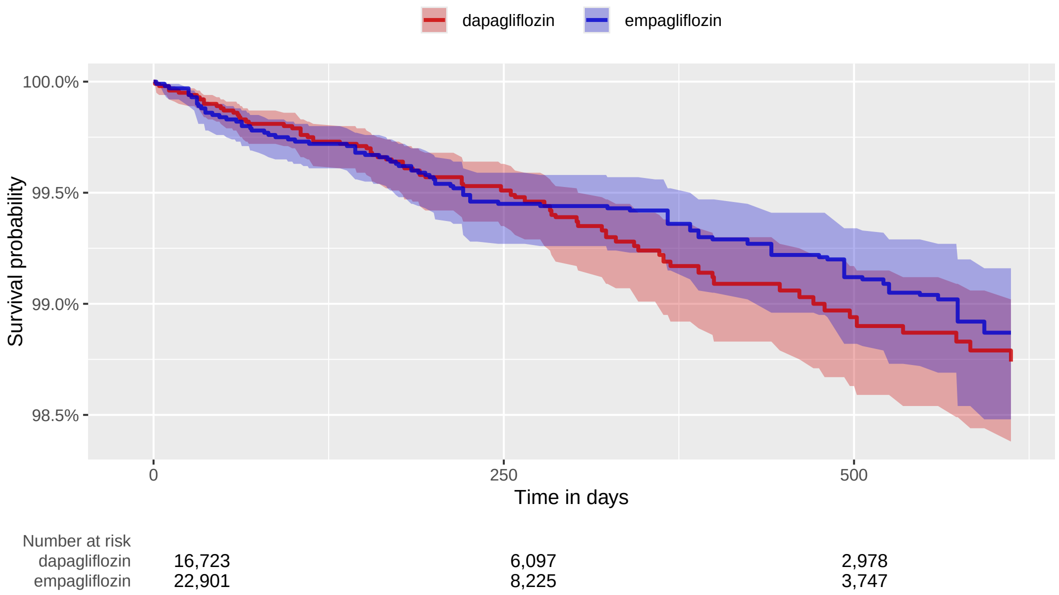

##### 3.6.2 GermanyDA

**Supplementary Table 33:** Coefficients of top-15 most influential covariates of the fitted propensity model. Positive coefficients indicate predictors of the target exposure.

| Coef | Covariate Name (total selected: 150) |
|---|---|
| -0.45 | index year: 2019.000000 |
| -0.45 | index year: 2018.000000 |
| 0.41 | index year: 2015.000000 |
| -0.35 | Diabetes Comorbidity Severity Index (DCSI) |
| 0.29 | measurement during day -365 through 0 days relative to index: Leukocytes [#/volume] in Body fluid |
| 0.27 | drug_era group during day 0 through 0 days relative to index: metformin |
| -0.22 | index year: 2017.000000 |
| -0.21 | age group: 75 - 79 |
| -0.19 | drug_era group during day -365 through 0 days relative to index: ALL OTHER THERAPEUTIC PRODUCTS |
| 0.19 | observation during day -365 through 0 days relative to index: Obesity monitoring |
| -0.19 | condition_era group during day -365 through 0 days relative to index: Heart disease |
| -0.17 | condition_era group during day -365 through 0 days relative to index: Peripheral circulatory disorder due to type 2 diabetes mellitus |
| -0.15 | condition_era group during day -365 through 0 days relative to index: Structural disorder of heart |
| 0.14 | measurement during day -30 through 0 days relative to index: Hemoglobin A1c/Hemoglobin.total in Blood |
| -0.14 | index year: 2020.000000 |

**Supplementary Figure 29:** Kaplan Meier curve (top) and number of subjects at risk (bottom) for 3-pt MACE comparing dapagliflozin vs empagliflozin new-users in the GermanyDA data source.

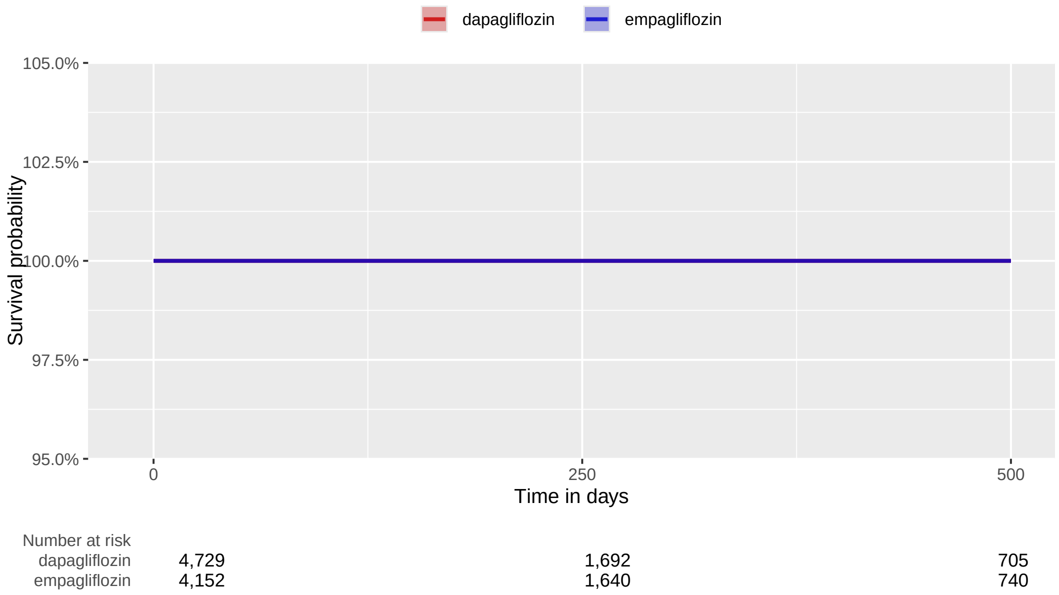

##### 3.6.3 OptumDOD

**Supplementary Table 34:** Coefficients of top-15 most influential covariates of the fitted propensity model. Positive coefficients indicate predictors of the target exposure.

| Coef | Covariate Name (total selected: 472) |
|---|---|
| 2.63 | index year: 2014 |
| -2.06 | index year: 2019 |
| -1.77 | index year: 2020 |
| 1.20 | index year: 2015 |
| -1.00 | index year: 2018 |
| -0.51 | index year: 2017 |
| -0.48 | drug_era group during day -365 through 0 days relative to index: Other viral vaccines |
| 0.41 | drug_era group during day -365 through 0 days relative to index: sacubitril |
| 0.32 | condition_era group during day -365 through 0 days relative to index: Chronic kidney disease stage 3B |
| 0.30 | condition_era group during day -30 through 0 days relative to index: Renal impairment |
| -0.24 | procedure_occurrence during day -365 through 0 days relative to index: Primary immunisation - second dose |
| 0.24 | measurement during day -365 through 0 days relative to index: Body height |
| -0.24 | drug_era group during day -365 through 0 days relative to index: aspirin |
| -0.23 | measurement during day -365 through 0 days relative to index: General health panel This panel must include the following: Comprehensive metabolic panel (80053) Blood count, complete (CBC), automated and automated differential WBC count (85025 or 85027 and 85004) OR Blood count, complete (CBC), automated (85027) and |
| 0.21 | procedure_occurrence during day -365 through 0 days relative to index: Advance care planning including the explanation and discussion of advance directives such as standard forms (with completion of such forms, when performed), by the physician or other qualified health care professional; first 30 minutes, face-to-face with |

**Supplementary Figure 30:** Kaplan Meier curve (top) and number of subjects at risk (bottom) for 3-pt MACE comparing dapagliflozin vs empagliflozin new-users in the OptumDOD data source.

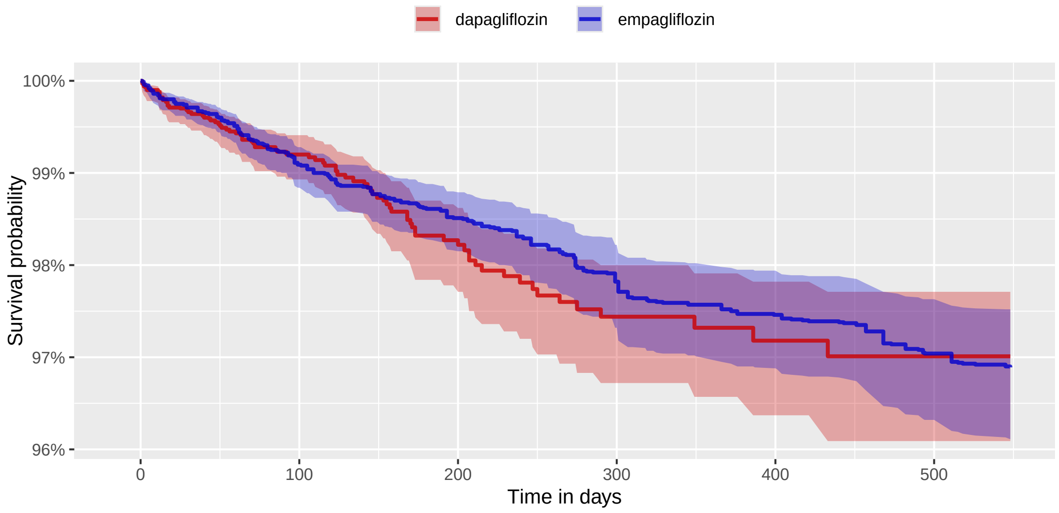

| Number at risk | | | | | | |
|---|---|---|---|---|---|---|
| dapagliflozin | 6,757 | 3,444 | 1,853 | 1,102 | 641 | 359 |
| empagliflozin | 28,219 | 16,574 | 10,876 | 7,532 | 5,270 | 3,862 |

##### 3.6.4 MDCR

**Supplementary Table 35:** Coefficients of top-15 most influential covariates of the fitted propensity model. Positive coefficients indicate predictors of the target exposure.

| Coef | Covariate Name (total selected: 119) |
|---|---|
| -0.94 | measurement within normal range during day -365 through 0 days relative to index: Potassium [Moles/volume] in Serum or Plasma |
| 0.71 | index year: 2016 |
| 0.70 | index year: 2015 |
| -0.36 | condition_era group during day -365 through 0 days relative to index: Atherosclerosis of coronary artery without angina pectoris |
| -0.22 | index year: 2021 |
| 0.19 | measurement during day -365 through 0 days relative to index: Urinalysis, by dip stick or tablet reagent for bilirubin, glucose, hemoglobin, ketones, leukocytes, nitrite, pH, protein, specific gravity, urobilinogen, any number of these constituents; automated, with microscopy |
| 0.19 | index year: 2020 |
| -0.19 | measurement during day -365 through 0 days relative to index: Albumin measurement, urine, quantitative |
| 0.17 | condition_era group during day -365 through 0 days relative to index: Lesion of joint |
| 0.17 | condition_era group during day -365 through 0 days relative to index: Benign hypertension |
| 0.16 | drug_era group during day 0 through 0 days relative to index: HMG CoA reductase inhibitors, other combinations |
| -0.15 | observation during day -365 through 0 days relative to index: Long-term current use of drug therapy |
| -0.15 | measurement within normal range during day -365 through 0 days relative to index: Creatinine [Mass/volume] in Serum or Plasma |
| 0.15 | drug_era group during day 0 through 0 days relative to index: ALIMENTARY TRACT AND METABOLISM |
| 0.14 | observation during day -365 through 0 days relative to index: Current tobacco non-user (CAD, CAP, COPD, PV) (DM) (IBD) |

**Supplementary Figure 31:** Kaplan Meier curve (top) and number of subjects at risk (bottom) for 3-pt MACE comparing dapagliflozin vs empagliflozin new-users in the MDCR data source.

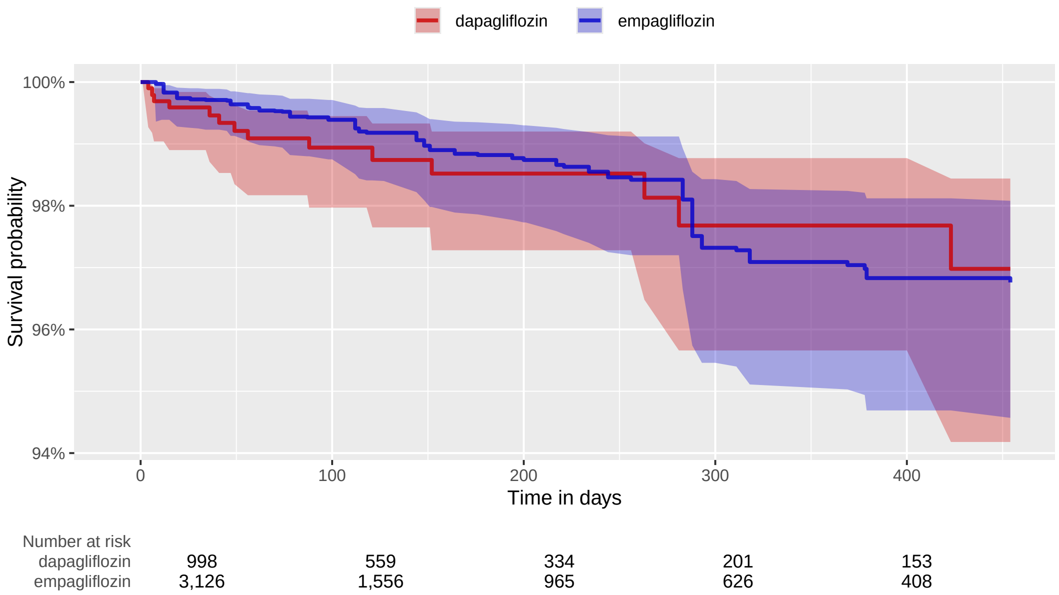

##### 3.6.5 MDCD

**Supplementary Table 36:** Coefficients of top-15 most influential covariates of the fitted propensity model. Positive coefficients indicate predictors of the target exposure.

| Coef | Covariate Name (total selected: 95) |
|---|---|
| -0.45 | index year: 2020 |
| 0.44 | observation during day -365 through 0 days relative to index: Free-Standing Clinic - Other Freestanding Clinic |
| 0.29 | measurement during day -365 through 0 days relative to index: General health panel This panel must include the following: Comprehensive metabolic panel (80053) Blood count, complete (CBC), automated and automated differential WBC count (85025 or 85027 and 85004) OR Blood count, complete (CBC), automated (85027) and |
| 0.22 | condition_era group during day -30 through 0 days relative to index: Systolic dysfunction |
| 0.16 | drug_era group during day -365 through 0 days relative to index: Other dermatologicals |
| -0.16 | race = White |
| -0.15 | drug_era group during day 0 through 0 days relative to index: atorvastatin |
| -0.14 | condition_era group during day -365 through 0 days relative to index: Musculoskeletal and connective tissue disorder |
| 0.14 | observation during day -30 through 0 days relative to index: Free-Standing Clinic - Rural Health-Clinic |
| -0.12 | drug_era group during day -365 through 0 days relative to index: isopropyl alcohol |
| 0.12 | condition_era group during day -30 through 0 days relative to index: Heart failure |
| 0.11 | drug_era group during day -30 through 0 days relative to index: NERVOUS SYSTEM |
| 0.11 | device_exposure during day -365 through 0 days relative to index: Blood glucose test or reagent strips for home blood glucose monitor, per 50 strips |
| 0.10 | drug_era group during day -365 through 0 days relative to index: Angiotensin II receptor blockers (ARBs), other combinations |
| 0.10 | drug_era group during day -30 through 0 days relative to index: Other centrally acting agents |

**Supplementary Figure 32:** Kaplan Meier curve (top) and number of subjects at risk (bottom) for 3-pt MACE comparing dapagliflozin vs empagliflozin new-users in the MDCD data source.

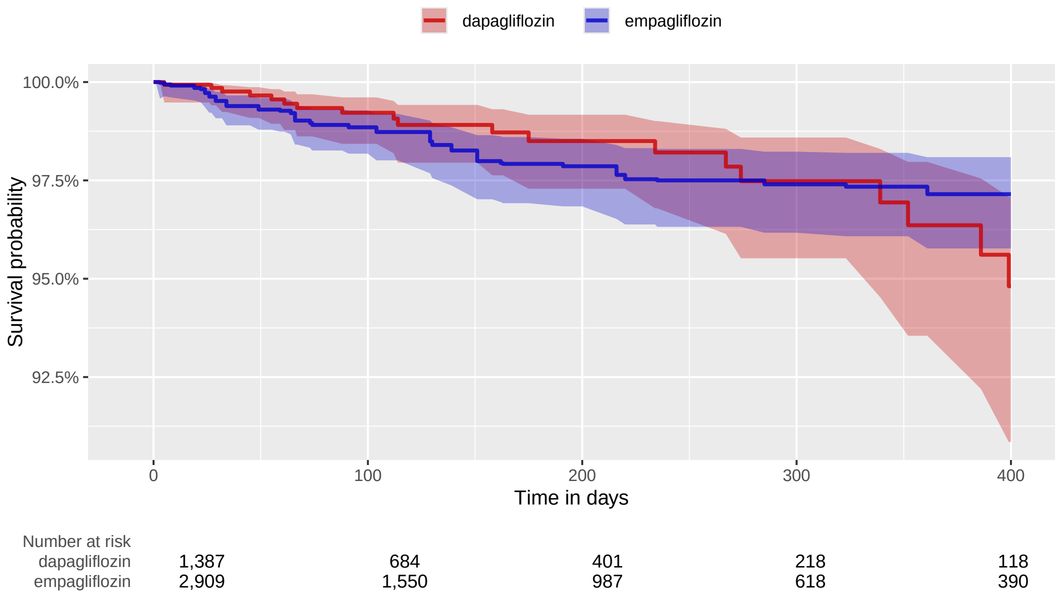

##### 3.6.6 Open Claims

**Supplementary Table 37:** Coefficients of top-15 most influential covariates of the fitted propensity model. Positive coefficients indicate predictors of the target exposure.

| Coef | Covariate Name (total selected: 775) |
|---|---|
| 1.74 | index year: 2014 |
| 1.50 | index year: 2015 |
| 0.97 | index year: 2016 |
| 0.59 | drug_era group (DrugGroupEraLongTerm) during day -365 through 0 days relative to index: sacubitril |
| -0.51 | index year: 2019 |
| 0.39 | condition_era group (ConditionGroupEraLongTerm) during day -365 through 0 days relative to index: Chronic kidney disease stage 3B |
| -0.31 | observation during day -365 through 0 days relative to index: Clinic visit/encounter, all-inclusive |
| 0.26 | index year: 2017 |
| 0.26 | condition_era group (ConditionGroupEraLongTerm) during day -365 through 0 days relative to index: Chronic kidney disease stage 3A |
| -0.22 | index year: 2020 |
| -0.19 | age group: 70 - 74 |
| -0.19 | device_exposure during day -365 through 0 days relative to index: ONE TOUCH VERIO TEST STRIP |
| 0.18 | drug_era group (DrugGroupEraLongTerm) during day -365 through 0 days relative to index: icosapent ethyl |
| -0.17 | device_exposure during day -365 through 0 days relative to index: ACCU-CHEK AVIVA PLUS TEST STRP |
| -0.16 | age group: 75 - 79 |

**Supplementary Figure 33:** Kaplan Meier curve (top) and number of subjects at risk (bottom) for 3-pt MACE comparing dapagliflozin vs empagliflozin new-users in the Open Claims data source.

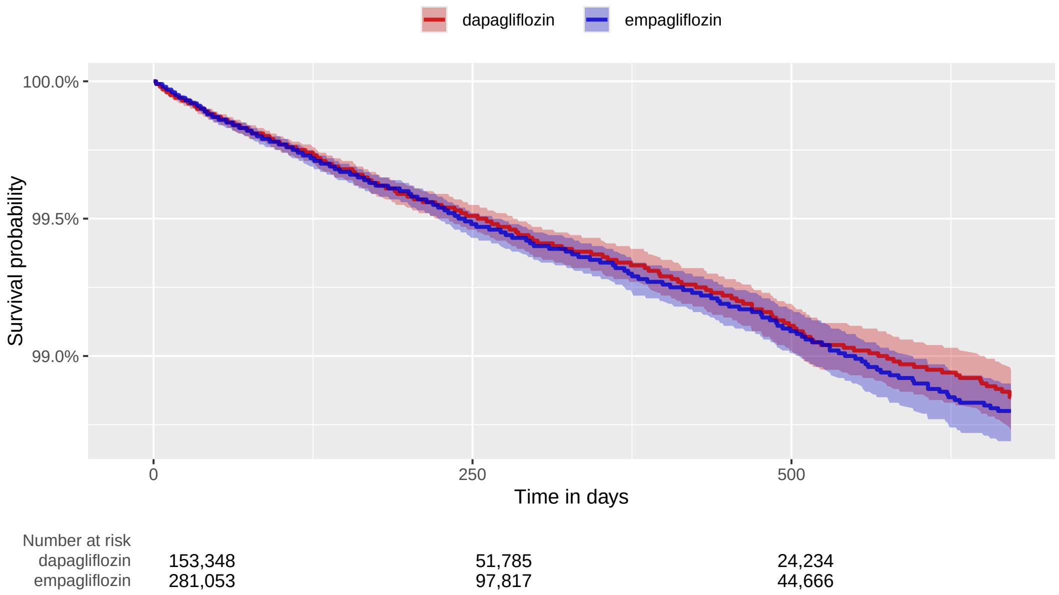

##### 3.6.7 IMRD

**Supplementary Table 38:** Coefficients of top-15 most influential covariates of the fitted propensity model. Positive coefficients indicate predictors of the target exposure.

| Coef | Covariate Name (total selected: 316) |
|---|---|
| 2.53 | index year: 2015 |
| -1.51 | observation during day -365 through 0 days relative to index: Provision of diabetes clinical summary |
| -1.45 | observation during day -365 through 0 days relative to index: Red blood cell color |
| 1.34 | index year: 2016 |
| 1.27 | observation during day -365 through 0 days relative to index: Diabetic leaflet given |
| -1.16 | observation during day -365 through 0 days relative to index: Blood sample sent to biochemistry laboratory |
| -0.87 | observation during day -365 through 0 days relative to index: Self-help advice leaflet given |
| 0.71 | observation during day -365 through 0 days relative to index: Total alkaline phosphatase level |
| -0.70 | observation during day -30 through 0 days relative to index: Serum GGT (gamma-glutamyl transferase) level |
| 0.67 | observation during day -365 through 0 days relative to index: Diabetic on oral treatment |
| -0.66 | observation during day -365 through 0 days relative to index: Chronic disease monitoring |
| 0.63 | observation during day -365 through 0 days relative to index: Seen by ophthalmologist |
| 0.58 | observation during day -365 through 0 days relative to index: Percentage basophils |
| -0.58 | observation during day -365 through 0 days relative to index: Promyelocyte count |
| 0.57 | observation during day -365 through 0 days relative to index: Influenza vaccination consent given |

**Supplementary Figure 34:** Kaplan Meier curve (top) and number of subjects at risk (bottom) for 3-pt MACE comparing dapagliflozin vs empagliflozin new-users in the IMRD data source.

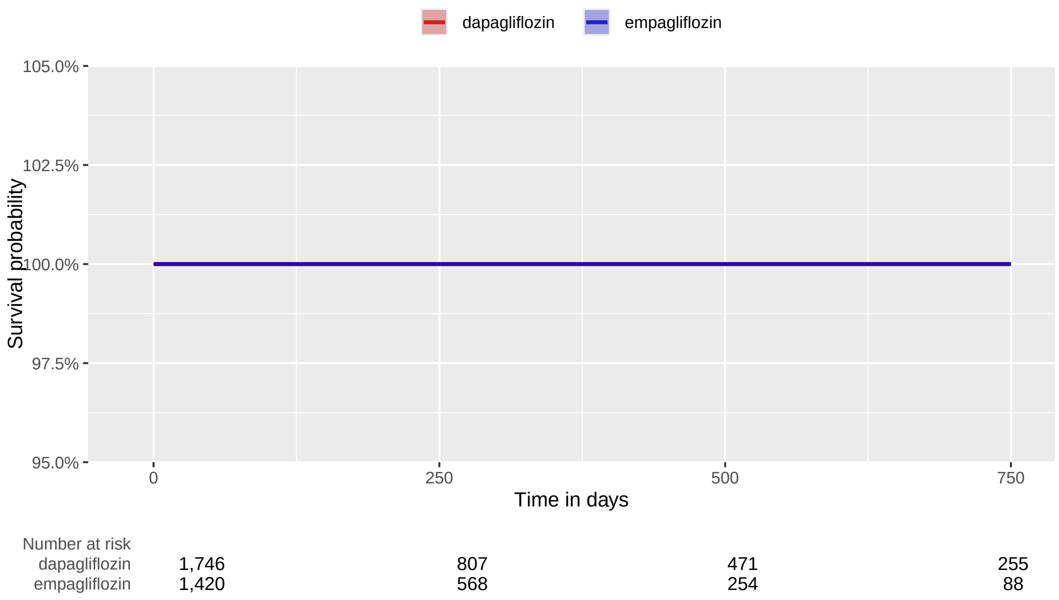

##### 3.6.8 OptumEHR

**Supplementary Table 39:** Coefficients of top-15 most influential covariates of the fitted propensity model. Positive coefficients indicate predictors of the target exposure.

| Coef | Covariate Name (total selected: 585) |
|---|---|
| 1.77 | index year: 2015 |
| 1.66 | index year: 2014 |
| 1.17 | index year: 2016 |
| -0.39 | observation during day -365 through 0 days relative to index: Most recent LDL-C less than 100 mg/dL (CAD) (DM) |
| 0.32 | index year: 2017 |
| -0.29 | observation during day -365 through 0 days relative to index: Discharge medications reconciled with the current medication list in outpatient medical record (COA) (GER) |
| -0.29 | measurement during day -365 through 0 days relative to index: Platelet mean volume [Entitic volume] in Blood |
| -0.27 | index year: 2019 |
| 0.26 | drug_era group during day -365 through 0 days relative to index: sacubitril |
| 0.25 | measurement during day -365 through 0 days relative to index: MCHC [Mass/volume] |
| 0.23 | condition_era group during day -365 through 0 days relative to index: Mitral valve disorder |
| -0.21 | Diabetes Comorbidity Severity Index (DCSI) |
| -0.21 | measurement within normal range during day -365 through 0 days relative to index: Oxygen [Partial pressure] in Blood |
| -0.21 | measurement during day -365 through 0 days relative to index: Urea nitrogen/Creatinine [Mass Ratio] in Blood |
| 0.19 | observation during day -365 through 0 days relative to index: Medication list documented in medical record (COA) |

**Supplementary Figure 35:** Kaplan Meier curve (top) and number of subjects at risk (bottom) for 3-pt MACE comparing dapagliflozin vs empagliflozin new-users in the OptumEHR data source.

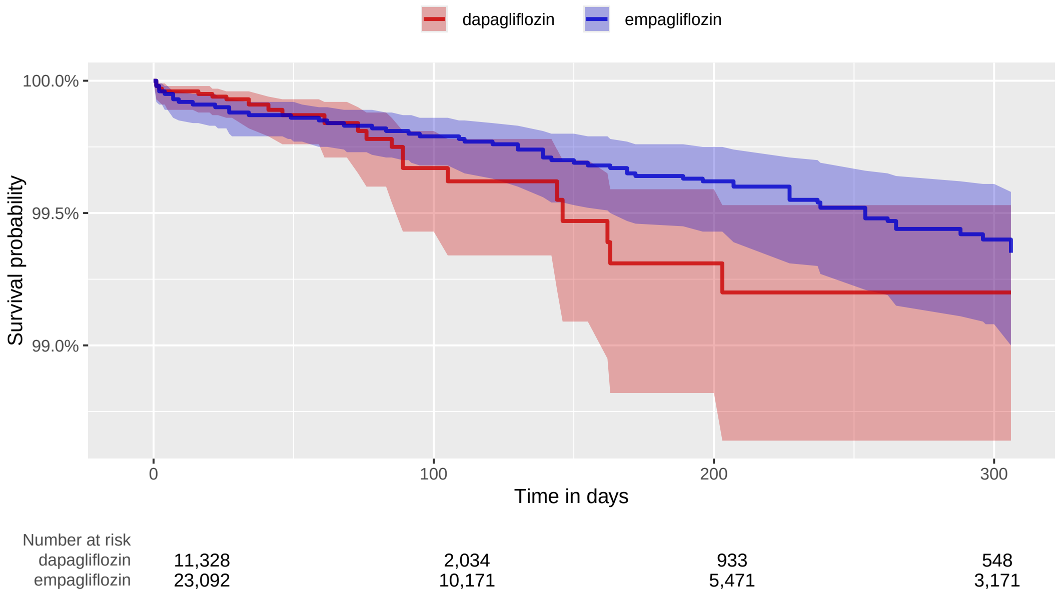

### 4 Negative control outcomes

We identified candidate negative control outcomes through a data-rich algorithm [21] that identifies prevalent OMOP condition concept occurrences that lack evidence of association with exposures of interest, and then adjudicated them by clinical review with a panel of experts. The data-rich algorithm was designed to automate the process to extract patterns from large observational datasets, without requiring a priori hypotheses. More specifically, it uses a combination of automated routines to search through published literature, drug-product labels and spontaneous reports, and then summarizes exposure-outcome relationships in accessible ways that are readable to both clinical and non-clinical experts for review and validation.

This two-stage process resulted in a list of 100 negative controls. We previously validated 60 of them in the LEGEND-HTN study [22].

The complete list of these controls are:

**Supplementary Table 40: Negative control outcomes.**

| Condition concept |
|---|
| Abnormal posture |
| Abrasion and/or friction burn of multiple sites |
| Absent kidney |
| Anomaly of jaw size |
| Benign paroxysmal positional vertigo |
| Bizarre personal appearance |
| Cachexia |
| Colostomy present |
| Complication of gastrostomy |
| Developmental delay |
| Deviated nasal septum |
| Epidermoid cyst |
| Exhaustion due to excessive exertion |
| Feces contents abnormal |
| Foreign body in ear |
| Foreskin deficient |
| Galactosemia |
| Ganglion cyst |
| Genetic disorder carrier |
| Impacted cerumen |
| Inadequate sleep hygiene |
| Jellyfish poisoning |
| Lagophthalmos |
| Lipid storage disease |
| Lymphangioma |
| Malingering |
| Marfan's syndrome |
| Mechanical complication of internal orthopedic device, implant AND/OR graft |
| Minimal cognitive impairment |
| Nicotine dependence |
| Nonspecific tuberculin test reaction |
| Opioid abuse |
| Opioid intoxication |
| Physiological development failure |
| Poisoning by tranquilizer |
| Social exclusion |

| |
|---|
| Symbolic dysfunction |
| Tooth loss |
| Toxic effect of lead compound |
| Toxic effect of tobacco and nicotine |
| Tracheostomy present |
| Unsatisfactory tooth restoration |
| Abnormal pupil |
| Abrasion and/or friction burn of trunk without infection |
| Absence of breast |
| Acquired hallux valgus |
| Acquired keratoderma |
| Anal and rectal polyp |
| Burn of forearm |
| Calcaneal spur |
| Cannabis abuse |
| Changes in skin texture |
| Chondromalacia of patella |
| Cocaine abuse |
| Complication due to Crohn’s disease |
| Contact dermatitis |
| Contusion of knee |
| Crohn’s disease |
| Derangement of knee |
| Difficulty sleeping |
| Disproportion of reconstructed breast |
| Effects of hunger |
| Endometriosis |
| Feces contents abnormal |
| Foreign body in orifice |
| Ganglion cyst |
| Hammer toe |
| Hereditary thrombophilia |
| High risk sexual behavior |
| Homocystinuria |
| Impacted cerumen |
| Impingement syndrome of shoulder region |
| Ingrowing nail |
| Injury of knee |
| Kwashiorkor |
| Late effect of contusion |
| Late effect of motor vehicle accident |
| Macular drusen |
| Melena |
| Nicotine dependence |
| Noise effects on inner ear |
| Non-toxic multinodular goiter |
| Nonspecific tuberculin test reaction |
| Opioid abuse |
| Passing flatus |
| Postviral fatigue syndrome |
| Presbyopia |
| Psychalgia |
| Ptotic breast |

| |
|---|
| Regular astigmatism |
| Senile hyperkeratosis |
| Somatic dysfunction of lumbar region |
| Splinter of face without major open wound |
| Sprain of ankle |
| Strain of rotator cuff capsule |
| Tear film insufficiency |
| Tobacco dependence syndrome |
| Verruca vulgaris |
| Wrist joint pain |
| Wristdrop |

### 5 Baseline patient characteristics across data sources

#### 5.1 dulaglutide vs semaglutide

##### 5.1.1 CCAE

**Supplementary Table 41:** Baseline patient characteristics for dulaglutide (T) and semaglutide (C) new-users in the CCAE data source. We report proportion of initiators satisfying selected base-line characteristics and the standardized difference of population proportions (StdDiff) before and after propensity score adjustment. Less extreme StdDiffs through matching and stratification suggest improved balance between patient cohorts through adjustment. The ”Medication use” entries refer to second-level ATC medication classes.

| | Before adjustment | | | After matching | | | After stratification | | |
|---|---|---|---|---|---|---|---|---|---|
| Characteristic | T | C | StdDiff | T | C | StdDiff | T | C | StdDiff |
| Age: mean | 51.3 | 50.9 | 0.04 | 51.3 | 51.2 | 0.01 | 51.3 | 51.1 | 0.02 |
| Age: std | 9.0 | 9.1 | | 9.2 | 9.1 | | 9.2 | 9.1 | |
| Characteristic (in proportions) | T (%) | C (%) | StdDiff | T (%) | C (%) | StdDiff | T (%) | C (%) | StdDiff |
| Gender: female | 56.2 | 61.1 | -0.10 | 56.3 | 55.9 | 0.01 | 58.8 | 59.0 | 0.00 |
| **Medical history: General** | | | | | | | | | |
| Chronic liver disease | 0.9 | 0.6 | 0.03 | 0.6 | 0.6 | 0.00 | 0.7 | 0.6 | 0.01 |
| Chronic obstructive lung disease | 2.4 | 2.2 | 0.01 | 2.3 | 2.2 | 0.01 | 2.4 | 2.2 | 0.01 |
| Dementia | 0.2 | 0.2 | 0.01 | 0.2 | 0.1 | 0.03 | 0.3 | 0.2 | 0.02 |
| Gastroesophageal reflux disease | 15.3 | 17.2 | -0.05 | 15.7 | 15.4 | 0.01 | 16.6 | 16.5 | 0.00 |
| Hyperlipidemia | 69.0 | 67.6 | 0.03 | 68.2 | 69.0 | -0.02 | 68.2 | 69.0 | -0.02 |
| Hypertensive disorder | 68.7 | 67.2 | 0.03 | 67.5 | 68.2 | -0.02 | 67.9 | 67.7 | 0.00 |
| Obesity | 42.2 | 48.9 | -0.14 | 44.5 | 44.1 | 0.01 | 46.5 | 46.8 | -0.01 |
| Renal impairment | 3.9 | 3.8 | 0.00 | 4.2 | 4.1 | 0.01 | 4.1 | 3.9 | 0.01 |
| Urinary tract infectious disease | 6.4 | 6.6 | 0.00 | 6.0 | 6.1 | 0.00 | 6.2 | 6.4 | -0.01 |
| Acute urinary tract infection | 0.3 | 0.3 | 0.00 | 0.3 | 0.3 | 0.01 | 0.3 | 0.3 | 0.01 |
| Closed fracture of lower limb | 1.2 | 1.1 | 0.01 | 1.1 | 1.2 | 0.00 | 1.2 | 1.2 | 0.00 |
| **Medical history: Cardiovascular disease** | | | | | | | | | |
| Cerebrovascular disease | 1.7 | 1.8 | 0.00 | 1.6 | 1.8 | -0.01 | 1.6 | 1.8 | -0.02 |
| Coronary arteriosclerosis | 5.6 | 5.2 | 0.02 | 5.2 | 5.5 | -0.01 | 5.3 | 5.3 | 0.00 |
| Heart disease | 15.6 | 15.2 | 0.01 | 15.3 | 15.0 | 0.01 | 15.6 | 15.1 | 0.01 |
| Heart failure | 2.0 | 1.8 | 0.01 | 2.0 | 1.9 | 0.01 | 2.0 | 1.9 | 0.01 |
| Ischemic heart disease | 3.2 | 2.9 | 0.02 | 3.3 | 3.0 | 0.02 | 3.3 | 3.0 | 0.02 |
| Peripheral vascular disease | 4.1 | 4.0 | 0.00 | 4.4 | 4.3 | 0.00 | 4.4 | 4.3 | 0.00 |
| Pulmonary embolism | 0.6 | 0.7 | -0.01 | 0.7 | 0.7 | 0.00 | 0.7 | 0.7 | 0.00 |
| Venous thrombosis | 1.0 | 1.0 | 0.01 | 1.0 | 1.0 | 0.00 | 1.1 | 1.0 | 0.01 |
| Medical history: Neoplasms | 5.6 | 5.5 | 0.00 | 5.5 | 5.3 | 0.01 | 5.7 | 5.5 | 0.01 |
| **Medication use** | | | | | | | | | |
| Agents acting on the renin-angiotensin system | 62.4 | 58.8 | 0.07 | 61.0 | 61.6 | -0.01 | 60.4 | 60.1 | 0.01 |
| Antibacterials for systemic use | 58.3 | 57.7 | 0.01 | 55.6 | 55.8 | 0.00 | 57.2 | 57.1 | 0.00 |
| Antithrombotic agents | 9.4 | 9.2 | 0.01 | 9.8 | 9.5 | 0.01 | 9.7 | 9.3 | 0.01 |
| Beta blocking agents | 23.5 | 23.2 | 0.01 | 23.4 | 23.3 | 0.00 | 24.1 | 23.3 | 0.02 |
| Calcium channel blockers | 20.0 | 20.4 | -0.01 | 20.2 | 20.4 | 0.00 | 20.9 | 20.5 | 0.01 |
| Diuretics | 38.3 | 39.5 | -0.03 | 38.0 | 38.2 | 0.00 | 38.0 | 38.2 | 0.00 |
| Immunosuppressants | 3.1 | 3.9 | -0.05 | 3.2 | 3.4 | -0.01 | 3.5 | 3.7 | -0.01 |
| Lipid modifying agents | 63.8 | 60.9 | 0.06 | 63.9 | 64.1 | 0.00 | 62.6 | 62.2 | 0.01 |

##### 5.1.2 OptumDOD

**Supplementary Table 42:** Baseline patient characteristics for dulaglutide (T) and semaglutide (C) new-users in the OptumDOD data source. We report proportion of initiators satisfying selected base-line characteristics and the standardized difference of population proportions (StdDiff) before and after propensity score adjustment. Less extreme StdDiffs through matching and stratification suggest improved balance between patient cohorts through adjustment. The ”Medication use” entries refer to second-level ATC medication classes.

| | Before adjustment | | | After matching | | | After stratification | | |
|---|---|---|---|---|---|---|---|---|---|
| Characteristic | T | C | StdDiff | T | C | StdDiff | T | C | StdDiff |
| Age: mean | 58.5 | 58.3 | 0.02 | 59.0 | 58.6 | 0.03 | 58.6 | 58.3 | 0.03 |
| Age: std | 12.3 | 12.3 | | 12.1 | 12.1 | | 12.1 | 12.1 | |
| Characteristic (in proportions) | T (%) | C (%) | StdDiff | T (%) | C (%) | StdDiff | T (%) | C (%) | StdDiff |
| Gender: female | 55.5 | 60.3 | -0.10 | 56.1 | 56.0 | 0.00 | 58.4 | 58.1 | 0.01 |
| **Medical history: General** | | | | | | | | | |
| Chronic liver disease | 1.2 | 1.1 | 0.01 | 1.2 | 1.2 | 0.00 | 1.1 | 1.2 | -0.01 |
| Chronic obstructive lung disease | 8.4 | 8.1 | 0.01 | 8.4 | 8.3 | 0.00 | 8.3 | 8.1 | 0.01 |
| Dementia | 1.2 | 0.7 | 0.05 | 1.1 | 0.8 | 0.03 | 1.0 | 0.8 | 0.03 |
| Gastroesophageal reflux disease | 21.8 | 23.4 | -0.04 | 22.6 | 22.1 | 0.01 | 22.9 | 22.6 | 0.01 |
| Hyperlipidemia | 75.7 | 77.5 | -0.04 | 75.9 | 76.6 | -0.02 | 76.5 | 76.9 | -0.01 |
| Hypertensive disorder | 76.3 | 76.3 | 0.00 | 76.5 | 76.6 | 0.00 | 76.2 | 76.1 | 0.00 |
| Obesity | 45.4 | 53.6 | -0.16 | 47.3 | 46.7 | 0.01 | 51.0 | 50.2 | 0.01 |
| Renal impairment | 11.3 | 11.0 | 0.01 | 11.6 | 11.6 | 0.00 | 11.6 | 11.6 | 0.00 |
| Urinary tract infectious disease | 8.6 | 8.8 | 0.00 | 8.3 | 8.4 | 0.00 | 8.6 | 8.6 | 0.00 |
| Acute urinary tract infection | 0.3 | 0.3 | 0.01 | 0.3 | 0.3 | 0.01 | 0.3 | 0.3 | 0.00 |
| Closed fracture of hip | 0.1 | 0.1 | 0.00 | 0.2 | 0.1 | 0.00 | 0.1 | 0.1 | 0.00 |
| Closed fracture of lower limb | 1.5 | 1.5 | -0.01 | 1.5 | 1.4 | 0.00 | 1.6 | 1.5 | 0.01 |
| **Medical history: Cardiovascular disease** | | | | | | | | | |
| Atrial fibrillation | 5.0 | 4.9 | 0.00 | 5.1 | 5.0 | 0.00 | 5.2 | 4.9 | 0.01 |
| Cerebrovascular disease | 4.5 | 4.5 | 0.00 | 4.5 | 4.6 | 0.00 | 4.6 | 4.5 | 0.00 |
| Coronary arteriosclerosis | 12.5 | 12.2 | 0.01 | 12.6 | 12.5 | 0.00 | 12.4 | 12.3 | 0.00 |
| Heart disease | 26.2 | 27.1 | -0.02 | 26.6 | 26.5 | 0.00 | 26.7 | 26.7 | 0.00 |
| Heart failure | 6.0 | 5.9 | 0.00 | 5.9 | 5.8 | 0.01 | 6.1 | 5.8 | 0.01 |
| Ischemic heart disease | 6.5 | 6.3 | 0.01 | 6.7 | 6.5 | 0.01 | 6.5 | 6.3 | 0.00 |
| Peripheral vascular disease | 10.4 | 10.2 | 0.01 | 11.1 | 10.8 | 0.01 | 10.6 | 10.4 | 0.01 |
| Pulmonary embolism | 0.9 | 1.1 | -0.03 | 1.0 | 1.1 | -0.01 | 1.0 | 1.1 | -0.01 |
| Venous thrombosis | 1.4 | 1.2 | 0.02 | 1.5 | 1.2 | 0.03 | 1.4 | 1.1 | 0.02 |
| Medical history: Neoplasms | 8.8 | 8.6 | 0.01 | 8.9 | 8.4 | 0.02 | 8.7 | 8.4 | 0.01 |
| **Medication use** | | | | | | | | | |
| Agents acting on the renin-angiotensin system | 66.7 | 65.2 | 0.03 | 66.2 | 66.8 | -0.01 | 65.1 | 65.9 | -0.02 |
| Antibacterials for systemic use | 59.1 | 58.8 | 0.00 | 57.5 | 57.4 | 0.00 | 58.6 | 58.4 | 0.00 |
| Antithrombotic agents | 14.1 | 14.2 | 0.00 | 14.5 | 14.6 | 0.00 | 14.6 | 14.3 | 0.01 |
| Beta blocking agents | 31.5 | 31.2 | 0.00 | 31.8 | 31.4 | 0.01 | 31.6 | 31.1 | 0.01 |
| Calcium channel blockers | 25.1 | 25.4 | -0.01 | 25.8 | 25.2 | 0.01 | 25.9 | 25.0 | 0.02 |
| Diuretics | 42.9 | 45.0 | -0.04 | 43.2 | 43.0 | 0.00 | 44.5 | 43.9 | 0.01 |
| Immunosuppressants | 3.8 | 4.2 | -0.02 | 4.0 | 3.6 | 0.02 | 4.1 | 3.9 | 0.01 |
| Lipid modifying agents | 72.3 | 71.1 | 0.03 | 73.5 | 73.0 | 0.01 | 72.0 | 71.8 | 0.01 |

##### 5.1.3 MDCR

**Supplementary Table 43:** Baseline patient characteristics for dulaglutide (T) and semaglutide (C) new-users in the MDCR data source. We report proportion of initiators satisfying selected base-line characteristics and the standardized difference of population proportions (StdDiff) before and after propensity score adjustment. Less extreme StdDiffs through matching and stratification suggest improved balance between patient cohorts through adjustment. The ”Medication use” entries refer to second-level ATC medication classes.

| | Before adjustment | | | After matching | | | After stratification | | |
|---|---|---|---|---|---|---|---|---|---|
| Characteristic | T | C | StdDiff | T | C | StdDiff | T | C | StdDiff |
| Age: mean | 71.5 | 71.2 | 0.06 | 71.8 | 71.2 | 0.11 | 71.6 | 71.3 | 0.07 |
| Age: std | 5.0 | 5.0 | | 5.0 | 5.2 | | 5.0 | 5.1 | |
| Characteristic (in proportions) | T (%) | C (%) | StdDiff | T (%) | C (%) | StdDiff | T (%) | C (%) | StdDiff |
| Gender: female | 54.0 | 60.0 | -0.12 | 54.8 | 55.0 | 0.00 | 56.0 | 56.9 | -0.02 |
| **Medical history: General** | | | | | | | | | |
| Chronic liver disease | 0.9 | 1.6 | -0.06 | <0.7 | 1.7 | -0.10 | 0.6 | 1.7 | -0.09 |
| Chronic obstructive lung disease | 10.3 | 9.1 | 0.04 | 11.5 | 9.4 | 0.07 | 10.8 | 8.9 | 0.06 |
| Dementia | 1.7 | 1.0 | 0.06 | 1.7 | 1.4 | 0.03 | 1.7 | 1.1 | 0.05 |
| Gastroesophageal reflux disease | 20.9 | 21.7 | -0.02 | 22.1 | 20.5 | 0.04 | 22.3 | 21.3 | 0.02 |
| Hyperlipidemia | 82.7 | 83.1 | -0.01 | 82.6 | 83.3 | -0.02 | 82.0 | 83.5 | -0.04 |
| Hypertensive disorder | 83.1 | 85.2 | -0.06 | 81.7 | 84.1 | -0.06 | 83.1 | 85.2 | -0.06 |
| Obesity | 41.5 | 48.6 | -0.14 | 43.0 | 44.3 | -0.03 | 45.7 | 46.6 | -0.02 |
| Renal impairment | 16.3 | 16.6 | -0.01 | 17.6 | 16.5 | 0.03 | 17.2 | 16.3 | 0.02 |
| Urinary tract infectious disease | 10.3 | 9.0 | 0.04 | 9.4 | 9.5 | 0.00 | 9.4 | 9.5 | 0.00 |
| Acute urinary tract infection | <0.4 | <0.4 | -0.01 | <0.7 | 0.4 | -0.02 | <0.4 | <0.4 | 0.00 |
| Closed fracture of hip | <0.4 | <0.4 | 0.03 | <0.7 | <0.3 | 0.03 | <0.4 | <0.4 | 0.02 |
| Closed fracture of lower limb | 2.5 | 1.7 | 0.06 | 2.8 | 1.9 | 0.06 | 2.4 | 1.9 | 0.04 |
| **Medical history: Cardiovascular disease** | | | | | | | | | |
| Atrial fibrillation | 9.7 | 9.9 | -0.01 | 9.0 | 10.4 | -0.05 | 8.5 | 10.0 | -0.05 |
| Cerebrovascular disease | 8.7 | 8.5 | 0.01 | 7.6 | 9.1 | -0.06 | 9.2 | 9.1 | 0.00 |
| Coronary arteriosclerosis | 22.8 | 20.6 | 0.05 | 23.0 | 21.5 | 0.04 | 21.2 | 21.1 | 0.00 |
| Heart disease | 40.8 | 39.5 | 0.03 | 40.6 | 39.8 | 0.02 | 38.6 | 39.5 | -0.02 |
| Heart failure | 7.8 | 7.6 | 0.01 | 8.4 | 8.0 | 0.01 | 8.0 | 7.7 | 0.01 |
| Ischemic heart disease | 10.3 | 8.4 | 0.06 | 9.7 | 9.0 | 0.03 | 9.3 | 8.4 | 0.03 |
| Peripheral vascular disease | 13.0 | 14.3 | -0.04 | 12.5 | 14.3 | -0.05 | 12.4 | 14.6 | -0.06 |
| Pulmonary embolism | 1.3 | 0.7 | 0.06 | 2.0 | 0.8 | 0.11 | 2.0 | 0.7 | 0.13 |
| Venous thrombosis | 2.3 | 1.2 | 0.09 | 2.3 | 1.0 | 0.11 | 2.2 | 1.0 | 0.09 |
| Medical history: Neoplasms | 17.4 | 16.6 | 0.02 | 15.8 | 17.8 | -0.05 | 15.3 | 17.3 | -0.05 |
| **Medication use** | | | | | | | | | |
| Agents acting on the renin-angiotensin system | 73.9 | 74.4 | -0.01 | 74.4 | 71.4 | 0.07 | 75.2 | 72.0 | 0.07 |
| Antibacterials for systemic use | 60.3 | 59.5 | 0.02 | 59.7 | 58.8 | 0.02 | 60.4 | 59.6 | 0.01 |
| Antithrombotic agents | 22.5 | 20.2 | 0.06 | 21.8 | 21.5 | 0.01 | 20.6 | 20.8 | -0.01 |
| Beta blocking agents | 44.0 | 44.3 | -0.01 | 45.8 | 42.8 | 0.06 | 44.5 | 43.5 | 0.02 |
| Calcium channel blockers | 31.6 | 34.1 | -0.05 | 34.7 | 30.9 | 0.08 | 35.3 | 31.7 | 0.07 |
| Diuretics | 48.6 | 52.2 | -0.07 | 50.4 | 46.8 | 0.07 | 51.9 | 48.8 | 0.06 |
| Immunosuppressants | 5.8 | 5.3 | 0.02 | 5.1 | 4.5 | 0.03 | 5.7 | 4.8 | 0.04 |
| Lipid modifying agents | 83.4 | 84.8 | -0.04 | 85.5 | 81.6 | 0.10 | 85.7 | 82.1 | 0.10 |

###### 5.1.4 Open Claims

##### 5.1.5 OptumEHR

**Supplementary Table 44:** Baseline patient characteristics for dulaglutide (T) and semaglutide (C) new-users in the OptumEHR data source. We report proportion of initiators satisfying selected base-line characteristics and the standardized difference of population proportions (StdDiff) before and after propensity score adjustment. Less extreme StdDiffs through matching and stratification suggest improved balance between patient cohorts through adjustment. The ”Medication use” entries refer to second-level ATC medication classes.

| | Before adjustment | | | After matching | | | After stratification | | |
|---|---|---|---|---|---|---|---|---|---|
| Characteristic | T | C | StdDiff | T | C | StdDiff | T | C | StdDiff |
| Age: mean | 53.7 | 53.4 | 0.02 | 54.0 | 53.5 | 0.04 | 53.7 | 53.4 | 0.03 |
| Age: std | 11.7 | 12.0 | | 11.9 | 11.7 | | 11.9 | 11.7 | |
| Characteristic (in proportions) | T (%) | C (%) | StdDiff | T (%) | C (%) | StdDiff | T (%) | C (%) | StdDiff |
| Gender: female | 58.4 | 62.6 | -0.09 | 59.6 | 59.2 | 0.01 | 61.0 | 59.9 | 0.02 |
| **Race** | | | | | | | | | |
| Asian | 1.7 | 1.9 | -0.02 | 1.8 | 1.8 | 0.00 | 1.7 | 1.9 | -0.02 |
| Black or African American | 12.6 | 13.9 | -0.04 | 13.5 | 12.8 | 0.02 | 13.5 | 13.0 | 0.01 |
| White | 78.2 | 77.1 | 0.03 | 77.1 | 77.9 | -0.02 | 77.6 | 77.8 | 0.00 |
| **Ethnicity** | | | | | | | | | |
| Hispanic or Latino | 6.5 | 5.6 | 0.04 | 6.4 | 6.2 | 0.01 | 6.2 | 6.1 | 0.00 |
| Not Hispanic or Latino | 84.8 | 84.6 | 0.01 | 83.6 | 84.2 | -0.02 | 84.2 | 84.6 | -0.01 |
| **Medical history: General** | | | | | | | | | |
| Chronic liver disease | 1.7 | 1.1 | 0.05 | 1.2 | 1.2 | 0.00 | 1.2 | 1.2 | 0.01 |
| Chronic obstructive lung disease | 4.3 | 4.0 | 0.02 | 4.4 | 4.1 | 0.01 | 4.4 | 4.0 | 0.02 |
| Dementia | 0.4 | 0.3 | 0.00 | 0.4 | 0.4 | 0.00 | 0.4 | 0.3 | 0.00 |
| Gastroesophageal reflux disease | 18.5 | 20.8 | -0.06 | 19.2 | 19.3 | 0.00 | 19.9 | 19.7 | 0.00 |
| Hyperlipidemia | 61.3 | 62.3 | -0.02 | 60.8 | 60.1 | 0.01 | 61.8 | 60.5 | 0.03 |
| Hypertensive disorder | 62.4 | 63.6 | -0.02 | 61.6 | 61.5 | 0.00 | 63.0 | 62.0 | 0.02 |
| Obesity | 43.7 | 51.6 | -0.16 | 46.5 | 45.8 | 0.01 | 48.1 | 46.9 | 0.02 |
| Renal impairment | 5.2 | 5.4 | -0.01 | 5.3 | 5.2 | 0.00 | 5.2 | 5.5 | -0.01 |
| Urinary tract infectious disease | 4.2 | 4.3 | -0.01 | 4.0 | 3.9 | 0.01 | 4.3 | 4.1 | 0.01 |
| Primary osteoporosis | 0.1 | 0.1 | -0.01 | 0.1 | 0.1 | -0.01 | 0.1 | 0.1 | -0.01 |
| Acute urinary tract infection | 0.6 | 1.1 | -0.06 | 0.7 | 0.9 | -0.03 | 0.8 | 0.9 | -0.01 |
| Closed fracture of hip | 0.0 | 0.0 | 0.01 | <0.1 | 0.0 | 0.00 | <0.1 | 0.0 | 0.00 |
| Closed fracture of lower limb | 0.8 | 0.8 | 0.00 | 0.8 | 0.7 | 0.00 | 0.9 | 1.0 | -0.01 |
| **Medical history: Cardiovascular disease** | | | | | | | | | |
| Atrial fibrillation | 2.8 | 3.0 | -0.01 | 2.9 | 2.9 | 0.00 | 3.0 | 2.9 | 0.00 |
| Cerebrovascular disease | 1.9 | 2.1 | -0.01 | 2.1 | 1.9 | 0.01 | 2.1 | 1.8 | 0.02 |
| Coronary arteriosclerosis | 7.2 | 7.1 | 0.01 | 7.4 | 7.2 | 0.01 | 7.4 | 6.8 | 0.02 |
| Heart disease | 16.5 | 17.2 | -0.02 | 16.7 | 16.6 | 0.00 | 17.2 | 16.5 | 0.02 |
| Heart failure | 2.3 | 2.5 | -0.01 | 2.3 | 2.6 | -0.02 | 2.4 | 2.4 | 0.00 |
| Ischemic heart disease | 3.4 | 3.8 | -0.02 | 3.5 | 3.8 | -0.01 | 3.5 | 3.6 | -0.01 |
| Peripheral vascular disease | 4.3 | 5.6 | -0.06 | 4.7 | 4.8 | 0.00 | 4.8 | 5.1 | -0.01 |
| Pulmonary embolism | 0.7 | 0.7 | -0.01 | 0.7 | 0.6 | 0.01 | 0.7 | 0.6 | 0.01 |
| Venous thrombosis | 0.8 | 0.9 | -0.01 | 0.8 | 0.8 | -0.01 | 0.9 | 0.9 | 0.00 |
| Chronic urinary tract infection | 0.1 | 0.1 | -0.02 | 0.1 | 0.1 | 0.00 | 0.1 | 0.1 | 0.00 |
| Medical history: Neoplasms | 5.3 | 5.1 | 0.01 | 5.6 | 5.0 | 0.03 | 5.4 | 5.3 | 0.00 |
| **Medication use** | | | | | | | | | |
| Agents acting on the renin-angiotensin system | 61.2 | 58.4 | 0.06 | 59.2 | 59.4 | 0.00 | 59.7 | 59.2 | 0.01 |
| Antibacterials for systemic use | 44.0 | 46.3 | -0.05 | 43.7 | 43.5 | 0.00 | 45.4 | 44.2 | 0.03 |
| Antithrombotic agents | 31.7 | 27.7 | 0.09 | 29.6 | 29.5 | 0.00 | 29.7 | 30.7 | -0.02 |
| Beta blocking agents | 27.4 | 28.5 | -0.03 | 28.1 | 28.1 | 0.00 | 27.9 | 28.7 | -0.02 |
| Calcium channel blockers | 20.5 | 22.3 | -0.04 | 21.6 | 21.2 | 0.01 | 21.5 | 21.5 | 0.00 |
| Diuretics | 38.9 | 40.5 | -0.03 | 39.8 | 38.7 | 0.02 | 39.9 | 38.9 | 0.02 |
| Immunosuppressants | 3.6 | 3.7 | 0.00 | 3.9 | 3.3 | 0.03 | 3.8 | 3.4 | 0.02 |
| Lipid modifying agents | 64.2 | 63.0 | 0.02 | 64.5 | 63.9 | 0.01 | 64.0 | 63.5 | 0.01 |

#### 5.2 dulaglutide vs dapagliflozin

##### 5.2.1 CCAE

**Supplementary Table 45:** Baseline patient characteristics for dulaglutide (T) and dapagliflozin (C) new-users in the CCAE data source. We report proportion of initiators satisfying selected base-line characteristics and the standardized difference of population proportions (StdDiff) before and after propensity score adjustment. Less extreme StdDiffs through matching and stratification suggest improved balance between patient cohorts through adjustment. The ”Medication use” entries refer to second-level ATC medication classes.

| | Before adjustment | | | After matching | | | After stratification | | |
|---|---|---|---|---|---|---|---|---|---|
| Characteristic | T | C | StdDiff | T | C | StdDiff | T | C | StdDiff |
| Age: mean | 51.3 | 52.5 | -0.14 | 51.9 | 52.0 | -0.01 | 51.9 | 51.9 | -0.01 |
| Age: std | 9.0 | 8.7 | | 8.5 | 8.7 | | 8.5 | 8.8 | |
| Characteristic (in proportions) | T (%) | C (%) | StdDiff | T (%) | C (%) | StdDiff | T (%) | C (%) | StdDiff |
| Gender: female | 56.2 | 43.7 | 0.25 | 48.2 | 50.1 | -0.04 | 48.6 | 50.0 | -0.03 |
| **Medical history: General** | | | | | | | | | |
| Chronic liver disease | 0.9 | 1.4 | -0.05 | 0.9 | 1.0 | -0.01 | 1.1 | 1.2 | -0.01 |
| Dementia | 0.2 | 0.1 | 0.03 | 0.2 | 0.1 | 0.02 | 0.2 | 0.1 | 0.01 |
| Gastroesophageal reflux disease | 15.3 | 13.3 | 0.06 | 14.0 | 14.3 | -0.01 | 14.0 | 14.3 | -0.01 |
| Hyperlipidemia | 69.0 | 73.3 | -0.10 | 71.5 | 71.0 | 0.01 | 71.5 | 71.2 | 0.01 |
| Hypertensive disorder | 68.7 | 71.2 | -0.05 | 70.4 | 70.1 | 0.01 | 70.4 | 70.0 | 0.01 |
| Obesity | 42.2 | 28.3 | 0.29 | 33.6 | 35.3 | -0.04 | 34.7 | 36.2 | -0.03 |
| Renal impairment | 3.9 | 4.7 | -0.04 | 4.2 | 4.2 | 0.00 | 4.4 | 4.4 | 0.00 |
| Urinary tract infectious disease | 6.4 | 5.9 | 0.02 | 5.8 | 6.2 | -0.02 | 5.9 | 6.3 | -0.02 |
| Acute urinary tract infection | 0.3 | 0.2 | 0.03 | 0.3 | 0.2 | 0.01 | 0.3 | 0.2 | 0.00 |
| Closed fracture of lower limb | 1.2 | 1.0 | 0.02 | 1.1 | 1.0 | 0.01 | 1.1 | 1.0 | 0.01 |
| **Medical history: Cardiovascular disease** | | | | | | | | | |
| Atrial fibrillation | 1.9 | 2.3 | -0.03 | 2.0 | 2.1 | 0.00 | 2.0 | 2.2 | -0.01 |
| Cerebrovascular disease | 1.7 | 2.3 | -0.04 | 1.8 | 2.0 | -0.01 | 1.9 | 2.2 | -0.02 |
| Coronary arteriosclerosis | 5.6 | 7.3 | -0.07 | 6.2 | 6.1 | 0.01 | 6.5 | 6.6 | 0.00 |
| Heart disease | 15.6 | 17.3 | -0.05 | 15.9 | 15.4 | 0.01 | 16.6 | 16.3 | 0.01 |
| Heart failure | 2.0 | 3.2 | -0.07 | 2.2 | 2.1 | 0.01 | 2.6 | 2.6 | 0.00 |
| Ischemic heart disease | 3.2 | 4.2 | -0.05 | 3.4 | 3.5 | 0.00 | 3.6 | 3.8 | -0.01 |
| Peripheral vascular disease | 4.1 | 3.8 | 0.01 | 3.8 | 3.9 | 0.00 | 3.9 | 4.0 | 0.00 |
| Pulmonary embolism | 0.6 | 0.4 | 0.03 | 0.5 | 0.4 | 0.02 | 0.6 | 0.4 | 0.02 |
| Venous thrombosis | 1.0 | 0.9 | 0.02 | 1.0 | 0.9 | 0.02 | 1.0 | 0.9 | 0.02 |
| Medical history: Neoplasms | 5.6 | 5.2 | 0.02 | 5.2 | 5.4 | -0.01 | 5.4 | 5.5 | -0.01 |
| **Medication use** | | | | | | | | | |
| Agents acting on the renin-angiotensin system | 62.4 | 67.4 | -0.10 | 65.5 | 64.7 | 0.02 | 65.5 | 64.9 | 0.01 |
| Antibacterials for systemic use | 58.3 | 56.2 | 0.04 | 56.1 | 56.8 | -0.01 | 57.1 | 57.6 | -0.01 |
| Antithrombotic agents | 9.4 | 10.0 | -0.02 | 9.3 | 9.2 | 0.00 | 9.7 | 9.6 | 0.00 |
| Beta blocking agents | 23.5 | 25.5 | -0.05 | 24.2 | 24.1 | 0.00 | 24.6 | 24.7 | 0.00 |
| Calcium channel blockers | 20.0 | 21.6 | -0.04 | 21.1 | 20.9 | 0.00 | 21.1 | 20.9 | 0.00 |
| Diuretics | 38.3 | 37.8 | 0.01 | 37.3 | 37.8 | -0.01 | 37.9 | 38.1 | 0.00 |
| Immunosuppressants | 3.1 | 2.3 | 0.05 | 2.5 | 2.5 | 0.00 | 2.7 | 2.6 | 0.01 |
| Lipid modifying agents | 63.8 | 68.2 | -0.09 | 67.1 | 66.4 | 0.01 | 66.6 | 66.4 | 0.00 |

##### 5.2.2 OptumDOD

**Supplementary Table 46:** Baseline patient characteristics for dulaglutide (T) and dapagliflozin (C) new-users in the OptumDOD data source. We report proportion of initiators satisfying selected base-line characteristics and the standardized difference of population proportions (StdDiff) before and after propensity score adjustment. Less extreme StdDiffs through matching and stratification suggest improved balance between patient cohorts through adjustment. The ”Medication use” entries refer to second-level ATC medication classes.

| | Before adjustment | | | After matching | | | After stratification | | |
|---|---|---|---|---|---|---|---|---|---|
| Characteristic | T | C | StdDiff | T | C | StdDiff | T | C | StdDiff |
| Age: mean | 58.5 | 67.2 | -0.69 | 64.1 | 64.3 | -0.02 | 62.0 | 61.8 | 0.02 |
| Age: std | 12.3 | 11.9 | | 11.4 | 11.3 | | 11.4 | 12.5 | |
| Characteristic (in proportions) | T (%) | C (%) | StdDiff | T (%) | C (%) | StdDiff | T (%) | C (%) | StdDiff |
| Gender: female | 55.5 | 45.4 | 0.20 | 50.3 | 50.1 | 0.00 | 51.3 | 49.9 | 0.03 |
| **Medical history: General** | | | | | | | | | |
| Chronic liver disease | 1.2 | 2.3 | -0.09 | 1.5 | 1.7 | -0.02 | 1.5 | 1.5 | 0.00 |
| Chronic obstructive lung disease | 8.4 | 14.6 | -0.20 | 11.0 | 10.6 | 0.01 | 11.1 | 10.7 | 0.01 |
| Dementia | 1.2 | 2.6 | -0.11 | 1.9 | 1.7 | 0.01 | 1.9 | 1.7 | 0.01 |
| Gastroesophageal reflux disease | 21.8 | 23.6 | -0.04 | 22.2 | 22.6 | -0.01 | 23.4 | 22.5 | 0.02 |
| Hyperlipidemia | 75.7 | 84.7 | -0.22 | 81.6 | 80.8 | 0.02 | 79.8 | 78.8 | 0.02 |
| Hypertensive disorder | 76.3 | 85.5 | -0.23 | 82.5 | 82.1 | 0.01 | 80.4 | 79.9 | 0.01 |
| Obesity | 45.4 | 29.6 | 0.32 | 33.2 | 34.8 | -0.03 | 39.0 | 39.5 | -0.01 |
| Renal impairment | 11.3 | 30.3 | -0.49 | 19.3 | 17.2 | 0.05 | 19.4 | 17.6 | 0.05 |
| Urinary tract infectious disease | 8.6 | 10.0 | -0.05 | 9.2 | 8.8 | 0.01 | 9.4 | 9.0 | 0.01 |
| Acute urinary tract infection | 0.3 | 0.2 | 0.04 | 0.3 | 0.2 | 0.03 | 0.3 | 0.4 | -0.02 |
| Closed fracture of hip | 0.1 | 0.4 | -0.05 | 0.3 | 0.3 | 0.01 | 0.2 | 0.2 | -0.01 |
| Closed fracture of lower limb | 1.5 | 1.6 | -0.01 | 1.7 | 1.6 | 0.01 | 1.4 | 1.6 | -0.02 |
| **Medical history: Cardiovascular disease** | | | | | | | | | |
| Atrial fibrillation | 5.0 | 14.1 | -0.32 | 8.2 | 7.1 | 0.04 | 8.4 | 7.9 | 0.02 |
| Cerebrovascular disease | 4.5 | 9.5 | -0.20 | 6.7 | 6.6 | 0.00 | 7.4 | 6.1 | 0.05 |
| Coronary arteriosclerosis | 12.5 | 27.5 | -0.39 | 19.2 | 17.4 | 0.05 | 18.7 | 17.3 | 0.04 |
| Heart disease | 26.2 | 46.9 | -0.44 | 36.9 | 33.8 | 0.07 | 35.1 | 33.0 | 0.04 |
| Heart failure | 6.0 | 21.9 | -0.49 | 11.1 | 8.7 | 0.08 | 11.9 | 11.5 | 0.01 |
| Ischemic heart disease | 6.5 | 15.7 | -0.30 | 9.8 | 8.9 | 0.03 | 9.9 | 9.8 | 0.00 |
| Peripheral vascular disease | 10.4 | 19.1 | -0.25 | 14.7 | 14.3 | 0.01 | 14.4 | 13.5 | 0.03 |
| Pulmonary embolism | 0.9 | 1.3 | -0.04 | 1.1 | 1.0 | 0.02 | 1.0 | 1.1 | -0.01 |
| Venous thrombosis | 1.4 | 2.1 | -0.05 | 1.6 | 1.6 | 0.00 | 2.0 | 1.5 | 0.04 |
| Medical history: Neoplasms | 8.8 | 12.3 | -0.12 | 10.1 | 10.3 | -0.01 | 10.0 | 9.0 | 0.04 |
| **Medication use** | | | | | | | | | |
| Agents acting on the renin-angiotensin system | 66.7 | 78.7 | -0.27 | 73.4 | 73.3 | 0.00 | 71.2 | 70.8 | 0.01 |
| Antibacterials for systemic use | 59.1 | 55.9 | 0.06 | 55.8 | 55.3 | 0.01 | 58.5 | 56.6 | 0.04 |
| Antithrombotic agents | 14.1 | 27.0 | -0.33 | 19.3 | 18.0 | 0.03 | 19.3 | 18.0 | 0.03 |
| Beta blocking agents | 31.5 | 48.8 | -0.36 | 40.0 | 38.0 | 0.04 | 38.2 | 37.0 | 0.03 |
| Calcium channel blockers | 25.1 | 34.3 | -0.20 | 30.9 | 30.1 | 0.02 | 28.8 | 27.4 | 0.03 |
| Diuretics | 42.9 | 52.8 | -0.20 | 46.6 | 45.9 | 0.01 | 47.1 | 45.7 | 0.03 |
| Immunosuppressants | 3.8 | 3.3 | 0.03 | 3.6 | 3.7 | -0.01 | 3.6 | 3.7 | -0.01 |
| Lipid modifying agents | 72.3 | 83.8 | -0.27 | 80.2 | 79.7 | 0.01 | 77.1 | 75.6 | 0.04 |

##### 5.2.3 MDCR

**Supplementary Table 47:** Baseline patient characteristics for dulaglutide (T) and dapagliflozin (C) new-users in the MDCR data source. We report proportion of initiators satisfying selected base-line characteristics and the standardized difference of population proportions (StdDiff) before and after propensity score adjustment. Less extreme StdDiffs through matching and stratification suggest improved balance between patient cohorts through adjustment. The "Medication use" entries refer to second-level ATC medication classes.

| | Before adjustment | | | After matching | | | After stratification | | |
|---|---|---|---|---|---|---|---|---|---|
| Characteristic | T | C | StdDiff | T | C | StdDiff | T | C | StdDiff |
| Age: mean | 71.5 | 72.8 | -0.23 | 71.7 | 72.1 | -0.07 | 72.0 | 72.4 | -0.08 |
| Age: std | 5.0 | 5.4 | | 5.9 | 5.6 | | 5.9 | 5.7 | |
| Characteristic (in proportions) | T (%) | C (%) | StdDiff | T (%) | C (%) | StdDiff | T (%) | C (%) | StdDiff |
| Gender: female | 54.0 | 42.5 | 0.23 | 46.0 | 49.2 | -0.06 | 46.3 | 50.0 | -0.07 |
| **Medical history: General** | | | | | | | | | |
| Chronic liver disease | 0.9 | 1.8 | -0.07 | 1.0 | 2.3 | -0.11 | 1.2 | 1.7 | -0.04 |
| Chronic obstructive lung disease | 10.3 | 11.5 | -0.04 | 9.2 | 9.6 | -0.01 | 11.2 | 10.4 | 0.03 |
| Dementia | 1.7 | 1.9 | -0.01 | 1.3 | 1.5 | -0.02 | 2.0 | 2.5 | -0.04 |
| Gastroesophageal reflux disease | 20.9 | 17.0 | 0.10 | 17.5 | 18.3 | -0.02 | 17.5 | 18.3 | -0.02 |
| Hyperlipidemia | 82.7 | 82.5 | 0.00 | 82.2 | 83.2 | -0.03 | 82.2 | 85.0 | -0.07 |
| Hypertensive disorder | 83.1 | 85.8 | -0.07 | 84.5 | 86.5 | -0.06 | 84.4 | 87.1 | -0.08 |
| Obesity | 41.5 | 23.4 | 0.39 | 24.9 | 30.6 | -0.13 | 30.6 | 32.6 | -0.04 |
| Renal impairment | 16.3 | 22.6 | -0.16 | 18.0 | 16.6 | 0.04 | 18.5 | 18.6 | 0.00 |
| Urinary tract infectious disease | 10.3 | 8.1 | 0.07 | 10.6 | 8.7 | 0.06 | 10.9 | 8.4 | 0.09 |
| Primary osteoporosis | <0.4 | <0.4 | -0.04 | <0.8 | <0.3 | 0.01 | <0.4 | <0.4 | -0.01 |
| Acute urinary tract infection | <0.4 | <0.4 | 0.00 | <0.8 | <0.3 | 0.01 | <0.4 | <0.4 | 0.00 |
| Closed fracture of hip | <0.4 | <0.4 | 0.00 | <0.8 | <0.3 | 0.01 | <0.4 | <0.4 | 0.01 |
| Closed fracture of lower limb | 2.5 | 1.2 | 0.09 | 1.8 | 1.3 | 0.04 | 1.8 | 2.1 | -0.02 |
| **Medical history: Cardiovascular disease** | | | | | | | | | |
| Atrial fibrillation | 9.7 | 14.0 | -0.13 | 10.3 | 10.4 | 0.00 | 11.7 | 12.0 | -0.01 |
| Cerebrovascular disease | 8.7 | 13.0 | -0.14 | 9.7 | 9.6 | 0.00 | 10.4 | 10.3 | 0.00 |
| Coronary arteriosclerosis | 22.8 | 26.4 | -0.08 | 24.7 | 19.8 | 0.12 | 26.6 | 21.9 | 0.11 |
| Heart disease | 40.8 | 49.1 | -0.17 | 43.2 | 40.9 | 0.05 | 45.9 | 43.4 | 0.05 |
| Heart failure | 7.8 | 15.4 | -0.24 | 9.5 | 8.4 | 0.04 | 12.1 | 11.7 | 0.01 |
| Ischemic heart disease | 10.3 | 12.2 | -0.06 | 11.0 | 8.2 | 0.10 | 11.5 | 9.4 | 0.07 |
| Peripheral vascular disease | 13.0 | 15.9 | -0.08 | 12.6 | 13.6 | -0.03 | 14.9 | 15.0 | 0.00 |
| Pulmonary embolism | 1.3 | 1.1 | 0.02 | 1.5 | 1.2 | 0.02 | 1.6 | 1.3 | 0.03 |
| Venous thrombosis | 2.3 | 2.6 | -0.01 | 2.8 | 2.3 | 0.03 | 2.2 | 2.3 | 0.00 |
| Medical history: Neoplasms | 17.4 | 18.3 | -0.02 | 16.4 | 17.1 | -0.02 | 17.1 | 16.6 | 0.01 |
| **Medication use** | | | | | | | | | |
| Agents acting on the renin-angiotensin system | 73.9 | 76.8 | -0.07 | 77.9 | 74.0 | 0.09 | 76.7 | 72.9 | 0.09 |
| Antibacterials for systemic use | 60.3 | 60.2 | 0.00 | 56.6 | 59.7 | -0.06 | 58.9 | 59.9 | -0.02 |
| Antithrombotic agents | 22.5 | 26.8 | -0.10 | 23.7 | 22.0 | 0.04 | 26.0 | 24.6 | 0.03 |
| Beta blocking agents | 44.0 | 51.1 | -0.14 | 46.2 | 44.4 | 0.04 | 49.3 | 47.0 | 0.05 |
| Calcium channel blockers | 31.6 | 34.7 | -0.07 | 34.2 | 31.8 | 0.05 | 33.8 | 29.5 | 0.09 |
| Diuretics | 48.6 | 51.6 | -0.06 | 50.6 | 49.3 | 0.03 | 51.8 | 50.3 | 0.03 |
| Immunosuppressants | 5.8 | 4.2 | 0.07 | 3.8 | 4.1 | -0.02 | 5.0 | 4.9 | 0.00 |
| Lipid modifying agents | 83.4 | 85.3 | -0.05 | 84.8 | 82.0 | 0.07 | 85.8 | 81.2 | 0.13 |

##### 5.2.4 MDCD

**Supplementary Table 48:** Baseline patient characteristics for dulaglutide (T) and dapagliflozin (C) new-users in the MDCD data source. We report proportion of initiators satisfying selected base-line characteristics and the standardized difference of population proportions (StdDiff) before and after propensity score adjustment. Less extreme StdDiffs through matching and stratification suggest improved balance between patient cohorts through adjustment. The ”Medication use” entries refer to second-level ATC medication classes.

| | Before adjustment | | | After matching | | | After stratification | | |
|---|---|---|---|---|---|---|---|---|---|
| Characteristic | T | C | StdDiff | T | C | StdDiff | T | C | StdDiff |
| Age: mean | 46.9 | 51.0 | -0.35 | 50.0 | 49.4 | 0.05 | 48.3 | 48.4 | -0.01 |
| Age: std | 11.5 | 11.0 | | 11.7 | 11.8 | | 11.7 | 11.8 | |
| Characteristic (in proportions) | T (%) | C (%) | StdDiff | T (%) | C (%) | StdDiff | T (%) | C (%) | StdDiff |
| Gender: female | 70.9 | 56.3 | 0.31 | 59.5 | 64.7 | -0.11 | 66.6 | 68.7 | -0.04 |
| **Race** | | | | | | | | | |
| Black or African American | 21.5 | 25.2 | -0.09 | 22.0 | 21.3 | 0.02 | 21.5 | 21.5 | 0.00 |
| White | 59.6 | 50.9 | 0.17 | 56.5 | 57.7 | -0.02 | 57.3 | 60.4 | -0.06 |
| **Ethnicity** | | | | | | | | | |
| Hispanic or Latino | 7.1 | 7.9 | -0.03 | 8.3 | 6.7 | 0.06 | 8.2 | 5.7 | 0.09 |
| **Medical history: General** | | | | | | | | | |
| Chronic liver disease | 2.0 | 2.9 | -0.06 | 1.8 | 2.4 | -0.04 | 2.2 | 2.4 | -0.01 |
| Chronic obstructive lung disease | 15.2 | 20.0 | -0.13 | 17.8 | 16.8 | 0.03 | 17.2 | 16.7 | 0.01 |
| Dementia | 0.5 | 0.8 | -0.04 | 0.5 | 0.7 | -0.02 | 0.6 | 0.5 | 0.01 |
| Gastroesophageal reflux disease | 28.1 | 26.9 | 0.03 | 26.9 | 28.8 | -0.04 | 26.9 | 28.8 | -0.04 |
| Hyperlipidemia | 59.4 | 67.5 | -0.17 | 64.2 | 62.7 | 0.03 | 62.2 | 62.0 | 0.00 |
| Hypertensive disorder | 69.9 | 75.6 | -0.13 | 73.1 | 70.3 | 0.06 | 73.0 | 69.9 | 0.07 |
| Obesity | 57.7 | 43.6 | 0.28 | 44.3 | 48.7 | -0.09 | 52.6 | 56.9 | -0.09 |
| Renal impairment | 6.6 | 12.4 | -0.21 | 8.9 | 8.6 | 0.01 | 8.4 | 9.0 | -0.02 |
| Urinary tract infectious disease | 9.8 | 8.9 | 0.03 | 9.1 | 10.9 | -0.06 | 9.1 | 10.9 | -0.06 |
| Acute urinary tract infection | 0.6 | 0.5 | 0.01 | 0.5 | 0.5 | 0.00 | 0.6 | 0.5 | 0.01 |
| Closed fracture of hip | <0.2 | <0.3 | -0.05 | <0.2 | <0.2 | -0.04 | <0.2 | <0.2 | -0.04 |
| Closed fracture of lower limb | 1.5 | 1.9 | -0.03 | 1.4 | 1.5 | -0.01 | 1.4 | 1.2 | 0.01 |
| **Medical history: Cardiovascular disease** | | | | | | | | | |
| Atrial fibrillation | 2.7 | 5.7 | -0.15 | 3.7 | 3.4 | 0.02 | 3.6 | 4.0 | -0.02 |
| Cerebrovascular disease | 3.2 | 5.4 | -0.11 | 3.4 | 3.9 | -0.03 | 3.7 | 4.2 | -0.03 |
| Coronary arteriosclerosis | 8.2 | 14.8 | -0.21 | 12.3 | 8.8 | 0.12 | 11.2 | 9.0 | 0.07 |
| Heart disease | 23.7 | 32.9 | -0.20 | 28.6 | 23.9 | 0.11 | 28.9 | 25.4 | 0.08 |
| Heart failure | 6.3 | 17.7 | -0.37 | 10.5 | 7.9 | 0.09 | 11.2 | 10.7 | 0.02 |
| Ischemic heart disease | 5.2 | 12.5 | -0.27 | 7.8 | 6.5 | 0.05 | 7.8 | 6.5 | 0.05 |
| Peripheral vascular disease | 6.8 | 9.3 | -0.09 | 7.3 | 7.2 | 0.00 | 8.2 | 7.7 | 0.02 |
| Pulmonary embolism | 1.6 | 2.1 | -0.04 | 1.2 | 1.3 | -0.01 | 1.4 | 1.9 | -0.04 |
| Venous thrombosis | 1.9 | 1.4 | 0.04 | 1.9 | 1.0 | 0.07 | 2.0 | 1.2 | 0.06 |
| Medical history: Neoplasms | 4.8 | 6.4 | -0.07 | 5.5 | 6.9 | -0.06 | 5.4 | 7.5 | -0.09 |
| **Medication use** | | | | | | | | | |
| Agents acting on the renin-angiotensin system | 55.7 | 63.8 | -0.17 | 61.1 | 58.1 | 0.06 | 59.6 | 58.1 | 0.03 |
| Antibacterials for systemic use | 67.1 | 62.8 | 0.09 | 63.0 | 63.0 | 0.00 | 65.8 | 66.0 | 0.00 |
| Antithrombotic agents | 22.6 | 27.9 | -0.12 | 24.8 | 22.3 | 0.06 | 26.4 | 24.2 | 0.05 |
| Beta blocking agents | 29.1 | 39.8 | -0.23 | 33.9 | 31.5 | 0.05 | 34.1 | 31.0 | 0.07 |
| Calcium channel blockers | 21.7 | 26.9 | -0.12 | 25.2 | 24.4 | 0.02 | 23.9 | 23.4 | 0.01 |
| Diuretics | 41.3 | 48.9 | -0.16 | 44.3 | 42.3 | 0.04 | 45.2 | 43.9 | 0.03 |
| Immunosuppressants | 3.0 | 2.7 | 0.02 | 3.7 | 3.0 | 0.04 | 3.8 | 2.8 | 0.06 |
| Lipid modifying agents | 60.2 | 70.3 | -0.21 | 68.9 | 65.5 | 0.07 | 64.9 | 63.6 | 0.03 |

##### 5.2.5 Open Claims

**Supplementary Table 49:** Baseline patient characteristics for dulaglutide (T) and dapagliflozin (C) new-users in the Open Claims data source. We report proportion of initiators satisfying selected base-line characteristics and the standardized difference of population proportions (StdDiff) before and after propensity score adjustment. Less extreme StdDiffs through matching and stratification suggest improved balance between patient cohorts through adjustment. The ”Medication use” entries refer to second-level ATC medication classes.

| | Before adjustment | | | After matching | | | After stratification | | |
|---|---|---|---|---|---|---|---|---|---|
| Characteristic | T | C | StdDiff | T | C | StdDiff | T | C | StdDiff |
| Age: mean | 55.3 | 57.8 | -0.20 | 56.8 | 56.9 | -0.01 | 56.6 | 56.8 | -0.01 |
| Age: std | 12.1 | 11.8 | | 11.8 | 11.8 | | 11.8 | 12.0 | |
| Characteristic (in proportions) | T (%) | C (%) | StdDiff | T (%) | C (%) | StdDiff | T (%) | C (%) | StdDiff |
| Gender: female | 57.9 | 45.9 | 0.24 | 50.3 | 50.9 | -0.01 | 51.6 | 51.9 | 0.00 |
| **Medical history: General** | | | | | | | | | |
| Chronic liver disease | 0.7 | 1.0 | -0.04 | 0.7 | 0.7 | 0.00 | 0.8 | 0.8 | 0.00 |
| Chronic obstructive lung disease | 4.3 | 4.2 | 0.00 | 3.9 | 4.0 | 0.00 | 4.3 | 4.4 | 0.00 |
| Dementia | 0.4 | 0.4 | -0.01 | 0.4 | 0.4 | 0.00 | 0.4 | 0.4 | 0.00 |
| Gastroesophageal reflux disease | 9.4 | 8.0 | 0.05 | 8.3 | 8.2 | 0.00 | 8.8 | 8.7 | 0.00 |
| Hyperlipidemia | 39.7 | 44.4 | -0.10 | 41.2 | 40.8 | 0.01 | 42.4 | 42.0 | 0.01 |
| Hypertensive disorder | 46.6 | 49.3 | -0.06 | 46.7 | 46.6 | 0.00 | 48.2 | 48.0 | 0.00 |
| Obesity | 19.1 | 12.3 | 0.19 | 13.9 | 13.9 | 0.00 | 16.0 | 15.6 | 0.01 |
| Renal impairment | 4.5 | 6.0 | -0.07 | 4.7 | 4.8 | 0.00 | 5.3 | 5.4 | 0.00 |
| Urinary tract infectious disease | 4.7 | 4.0 | 0.04 | 4.0 | 4.0 | 0.00 | 4.3 | 4.3 | 0.00 |
| Acute urinary tract infection | 0.2 | 0.1 | 0.02 | 0.1 | 0.1 | 0.00 | 0.1 | 0.1 | 0.00 |
| Closed fracture of hip | 0.1 | 0.1 | 0.00 | 0.1 | 0.1 | 0.00 | 0.1 | 0.1 | 0.00 |
| Closed fracture of lower limb | 0.9 | 0.7 | 0.02 | 0.7 | 0.8 | 0.00 | 0.8 | 0.8 | 0.00 |
| **Medical history: Cardiovascular disease** | | | | | | | | | |
| Atrial fibrillation | 2.6 | 3.9 | -0.07 | 2.8 | 2.9 | 0.00 | 3.1 | 3.3 | -0.01 |
| Cerebrovascular disease | 1.9 | 2.4 | -0.03 | 2.1 | 2.0 | 0.00 | 2.2 | 2.2 | 0.00 |
| Coronary arteriosclerosis | 5.6 | 8.2 | -0.10 | 6.3 | 6.3 | 0.00 | 6.9 | 7.0 | 0.00 |
| Heart disease | 13.4 | 17.8 | -0.12 | 14.2 | 14.3 | 0.00 | 15.6 | 16.0 | -0.01 |
| Heart failure | 2.7 | 5.6 | -0.15 | 3.0 | 3.1 | 0.00 | 3.8 | 4.4 | -0.03 |
| Ischemic heart disease | 2.7 | 4.2 | -0.09 | 3.0 | 3.0 | 0.00 | 3.3 | 3.6 | -0.01 |
| Peripheral vascular disease | 3.7 | 4.1 | -0.02 | 3.7 | 3.7 | 0.00 | 4.0 | 4.0 | 0.00 |
| Pulmonary embolism | 0.5 | 0.4 | 0.01 | 0.4 | 0.4 | 0.00 | 0.5 | 0.5 | 0.00 |
| Venous thrombosis | 0.7 | 0.7 | 0.00 | 0.6 | 0.6 | 0.00 | 0.7 | 0.7 | 0.00 |
| Medical history: Neoplasms | 4.4 | 4.8 | -0.02 | 4.4 | 4.5 | 0.00 | 4.7 | 4.7 | 0.00 |
| **Medication use** | | | | | | | | | |
| Agents acting on the renin-angiotensin system | 62.3 | 69.2 | -0.14 | 66.2 | 65.9 | 0.01 | 65.8 | 66.0 | 0.00 |
| Antibacterials for systemic use | 57.4 | 54.9 | 0.05 | 54.9 | 55.1 | 0.00 | 56.3 | 56.3 | 0.00 |
| Antithrombotic agents | 14.2 | 17.4 | -0.09 | 15.0 | 15.0 | 0.00 | 15.8 | 16.0 | -0.01 |
| Beta blocking agents | 28.3 | 33.2 | -0.11 | 29.9 | 29.9 | 0.00 | 30.9 | 31.1 | 0.00 |
| Calcium channel blockers | 22.3 | 24.5 | -0.05 | 23.7 | 23.7 | 0.00 | 23.8 | 23.5 | 0.00 |
| Diuretics | 39.8 | 41.1 | -0.03 | 39.7 | 39.8 | 0.00 | 40.3 | 40.8 | -0.01 |
| Immunosuppressants | 3.2 | 2.6 | 0.04 | 2.7 | 2.8 | 0.00 | 2.9 | 3.0 | 0.00 |
| Lipid modifying agents | 65.7 | 71.0 | -0.11 | 69.4 | 69.1 | 0.00 | 68.8 | 68.7 | 0.00 |

##### 5.2.6 OptumEHR

**Supplementary Table 50:** Baseline patient characteristics for dulaglutide (T) and dapagliflozin (C) new-users in the OptumEHR data source. We report proportion of initiators satisfying selected base-line characteristics and the standardized difference of population proportions (StdDiff) before and after propensity score adjustment. Less extreme StdDiffs through matching and stratification suggest improved balance between patient cohorts through adjustment. The "Medication use" entries refer to second-level ATC medication classes.

| | Before adjustment | | | After matching | | | After stratification | | |
|---|---|---|---|---|---|---|---|---|---|
| Characteristic | T | C | StdDiff | T | C | StdDiff | T | C | StdDiff |
| Age: mean | 53.7 | 55.7 | -0.17 | 54.9 | 55.3 | -0.04 | 54.4 | 54.8 | -0.03 |
| Age: std | 11.7 | 11.6 | | 11.4 | 11.5 | | 11.4 | 11.6 | |
| Characteristic (in proportions) | T (%) | C (%) | StdDiff | T (%) | C (%) | StdDiff | T (%) | C (%) | StdDiff |
| Gender: female | 58.4 | 46.0 | 0.25 | 48.7 | 51.2 | -0.05 | 51.9 | 53.4 | -0.03 |
| **Race** | | | | | | | | | |
| Asian | 1.7 | 2.1 | -0.03 | 1.8 | 2.0 | -0.02 | 1.7 | 2.0 | -0.02 |
| Black or African American | 12.6 | 12.3 | 0.01 | 12.3 | 12.3 | 0.00 | 12.5 | 12.1 | 0.01 |
| White | 78.2 | 78.4 | 0.00 | 78.6 | 78.3 | 0.01 | 78.6 | 78.7 | 0.00 |
| **Ethnicity** | | | | | | | | | |
| Hispanic or Latino | 6.5 | 6.1 | 0.02 | 5.8 | 6.3 | -0.02 | 6.1 | 6.1 | 0.00 |
| Not Hispanic or Latino | 84.8 | 85.9 | -0.03 | 85.1 | 85.4 | -0.01 | 85.3 | 85.9 | -0.02 |
| **Medical history: General** | | | | | | | | | |
| Chronic liver disease | 1.7 | 1.7 | 0.00 | 1.5 | 1.5 | 0.00 | 1.6 | 1.7 | 0.00 |
| Chronic obstructive lung disease | 4.3 | 4.5 | -0.01 | 4.5 | 4.3 | 0.01 | 4.5 | 4.3 | 0.01 |
| Dementia | 0.4 | 0.3 | 0.01 | 0.3 | 0.3 | 0.01 | 0.4 | 0.3 | 0.01 |
| Gastroesophageal reflux disease | 18.5 | 17.2 | 0.03 | 16.2 | 17.2 | -0.03 | 17.6 | 17.9 | -0.01 |
| Hyperlipidemia | 61.3 | 65.9 | -0.10 | 63.3 | 62.3 | 0.02 | 63.6 | 62.7 | 0.02 |
| Hypertensive disorder | 62.4 | 67.6 | -0.11 | 65.0 | 64.3 | 0.02 | 65.0 | 64.3 | 0.01 |
| Obesity | 43.7 | 32.6 | 0.23 | 34.3 | 36.3 | -0.04 | 38.2 | 39.8 | -0.03 |
| Renal impairment | 5.2 | 5.4 | -0.01 | 5.3 | 5.2 | 0.00 | 5.3 | 5.4 | 0.00 |
| Urinary tract infectious disease | 4.2 | 3.5 | 0.03 | 3.6 | 3.5 | 0.00 | 3.9 | 3.7 | 0.01 |
| Primary osteoporosis | 0.1 | 0.1 | -0.01 | <0.1 | 0.1 | -0.03 | 0.1 | 0.2 | -0.02 |
| Acute urinary tract infection | 0.6 | 0.4 | 0.02 | 0.5 | 0.4 | 0.00 | 0.5 | 0.5 | -0.01 |
| Closed fracture of hip | 0.0 | 0.0 | 0.00 | <0.1 | 0.0 | 0.00 | <0.1 | 0.0 | 0.00 |
| Closed fracture of lower limb | 0.8 | 0.7 | 0.01 | 0.5 | 0.8 | -0.04 | 0.7 | 0.8 | -0.01 |
| **Medical history: Cardiovascular disease** | | | | | | | | | |
| Atrial fibrillation | 2.8 | 3.8 | -0.05 | 3.3 | 3.1 | 0.01 | 3.4 | 3.2 | 0.01 |
| Cerebrovascular disease | 1.9 | 2.1 | -0.01 | 2.2 | 2.0 | 0.01 | 2.1 | 1.9 | 0.01 |
| Coronary arteriosclerosis | 7.2 | 10.9 | -0.13 | 9.2 | 8.7 | 0.02 | 8.8 | 8.7 | 0.00 |
| Heart disease | 16.5 | 20.2 | -0.10 | 18.5 | 17.3 | 0.03 | 18.5 | 17.8 | 0.02 |
| Heart failure | 2.3 | 4.9 | -0.14 | 2.9 | 2.9 | 0.00 | 3.3 | 3.5 | -0.01 |
| Ischemic heart disease | 3.4 | 5.3 | -0.09 | 4.1 | 4.0 | 0.01 | 4.0 | 4.2 | -0.01 |
| Peripheral vascular disease | 4.3 | 5.6 | -0.06 | 4.9 | 4.9 | 0.00 | 4.8 | 5.0 | -0.01 |
| Pulmonary embolism | 0.7 | 0.6 | 0.01 | 0.5 | 0.5 | 0.00 | 0.6 | 0.5 | 0.01 |
| Venous thrombosis | 0.8 | 0.9 | -0.01 | 0.8 | 1.0 | -0.02 | 0.8 | 1.0 | -0.02 |
| Chronic urinary tract infection | 0.1 | 0.1 | 0.01 | 0.1 | 0.1 | 0.00 | 0.1 | 0.1 | 0.00 |
| Medical history: Neoplasms | 5.3 | 4.9 | 0.02 | 4.9 | 5.1 | -0.01 | 5.2 | 5.1 | 0.01 |
| **Medication use** | | | | | | | | | |
| Agents acting on the renin-angiotensin system | 61.2 | 67.1 | -0.12 | 65.6 | 64.4 | 0.03 | 64.4 | 63.6 | 0.02 |
| Antibacterials for systemic use | 44.0 | 42.9 | 0.02 | 42.4 | 42.8 | -0.01 | 43.8 | 43.9 | 0.00 |
| Antithrombotic agents | 31.7 | 34.1 | -0.05 | 33.3 | 32.1 | 0.03 | 32.6 | 32.4 | 0.01 |
| Beta blocking agents | 27.4 | 31.9 | -0.10 | 29.9 | 29.3 | 0.01 | 29.6 | 29.4 | 0.00 |
| Calcium channel blockers | 20.5 | 22.4 | -0.05 | 22.4 | 21.7 | 0.02 | 21.7 | 21.2 | 0.01 |
| Diuretics | 38.9 | 39.3 | -0.01 | 38.1 | 38.4 | 0.00 | 39.2 | 38.8 | 0.01 |
| Immunosuppressants | 3.6 | 2.5 | 0.06 | 3.0 | 2.8 | 0.01 | 3.2 | 2.9 | 0.01 |
| Lipid modifying agents | 64.2 | 68.1 | -0.08 | 67.6 | 66.3 | 0.03 | 66.1 | 65.5 | 0.01 |

#### 5.3 dulaglutide vs empagliflozin

##### 5.3.1 CCAE

**Supplementary Table 51:** Baseline patient characteristics for dulaglutide (T) and empagliflozin (C) new-users in the CCAE data source. We report proportion of initiators satisfying selected base-line characteristics and the standardized difference of population proportions (StdDiff) before and after propensity score adjustment. Less extreme StdDiffs through matching and stratification suggest improved balance between patient cohorts through adjustment. The "Medication use" entries refer to second-level ATC medication classes.

| | Before adjustment | | | After matching | | | After stratification | | |
|---|---|---|---|---|---|---|---|---|---|
| Characteristic | T | C | StdDiff | T | C | StdDiff | T | C | StdDiff |
| Age: mean | 51.3 | 53.2 | -0.22 | 51.8 | 52.0 | -0.02 | 52.3 | 52.5 | -0.03 |
| Age: std | 9.0 | 8.8 | | 8.3 | 8.6 | | 8.3 | 8.5 | |
| Characteristic (in proportions) | T (%) | C (%) | StdDiff | T (%) | C (%) | StdDiff | T (%) | C (%) | StdDiff |
| Gender: female | 56.2 | 40.3 | 0.32 | 51.2 | 53.0 | -0.04 | 45.3 | 47.1 | -0.04 |
| **Medical history: General** | | | | | | | | | |
| Chronic liver disease | 0.9 | 0.9 | 0.00 | 0.9 | 0.9 | -0.01 | 0.8 | 0.9 | -0.01 |
| Chronic obstructive lung disease | 2.4 | 2.6 | -0.02 | 2.4 | 2.6 | -0.01 | 2.5 | 2.7 | -0.01 |
| Dementia | 0.2 | 0.2 | 0.01 | 0.2 | 0.2 | 0.00 | 0.2 | 0.2 | 0.00 |
| Gastroesophageal reflux disease | 15.3 | 13.6 | 0.05 | 14.4 | 14.8 | -0.01 | 14.1 | 14.3 | -0.01 |
| Hyperlipidemia | 69.0 | 74.3 | -0.12 | 71.0 | 70.5 | 0.01 | 71.0 | 70.5 | 0.01 |
| Hypertensive disorder | 68.7 | 70.9 | -0.05 | 69.9 | 69.3 | 0.01 | 70.3 | 70.0 | 0.01 |
| Obesity | 42.2 | 30.2 | 0.25 | 37.4 | 39.0 | -0.03 | 34.6 | 35.5 | -0.02 |
| Renal impairment | 3.9 | 4.3 | -0.02 | 3.9 | 3.9 | 0.00 | 4.2 | 4.1 | 0.01 |
| Urinary tract infectious disease | 6.4 | 4.5 | 0.09 | 5.5 | 5.7 | -0.01 | 5.1 | 5.3 | 0.00 |
| Acute urinary tract infection | 0.3 | 0.1 | 0.05 | 0.2 | 0.2 | 0.02 | 0.3 | 0.1 | 0.03 |
| Closed fracture of lower limb | 1.2 | 0.9 | 0.03 | 1.0 | 1.1 | -0.01 | 1.0 | 1.1 | -0.01 |
| **Medical history: Cardiovascular disease** | | | | | | | | | |
| Atrial fibrillation | 1.9 | 2.5 | -0.04 | 2.0 | 1.9 | 0.00 | 2.3 | 2.3 | 0.00 |
| Cerebrovascular disease | 1.7 | 2.5 | -0.05 | 1.9 | 2.0 | -0.01 | 2.1 | 2.3 | -0.01 |
| Coronary arteriosclerosis | 5.6 | 10.7 | -0.18 | 6.3 | 5.7 | 0.02 | 8.4 | 8.6 | -0.01 |
| Heart disease | 15.6 | 20.4 | -0.12 | 16.0 | 15.1 | 0.03 | 18.4 | 18.2 | 0.01 |
| Heart failure | 2.0 | 3.4 | -0.08 | 2.1 | 2.1 | -0.01 | 2.6 | 2.9 | -0.02 |
| Ischemic heart disease | 3.2 | 5.7 | -0.12 | 3.5 | 3.2 | 0.02 | 4.4 | 4.7 | -0.01 |
| Peripheral vascular disease | 4.1 | 4.5 | -0.02 | 4.2 | 3.9 | 0.01 | 4.6 | 4.3 | 0.02 |
| Pulmonary embolism | 0.6 | 0.4 | 0.03 | 0.6 | 0.4 | 0.02 | 0.6 | 0.4 | 0.03 |
| Venous thrombosis | 1.0 | 0.8 | 0.02 | 0.9 | 0.9 | 0.01 | 1.0 | 0.9 | 0.02 |
| **Medication use** | | | | | | | | | |
| Agents acting on the renin-angiotensin system | 62.4 | 66.9 | -0.10 | 64.6 | 64.1 | 0.01 | 65.6 | 65.2 | 0.01 |
| Antibacterials for systemic use | 58.3 | 52.3 | 0.12 | 56.0 | 56.8 | -0.02 | 54.1 | 54.9 | -0.02 |
| Antithrombotic agents | 9.4 | 12.6 | -0.10 | 9.7 | 9.3 | 0.01 | 11.1 | 11.3 | -0.01 |
| Beta blocking agents | 23.5 | 26.9 | -0.08 | 23.8 | 23.5 | 0.01 | 25.3 | 25.7 | -0.01 |
| Calcium channel blockers | 20.0 | 21.8 | -0.04 | 20.8 | 20.3 | 0.01 | 21.1 | 21.0 | 0.00 |
| Diuretics | 38.3 | 35.8 | 0.05 | 37.7 | 38.0 | -0.01 | 36.3 | 37.0 | -0.01 |
| Immunosuppressants | 3.1 | 2.8 | 0.01 | 2.9 | 3.2 | -0.02 | 2.8 | 3.1 | -0.02 |
| Lipid modifying agents | 63.8 | 72.5 | -0.19 | 67.5 | 66.4 | 0.02 | 69.5 | 69.1 | 0.01 |

##### 5.3.2 OptumDOD

**Supplementary Table 52:** Baseline patient characteristics for dulaglutide (T) and empagliflozin (C) new-users in the OptumDOD data source. We report proportion of initiators satisfying selected base-line characteristics and the standardized difference of population proportions (StdDiff) before and after propensity score adjustment. Less extreme StdDiffs through matching and stratification suggest improved balance between patient cohorts through adjustment. The ”Medication use” entries refer to second-level ATC medication classes.

| | Before adjustment | | | After matching | | | After stratification | | |
|---|---|---|---|---|---|---|---|---|---|
| Characteristic | T | C | StdDiff | T | C | StdDiff | T | C | StdDiff |
| Age: mean | 58.5 | 62.3 | -0.31 | 59.0 | 59.2 | -0.01 | 60.9 | 61.3 | -0.04 |
| Age: std | 12.3 | 12.2 | | 11.9 | 12.0 | | 11.9 | 12.1 | |
| Characteristic (in proportions) | T (%) | C (%) | StdDiff | T (%) | C (%) | StdDiff | T (%) | C (%) | StdDiff |
| Gender: female | 55.5 | 41.2 | 0.29 | 53.2 | 55.0 | -0.04 | 43.6 | 45.6 | -0.04 |
| **Medical history: General** | | | | | | | | | |
| Chronic liver disease | 1.2 | 1.3 | 0.00 | 1.2 | 1.2 | 0.00 | 1.2 | 1.3 | 0.00 |
| Chronic obstructive lung disease | 8.4 | 9.1 | -0.02 | 8.2 | 8.1 | 0.00 | 9.1 | 8.9 | 0.01 |
| Dementia | 1.2 | 1.3 | -0.01 | 1.1 | 1.1 | 0.00 | 1.1 | 1.2 | 0.00 |
| Gastroesophageal reflux disease | 21.8 | 20.3 | 0.04 | 21.2 | 21.9 | -0.02 | 20.5 | 20.9 | -0.01 |
| Hyperlipidemia | 75.7 | 81.8 | -0.15 | 76.9 | 76.1 | 0.02 | 80.4 | 79.9 | 0.01 |
| Hypertensive disorder | 76.3 | 79.8 | -0.09 | 76.7 | 76.5 | 0.00 | 79.3 | 78.7 | 0.01 |
| Obesity | 45.4 | 32.5 | 0.27 | 42.6 | 44.5 | -0.04 | 35.6 | 36.6 | -0.02 |
| Renal impairment | 11.3 | 14.2 | -0.09 | 11.5 | 11.0 | 0.01 | 13.4 | 13.3 | 0.00 |
| Urinary tract infectious disease | 8.6 | 6.9 | 0.06 | 8.2 | 8.3 | 0.00 | 7.4 | 7.4 | 0.00 |
| Acute urinary tract infection | 0.3 | 0.2 | 0.03 | 0.3 | 0.3 | 0.01 | 0.3 | 0.2 | 0.01 |
| Closed fracture of hip | 0.1 | 0.2 | -0.01 | 0.1 | 0.2 | 0.00 | 0.2 | 0.2 | -0.01 |
| Closed fracture of lower limb | 1.5 | 1.3 | 0.02 | 1.4 | 1.3 | 0.00 | 1.3 | 1.3 | 0.00 |
| **Medical history: Cardiovascular disease** | | | | | | | | | |
| Atrial fibrillation | 5.0 | 8.5 | -0.14 | 5.2 | 4.9 | 0.01 | 7.4 | 7.5 | 0.00 |
| Cerebrovascular disease | 4.5 | 6.6 | -0.09 | 4.7 | 4.4 | 0.01 | 6.3 | 6.0 | 0.01 |
| Coronary arteriosclerosis | 12.5 | 21.8 | -0.24 | 13.1 | 12.4 | 0.02 | 19.7 | 19.0 | 0.02 |
| Heart disease | 26.2 | 36.7 | -0.22 | 26.9 | 25.8 | 0.02 | 34.4 | 33.4 | 0.02 |
| Heart failure | 6.0 | 11.2 | -0.18 | 6.1 | 5.8 | 0.01 | 9.3 | 9.7 | -0.01 |
| Ischemic heart disease | 6.5 | 12.0 | -0.18 | 6.9 | 6.6 | 0.01 | 10.3 | 10.4 | 0.00 |
| Peripheral vascular disease | 10.4 | 13.4 | -0.09 | 10.5 | 10.3 | 0.01 | 12.5 | 12.5 | 0.00 |
| Pulmonary embolism | 0.9 | 1.0 | -0.01 | 0.8 | 0.8 | 0.00 | 0.9 | 0.9 | -0.01 |
| Venous thrombosis | 1.4 | 1.6 | -0.01 | 1.4 | 1.4 | 0.00 | 1.6 | 1.5 | 0.00 |
| Medical history: Neoplasms | 8.8 | 10.4 | -0.05 | 8.9 | 8.7 | 0.01 | 9.9 | 9.9 | 0.00 |
| **Medication use** | | | | | | | | | |
| Agents acting on the renin-angiotensin system | 66.7 | 71.6 | -0.11 | 67.7 | 67.4 | 0.01 | 70.8 | 70.3 | 0.01 |
| Antibacterials for systemic use | 59.1 | 53.1 | 0.12 | 57.7 | 58.9 | -0.03 | 54.8 | 55.1 | -0.01 |
| Antithrombotic agents | 14.1 | 21.1 | -0.18 | 14.2 | 14.0 | 0.01 | 19.2 | 19.0 | 0.01 |
| Beta blocking agents | 31.5 | 39.0 | -0.16 | 31.9 | 31.3 | 0.01 | 37.0 | 36.7 | 0.01 |
| Calcium channel blockers | 25.1 | 28.4 | -0.07 | 25.6 | 25.2 | 0.01 | 27.5 | 27.3 | 0.00 |
| Diuretics | 42.9 | 42.6 | 0.01 | 42.6 | 42.8 | 0.00 | 42.6 | 42.8 | 0.00 |
| Immunosuppressants | 3.8 | 3.0 | 0.04 | 3.5 | 3.5 | 0.00 | 3.0 | 3.2 | -0.01 |
| Lipid modifying agents | 72.3 | 80.3 | -0.19 | 73.9 | 72.8 | 0.02 | 78.5 | 77.8 | 0.02 |

##### 5.3.3 MDCR

**Supplementary Table 53:** Baseline patient characteristics for dulaglutide (T) and empagliflozin (C) new-users in the MDCR data source. We report proportion of initiators satisfying selected base-line characteristics and the standardized difference of population proportions (StdDiff) before and after propensity score adjustment. Less extreme StdDiffs through matching and stratification suggest improved balance between patient cohorts through adjustment. The ”Medication use” entries refer to second-level ATC medication classes.

| | Before adjustment | | | After matching | | | After stratification | | |
|---|---|---|---|---|---|---|---|---|---|
| Characteristic | T | C | StdDiff | T | C | StdDiff | T | C | StdDiff |
| Age: mean | 71.5 | 72.9 | -0.26 | 71.7 | 71.8 | -0.02 | 72.4 | 72.6 | -0.03 |
| Age: std | 5.0 | 5.0 | | 5.6 | 5.3 | | 5.6 | 5.6 | |
| Characteristic (in proportions) | T (%) | C (%) | StdDiff | T (%) | C (%) | StdDiff | T (%) | C (%) | StdDiff |
| Gender: female | 54.0 | 40.9 | 0.26 | 52.0 | 54.4 | -0.05 | 42.2 | 44.6 | -0.05 |
| **Medical history: General** | | | | | | | | | |
| Chronic liver disease | 0.9 | 1.3 | -0.04 | 0.9 | 1.4 | -0.04 | 0.9 | 1.4 | -0.04 |
| Chronic obstructive lung disease | 10.3 | 11.9 | -0.05 | 10.1 | 10.4 | -0.01 | 11.1 | 11.4 | -0.01 |
| Dementia | 1.7 | 1.6 | 0.01 | 1.8 | 1.7 | 0.01 | 2.5 | 1.6 | 0.07 |
| Gastroesophageal reflux disease | 20.9 | 19.4 | 0.04 | 20.1 | 22.1 | -0.05 | 19.1 | 20.1 | -0.02 |
| Hyperlipidemia | 82.7 | 84.5 | -0.05 | 82.7 | 82.5 | 0.01 | 84.1 | 83.9 | 0.00 |
| Hypertensive disorder | 83.1 | 86.4 | -0.09 | 83.0 | 84.5 | -0.04 | 84.8 | 85.9 | -0.03 |
| Obesity | 41.5 | 29.3 | 0.26 | 38.5 | 40.8 | -0.05 | 31.7 | 32.8 | -0.02 |
| Renal impairment | 16.3 | 20.8 | -0.11 | 16.4 | 17.3 | -0.02 | 19.1 | 19.9 | -0.02 |
| Urinary tract infectious disease | 10.3 | 7.4 | 0.10 | 9.5 | 8.1 | 0.05 | 9.1 | 7.6 | 0.06 |
| Acute urinary tract infection | <0.4 | 0.2 | 0.01 | <0.5 | 0.3 | 0.00 | <0.4 | 0.2 | -0.01 |
| Closed fracture of hip | <0.4 | 0.4 | -0.04 | <0.5 | 0.4 | -0.03 | <0.4 | 0.4 | -0.04 |
| Closed fracture of lower limb | 2.5 | 1.6 | 0.07 | 2.2 | 1.7 | 0.04 | 2.0 | 1.6 | 0.03 |
| **Medical history: Cardiovascular disease** | | | | | | | | | |
| Atrial fibrillation | 9.7 | 16.8 | -0.20 | 10.0 | 9.6 | 0.01 | 14.1 | 14.9 | -0.02 |
| Cerebrovascular disease | 8.7 | 10.3 | -0.06 | 9.0 | 8.1 | 0.03 | 9.0 | 8.1 | 0.03 |
| Coronary arteriosclerosis | 22.8 | 35.3 | -0.27 | 23.7 | 20.8 | 0.07 | 34.3 | 31.4 | 0.06 |
| Heart disease | 40.8 | 55.3 | -0.29 | 41.8 | 39.4 | 0.05 | 53.5 | 51.0 | 0.05 |
| Heart failure | 7.8 | 17.7 | -0.28 | 8.2 | 8.0 | 0.01 | 14.7 | 15.1 | -0.01 |
| Ischemic heart disease | 10.3 | 17.0 | -0.19 | 10.7 | 10.0 | 0.03 | 15.3 | 15.2 | 0.00 |
| Peripheral vascular disease | 13.0 | 16.9 | -0.10 | 13.2 | 12.7 | 0.01 | 16.1 | 15.7 | 0.01 |
| Pulmonary embolism | 1.3 | 1.1 | 0.02 | 1.3 | 1.0 | 0.03 | 1.9 | 1.1 | 0.08 |
| Venous thrombosis | 2.3 | 2.2 | 0.01 | 2.3 | 2.0 | 0.02 | 2.8 | 2.1 | 0.05 |
| Medical history: Neoplasms | 17.4 | 18.0 | -0.01 | 17.3 | 17.4 | 0.00 | 18.2 | 17.9 | 0.01 |
| **Medication use** | | | | | | | | | |
| Agents acting on the renin-angiotensin system | 73.9 | 77.8 | -0.09 | 74.6 | 75.5 | -0.02 | 74.6 | 75.5 | -0.02 |
| Antibacterials for systemic use | 60.3 | 57.4 | 0.06 | 59.4 | 62.4 | -0.06 | 55.7 | 58.8 | -0.06 |
| Antithrombotic agents | 22.5 | 31.8 | -0.20 | 22.4 | 21.3 | 0.03 | 29.4 | 29.0 | 0.01 |
| Beta blocking agents | 44.0 | 54.0 | -0.20 | 45.0 | 42.0 | 0.06 | 54.3 | 50.7 | 0.07 |
| Calcium channel blockers | 31.6 | 36.0 | -0.09 | 31.7 | 33.4 | -0.04 | 33.4 | 35.4 | -0.04 |
| Diuretics | 48.6 | 51.8 | -0.06 | 50.2 | 50.9 | -0.01 | 50.2 | 50.9 | -0.01 |
| Immunosuppressants | 5.8 | 3.9 | 0.09 | 5.5 | 4.8 | 0.03 | 4.2 | 4.1 | 0.00 |
| Lipid modifying agents | 83.4 | 86.5 | -0.09 | 83.7 | 83.2 | 0.02 | 85.9 | 85.6 | 0.01 |

##### 5.3.4 MDCD

**Supplementary Table 54:** Baseline patient characteristics for dulaglutide (T) and empagliflozin (C) new-users in the MDCD data source. We report proportion of initiators satisfying selected base-line characteristics and the standardized difference of population proportions (StdDiff) before and after propensity score adjustment. Less extreme StdDiffs through matching and stratification suggest improved balance between patient cohorts through adjustment. The "Medication use" entries refer to second-level ATC medication classes.

| | Before adjustment | | | After matching | | | After stratification | | |
|---|---|---|---|---|---|---|---|---|---|
| Characteristic | T | C | StdDiff | T | C | StdDiff | T | C | StdDiff |
| Age: mean | 46.9 | 51.0 | -0.35 | 48.7 | 49.0 | -0.02 | 49.1 | 49.4 | -0.02 |
| Age: std | 11.5 | 11.2 | | 11.3 | 11.6 | | 11.3 | 11.6 | |
| Characteristic (in proportions) | T (%) | C (%) | StdDiff | T (%) | C (%) | StdDiff | T (%) | C (%) | StdDiff |
| Gender: female | 70.9 | 56.0 | 0.31 | 63.5 | 67.3 | -0.08 | 60.7 | 64.0 | -0.07 |
| **Race** | | | | | | | | | |
| Black or African American | 21.5 | 23.4 | -0.05 | 21.8 | 21.6 | 0.00 | 20.6 | 22.5 | -0.04 |
| White | 59.6 | 57.0 | 0.05 | 59.3 | 59.7 | -0.01 | 60.5 | 58.5 | 0.04 |
| **Ethnicity** | | | | | | | | | |
| Hispanic or Latino | 7.1 | 5.2 | 0.08 | 6.3 | 5.4 | 0.04 | 6.2 | 5.5 | 0.03 |
| **Medical history: General** | | | | | | | | | |
| Chronic liver disease | 2.0 | 3.1 | -0.07 | 2.5 | 3.0 | -0.03 | 2.2 | 3.0 | -0.05 |
| Chronic obstructive lung disease | 15.2 | 18.1 | -0.08 | 15.8 | 16.4 | -0.02 | 16.4 | 16.8 | -0.01 |
| Dementia | 0.5 | 1.1 | -0.06 | 0.6 | 1.0 | -0.04 | 1.1 | 1.1 | 0.00 |
| Gastroesophageal reflux disease | 28.1 | 28.5 | -0.01 | 27.2 | 28.3 | -0.03 | 27.5 | 29.2 | -0.04 |
| Hyperlipidemia | 59.4 | 66.8 | -0.15 | 62.3 | 61.1 | 0.03 | 63.3 | 62.9 | 0.01 |
| Hypertensive disorder | 69.9 | 75.1 | -0.12 | 72.3 | 71.5 | 0.02 | 73.6 | 73.2 | 0.01 |
| Obesity | 57.7 | 42.2 | 0.31 | 47.7 | 51.8 | -0.08 | 47.3 | 50.8 | -0.07 |
| Renal impairment | 6.6 | 8.9 | -0.09 | 7.5 | 6.5 | 0.04 | 7.8 | 7.5 | 0.01 |
| Urinary tract infectious disease | 9.8 | 7.4 | 0.09 | 7.8 | 8.9 | -0.04 | 8.3 | 8.8 | -0.02 |
| Acute urinary tract infection | 0.6 | 0.4 | 0.04 | 0.7 | 0.5 | 0.02 | 0.5 | 0.4 | 0.02 |
| Closed fracture of hip | <0.2 | 0.2 | -0.05 | <0.2 | 0.2 | -0.04 | <0.2 | 0.2 | -0.04 |
| Closed fracture of lower limb | 1.5 | 1.9 | -0.03 | 1.5 | 1.9 | -0.04 | 1.4 | 2.1 | -0.05 |
| **Medical history: Cardiovascular disease** | | | | | | | | | |
| Atrial fibrillation | 2.7 | 4.5 | -0.09 | 3.7 | 3.0 | 0.04 | 3.7 | 3.6 | 0.01 |
| Cerebrovascular disease | 3.2 | 4.6 | -0.07 | 3.6 | 3.1 | 0.03 | 4.0 | 3.7 | 0.01 |
| Coronary arteriosclerosis | 8.2 | 15.7 | -0.23 | 10.4 | 9.6 | 0.03 | 13.2 | 12.2 | 0.03 |
| Heart disease | 23.7 | 32.5 | -0.19 | 27.4 | 25.1 | 0.05 | 29.7 | 28.1 | 0.03 |
| Heart failure | 6.3 | 13.0 | -0.22 | 7.6 | 7.6 | 0.00 | 10.0 | 10.2 | -0.01 |
| Ischemic heart disease | 5.2 | 11.1 | -0.21 | 7.1 | 6.2 | 0.04 | 8.3 | 8.2 | 0.00 |
| Peripheral vascular disease | 6.8 | 11.1 | -0.15 | 7.9 | 8.5 | -0.02 | 9.9 | 9.5 | 0.01 |
| Pulmonary embolism | 1.6 | 1.6 | 0.00 | 1.3 | 1.2 | 0.01 | 1.6 | 1.5 | 0.01 |
| Venous thrombosis | 1.9 | 1.8 | 0.01 | 1.9 | 1.6 | 0.02 | 1.9 | 1.9 | 0.00 |
| Medical history: Neoplasms | 4.8 | 5.7 | -0.04 | 5.2 | 5.8 | -0.03 | 6.3 | 6.0 | 0.01 |
| **Medication use** | | | | | | | | | |
| Agents acting on the renin-angiotensin system | 55.7 | 62.0 | -0.13 | 60.2 | 56.9 | 0.07 | 60.8 | 58.7 | 0.04 |
| Antibacterials for systemic use | 67.1 | 61.4 | 0.12 | 62.4 | 65.2 | -0.06 | 62.6 | 64.8 | -0.05 |
| Antithrombotic agents | 22.6 | 29.0 | -0.15 | 24.8 | 23.4 | 0.03 | 27.0 | 26.6 | 0.01 |
| Beta blocking agents | 29.1 | 36.1 | -0.15 | 31.9 | 30.4 | 0.03 | 32.7 | 32.3 | 0.01 |
| Calcium channel blockers | 21.7 | 24.6 | -0.07 | 23.9 | 21.8 | 0.05 | 24.2 | 22.5 | 0.04 |
| Diuretics | 41.3 | 43.7 | -0.05 | 41.4 | 41.6 | 0.00 | 41.7 | 42.7 | -0.02 |
| Immunosuppressants | 3.0 | 2.4 | 0.04 | 3.2 | 2.6 | 0.04 | 3.3 | 2.7 | 0.04 |
| Lipid modifying agents | 60.2 | 71.3 | -0.23 | 66.5 | 63.0 | 0.07 | 67.2 | 65.2 | 0.04 |

##### 5.3.5 Open Claims

**Supplementary Table 55:** Baseline patient characteristics for dulaglutide (T) and empagliflozin (C) new-users in the Open Claims data source. We report proportion of initiators satisfying selected base-line characteristics and the standardized difference of population proportions (StdDiff) before and after propensity score adjustment. Less extreme StdDiffs through matching and stratification suggest improved balance between patient cohorts through adjustment. The "Medication use" entries refer to second-level ATC medication classes.

| | Before adjustment | | | After matching | | | After stratification | | |
|---|---|---|---|---|---|---|---|---|---|
| Characteristic | T | C | StdDiff | T | C | StdDiff | T | C | StdDiff |
| Age: mean | 55.3 | 59.4 | -0.33 | 56.1 | 56.1 | 0.00 | 57.9 | 58.0 | -0.01 |
| Age: std | 12.1 | 11.9 | | 11.9 | 11.9 | | 11.9 | 12.1 | |
| Characteristic (in proportions) | T (%) | C (%) | StdDiff | T (%) | C (%) | StdDiff | T (%) | C (%) | StdDiff |
| Gender: female | 57.9 | 43.4 | 0.29 | 55.4 | 55.5 | 0.00 | 48.1 | 48.4 | -0.01 |
| **Medical history: General** | | | | | | | | | |
| Chronic liver disease | 0.7 | 0.7 | 0.00 | 0.7 | 0.7 | 0.00 | 0.7 | 0.7 | 0.00 |
| Chronic obstructive lung disease | 4.3 | 4.7 | -0.02 | 4.3 | 4.3 | 0.00 | 4.6 | 4.6 | 0.00 |
| Dementia | 0.4 | 0.4 | -0.01 | 0.4 | 0.3 | 0.00 | 0.4 | 0.4 | 0.00 |
| Gastroesophageal reflux disease | 9.4 | 8.4 | 0.04 | 9.0 | 9.1 | 0.00 | 8.9 | 8.8 | 0.00 |
| Hyperlipidemia | 39.7 | 43.8 | -0.08 | 40.1 | 40.0 | 0.00 | 42.6 | 42.4 | 0.01 |
| Hypertensive disorder | 46.6 | 48.6 | -0.04 | 46.4 | 46.3 | 0.00 | 48.2 | 47.9 | 0.00 |
| Obesity | 19.1 | 12.4 | 0.19 | 16.8 | 16.7 | 0.00 | 15.1 | 14.5 | 0.02 |
| Renal impairment | 4.5 | 5.4 | -0.04 | 4.5 | 4.4 | 0.00 | 5.1 | 5.0 | 0.00 |
| Urinary tract infectious disease | 4.7 | 3.5 | 0.06 | 4.3 | 4.3 | 0.00 | 4.0 | 3.9 | 0.01 |
| Acute urinary tract infection | 0.2 | 0.1 | 0.02 | 0.2 | 0.1 | 0.01 | 0.1 | 0.1 | 0.01 |
| Closed fracture of hip | 0.1 | 0.1 | -0.01 | 0.1 | 0.1 | 0.00 | 0.1 | 0.1 | 0.00 |
| Closed fracture of lower limb | 0.9 | 0.7 | 0.02 | 0.8 | 0.9 | 0.00 | 0.8 | 0.8 | 0.00 |
| **Medical history: Cardiovascular disease** | | | | | | | | | |
| Atrial fibrillation | 2.6 | 4.3 | -0.09 | 2.7 | 2.7 | 0.00 | 3.6 | 3.7 | -0.01 |
| Cerebrovascular disease | 1.9 | 2.8 | -0.06 | 2.0 | 1.9 | 0.00 | 2.5 | 2.5 | 0.00 |
| Coronary arteriosclerosis | 5.6 | 10.8 | -0.18 | 5.9 | 5.8 | 0.00 | 8.9 | 9.1 | 0.00 |
| Heart disease | 13.4 | 20.3 | -0.18 | 13.8 | 13.6 | 0.00 | 17.8 | 18.0 | -0.01 |
| Heart failure | 2.7 | 5.2 | -0.12 | 2.8 | 2.7 | 0.00 | 4.0 | 4.4 | -0.02 |
| Ischemic heart disease | 2.7 | 5.3 | -0.13 | 2.8 | 2.8 | 0.00 | 4.2 | 4.4 | -0.01 |
| Peripheral vascular disease | 3.7 | 4.9 | -0.06 | 3.7 | 3.8 | 0.00 | 4.5 | 4.5 | 0.00 |
| Pulmonary embolism | 0.5 | 0.5 | 0.01 | 0.5 | 0.5 | 0.00 | 0.5 | 0.5 | 0.01 |
| Venous thrombosis | 0.7 | 0.7 | -0.01 | 0.7 | 0.7 | 0.00 | 0.7 | 0.7 | 0.00 |
| Medical history: Neoplasms | 4.4 | 5.2 | -0.04 | 4.5 | 4.5 | 0.00 | 4.9 | 5.0 | 0.00 |
| **Medication use** | | | | | | | | | |
| Agents acting on the renin-angiotensin system | 62.3 | 68.3 | -0.13 | 63.6 | 63.6 | 0.00 | 66.2 | 66.4 | 0.00 |
| Antibacterials for systemic use | 57.4 | 52.5 | 0.10 | 56.0 | 56.3 | -0.01 | 54.3 | 54.3 | 0.00 |
| Antithrombotic agents | 14.2 | 21.0 | -0.18 | 14.6 | 14.4 | 0.00 | 18.4 | 18.7 | -0.01 |
| Beta blocking agents | 28.3 | 35.5 | -0.15 | 28.8 | 28.7 | 0.00 | 32.9 | 33.1 | 0.00 |
| Calcium channel blockers | 22.3 | 25.8 | -0.08 | 22.8 | 22.7 | 0.00 | 24.7 | 24.5 | 0.00 |
| Diuretics | 39.8 | 39.3 | 0.01 | 39.4 | 39.5 | 0.00 | 39.3 | 39.6 | 0.00 |
| Immunosuppressants | 3.2 | 2.8 | 0.03 | 3.1 | 3.2 | 0.00 | 2.9 | 3.0 | 0.00 |
| Lipid modifying agents | 65.7 | 74.9 | -0.20 | 67.7 | 67.4 | 0.01 | 71.8 | 71.8 | 0.00 |

##### 5.3.6 OptumEHR

**Supplementary Table 56:** Baseline patient characteristics for dulaglutide (T) and empagliflozin (C) new-users in the OptumEHR data source. We report proportion of initiators satisfying selected base-line characteristics and the standardized difference of population proportions (StdDiff) before and after propensity score adjustment. Less extreme StdDiffs through matching and stratification suggest improved balance between patient cohorts through adjustment. The "Medication use" entries refer to second-level ATC medication classes.

| | Before adjustment | | | After matching | | | After stratification | | |
|---|---|---|---|---|---|---|---|---|---|
| Characteristic | T | C | StdDiff | T | C | StdDiff | T | C | StdDiff |
| Age: mean | 53.7 | 57.8 | -0.35 | 54.9 | 54.9 | 0.00 | 56.2 | 56.4 | -0.02 |
| Age: std | 11.7 | 11.5 | | 11.6 | 11.3 | | 11.6 | 11.7 | |
| Characteristic (in proportions) | T (%) | C (%) | StdDiff | T (%) | C (%) | StdDiff | T (%) | C (%) | StdDiff |
| Gender: female | 58.4 | 42.4 | 0.32 | 53.5 | 55.9 | -0.05 | 47.0 | 49.2 | -0.04 |
| **Race** | | | | | | | | | |
| Asian | 1.7 | 3.0 | -0.08 | 1.9 | 1.9 | 0.00 | 2.4 | 2.5 | 0.00 |
| Black or African American | 12.6 | 10.8 | 0.06 | 11.9 | 12.0 | -0.01 | 11.2 | 11.4 | -0.01 |
| White | 78.2 | 78.3 | 0.00 | 78.8 | 78.4 | 0.01 | 78.7 | 78.4 | 0.01 |
| **Ethnicity** | | | | | | | | | |
| Hispanic or Latino | 6.5 | 6.0 | 0.02 | 6.5 | 6.6 | 0.00 | 6.3 | 6.3 | 0.00 |
| Not Hispanic or Latino | 84.8 | 83.5 | 0.04 | 84.3 | 84.5 | 0.00 | 84.0 | 83.9 | 0.00 |
| **Medical history: General** | | | | | | | | | |
| Chronic liver disease | 1.7 | 1.3 | 0.03 | 1.6 | 1.5 | 0.01 | 1.5 | 1.5 | 0.00 |
| Chronic obstructive lung disease | 4.3 | 5.1 | -0.04 | 4.5 | 4.4 | 0.01 | 4.9 | 4.9 | 0.00 |
| Dementia | 0.4 | 0.5 | -0.02 | 0.4 | 0.4 | 0.00 | 0.4 | 0.4 | 0.00 |
| Gastroesophageal reflux disease | 18.5 | 17.0 | 0.04 | 17.4 | 17.7 | -0.01 | 17.3 | 17.7 | -0.01 |
| Hyperlipidemia | 61.3 | 67.8 | -0.14 | 62.7 | 61.6 | 0.02 | 65.7 | 65.0 | 0.02 |
| Hypertensive disorder | 62.4 | 67.2 | -0.10 | 63.2 | 62.5 | 0.02 | 66.0 | 65.2 | 0.02 |
| Obesity | 43.7 | 33.0 | 0.22 | 38.9 | 40.5 | -0.03 | 36.6 | 37.4 | -0.01 |
| Renal impairment | 5.2 | 6.0 | -0.04 | 5.4 | 5.0 | 0.02 | 6.0 | 5.6 | 0.02 |
| Urinary tract infectious disease | 4.2 | 3.0 | 0.07 | 3.7 | 3.8 | -0.01 | 3.3 | 3.4 | -0.01 |
| Primary osteoporosis | 0.1 | 0.1 | 0.00 | 0.1 | 0.1 | 0.01 | 0.1 | 0.1 | 0.01 |
| Acute urinary tract infection | 0.6 | 0.4 | 0.02 | 0.5 | 0.5 | -0.01 | 0.4 | 0.5 | 0.00 |
| Closed fracture of hip | 0.0 | 0.1 | 0.00 | 0.1 | 0.1 | 0.00 | 0.0 | 0.1 | 0.00 |
| Closed fracture of lower limb | 0.8 | 0.6 | 0.03 | 0.8 | 0.7 | 0.01 | 0.6 | 0.6 | 0.00 |
| **Medical history: Cardiovascular disease** | | | | | | | | | |
| Atrial fibrillation | 2.8 | 4.9 | -0.10 | 3.1 | 2.8 | 0.02 | 4.3 | 4.0 | 0.02 |
| Cerebrovascular disease | 1.9 | 3.0 | -0.07 | 2.1 | 2.0 | 0.01 | 2.6 | 2.6 | 0.00 |
| Coronary arteriosclerosis | 7.2 | 15.2 | -0.24 | 8.3 | 7.3 | 0.04 | 12.7 | 12.0 | 0.02 |
| Heart disease | 16.5 | 25.6 | -0.22 | 17.6 | 16.2 | 0.04 | 23.0 | 21.8 | 0.03 |
| Heart failure | 2.3 | 4.9 | -0.13 | 2.6 | 2.4 | 0.01 | 3.7 | 4.0 | -0.01 |
| Ischemic heart disease | 3.4 | 6.8 | -0.15 | 3.8 | 3.6 | 0.01 | 5.6 | 5.5 | 0.00 |
| Peripheral vascular disease | 4.3 | 6.8 | -0.11 | 4.7 | 4.5 | 0.01 | 5.8 | 5.9 | 0.00 |
| Pulmonary embolism | 0.7 | 0.5 | 0.02 | 0.6 | 0.5 | 0.02 | 0.6 | 0.5 | 0.01 |
| Venous thrombosis | 0.8 | 0.9 | -0.01 | 0.8 | 0.9 | 0.00 | 0.8 | 0.9 | -0.01 |
| Chronic urinary tract infection | 0.1 | 0.1 | 0.01 | 0.1 | 0.1 | -0.01 | 0.1 | 0.1 | -0.01 |
| Medical history: Neoplasms | 5.3 | 5.9 | -0.02 | 5.5 | 5.1 | 0.02 | 5.9 | 5.6 | 0.01 |
| **Medication use** | | | | | | | | | |
| Agents acting on the renin-angiotensin system | 61.2 | 66.5 | -0.11 | 63.1 | 62.4 | 0.01 | 64.7 | 64.4 | 0.01 |
| Antibacterials for systemic use | 44.0 | 40.5 | 0.07 | 42.0 | 42.9 | -0.02 | 41.1 | 41.9 | -0.02 |
| Antithrombotic agents | 31.7 | 39.4 | -0.16 | 33.1 | 32.1 | 0.02 | 36.7 | 36.3 | 0.01 |
| Beta blocking agents | 27.4 | 34.7 | -0.16 | 28.8 | 27.6 | 0.03 | 32.0 | 31.9 | 0.00 |
| Calcium channel blockers | 20.5 | 24.3 | -0.09 | 21.5 | 21.1 | 0.01 | 22.7 | 22.8 | 0.00 |
| Diuretics | 38.9 | 38.0 | 0.02 | 38.6 | 38.5 | 0.00 | 38.4 | 38.4 | 0.00 |
| Immunosuppressants | 3.6 | 2.9 | 0.04 | 3.3 | 3.1 | 0.01 | 3.2 | 3.1 | 0.01 |
| Lipid modifying agents | 64.2 | 74.5 | -0.23 | 68.0 | 66.5 | 0.03 | 71.3 | 70.4 | 0.02 |

#### 5.4 semaglutide vs dapagliflozin

##### 5.4.1 CCAE

**Supplementary Table 57:** Baseline patient characteristics for semaglutide (T) and dapagliflozin (C) new-users in the CCAE data source. We report proportion of initiators satisfying selected base-line characteristics and the standardized difference of population proportions (StdDiff) before and after propensity score adjustment. Less extreme StdDiffs through matching and stratification suggest improved balance between patient cohorts through adjustment. The ”Medication use” entries refer to second-level ATC medication classes.

| | Before adjustment | | | After matching | | | After stratification | | |
|---|---|---|---|---|---|---|---|---|---|
| Characteristic | T | C | StdDiff | T | C | StdDiff | T | C | StdDiff |
| Age: mean | 50.9 | 52.5 | -0.17 | 52.3 | 52.5 | -0.03 | 51.5 | 51.8 | -0.03 |
| Age: std | 9.2 | 8.7 | | 8.5 | 8.5 | | 8.5 | 8.9 | |
| Characteristic (in proportions) | T (%) | C (%) | StdDiff | T (%) | C (%) | StdDiff | T (%) | C (%) | StdDiff |
| Gender: female | 61.1 | 43.7 | 0.35 | 43.9 | 45.7 | -0.04 | 53.9 | 55.0 | -0.02 |
| **Medical history: General** | | | | | | | | | |
| Chronic liver disease | 0.6 | 1.4 | -0.08 | 0.6 | 0.6 | -0.01 | 0.6 | 0.7 | -0.01 |
| Chronic obstructive lung disease | 2.2 | 2.4 | -0.01 | 2.1 | 2.3 | -0.01 | 2.2 | 2.6 | -0.02 |
| Dementia | 0.2 | 0.1 | 0.01 | 0.1 | 0.1 | 0.00 | 0.1 | 0.2 | 0.00 |
| Gastroesophageal reflux disease | 17.2 | 13.3 | 0.11 | 13.7 | 14.4 | -0.02 | 16.1 | 16.6 | -0.01 |
| Hyperlipidemia | 67.6 | 73.3 | -0.12 | 73.0 | 73.1 | 0.00 | 69.9 | 71.0 | -0.02 |
| Hypertensive disorder | 67.2 | 71.2 | -0.09 | 71.2 | 70.9 | 0.00 | 68.9 | 69.0 | 0.00 |
| Obesity | 48.9 | 28.3 | 0.42 | 32.5 | 34.4 | -0.04 | 42.5 | 44.4 | -0.04 |
| Renal impairment | 3.8 | 4.7 | -0.04 | 5.5 | 5.7 | -0.01 | 4.8 | 5.1 | -0.01 |
| Urinary tract infectious disease | 6.6 | 5.9 | 0.03 | 5.0 | 5.6 | -0.03 | 6.1 | 6.3 | -0.01 |
| Acute urinary tract infection | 0.3 | 0.2 | 0.03 | 0.3 | 0.2 | 0.01 | 0.3 | 0.2 | 0.01 |
| Closed fracture of lower limb | 1.1 | 1.0 | 0.01 | 1.0 | 0.9 | 0.01 | 1.1 | 0.8 | 0.03 |
| **Medical history: Cardiovascular disease** | | | | | | | | | |
| Atrial fibrillation | 1.9 | 2.3 | -0.03 | 2.3 | 2.4 | -0.01 | 2.2 | 2.5 | -0.02 |
| Cerebrovascular disease | 1.8 | 2.3 | -0.03 | 2.0 | 2.2 | -0.01 | 1.9 | 2.1 | -0.01 |
| Coronary arteriosclerosis | 5.2 | 7.3 | -0.09 | 7.2 | 7.0 | 0.01 | 6.3 | 6.2 | 0.00 |
| Heart disease | 15.2 | 17.3 | -0.06 | 16.8 | 17.2 | -0.01 | 16.6 | 16.4 | 0.00 |
| Heart failure | 1.8 | 3.2 | -0.09 | 2.6 | 2.8 | -0.01 | 2.6 | 2.7 | -0.01 |
| Ischemic heart disease | 2.9 | 4.2 | -0.07 | 3.6 | 3.9 | -0.02 | 3.6 | 3.9 | -0.02 |
| Peripheral vascular disease | 4.0 | 3.8 | 0.01 | 4.3 | 4.4 | 0.00 | 4.3 | 4.0 | 0.01 |
| Pulmonary embolism | 0.7 | 0.4 | 0.04 | 0.6 | 0.5 | 0.02 | 0.7 | 0.4 | 0.03 |
| Venous thrombosis | 1.0 | 0.9 | 0.01 | 0.9 | 0.9 | 0.00 | 1.0 | 0.8 | 0.02 |
| Medical history: Neoplasms | 5.5 | 5.2 | 0.01 | 5.3 | 5.5 | -0.01 | 5.6 | 6.1 | -0.02 |
| **Medication use** | | | | | | | | | |
| Agents acting on the renin-angiotensin system | 58.8 | 67.4 | -0.18 | 66.4 | 65.9 | 0.01 | 61.9 | 62.9 | -0.02 |
| Antibacterials for systemic use | 57.7 | 56.2 | 0.03 | 53.2 | 54.3 | -0.02 | 56.3 | 57.9 | -0.03 |
| Antithrombotic agents | 9.2 | 10.0 | -0.03 | 9.9 | 10.2 | -0.01 | 10.0 | 9.7 | 0.01 |
| Beta blocking agents | 23.2 | 25.5 | -0.05 | 24.5 | 25.1 | -0.01 | 24.4 | 25.6 | -0.03 |
| Calcium channel blockers | 20.4 | 21.6 | -0.03 | 22.7 | 22.6 | 0.00 | 21.4 | 21.5 | 0.00 |
| Diuretics | 39.5 | 37.8 | 0.04 | 37.6 | 37.4 | 0.00 | 38.8 | 39.3 | -0.01 |
| Immunosuppressants | 3.9 | 2.3 | 0.09 | 3.1 | 2.7 | 0.02 | 3.5 | 3.5 | 0.00 |
| Lipid modifying agents | 60.9 | 68.2 | -0.15 | 69.4 | 69.5 | 0.00 | 64.6 | 65.7 | -0.02 |

##### 5.4.2 OptumDOD

**Supplementary Table 58:** Baseline patient characteristics for semaglutide (T) and dapagliflozin (C) new-users in the OptumDOD data source. We report proportion of initiators satisfying selected base-line characteristics and the standardized difference of population proportions (StdDiff) before and after propensity score adjustment. Less extreme StdDiffs through matching and stratification suggest improved balance between patient cohorts through adjustment. The ”Medication use” entries refer to second-level ATC medication classes.

| | Before adjustment | | | After matching | | | After stratification | | |
|---|---|---|---|---|---|---|---|---|---|
| Characteristic | T | C | StdDiff | T | C | StdDiff | T | C | StdDiff |
| Age: mean | 58.3 | 67.2 | -0.71 | 67.7 | 67.8 | -0.01 | 61.2 | 61.8 | -0.05 |
| Age: std | 12.1 | 10.1 | | 11.4 | 9.7 | | 11.4 | 11.9 | |
| Characteristic (in proportions) | T (%) | C (%) | StdDiff | T (%) | C (%) | StdDiff | T (%) | C (%) | StdDiff |
| Gender: female | 60.3 | 45.4 | 0.30 | 51.6 | 51.2 | 0.01 | 56.7 | 54.9 | 0.04 |
| **Medical history: General** | | | | | | | | | |
| Chronic liver disease | 1.1 | 2.3 | -0.10 | 1.2 | 1.6 | -0.04 | 1.2 | 1.3 | -0.01 |
| Chronic obstructive lung disease | 8.1 | 14.6 | -0.22 | 13.4 | 12.9 | 0.01 | 10.0 | 10.8 | -0.02 |
| Dementia | 0.7 | 2.6 | -0.17 | 1.5 | 2.0 | -0.04 | 1.1 | 1.4 | -0.03 |
| Gastroesophageal reflux disease | 23.4 | 23.6 | -0.01 | 24.8 | 24.8 | 0.00 | 24.0 | 25.3 | -0.03 |
| Hyperlipidemia | 77.5 | 84.7 | -0.18 | 85.1 | 83.3 | 0.05 | 79.8 | 79.7 | 0.00 |
| Hypertensive disorder | 76.3 | 85.5 | -0.23 | 85.8 | 85.2 | 0.02 | 79.4 | 79.2 | 0.00 |
| Obesity | 53.6 | 29.6 | 0.48 | 32.8 | 34.1 | -0.03 | 47.3 | 47.4 | 0.00 |
| Renal impairment | 11.0 | 30.3 | -0.52 | 25.5 | 23.7 | 0.04 | 17.7 | 16.9 | 0.02 |
| Urinary tract infectious disease | 8.8 | 10.0 | -0.04 | 9.5 | 9.3 | 0.00 | 9.1 | 9.6 | -0.02 |
| Acute urinary tract infection | 0.3 | 0.2 | 0.03 | 0.3 | 0.1 | 0.04 | 0.3 | 0.1 | 0.04 |
| Closed fracture of hip | 0.1 | 0.4 | -0.06 | 0.3 | 0.3 | -0.01 | 0.2 | 0.2 | 0.00 |
| Closed fracture of lower limb | 1.5 | 1.6 | 0.00 | 1.6 | 1.8 | -0.01 | 1.5 | 2.4 | -0.07 |
| **Medical history: Cardiovascular disease** | | | | | | | | | |
| Atrial fibrillation | 4.9 | 14.1 | -0.35 | 9.7 | 9.3 | 0.01 | 7.2 | 7.4 | -0.01 |
| Cerebrovascular disease | 4.5 | 9.5 | -0.21 | 8.8 | 8.2 | 0.02 | 6.4 | 5.9 | 0.02 |
| Coronary arteriosclerosis | 12.2 | 27.5 | -0.41 | 23.9 | 21.5 | 0.06 | 16.9 | 17.5 | -0.02 |
| Heart disease | 27.1 | 46.9 | -0.42 | 43.8 | 40.8 | 0.06 | 33.8 | 33.7 | 0.00 |
| Heart failure | 5.9 | 21.9 | -0.52 | 13.8 | 12.3 | 0.05 | 10.2 | 10.4 | -0.01 |
| Ischemic heart disease | 6.3 | 15.7 | -0.33 | 12.1 | 11.0 | 0.04 | 8.9 | 9.0 | 0.00 |
| Peripheral vascular disease | 10.2 | 19.1 | -0.27 | 18.3 | 17.4 | 0.02 | 13.2 | 13.0 | 0.01 |
| Pulmonary embolism | 1.1 | 1.3 | -0.01 | 1.6 | 1.1 | 0.04 | 1.3 | 0.8 | 0.05 |
| Venous thrombosis | 1.2 | 2.1 | -0.08 | 1.9 | 1.7 | 0.01 | 1.4 | 1.4 | 0.00 |
| Medical history: Neoplasms | 8.6 | 12.3 | -0.13 | 11.9 | 11.8 | 0.00 | 9.8 | 8.8 | 0.03 |
| **Medication use** | | | | | | | | | |
| Agents acting on the renin-angiotensin system | 65.2 | 78.7 | -0.29 | 77.0 | 76.8 | 0.01 | 77.0 | 76.8 | 0.01 |
| Antibacterials for systemic use | 58.8 | 55.9 | 0.06 | 54.4 | 55.0 | -0.01 | 57.8 | 58.3 | -0.01 |
| Antithrombotic agents | 14.2 | 27.0 | -0.33 | 23.0 | 21.5 | 0.04 | 17.8 | 16.5 | 0.04 |
| Beta blocking agents | 31.2 | 48.8 | -0.36 | 45.8 | 43.9 | 0.04 | 36.7 | 36.1 | 0.01 |
| Calcium channel blockers | 25.4 | 34.3 | -0.20 | 35.1 | 34.2 | 0.02 | 28.7 | 30.1 | -0.03 |
| Diuretics | 45.0 | 52.8 | -0.16 | 51.8 | 49.4 | 0.05 | 47.6 | 48.1 | -0.01 |
| Immunosuppressants | 4.2 | 3.3 | 0.04 | 3.6 | 3.8 | -0.01 | 4.0 | 4.5 | -0.02 |
| Lipid modifying agents | 71.1 | 83.8 | -0.29 | 86.6 | 84.7 | 0.05 | 75.5 | 75.3 | 0.00 |

##### 5.4.3 MDCR

**Supplementary Table 59:** Baseline patient characteristics for semaglutide (T) and dapagliflozin (C) new-users in the MDCR data source. We report proportion of initiators satisfying selected base-line characteristics and the standardized difference of population proportions (StdDiff) before and after propensity score adjustment. Less extreme StdDiffs through matching and stratification suggest improved balance between patient cohorts through adjustment. The ”Medication use” entries refer to second-level ATC medication classes.

| | Before adjustment | | | After matching | | | After stratification | | |
|---|---|---|---|---|---|---|---|---|---|
| Characteristic | T | C | StdDiff | T | C | StdDiff | T | C | StdDiff |
| Age: mean | 71.2 | 72.8 | -0.28 | 71.9 | 72.5 | -0.11 | 71.8 | 72.0 | -0.04 |
| Age: std | 5.0 | 5.5 | | 5.9 | 5.7 | | 5.9 | 5.6 | |
| Characteristic (in proportions) | T (%) | C (%) | StdDiff | T (%) | C (%) | StdDiff | T (%) | C (%) | StdDiff |
| Gender: female | 60.0 | 42.5 | 0.35 | 46.5 | 50.3 | -0.08 | 51.1 | 58.7 | -0.15 |
| **Medical history: General** | | | | | | | | | |
| Chronic liver disease | 1.6 | 1.8 | -0.01 | 1.4 | 1.5 | -0.01 | 1.5 | 1.6 | -0.01 |
| Chronic obstructive lung disease | 9.1 | 11.5 | -0.08 | 9.5 | 10.4 | -0.03 | 9.3 | 10.6 | -0.04 |
| Dementia | 1.0 | 1.9 | -0.07 | 1.4 | 1.8 | -0.03 | 1.3 | 2.4 | -0.09 |
| Gastroesophageal reflux disease | 21.7 | 17.0 | 0.12 | 17.7 | 19.8 | -0.06 | 18.9 | 22.8 | -0.10 |
| Hyperlipidemia | 83.1 | 82.5 | 0.01 | 81.5 | 84.2 | -0.07 | 83.7 | 85.9 | -0.06 |
| Hypertensive disorder | 85.2 | 85.8 | -0.02 | 85.8 | 85.9 | 0.00 | 86.2 | 89.3 | -0.09 |
| Obesity | 48.6 | 23.4 | 0.52 | 25.3 | 33.3 | -0.17 | 37.8 | 43.4 | -0.11 |
| Renal impairment | 16.6 | 22.6 | -0.15 | 21.4 | 19.6 | 0.04 | 21.9 | 19.3 | 0.06 |
| Urinary tract infectious disease | 9.0 | 8.1 | 0.03 | 9.1 | 8.0 | 0.04 | 9.2 | 8.7 | 0.02 |
| Acute urinary tract infection | <0.4 | <0.4 | 0.01 | <1.0 | <0.4 | 0.07 | 0.4 | <0.5 | 0.06 |
| Closed fracture of hip | <0.4 | <0.4 | -0.03 | <0.3 | <0.5 | -0.03 | <0.3 | <0.5 | -0.03 |
| Closed fracture of lower limb | 1.7 | 1.2 | 0.04 | <1.0 | 1.6 | -0.10 | 1.7 | 1.8 | -0.01 |
| **Medical history: Cardiovascular disease** | | | | | | | | | |
| Atrial fibrillation | 9.9 | 14.0 | -0.13 | 12.1 | 10.4 | 0.05 | 12.3 | 10.7 | 0.05 |
| Cerebrovascular disease | 8.5 | 13.0 | -0.14 | 11.5 | 10.8 | 0.02 | 10.2 | 11.6 | -0.05 |
| Coronary arteriosclerosis | 20.6 | 26.4 | -0.14 | 25.1 | 19.7 | 0.13 | 23.4 | 18.2 | 0.12 |
| Heart disease | 39.5 | 49.1 | -0.19 | 46.5 | 43.3 | 0.06 | 43.9 | 42.9 | 0.02 |
| Heart failure | 7.6 | 15.4 | -0.25 | 10.1 | 9.3 | 0.03 | 10.0 | 10.7 | -0.02 |
| Ischemic heart disease | 8.4 | 12.2 | -0.12 | 11.3 | 8.2 | 0.10 | 10.2 | 8.3 | 0.06 |
| Peripheral vascular disease | 14.3 | 15.9 | -0.04 | 17.1 | 14.7 | 0.06 | 16.0 | 12.8 | 0.09 |
| Pulmonary embolism | 0.7 | 1.1 | -0.04 | <1.0 | 1.4 | -0.06 | 0.7 | 2.3 | -0.17 |
| Venous thrombosis | 1.2 | 2.6 | -0.10 | 1.4 | 2.1 | -0.06 | 1.4 | 2.0 | -0.05 |
| Medical history: Neoplasms | 16.6 | 18.3 | -0.04 | 16.9 | 17.5 | -0.02 | 16.7 | 15.6 | 0.03 |
| **Medication use** | | | | | | | | | |
| Agents acting on the renin-angiotensin system | 74.4 | 76.8 | -0.06 | 75.5 | 76.0 | -0.01 | 75.7 | 76.8 | -0.03 |
| Antibacterials for systemic use | 59.5 | 60.2 | -0.01 | 57.6 | 57.3 | 0.01 | 58.2 | 60.9 | -0.05 |
| Antithrombotic agents | 20.2 | 26.8 | -0.16 | 22.8 | 22.5 | 0.01 | 22.6 | 24.0 | -0.03 |
| Beta blocking agents | 44.3 | 51.1 | -0.14 | 51.4 | 49.5 | 0.04 | 49.0 | 49.7 | -0.01 |
| Calcium channel blockers | 34.1 | 34.7 | -0.01 | 33.9 | 35.3 | -0.03 | 35.2 | 34.0 | 0.03 |
| Diuretics | 52.2 | 51.6 | 0.01 | 49.8 | 49.3 | 0.01 | 51.7 | 52.7 | -0.02 |
| Immunosuppressants | 5.3 | 4.2 | 0.05 | 4.5 | 4.4 | 0.00 | 4.4 | 5.2 | -0.04 |
| Lipid modifying agents | 84.8 | 85.3 | -0.01 | 85.8 | 85.3 | 0.01 | 85.0 | 83.1 | 0.06 |

##### 5.4.4 Open Claims

**Supplementary Table 60:** Baseline patient characteristics for semaglutide (T) and dapagliflozin (C) new-users in the Open Claims data source. We report proportion of initiators satisfying selected base-line characteristics and the standardized difference of population proportions (StdDiff) before and after propensity score adjustment. Less extreme StdDiffs through matching and stratification suggest improved balance between patient cohorts through adjustment. The "Medication use" entries refer to second-level ATC medication classes.

| | Before adjustment | | | After matching | | | After stratification | | |
|---|---|---|---|---|---|---|---|---|---|
| Characteristic | T | C | StdDiff | T | C | StdDiff | T | C | StdDiff |
| Age: mean | 55.3 | 57.8 | -0.20 | 58.1 | 58.1 | 0.00 | 56.9 | 57.0 | -0.01 |
| Age: std | 12.1 | 11.9 | | 11.8 | 11.9 | | 11.8 | 12.2 | |
| Characteristic (in proportions) | T (%) | C (%) | StdDiff | T (%) | C (%) | StdDiff | T (%) | C (%) | StdDiff |
| Gender: female | 61.3 | 45.9 | 0.31 | 47.8 | 48.8 | -0.02 | 54.4 | 54.8 | -0.01 |
| **Medical history: General** | | | | | | | | | |
| Chronic liver disease | 0.5 | 1.0 | -0.06 | 0.5 | 0.5 | 0.00 | 0.6 | 0.6 | 0.00 |
| Chronic obstructive lung disease | 3.7 | 4.2 | -0.03 | 4.0 | 4.1 | 0.00 | 4.2 | 4.4 | -0.01 |
| Dementia | 0.3 | 0.4 | -0.03 | 0.3 | 0.4 | 0.00 | 0.3 | 0.4 | 0.00 |
| Gastroesophageal reflux disease | 9.8 | 8.0 | 0.06 | 8.3 | 8.2 | 0.00 | 9.2 | 9.2 | 0.00 |
| Hyperlipidemia | 39.2 | 44.4 | -0.10 | 41.2 | 41.0 | 0.00 | 41.1 | 41.3 | 0.00 |
| Hypertensive disorder | 46.0 | 49.3 | -0.07 | 46.8 | 46.6 | 0.00 | 47.4 | 47.8 | -0.01 |
| Obesity | 21.2 | 12.3 | 0.24 | 12.8 | 13.0 | 0.00 | 17.7 | 17.8 | 0.00 |
| Renal impairment | 4.3 | 6.0 | -0.08 | 5.8 | 5.8 | 0.00 | 5.9 | 6.0 | -0.01 |
| Urinary tract infectious disease | 4.5 | 4.0 | 0.03 | 3.8 | 3.7 | 0.00 | 4.3 | 4.4 | 0.00 |
| Acute urinary tract infection | 0.1 | 0.1 | 0.01 | 0.1 | 0.1 | 0.00 | 0.1 | 0.1 | -0.01 |
| Closed fracture of hip | 0.1 | 0.1 | -0.01 | 0.1 | 0.1 | 0.00 | 0.1 | 0.1 | 0.00 |
| Closed fracture of lower limb | 0.8 | 0.7 | 0.01 | 0.7 | 0.7 | -0.01 | 0.8 | 0.8 | -0.01 |
| **Medical history: Cardiovascular disease** | | | | | | | | | |
| Atrial fibrillation | 2.5 | 3.9 | -0.07 | 3.2 | 3.2 | 0.00 | 3.3 | 3.5 | -0.01 |
| Cerebrovascular disease | 1.7 | 2.4 | -0.05 | 2.1 | 2.1 | 0.00 | 2.1 | 2.2 | -0.01 |
| Coronary arteriosclerosis | 5.2 | 8.2 | -0.12 | 6.9 | 7.0 | 0.00 | 6.8 | 7.2 | -0.01 |
| Heart disease | 13.1 | 17.8 | -0.13 | 15.5 | 15.6 | 0.00 | 15.8 | 16.5 | -0.02 |
| Heart failure | 2.5 | 5.6 | -0.16 | 3.5 | 3.6 | -0.01 | 3.9 | 4.8 | -0.04 |
| Ischemic heart disease | 2.5 | 4.2 | -0.10 | 3.3 | 3.4 | 0.00 | 3.4 | 3.6 | -0.01 |
| Peripheral vascular disease | 3.6 | 4.1 | -0.02 | 4.0 | 4.1 | 0.00 | 4.1 | 4.3 | -0.01 |
| Pulmonary embolism | 0.5 | 0.4 | 0.01 | 0.5 | 0.4 | 0.01 | 0.5 | 0.5 | 0.00 |
| Venous thrombosis | 0.6 | 0.7 | -0.01 | 0.6 | 0.6 | 0.00 | 0.6 | 0.6 | 0.00 |
| Medical history: Neoplasms | 4.4 | 4.8 | -0.02 | 4.6 | 4.7 | 0.00 | 4.7 | 4.8 | 0.00 |
| **Medication use** | | | | | | | | | |
| Agents acting on the renin-angiotensin system | 60.7 | 69.2 | -0.18 | 67.6 | 67.0 | 0.01 | 64.5 | 65.0 | -0.01 |
| Antibacterials for systemic use | 57.5 | 54.9 | 0.05 | 53.2 | 53.2 | 0.00 | 55.8 | 56.4 | -0.01 |
| Antithrombotic agents | 13.5 | 17.4 | -0.11 | 16.6 | 16.8 | 0.00 | 16.2 | 16.6 | -0.01 |
| Beta blocking agents | 28.6 | 33.2 | -0.10 | 32.2 | 32.3 | 0.00 | 31.7 | 32.3 | -0.01 |
| Calcium channel blockers | 23.1 | 24.5 | -0.03 | 26.2 | 25.9 | 0.01 | 24.9 | 24.7 | 0.00 |
| Diuretics | 41.3 | 41.1 | 0.00 | 40.5 | 40.5 | 0.00 | 41.7 | 42.4 | -0.01 |
| Immunosuppressants | 3.6 | 2.6 | 0.06 | 2.8 | 2.9 | 0.00 | 3.3 | 3.3 | 0.00 |
| Lipid modifying agents | 64.5 | 71.0 | -0.14 | 72.7 | 72.0 | 0.01 | 68.6 | 69.0 | -0.01 |

##### 5.4.5 OptumEHR

**Supplementary Table 61:** Baseline patient characteristics for semaglutide (T) and dapagliflozin (C) new-users in the OptumEHR data source. We report proportion of initiators satisfying selected base-line characteristics and the standardized difference of population proportions (StdDiff) before and after propensity score adjustment. Less extreme StdDiffs through matching and stratification suggest improved balance between patient cohorts through adjustment. The ”Medication use” entries refer to second-level ATC medication classes.

| | Before adjustment | | | After matching | | | After stratification | | |
|---|---|---|---|---|---|---|---|---|---|
| Characteristic | T | C | StdDiff | T | C | StdDiff | T | C | StdDiff |
| Age: mean | 53.4 | 55.7 | -0.20 | 56.0 | 56.3 | -0.03 | 54.7 | 55.0 | -0.03 |
| Age: std | 11.9 | 11.7 | | 11.4 | 11.7 | | 11.4 | 11.7 | |
| Characteristic (in proportions) | T (%) | C (%) | StdDiff | T (%) | C (%) | StdDiff | T (%) | C (%) | StdDiff |
| Gender: female | 62.6 | 46.0 | 0.33 | 47.7 | 50.0 | -0.05 | 53.5 | 56.8 | -0.07 |
| **Race** | | | | | | | | | |
| Asian | 1.9 | 2.1 | -0.01 | 1.8 | 2.4 | -0.04 | 2.4 | 1.9 | 0.04 |
| Black or African American | 13.9 | 12.3 | 0.05 | 13.1 | 13.4 | -0.01 | 13.4 | 13.2 | 0.01 |
| White | 77.1 | 78.4 | -0.03 | 77.0 | 76.2 | 0.02 | 76.4 | 77.9 | -0.04 |
| **Ethnicity** | | | | | | | | | |
| Hispanic or Latino | 5.6 | 6.1 | -0.02 | 6.1 | 6.1 | 0.00 | 5.7 | 5.9 | -0.01 |
| Not Hispanic or Latino | 84.6 | 85.9 | -0.04 | 83.4 | 83.8 | -0.01 | 83.9 | 85.0 | -0.03 |
| **Medical history: General** | | | | | | | | | |
| Chronic liver disease | 1.1 | 1.7 | -0.05 | 0.9 | 1.0 | -0.01 | 1.0 | 1.1 | -0.01 |
| Chronic obstructive lung disease | 4.0 | 4.5 | -0.03 | 4.6 | 4.7 | 0.00 | 4.3 | 4.6 | -0.01 |
| Dementia | 0.3 | 0.3 | 0.00 | 0.5 | 0.2 | 0.04 | 0.5 | 0.3 | 0.04 |
| Gastroesophageal reflux disease | 20.8 | 17.2 | 0.09 | 17.9 | 18.0 | 0.00 | 19.4 | 19.5 | 0.00 |
| Hyperlipidemia | 62.3 | 65.9 | -0.08 | 64.3 | 64.3 | 0.00 | 63.7 | 63.4 | 0.01 |
| Hypertensive disorder | 63.6 | 67.6 | -0.09 | 66.5 | 66.3 | 0.00 | 65.0 | 65.9 | -0.02 |
| Obesity | 51.6 | 32.6 | 0.38 | 34.5 | 37.2 | -0.06 | 42.9 | 46.0 | -0.06 |
| Renal impairment | 5.4 | 5.4 | 0.00 | 6.6 | 6.7 | -0.01 | 6.6 | 6.4 | 0.01 |
| Urinary tract infectious disease | 4.3 | 3.5 | 0.04 | 3.6 | 3.3 | 0.02 | 3.8 | 3.8 | 0.00 |
| Primary osteoporosis | 0.1 | 0.1 | 0.00 | 0.2 | 0.2 | 0.00 | 0.1 | 0.2 | -0.01 |
| Acute urinary tract infection | 1.1 | 0.4 | 0.08 | 0.9 | 0.6 | 0.04 | 0.9 | 0.6 | 0.04 |
| Closed fracture of hip | 0.0 | 0.0 | 0.00 | <0.1 | 0.0 | 0.00 | <0.1 | 0.0 | 0.00 |
| Closed fracture of lower limb | 0.8 | 0.7 | 0.01 | 0.7 | 0.8 | -0.01 | 0.9 | 0.8 | 0.01 |
| **Medical history: Cardiovascular disease** | | | | | | | | | |
| Atrial fibrillation | 3.0 | 3.8 | -0.04 | 4.2 | 3.7 | 0.03 | 4.2 | 3.5 | 0.04 |
| Cerebrovascular disease | 2.1 | 2.1 | 0.00 | 2.5 | 2.3 | 0.01 | 2.4 | 2.3 | 0.01 |
| Coronary arteriosclerosis | 7.1 | 10.9 | -0.13 | 10.3 | 9.9 | 0.01 | 10.0 | 8.3 | 0.06 |
| Heart disease | 17.2 | 20.2 | -0.08 | 21.1 | 20.0 | 0.03 | 20.9 | 18.7 | 0.06 |
| Heart failure | 2.5 | 4.9 | -0.13 | 3.9 | 3.9 | 0.00 | 4.5 | 3.7 | 0.04 |
| Ischemic heart disease | 3.8 | 5.3 | -0.07 | 4.9 | 4.8 | 0.00 | 4.8 | 4.2 | 0.03 |
| Peripheral vascular disease | 5.6 | 5.6 | 0.00 | 6.0 | 6.4 | -0.01 | 6.1 | 6.3 | -0.01 |
| Pulmonary embolism | 0.7 | 0.6 | 0.02 | 0.5 | 0.5 | 0.00 | 0.6 | 0.5 | 0.01 |
| Venous thrombosis | 0.9 | 0.9 | 0.00 | 0.8 | 0.9 | -0.02 | 0.9 | 1.0 | -0.01 |
| Chronic urinary tract infection | 0.1 | 0.1 | 0.03 | 0.1 | 0.1 | -0.01 | 0.1 | 0.1 | -0.01 |
| Medical history: Neoplasms | 5.1 | 4.9 | 0.01 | 5.3 | 5.8 | -0.02 | 5.2 | 5.2 | 0.00 |
| **Medication use** | | | | | | | | | |
| Agents acting on the renin-angiotensin system | 58.4 | 67.1 | -0.18 | 66.0 | 64.7 | 0.03 | 63.1 | 62.7 | 0.01 |
| Antibacterials for systemic use | 46.3 | 42.9 | 0.07 | 41.3 | 42.7 | -0.03 | 43.9 | 45.8 | -0.04 |
| Antithrombotic agents | 27.7 | 34.1 | -0.14 | 32.4 | 31.6 | 0.02 | 32.1 | 30.6 | 0.03 |
| Beta blocking agents | 28.5 | 31.9 | -0.07 | 32.4 | 31.9 | 0.01 | 33.2 | 30.7 | 0.06 |
| Calcium channel blockers | 22.3 | 22.4 | 0.00 | 24.6 | 24.1 | 0.01 | 23.4 | 22.0 | 0.03 |
| Diuretics | 40.5 | 39.3 | 0.03 | 38.8 | 39.0 | 0.00 | 40.7 | 41.2 | -0.01 |
| Immunosuppressants | 3.7 | 2.5 | 0.07 | 2.9 | 3.1 | -0.01 | 3.1 | 3.0 | 0.00 |
| Lipid modifying agents | 63.0 | 68.1 | -0.11 | 70.1 | 69.4 | 0.01 | 67.4 | 65.9 | 0.03 |

#### 5.5 semaglutide vs empagliflozin

##### 5.5.1 CCAE

**Supplementary Table 62:** Baseline patient characteristics for semaglutide (T) and empagliflozin (C) new-users in the CCAE data source. We report proportion of initiators satisfying selected base-line characteristics and the standardized difference of population proportions (StdDiff) before and after propensity score adjustment. Less extreme StdDiffs through matching and stratification suggest improved balance between patient cohorts through adjustment. The "Medication use" entries refer to second-level ATC medication classes.

| | Before adjustment | | | After matching | | | After stratification | | |
|---|---|---|---|---|---|---|---|---|---|
| Characteristic | T | C | StdDiff | T | C | StdDiff | T | C | StdDiff |
| Age: mean | 50.9 | 53.2 | -0.26 | 52.2 | 52.5 | -0.03 | 51.9 | 52.2 | -0.03 |
| Age: std | 9.2 | 8.7 | | 8.3 | 8.5 | | 8.3 | 8.7 | |
| Characteristic (in proportions) | T (%) | C (%) | StdDiff | T (%) | C (%) | StdDiff | T (%) | C (%) | StdDiff |
| Gender: female | 61.1 | 40.3 | 0.42 | 47.5 | 48.7 | -0.03 | 50.0 | 50.9 | -0.02 |
| **Medical history: General** | | | | | | | | | |
| Chronic liver disease | 0.6 | 0.9 | -0.03 | 0.7 | 0.6 | 0.00 | 0.6 | 0.6 | 0.00 |
| Chronic obstructive lung disease | 2.2 | 2.6 | -0.03 | 2.2 | 2.5 | -0.03 | 2.4 | 2.7 | -0.02 |
| Dementia | 0.2 | 0.2 | 0.00 | 0.1 | 0.2 | -0.01 | 0.1 | 0.2 | -0.01 |
| Gastroesophageal reflux disease | 17.2 | 13.6 | 0.10 | 14.5 | 14.9 | -0.01 | 15.6 | 15.7 | 0.00 |
| Hyperlipidemia | 67.6 | 74.3 | -0.15 | 73.0 | 72.1 | 0.02 | 71.0 | 71.6 | -0.01 |
| Hypertensive disorder | 67.2 | 70.9 | -0.08 | 70.1 | 69.6 | 0.01 | 69.1 | 69.7 | -0.01 |
| Obesity | 48.9 | 30.2 | 0.38 | 36.4 | 37.9 | -0.03 | 40.0 | 41.2 | -0.02 |
| Renal impairment | 3.8 | 4.3 | -0.02 | 4.4 | 4.3 | 0.01 | 4.4 | 4.5 | 0.00 |
| Urinary tract infectious disease | 6.6 | 4.5 | 0.09 | 4.9 | 5.0 | -0.01 | 5.5 | 5.2 | 0.01 |
| Acute urinary tract infection | 0.3 | 0.1 | 0.05 | 0.2 | 0.1 | 0.03 | 0.3 | 0.2 | 0.02 |
| Closed fracture of lower limb | 1.1 | 0.9 | 0.02 | 1.0 | 1.0 | 0.00 | 1.1 | 1.2 | -0.01 |
| **Medical history: Cardiovascular disease** | | | | | | | | | |
| Atrial fibrillation | 1.9 | 2.5 | -0.04 | 2.3 | 2.2 | 0.01 | 2.3 | 2.2 | 0.01 |
| Cerebrovascular disease | 1.8 | 2.5 | -0.04 | 2.0 | 2.0 | 0.00 | 2.0 | 2.2 | -0.01 |
| Coronary arteriosclerosis | 5.2 | 10.7 | -0.20 | 7.1 | 7.2 | 0.00 | 7.7 | 8.2 | -0.02 |
| Heart disease | 15.2 | 20.4 | -0.14 | 17.7 | 18.3 | -0.02 | 17.7 | 18.3 | -0.02 |
| Heart failure | 1.8 | 3.4 | -0.10 | 2.3 | 2.5 | -0.01 | 2.7 | 3.0 | -0.02 |
| Ischemic heart disease | 2.9 | 5.7 | -0.14 | 3.8 | 3.7 | 0.01 | 4.1 | 4.5 | -0.02 |
| Peripheral vascular disease | 4.0 | 4.5 | -0.02 | 4.5 | 4.3 | 0.01 | 4.6 | 4.3 | 0.01 |
| Pulmonary embolism | 0.7 | 0.4 | 0.04 | 0.6 | 0.4 | 0.03 | 0.6 | 0.4 | 0.04 |
| Venous thrombosis | 1.0 | 0.8 | 0.02 | 0.9 | 0.9 | 0.01 | 1.0 | 1.0 | 0.00 |
| Medical history: Neoplasms | 5.5 | 5.6 | 0.00 | 5.5 | 5.7 | -0.01 | 5.5 | 5.7 | -0.01 |
| **Medication use** | | | | | | | | | |
| Agents acting on the renin-angiotensin system | 58.8 | 66.9 | -0.17 | 64.7 | 64.3 | 0.01 | 62.5 | 63.6 | -0.02 |
| Antibacterials for systemic use | 57.7 | 52.3 | 0.11 | 51.9 | 53.0 | -0.02 | 53.9 | 54.4 | -0.01 |
| Antithrombotic agents | 9.2 | 12.6 | -0.11 | 10.0 | 10.3 | -0.01 | 10.9 | 11.3 | -0.01 |
| Beta blocking agents | 23.2 | 26.9 | -0.08 | 24.0 | 24.3 | -0.01 | 24.8 | 25.7 | -0.02 |
| Calcium channel blockers | 20.4 | 21.8 | -0.04 | 22.0 | 21.8 | 0.01 | 21.2 | 21.5 | -0.01 |
| Diuretics | 39.5 | 35.8 | 0.08 | 36.4 | 37.2 | -0.01 | 37.2 | 38.3 | -0.02 |
| Immunosuppressants | 3.9 | 2.8 | 0.06 | 3.2 | 3.3 | 0.00 | 3.5 | 3.7 | -0.01 |
| Lipid modifying agents | 60.9 | 72.5 | -0.25 | 70.6 | 69.7 | 0.02 | 67.3 | 68.0 | -0.01 |

##### 5.5.2 OptumDOD

**Supplementary Table 63:** Baseline patient characteristics for semaglutide (T) and empagliflozin (C) new-users in the OptumDOD data source. We report proportion of initiators satisfying selected base-line characteristics and the standardized difference of population proportions (StdDiff) before and after propensity score adjustment. Less extreme StdDiffs through matching and stratification suggest improved balance between patient cohorts through adjustment. The ”Medication use” entries refer to second-level ATC medication classes.

| | Before adjustment | | | After matching | | | After stratification | | |
|---|---|---|---|---|---|---|---|---|---|
| Characteristic | T | C | StdDiff | T | C | StdDiff | T | C | StdDiff |
| Age: mean | 58.3 | 62.3 | -0.33 | 60.2 | 60.5 | -0.03 | 60.5 | 61.0 | -0.05 |
| Age: std | 12.1 | 11.8 | | 11.9 | 11.7 | | 11.9 | 12.1 | |
| Characteristic (in proportions) | T (%) | C (%) | StdDiff | T (%) | C (%) | StdDiff | T (%) | C (%) | StdDiff |
| Gender: female | 60.3 | 41.2 | 0.38 | 50.9 | 52.1 | -0.02 | 47.8 | 49.2 | -0.03 |
| **Medical history: General** | | | | | | | | | |
| Chronic liver disease | 1.1 | 1.3 | -0.01 | 1.1 | 1.0 | 0.00 | 1.2 | 1.1 | 0.01 |
| Chronic obstructive lung disease | 8.1 | 9.1 | -0.04 | 8.2 | 8.3 | 0.00 | 8.9 | 8.9 | 0.00 |
| Dementia | 0.7 | 1.3 | -0.05 | 0.8 | 1.0 | -0.02 | 1.0 | 1.1 | -0.02 |
| Gastroesophageal reflux disease | 23.4 | 20.3 | 0.07 | 21.0 | 21.5 | -0.01 | 21.6 | 22.0 | -0.01 |
| Hyperlipidemia | 77.5 | 81.8 | -0.11 | 80.0 | 79.4 | 0.02 | 80.6 | 80.2 | 0.01 |
| Hypertensive disorder | 76.3 | 79.8 | -0.08 | 78.7 | 77.7 | 0.02 | 78.9 | 78.4 | 0.01 |
| Obesity | 53.6 | 32.5 | 0.43 | 41.9 | 44.0 | -0.04 | 41.4 | 42.1 | -0.02 |
| Renal impairment | 11.0 | 14.2 | -0.10 | 12.5 | 12.3 | 0.01 | 14.5 | 13.4 | 0.03 |
| Urinary tract infectious disease | 8.8 | 6.9 | 0.07 | 7.7 | 7.6 | 0.01 | 7.9 | 7.8 | 0.00 |
| Acute urinary tract infection | 0.3 | 0.2 | 0.02 | 0.2 | 0.3 | -0.01 | 0.3 | 0.2 | 0.01 |
| Closed fracture of hip | 0.1 | 0.2 | -0.02 | 0.1 | 0.2 | -0.01 | 0.1 | 0.2 | -0.01 |
| Closed fracture of lower limb | 1.5 | 1.3 | 0.02 | 1.2 | 1.3 | -0.01 | 1.2 | 1.4 | -0.02 |
| **Medical history: Cardiovascular disease** | | | | | | | | | |
| Atrial fibrillation | 4.9 | 8.5 | -0.14 | 5.7 | 5.7 | 0.00 | 7.2 | 7.4 | -0.01 |
| Cerebrovascular disease | 4.5 | 6.6 | -0.09 | 5.2 | 4.8 | 0.02 | 6.3 | 5.7 | 0.02 |
| Coronary arteriosclerosis | 12.2 | 21.8 | -0.25 | 14.7 | 14.6 | 0.00 | 19.2 | 18.3 | 0.03 |
| Heart disease | 27.1 | 36.7 | -0.20 | 29.6 | 29.4 | 0.00 | 34.6 | 33.4 | 0.03 |
| Heart failure | 5.9 | 11.2 | -0.18 | 6.9 | 7.1 | -0.01 | 10.3 | 9.6 | 0.02 |
| Ischemic heart disease | 6.3 | 12.0 | -0.19 | 7.4 | 7.6 | -0.01 | 10.7 | 9.9 | 0.03 |
| Peripheral vascular disease | 10.2 | 13.4 | -0.10 | 11.4 | 11.4 | 0.00 | 13.1 | 12.5 | 0.02 |
| Pulmonary embolism | 1.1 | 1.0 | 0.02 | 1.0 | 0.9 | 0.01 | 1.1 | 1.0 | 0.02 |
| Venous thrombosis | 1.2 | 1.6 | -0.04 | 1.2 | 1.4 | -0.02 | 1.3 | 1.6 | -0.02 |
| Medical history: Neoplasms | 8.6 | 10.4 | -0.06 | 9.1 | 9.2 | 0.00 | 9.1 | 9.2 | 0.00 |
| **Medication use** | | | | | | | | | |
| Agents acting on the renin-angiotensin system | 65.2 | 71.6 | -0.14 | 69.9 | 68.9 | 0.02 | 70.0 | 69.5 | 0.01 |
| Antibacterials for systemic use | 58.8 | 53.1 | 0.12 | 54.5 | 55.1 | -0.01 | 54.9 | 55.4 | -0.01 |
| Antithrombotic agents | 14.2 | 21.1 | -0.18 | 15.8 | 15.7 | 0.00 | 19.2 | 18.8 | 0.01 |
| Beta blocking agents | 31.2 | 39.0 | -0.16 | 33.3 | 33.3 | 0.00 | 36.8 | 36.3 | 0.01 |
| Calcium channel blockers | 25.4 | 28.4 | -0.07 | 27.4 | 27.5 | 0.00 | 27.6 | 27.8 | 0.00 |
| Diuretics | 45.0 | 42.6 | 0.05 | 43.9 | 43.8 | 0.00 | 44.5 | 44.3 | 0.00 |
| Lipid modifying agents | 71.1 | 80.3 | -0.22 | 77.8 | 76.7 | 0.03 | 77.7 | 77.3 | 0.01 |

##### 5.5.3 MDCR

**Supplementary Table 64:** Baseline patient characteristics for semaglutide (T) and empagliflozin (C) new-users in the MDCR data source. We report proportion of initiators satisfying selected base-line characteristics and the standardized difference of population proportions (StdDiff) before and after propensity score adjustment. Less extreme StdDiffs through matching and stratification suggest improved balance between patient cohorts through adjustment. The "Medication use" entries refer to second-level ATC medication classes.

| | Before adjustment | | | After matching | | | After stratification | | |
|---|---|---|---|---|---|---|---|---|---|
| Characteristic | T | C | StdDiff | T | C | StdDiff | T | C | StdDiff |
| Age: mean | 71.2 | 72.9 | -0.31 | 71.6 | 71.9 | -0.06 | 72.5 | 72.7 | -0.02 |
| Age: std | 5.0 | 5.1 | | 5.6 | 5.3 | | 5.6 | 5.6 | |
| Characteristic (in proportions) | T (%) | C (%) | StdDiff | T (%) | C (%) | StdDiff | T (%) | C (%) | StdDiff |
| Gender: female | 60.0 | 40.9 | 0.38 | 53.3 | 57.4 | -0.08 | 44.4 | 47.5 | -0.06 |
| **Medical history: General** | | | | | | | | | |
| Chronic liver disease | 1.6 | 1.3 | 0.02 | 1.2 | 1.5 | -0.02 | 1.2 | 1.5 | -0.02 |
| Chronic obstructive lung disease | 9.1 | 11.9 | -0.09 | 9.2 | 10.3 | -0.04 | 10.0 | 11.3 | -0.04 |
| Dementia | 1.0 | 1.6 | -0.05 | 1.2 | 1.7 | -0.04 | 2.0 | 1.7 | 0.02 |
| Gastroesophageal reflux disease | 21.7 | 19.4 | 0.06 | 19.4 | 21.8 | -0.06 | 17.7 | 20.5 | -0.07 |
| Hyperlipidemia | 83.1 | 84.5 | -0.04 | 83.1 | 82.7 | 0.01 | 85.5 | 84.2 | 0.04 |
| Hypertensive disorder | 85.2 | 86.4 | -0.04 | 85.2 | 85.9 | -0.02 | 87.5 | 86.6 | 0.03 |
| Obesity | 48.6 | 29.3 | 0.41 | 39.9 | 44.3 | -0.09 | 34.7 | 36.8 | -0.04 |
| Renal impairment | 16.6 | 20.8 | -0.11 | 18.8 | 17.1 | 0.04 | 20.6 | 20.3 | 0.01 |
| Urinary tract infectious disease | 9.0 | 7.4 | 0.06 | 8.8 | 8.4 | 0.01 | 10.6 | 7.9 | 0.10 |
| Acute urinary tract infection | <0.4 | 0.2 | 0.02 | <0.5 | 0.2 | 0.02 | 0.4 | 0.2 | 0.04 |
| Closed fracture of hip | <0.4 | 0.4 | -0.06 | <0.3 | 0.4 | -0.07 | <0.3 | 0.4 | -0.07 |
| Closed fracture of lower limb | 1.7 | 1.6 | 0.01 | 1.5 | 1.6 | 0.00 | 1.4 | 1.6 | -0.01 |
| **Medical history: Cardiovascular disease** | | | | | | | | | |
| Atrial fibrillation | 9.9 | 16.8 | -0.19 | 11.2 | 10.7 | 0.02 | 13.7 | 15.4 | -0.05 |
| Cerebrovascular disease | 8.5 | 10.3 | -0.06 | 9.4 | 8.9 | 0.02 | 13.1 | 9.7 | 0.11 |
| Coronary arteriosclerosis | 20.6 | 35.3 | -0.32 | 23.6 | 21.0 | 0.06 | 33.9 | 30.2 | 0.08 |
| Heart disease | 39.5 | 55.3 | -0.32 | 42.6 | 40.4 | 0.04 | 52.5 | 50.5 | 0.04 |
| Heart failure | 7.6 | 17.7 | -0.28 | 9.1 | 8.6 | 0.02 | 13.6 | 15.6 | -0.06 |
| Ischemic heart disease | 8.4 | 17.0 | -0.24 | 10.3 | 9.3 | 0.03 | 14.8 | 14.7 | 0.00 |
| Peripheral vascular disease | 14.3 | 16.9 | -0.07 | 14.8 | 13.3 | 0.05 | 17.2 | 16.1 | 0.03 |
| Pulmonary embolism | 0.7 | 1.1 | -0.04 | 0.8 | 0.8 | 0.00 | 0.5 | 1.1 | -0.06 |
| Venous thrombosis | 1.2 | 2.2 | -0.07 | 1.1 | 2.0 | -0.07 | 0.8 | 2.1 | -0.09 |
| Medical history: Neoplasms | 16.6 | 18.0 | -0.04 | 17.1 | 16.5 | 0.02 | 17.1 | 17.6 | -0.01 |
| **Medication use** | | | | | | | | | |
| Agents acting on the renin-angiotensin system | 74.4 | 77.8 | -0.08 | 74.4 | 76.6 | -0.05 | 77.9 | 77.3 | 0.01 |
| Antibacterials for systemic use | 59.5 | 57.4 | 0.04 | 57.4 | 60.1 | -0.05 | 53.7 | 58.1 | -0.09 |
| Antithrombotic agents | 20.2 | 31.8 | -0.26 | 23.0 | 21.3 | 0.04 | 23.0 | 21.3 | 0.04 |
| Beta blocking agents | 44.3 | 54.0 | -0.19 | 45.1 | 44.6 | 0.01 | 53.8 | 50.9 | 0.06 |
| Calcium channel blockers | 34.1 | 36.0 | -0.04 | 34.9 | 36.1 | -0.02 | 33.8 | 36.1 | -0.05 |
| Diuretics | 52.2 | 51.8 | 0.01 | 50.3 | 51.8 | -0.03 | 49.4 | 52.7 | -0.06 |
| Immunosuppressants | 5.3 | 3.9 | 0.07 | 4.9 | 4.6 | 0.02 | 3.4 | 4.0 | -0.03 |
| Lipid modifying agents | 84.8 | 86.5 | -0.05 | 85.1 | 86.2 | -0.03 | 85.1 | 86.6 | -0.04 |

##### 5.5.4 Open Claims

**Supplementary Table 65:** Baseline patient characteristics for semaglutide (T) and empagliflozin (C) new-users in the Open Claims data source. We report proportion of initiators satisfying selected base-line characteristics and the standardized difference of population proportions (StdDiff) before and after propensity score adjustment. Less extreme StdDiffs through matching and stratification suggest improved balance between patient cohorts through adjustment. The "Medication use" entries refer to second-level ATC medication classes.

| | Before adjustment | | | After matching | | | After stratification | | |
|---|---|---|---|---|---|---|---|---|---|
| Characteristic | T | C | StdDiff | T | C | StdDiff | T | C | StdDiff |
| Age: mean | 55.3 | 59.4 | -0.33 | 57.0 | 57.1 | 0.00 | 57.9 | 58.1 | -0.01 |
| Age: std | 12.1 | 11.8 | | 11.9 | 11.8 | | 11.9 | 12.2 | |
| Characteristic (in proportions) | T (%) | C (%) | StdDiff | T (%) | C (%) | StdDiff | T (%) | C (%) | StdDiff |
| Gender: female | 61.3 | 43.4 | 0.36 | 53.7 | 53.8 | 0.00 | 50.1 | 50.0 | 0.00 |
| **Medical history: General** | | | | | | | | | |
| Chronic liver disease | 0.5 | 0.7 | -0.03 | 0.5 | 0.6 | 0.00 | 0.6 | 0.6 | 0.00 |
| Chronic obstructive lung disease | 3.7 | 4.7 | -0.05 | 3.9 | 4.1 | -0.01 | 4.4 | 4.6 | -0.01 |
| Dementia | 0.3 | 0.4 | -0.03 | 0.3 | 0.3 | -0.01 | 0.4 | 0.4 | 0.00 |
| Gastroesophageal reflux disease | 9.8 | 8.4 | 0.05 | 8.7 | 8.8 | 0.00 | 9.0 | 9.1 | 0.00 |
| Hyperlipidemia | 39.2 | 43.7 | -0.09 | 39.7 | 40.0 | 0.00 | 41.7 | 41.9 | -0.01 |
| Hypertensive disorder | 46.0 | 48.6 | -0.05 | 45.6 | 45.8 | 0.00 | 47.7 | 47.8 | 0.00 |
| Obesity | 21.2 | 12.3 | 0.24 | 15.6 | 15.9 | -0.01 | 16.1 | 15.8 | 0.01 |
| Renal impairment | 4.3 | 5.4 | -0.05 | 4.6 | 4.7 | -0.01 | 5.4 | 5.3 | 0.00 |
| Urinary tract infectious disease | 4.5 | 3.5 | 0.05 | 3.8 | 3.8 | 0.00 | 4.0 | 3.9 | 0.00 |
| Acute urinary tract infection | 0.1 | 0.1 | 0.01 | 0.1 | 0.1 | 0.00 | 0.1 | 0.1 | 0.00 |
| Closed fracture of hip | 0.1 | 0.1 | -0.01 | 0.1 | 0.1 | -0.01 | 0.1 | 0.1 | 0.00 |
| Closed fracture of lower limb | 0.8 | 0.7 | 0.01 | 0.7 | 0.8 | -0.01 | 0.8 | 0.8 | 0.00 |
| **Medical history: Cardiovascular disease** | | | | | | | | | |
| Atrial fibrillation | 2.5 | 4.3 | -0.09 | 2.9 | 2.9 | 0.00 | 3.8 | 3.8 | 0.00 |
| Cerebrovascular disease | 1.7 | 2.8 | -0.07 | 1.9 | 1.9 | 0.00 | 2.3 | 2.4 | -0.01 |
| Coronary arteriosclerosis | 5.2 | 10.8 | -0.20 | 6.2 | 6.3 | 0.00 | 8.6 | 8.9 | -0.01 |
| Heart disease | 13.1 | 20.3 | -0.19 | 14.2 | 14.4 | 0.00 | 17.7 | 18.0 | -0.01 |
| Heart failure | 2.5 | 5.2 | -0.14 | 2.8 | 3.0 | -0.01 | 4.2 | 4.5 | -0.01 |
| Ischemic heart disease | 2.5 | 5.3 | -0.14 | 2.9 | 3.0 | 0.00 | 4.2 | 4.4 | -0.01 |
| Peripheral vascular disease | 3.6 | 4.9 | -0.06 | 3.9 | 4.0 | 0.00 | 4.5 | 4.6 | 0.00 |
| Pulmonary embolism | 0.5 | 0.5 | 0.01 | 0.5 | 0.5 | 0.00 | 0.6 | 0.5 | 0.00 |
| Venous thrombosis | 0.6 | 0.7 | -0.02 | 0.6 | 0.6 | -0.01 | 0.7 | 0.7 | 0.00 |
| Medical history: Neoplasms | 4.4 | 5.2 | -0.04 | 4.4 | 4.6 | -0.01 | 4.9 | 5.0 | 0.00 |
| **Medication use** | | | | | | | | | |
| Agents acting on the renin-angiotensin system | 60.7 | 68.4 | -0.16 | 64.4 | 64.2 | 0.00 | 65.6 | 65.8 | 0.00 |
| Antibacterials for systemic use | 57.5 | 52.5 | 0.10 | 53.9 | 54.3 | -0.01 | 53.9 | 54.1 | 0.00 |
| Antithrombotic agents | 13.5 | 21.1 | -0.20 | 15.2 | 15.3 | 0.00 | 18.5 | 18.8 | -0.01 |
| Beta blocking agents | 28.6 | 35.5 | -0.15 | 30.1 | 30.2 | 0.00 | 33.2 | 33.5 | -0.01 |
| Calcium channel blockers | 23.1 | 25.8 | -0.06 | 24.6 | 24.7 | 0.00 | 25.3 | 25.4 | 0.00 |
| Diuretics | 41.3 | 39.4 | 0.04 | 39.8 | 40.2 | -0.01 | 40.3 | 40.5 | 0.00 |
| Immunosuppressants | 3.6 | 2.8 | 0.05 | 3.1 | 3.2 | -0.01 | 3.1 | 3.2 | 0.00 |
| Lipid modifying agents | 64.5 | 74.9 | -0.23 | 70.1 | 69.8 | 0.01 | 71.7 | 71.7 | 0.00 |

##### 5.5.5 CUIMC

**Supplementary Table 66:** Baseline patient characteristics for semaglutide (T) and empagliflozin (C) new-users in the CUIMC data source. We report proportion of initiators satisfying selected base-line characteristics and the standardized difference of population proportions (StdDiff) before and after propensity score adjustment. Less extreme StdDiffs through matching and stratification suggest improved balance between patient cohorts through adjustment. The "Medication use" entries refer to second-level ATC medication classes.

| | Before adjustment | | | After matching | | | After stratification | | |
|---|---|---|---|---|---|---|---|---|---|
| Characteristic | T | C | StdDiff | T | C | StdDiff | T | C | StdDiff |
| Age: mean | 57.2 | 64.7 | -0.60 | 61.0 | 62.4 | -0.12 | 61.1 | 61.7 | -0.05 |
| Age: std | | | | | | | | | |
| Characteristic (in proportions) | T (%) | C (%) | StdDiff | T (%) | C (%) | StdDiff | T (%) | C (%) | StdDiff |
| Gender: female | 65.4 | 44.8 | 0.42 | 52.3 | 54.0 | -0.03 | 51.7 | 58.5 | -0.14 |
| **Race** | | | | | | | | | |
| Asian | 3.0 | 3.6 | -0.03 | 2.2 | 3.8 | -0.09 | 3.9 | 2.8 | 0.06 |
| Black or African American | 16.8 | 16.6 | 0.00 | 13.7 | 16.2 | -0.07 | 14.0 | 15.6 | -0.05 |
| White | 42.9 | 41.1 | 0.04 | 45.5 | 41.9 | 0.07 | 42.7 | 46.2 | -0.07 |
| Native Hawaiian or Other Pacific Islander | <0.4 | <0.4 | -0.02 | <1.0 | 0.2 | 0.00 | 0.4 | <0.3 | 0.05 |
| American Indian or Alaska Native | <0.4 | <0.4 | 0.02 | <0.3 | <0.3 | 0.01 | <0.3 | <0.3 | 0.01 |
| **Ethnicity** | | | | | | | | | |
| Hispanic or Latino | 22.0 | 24.8 | -0.07 | 20.9 | 22.6 | -0.04 | 24.7 | 23.9 | 0.02 |
| Not Hispanic or Latino | 58.4 | 56.0 | 0.05 | 59.2 | 58.2 | 0.02 | 58.1 | 57.0 | 0.02 |
| **Medical history: General** | | | | | | | | | |
| Chronic liver disease | 1.0 | 1.0 | 0.00 | 1.4 | <0.2 | 0.15 | 0.7 | 0.4 | 0.04 |
| Chronic obstructive lung disease | 3.4 | 4.4 | -0.06 | 5.0 | 2.8 | 0.11 | 3.0 | 4.2 | -0.06 |
| Dementia | <0.4 | 1.3 | -0.12 | <1.0 | 0.8 | -0.05 | 0.5 | 0.9 | -0.05 |
| Gastroesophageal reflux disease | 17.8 | 13.9 | 0.11 | 13.1 | 12.5 | 0.02 | 13.4 | 11.2 | 0.07 |
| Hyperlipidemia | 59.4 | 61.6 | -0.04 | 60.6 | 57.1 | 0.07 | 62.8 | 61.8 | 0.02 |
| Hypertensive disorder | 58.1 | 66.5 | -0.17 | 58.3 | 59.4 | -0.02 | 62.5 | 59.1 | 0.07 |
| Obesity | 58.3 | 26.8 | 0.67 | 28.6 | 37.2 | -0.18 | 37.5 | 46.0 | -0.17 |
| Renal impairment | 4.5 | 10.0 | -0.21 | 6.0 | 5.8 | 0.01 | 8.4 | 6.7 | 0.07 |
| Urinary tract infectious disease | 3.3 | 1.9 | 0.09 | 1.6 | 1.6 | 0.00 | 2.1 | 1.4 | 0.06 |
| Closed fracture of lower limb | 1.0 | 0.5 | 0.06 | 1.2 | 0.6 | 0.06 | 1.0 | 0.6 | 0.05 |
| **Medical history: Cardiovascular disease** | | | | | | | | | |
| Atrial fibrillation | 4.6 | 9.0 | -0.18 | 7.0 | 4.9 | 0.09 | 8.4 | 5.9 | 0.10 |
| Cerebrovascular disease | 4.1 | 7.9 | -0.16 | 5.8 | 5.2 | 0.03 | 5.8 | 6.6 | -0.03 |
| Coronary arteriosclerosis | 10.5 | 27.1 | -0.44 | 15.7 | 12.3 | 0.10 | 18.4 | 16.6 | 0.05 |
| Heart disease | 23.4 | 44.2 | -0.45 | 30.0 | 25.8 | 0.09 | 35.6 | 30.6 | 0.11 |
| Heart failure | 4.1 | 16.1 | -0.41 | 6.8 | 6.4 | 0.02 | 10.0 | 9.7 | 0.01 |
| Ischemic heart disease | 3.1 | 12.4 | -0.35 | 5.2 | 4.8 | 0.02 | 7.0 | 7.4 | -0.01 |
| Peripheral vascular disease | 3.2 | 7.5 | -0.19 | 4.4 | 3.7 | 0.03 | 4.7 | 6.1 | -0.06 |
| Pulmonary embolism | 0.6 | 0.6 | 0.01 | <1.0 | 0.5 | -0.01 | 0.7 | 0.4 | 0.04 |
| Venous thrombosis | 1.8 | 1.4 | 0.03 | 2.0 | 1.6 | 0.03 | 3.0 | 1.2 | 0.13 |
| Medical history: Neoplasms | 9.6 | 9.3 | 0.01 | 9.1 | 8.1 | 0.04 | 7.8 | 8.6 | -0.03 |
| **Medication use** | | | | | | | | | |
| Agents acting on the renin-angiotensin system | 43.8 | 61.9 | -0.37 | 51.7 | 50.2 | 0.03 | 52.8 | 46.9 | 0.12 |
| Antibacterials for systemic use | 34.2 | 30.3 | 0.08 | 27.0 | 28.3 | -0.03 | 31.3 | 32.6 | -0.03 |
| Antithrombotic agents | 21.6 | 39.9 | -0.40 | 26.6 | 22.3 | 0.10 | 30.0 | 30.3 | -0.01 |
| Beta blocking agents | 24.1 | 40.8 | -0.36 | 31.2 | 28.1 | 0.07 | 34.9 | 32.6 | 0.05 |
| Calcium channel blockers | 25.6 | 29.8 | -0.09 | 25.8 | 25.2 | 0.01 | 29.1 | 24.8 | 0.10 |
| Diuretics | 32.2 | 39.1 | -0.14 | 32.4 | 32.2 | 0.00 | 38.0 | 37.1 | 0.02 |
| Immunosuppressants | 5.4 | 4.7 | 0.04 | 4.2 | 3.9 | 0.01 | 4.1 | 3.6 | 0.03 |
| Lipid modifying agents | 57.3 | 72.3 | -0.32 | 66.0 | 63.9 | 0.04 | 65.2 | 61.1 | 0.09 |

##### 5.5.6 OptumEHR

**Supplementary Table 67:** Baseline patient characteristics for semaglutide (T) and empagliflozin (C) new-users in the OptumEHR data source. We report proportion of initiators satisfying selected base-line characteristics and the standardized difference of population proportions (StdDiff) before and after propensity score adjustment. Less extreme StdDiffs through matching and stratification suggest improved balance between patient cohorts through adjustment. The "Medication use" entries refer to second-level ATC medication classes.

| | Before adjustment | | | After matching | | | After stratification | | |
|---|---|---|---|---|---|---|---|---|---|
| Characteristic | T | C | StdDiff | T | C | StdDiff | T | C | StdDiff |
| Age: mean | 53.4 | 57.8 | -0.37 | 55.3 | 55.5 | -0.02 | 56.2 | 56.2 | 0.00 |
| Age: std | 11.9 | 11.6 | | 11.6 | 11.5 | | 11.6 | 11.9 | |
| Characteristic (in proportions) | T (%) | C (%) | StdDiff | T (%) | C (%) | StdDiff | T (%) | C (%) | StdDiff |
| Gender: female | 62.6 | 42.4 | 0.40 | 52.7 | 54.5 | -0.04 | 46.7 | 50.2 | -0.07 |
| **Race** | | | | | | | | | |
| Asian | 1.9 | 3.0 | -0.07 | 2.0 | 2.3 | -0.02 | 3.1 | 2.6 | 0.03 |
| Black or African American | 13.9 | 10.8 | 0.10 | 12.2 | 12.2 | 0.00 | 10.9 | 11.9 | -0.03 |
| White | 77.1 | 78.3 | -0.03 | 78.3 | 77.9 | 0.01 | 79.0 | 78.0 | 0.03 |
| **Ethnicity** | | | | | | | | | |
| Hispanic or Latino | 5.6 | 6.0 | -0.02 | 5.8 | 6.1 | -0.01 | 5.7 | 6.1 | -0.02 |
| Not Hispanic or Latino | 84.6 | 83.5 | 0.03 | 83.8 | 83.6 | 0.00 | 83.7 | 83.9 | 0.00 |
| **Medical history: General** | | | | | | | | | |
| Chronic liver disease | 1.1 | 1.3 | -0.02 | 1.2 | 1.2 | 0.01 | 1.1 | 1.2 | 0.00 |
| Chronic obstructive lung disease | 4.0 | 5.1 | -0.05 | 4.2 | 4.4 | 0.00 | 4.9 | 4.8 | 0.00 |
| Dementia | 0.3 | 0.5 | -0.02 | 0.3 | 0.4 | 0.00 | 0.8 | 0.4 | 0.06 |
| Gastroesophageal reflux disease | 20.8 | 17.0 | 0.10 | 18.1 | 18.5 | -0.01 | 19.0 | 18.6 | 0.01 |
| Hyperlipidemia | 62.3 | 67.8 | -0.12 | 64.3 | 63.5 | 0.02 | 65.6 | 65.3 | 0.01 |
| Hypertensive disorder | 63.6 | 67.2 | -0.08 | 64.4 | 64.1 | 0.01 | 66.0 | 66.0 | 0.00 |
| Obesity | 51.6 | 33.0 | 0.38 | 40.5 | 42.8 | -0.05 | 39.4 | 40.8 | -0.03 |
| Renal impairment | 5.4 | 6.0 | -0.03 | 5.7 | 5.6 | 0.00 | 6.8 | 6.0 | 0.03 |
| Urinary tract infectious disease | 4.3 | 3.0 | 0.07 | 3.5 | 3.5 | 0.00 | 3.5 | 3.2 | 0.02 |
| Primary osteoporosis | 0.1 | 0.1 | 0.01 | 0.2 | 0.1 | 0.03 | 0.2 | 0.1 | 0.03 |
| Acute urinary tract infection | 1.1 | 0.4 | 0.09 | 0.8 | 0.7 | 0.01 | 0.7 | 0.6 | 0.02 |
| Closed fracture of hip | 0.0 | 0.1 | -0.01 | <0.1 | 0.0 | 0.00 | 0.1 | 0.1 | 0.00 |
| Closed fracture of lower limb | 0.8 | 0.6 | 0.02 | 0.7 | 0.7 | 0.00 | 0.8 | 0.6 | 0.02 |
| **Medical history: Cardiovascular disease** | | | | | | | | | |
| Atrial fibrillation | 3.0 | 4.9 | -0.09 | 3.4 | 3.5 | 0.00 | 4.7 | 4.3 | 0.02 |
| Cerebrovascular disease | 2.1 | 3.0 | -0.06 | 2.5 | 2.3 | 0.01 | 2.9 | 2.6 | 0.02 |
| Coronary arteriosclerosis | 7.1 | 15.2 | -0.25 | 9.2 | 8.7 | 0.02 | 14.2 | 12.2 | 0.06 |
| Heart disease | 17.2 | 25.6 | -0.20 | 19.2 | 18.4 | 0.02 | 24.7 | 22.3 | 0.06 |
| Heart failure | 2.5 | 4.9 | -0.12 | 3.0 | 3.1 | -0.01 | 5.0 | 4.2 | 0.04 |
| Ischemic heart disease | 3.8 | 6.8 | -0.13 | 4.4 | 4.1 | 0.01 | 6.7 | 5.6 | 0.05 |
| Peripheral vascular disease | 5.6 | 6.8 | -0.05 | 5.7 | 5.7 | 0.00 | 7.1 | 6.7 | 0.02 |
| Pulmonary embolism | 0.7 | 0.5 | 0.03 | 0.6 | 0.6 | 0.00 | 0.6 | 0.6 | -0.01 |
| Venous thrombosis | 0.9 | 0.9 | 0.00 | 0.9 | 0.8 | 0.00 | 0.9 | 0.8 | 0.00 |
| Chronic urinary tract infection | 0.1 | 0.1 | 0.03 | 0.1 | 0.1 | 0.00 | 0.1 | 0.1 | 0.00 |
| Medical history: Neoplasms | 5.1 | 5.9 | -0.03 | 5.3 | 5.4 | 0.00 | 5.3 | 5.7 | -0.02 |
| **Medication use** | | | | | | | | | |
| Agents acting on the renin-angiotensin system | 58.4 | 66.5 | -0.17 | 62.6 | 61.8 | 0.02 | 63.5 | 63.6 | 0.00 |
| Antibacterials for systemic use | 46.3 | 40.5 | 0.12 | 42.0 | 42.2 | 0.00 | 41.1 | 42.5 | -0.03 |
| Antithrombotic agents | 27.7 | 39.4 | -0.25 | 31.1 | 30.3 | 0.02 | 36.6 | 34.7 | 0.04 |
| Beta blocking agents | 28.5 | 34.7 | -0.13 | 29.7 | 29.6 | 0.00 | 34.9 | 32.5 | 0.05 |
| Calcium channel blockers | 22.3 | 24.3 | -0.05 | 22.8 | 22.9 | 0.00 | 24.0 | 23.7 | 0.01 |
| Diuretics | 40.5 | 38.0 | 0.05 | 38.6 | 38.7 | 0.00 | 37.4 | 39.0 | -0.03 |
| Immunosuppressants | 3.7 | 2.9 | 0.04 | 3.3 | 3.2 | 0.00 | 2.9 | 3.2 | -0.02 |
| Lipid modifying agents | 63.0 | 74.5 | -0.25 | 69.8 | 68.6 | 0.03 | 71.3 | 70.3 | 0.02 |

#### 5.5.7 VA

**Supplementary Table 68:** Baseline patient characteristics for semaglutide (T) and empagliflozin (C) new-users in the VA data source. We report proportion of initiators satisfying selected base-line characteristics and the standardized difference of population proportions (StdDiff) before and after propensity score adjustment. Less extreme StdDiffs through matching and stratification suggest improved balance between patient cohorts through adjustment. The "Medication use" entries refer to second-level ATC medication classes.

| | Before adjustment | | | After matching | | | After stratification | | |
|---|---|---|---|---|---|---|---|---|---|
| Characteristic | T | C | StdDiff | T | C | StdDiff | T | C | StdDiff |
| Age: mean | 59.6 | 65.3 | -0.48 | 61.0 | 61.1 | 0.00 | 64.8 | 64.9 | -0.01 |
| Age: std | 11.6 | 11.7 | | 11.6 | 11.9 | | 11.6 | 11.7 | |
| Characteristic (in proportions) | T (%) | C (%) | StdDiff | T (%) | C (%) | StdDiff | T (%) | C (%) | StdDiff |
| Gender: female | 21.6 | 6.6 | 0.56 | 15.2 | 16.9 | -0.05 | 7.9 | 7.5 | 0.01 |
| **Race** | | | | | | | | | |
| Asian | 1.0 | 1.5 | -0.04 | 0.9 | 1.2 | -0.03 | 1.2 | 1.5 | -0.03 |
| Black or African American | 22.5 | 18.7 | 0.10 | 20.3 | 21.0 | -0.02 | 13.5 | 19.0 | -0.14 |
| White | 67.2 | 71.3 | -0.09 | 69.5 | 68.6 | 0.02 | 74.2 | 70.9 | 0.07 |
| Unknown | 7.1 | 6.5 | 0.03 | 7.2 | 6.9 | 0.01 | 9.6 | 6.5 | 0.12 |
| Native Hawaiian or Other Pacific Islander | 1.3 | 1.2 | 0.01 | 1.3 | 1.4 | -0.01 | 1.1 | 1.2 | -0.01 |
| American Indian or Alaska Native | 0.8 | 0.8 | 0.00 | 0.7 | 0.9 | -0.02 | 0.5 | 0.8 | -0.03 |
| **Ethnicity** | | | | | | | | | |
| Hispanic or Latino | 7.1 | 7.3 | -0.01 | 6.9 | 6.7 | 0.01 | 8.2 | 7.3 | 0.03 |
| Not Hispanic or Latino | 90.0 | 90.0 | 0.00 | 90.1 | 90.5 | -0.01 | 87.6 | 90.0 | -0.08 |
| **Medical history: General** | | | | | | | | | |
| Chronic liver disease | 2.4 | 2.1 | 0.02 | 2.5 | 2.1 | 0.03 | 1.2 | 2.1 | -0.06 |
| Chronic obstructive lung disease | 9.3 | 13.0 | -0.11 | 9.7 | 10.0 | -0.01 | 10.8 | 12.8 | -0.06 |
| Dementia | 0.8 | 1.3 | -0.04 | 0.8 | 0.9 | -0.01 | 0.7 | 1.2 | -0.05 |
| Gastroesophageal reflux disease | 25.0 | 22.6 | 0.06 | 23.7 | 24.1 | -0.01 | 23.2 | 22.8 | 0.01 |
| Hyperlipidemia | 67.2 | 74.9 | -0.17 | 68.9 | 69.3 | -0.01 | 76.7 | 74.4 | 0.05 |
| Hypertensive disorder | 71.6 | 76.6 | -0.12 | 73.7 | 72.5 | 0.03 | 77.0 | 76.2 | 0.02 |
| Obesity | 70.7 | 31.9 | 0.81 | 58.3 | 62.0 | -0.07 | 31.8 | 34.5 | -0.06 |
| Renal impairment | 7.4 | 12.1 | -0.14 | 8.1 | 8.2 | 0.00 | 14.4 | 11.9 | 0.08 |
| Urinary tract infectious disease | 3.7 | 1.7 | 0.14 | 3.1 | 2.9 | 0.01 | 2.1 | 1.8 | 0.02 |
| Acute urinary tract infection | <0.2 | 0.1 | -0.01 | <0.3 | 0.1 | -0.02 | <0.1 | 0.1 | -0.02 |
| Closed fracture of hip | <0.2 | 0.0 | -0.01 | 0.0 | 0.0 | 0.00 | <0.1 | 0.0 | -0.02 |
| Closed fracture of lower limb | 0.7 | 0.4 | 0.03 | 0.5 | 0.5 | 0.00 | 0.3 | 0.4 | -0.02 |
| **Medical history: Cardiovascular disease** | | | | | | | | | |
| Atrial fibrillation | 7.2 | 13.2 | -0.18 | 8.5 | 8.3 | 0.01 | 17.3 | 12.9 | 0.13 |
| Cerebrovascular disease | 3.1 | 4.2 | -0.05 | 3.2 | 2.9 | 0.02 | 3.7 | 4.1 | -0.02 |
| Coronary arteriosclerosis | 14.2 | 27.0 | -0.29 | 17.2 | 16.4 | 0.02 | 30.2 | 26.0 | 0.10 |
| Heart disease | 28.2 | 44.1 | -0.32 | 31.6 | 30.8 | 0.02 | 46.4 | 42.9 | 0.07 |
| Heart failure | 5.0 | 15.2 | -0.29 | 6.0 | 6.0 | 0.00 | 15.3 | 14.6 | 0.02 |
| Ischemic heart disease | 5.5 | 11.3 | -0.19 | 6.2 | 6.1 | 0.01 | 11.2 | 10.8 | 0.01 |
| Peripheral vascular disease | 4.0 | 6.0 | -0.09 | 4.5 | 4.4 | 0.00 | 6.6 | 5.9 | 0.03 |
| Pulmonary embolism | 0.9 | 1.0 | -0.01 | 0.7 | 0.8 | -0.01 | 2.0 | 1.0 | 0.10 |
| Venous thrombosis | 1.2 | 1.3 | -0.01 | 1.0 | 0.9 | 0.00 | 0.7 | 1.3 | -0.05 |
| Medical history: Neoplasms | 6.7 | 9.6 | -0.10 | 6.9 | 7.0 | 0.00 | 7.9 | 9.4 | -0.05 |
| **Medication use** | | | | | | | | | |
| Agents acting on the renin-angiotensin system | 57.4 | 67.2 | -0.21 | 60.3 | 59.0 | 0.03 | 65.9 | 66.5 | -0.01 |
| Antibacterials for systemic use | 31.2 | 26.1 | 0.12 | 27.3 | 28.2 | -0.02 | 22.7 | 26.3 | -0.08 |
| Antithrombotic agents | 31.5 | 42.7 | -0.23 | 33.2 | 32.8 | 0.01 | 44.3 | 41.9 | 0.05 |
| Beta blocking agents | 32.7 | 46.2 | -0.27 | 35.6 | 34.5 | 0.02 | 49.1 | 45.2 | 0.08 |
| Calcium channel blockers | 26.5 | 30.2 | -0.08 | 27.1 | 26.4 | 0.01 | 28.0 | 29.9 | -0.04 |
| Diuretics | 40.6 | 42.7 | -0.04 | 40.7 | 39.8 | 0.02 | 47.3 | 42.5 | 0.10 |
| Immunosuppressants | 4.6 | 3.3 | 0.07 | 4.2 | 4.0 | 0.01 | 2.9 | 3.3 | -0.03 |
| Lipid modifying agents | 75.1 | 84.0 | -0.24 | 76.9 | 76.4 | 0.01 | 81.8 | 83.4 | -0.04 |

#### 5.6 dapagliflozin vs empagliflozin

##### 5.6.1 CCAE

**Supplementary Table 69:** Baseline patient characteristics for dapagliflozin (T) and empagliflozin (C) new-users in the CCAE data source. We report proportion of initiators satisfying selected base-line characteristics and the standardized difference of population proportions (StdDiff) before and after propensity score adjustment. Less extreme StdDiffs through matching and stratification suggest improved balance between patient cohorts through adjustment. The "Medication use" entries refer to second-level ATC medication classes.

| | Before adjustment | | | After matching | | | After stratification | | |
|---|---|---|---|---|---|---|---|---|---|
| Characteristic | T | C | StdDiff | T | C | StdDiff | T | C | StdDiff |
| Age: mean | 52.5 | 53.2 | -0.08 | 52.6 | 52.8 | -0.02 | 52.8 | 53.0 | -0.02 |
| Age: std | 8.5 | 8.5 | | 8.3 | 8.4 | | 8.3 | 8.3 | |
| Characteristic (in proportions) | T (%) | C (%) | StdDiff | T (%) | C (%) | StdDiff | T (%) | C (%) | StdDiff |
| Gender: female | 43.7 | 40.3 | 0.07 | 41.6 | 41.6 | 0.00 | 41.6 | 41.6 | 0.00 |
| **Medical history: General** | | | | | | | | | |
| Chronic liver disease | 1.4 | 0.9 | 0.05 | 1.0 | 1.1 | -0.01 | 1.1 | 1.1 | 0.00 |
| Chronic obstructive lung disease | 2.4 | 2.6 | -0.02 | 2.4 | 2.5 | 0.00 | 2.5 | 2.6 | -0.01 |
| Dementia | 0.1 | 0.2 | -0.01 | 0.1 | 0.2 | -0.02 | 0.1 | 0.2 | -0.01 |
| Gastroesophageal reflux disease | 13.3 | 13.6 | -0.01 | 13.7 | 13.6 | 0.00 | 13.7 | 13.6 | 0.00 |
| Hyperlipidemia | 73.3 | 74.3 | -0.02 | 73.6 | 73.6 | 0.00 | 74.1 | 73.9 | 0.01 |
| Hypertensive disorder | 71.2 | 70.9 | 0.01 | 71.3 | 70.9 | 0.01 | 71.3 | 71.1 | 0.00 |
| Obesity | 28.3 | 30.2 | -0.04 | 29.6 | 29.7 | 0.00 | 29.6 | 29.7 | 0.00 |
| Renal impairment | 4.7 | 4.3 | 0.02 | 5.0 | 4.8 | 0.01 | 4.6 | 4.5 | 0.01 |
| Urinary tract infectious disease | 5.9 | 4.5 | 0.06 | 5.2 | 5.0 | 0.01 | 5.2 | 5.0 | 0.01 |
| Acute urinary tract infection | 0.2 | 0.1 | 0.02 | 0.2 | 0.1 | 0.01 | 0.2 | 0.1 | 0.02 |
| Closed fracture of lower limb | 1.0 | 0.9 | 0.01 | 1.0 | 1.0 | 0.00 | 1.0 | 0.9 | 0.00 |
| **Medical history: Cardiovascular disease** | | | | | | | | | |
| Atrial fibrillation | 2.3 | 2.5 | -0.01 | 2.5 | 2.2 | 0.02 | 2.6 | 2.4 | 0.01 |
| Cerebrovascular disease | 2.3 | 2.5 | -0.01 | 2.3 | 2.2 | 0.00 | 2.4 | 2.4 | 0.00 |
| Coronary arteriosclerosis | 7.3 | 10.7 | -0.12 | 8.0 | 7.6 | 0.01 | 9.3 | 9.3 | 0.00 |
| Heart disease | 17.3 | 20.4 | -0.08 | 17.9 | 17.7 | 0.00 | 19.1 | 19.1 | 0.00 |
| Heart failure | 3.2 | 3.4 | -0.01 | 3.5 | 3.4 | 0.01 | 3.4 | 3.3 | 0.00 |
| Ischemic heart disease | 4.2 | 5.7 | -0.07 | 4.5 | 4.3 | 0.01 | 5.1 | 5.0 | 0.00 |
| Peripheral vascular disease | 3.8 | 4.5 | -0.03 | 4.1 | 3.9 | 0.01 | 4.3 | 4.2 | 0.01 |
| Pulmonary embolism | 0.4 | 0.4 | 0.00 | 0.4 | 0.4 | 0.00 | 0.4 | 0.4 | 0.00 |
| Venous thrombosis | 0.9 | 0.8 | 0.00 | 0.9 | 0.8 | 0.01 | 0.9 | 0.8 | 0.01 |
| Medical history: Neoplasms | 5.2 | 5.6 | -0.02 | 5.3 | 5.3 | 0.00 | 5.5 | 5.5 | 0.00 |
| **Medication use** | | | | | | | | | |
| Agents acting on the renin-angiotensin system | 67.4 | 66.9 | 0.01 | 67.2 | 67.0 | 0.00 | 67.4 | 67.1 | 0.01 |
| Antibacterials for systemic use | 56.2 | 52.3 | 0.08 | 54.6 | 54.8 | 0.00 | 53.9 | 54.1 | 0.00 |
| Antithrombotic agents | 10.0 | 12.6 | -0.08 | 10.8 | 10.4 | 0.01 | 11.4 | 11.5 | 0.00 |
| Beta blocking agents | 25.5 | 26.9 | -0.03 | 25.8 | 25.3 | 0.01 | 26.4 | 26.3 | 0.00 |
| Calcium channel blockers | 21.6 | 21.8 | -0.01 | 21.7 | 21.9 | 0.00 | 21.9 | 21.8 | 0.00 |
| Diuretics | 37.8 | 35.8 | 0.04 | 37.2 | 37.3 | 0.00 | 36.6 | 36.8 | 0.00 |
| Immunosuppressants | 2.3 | 2.8 | -0.03 | 2.4 | 2.6 | -0.01 | 2.5 | 2.7 | -0.01 |
| Lipid modifying agents | 68.2 | 72.5 | -0.09 | 69.8 | 69.4 | 0.01 | 71.0 | 70.7 | 0.01 |

##### 5.6.2 GermanyDA

**Supplementary Table 70:** Baseline patient characteristics for dapagliflozin (T) and empagliflozin (C) new-users in the GermanyDA data source. We report proportion of initiators satisfying selected base-line characteristics and the standardized difference of population proportions (StdDiff) before and after propensity score adjustment. Less extreme StdDiffs through matching and stratification suggest improved balance between patient cohorts through adjustment. The "Medication use" entries refer to second-level ATC medication classes.

| | Before adjustment | | | After matching | | | After stratification | | |
|---|---|---|---|---|---|---|---|---|---|
| Characteristic | T | C | StdDiff | T | C | StdDiff | T | C | StdDiff |
| Age: mean | 62.4 | 64.7 | -0.20 | 63.5 | 63.8 | -0.02 | 63.4 | 63.7 | -0.03 |
| Age: std | | | | | | | | | |
| Characteristic (in proportions) | T (%) | C (%) | StdDiff | T (%) | C (%) | StdDiff | T (%) | C (%) | StdDiff |
| Gender: female | 37.4 | 34.4 | 0.06 | 36.2 | 36.0 | 0.00 | 36.0 | 35.8 | 0.00 |
| **Medical history: General** | | | | | | | | | |
| Chronic liver disease | 0.5 | 0.4 | 0.03 | 0.7 | 0.3 | 0.05 | 0.6 | 0.3 | 0.04 |
| Chronic obstructive lung disease | 5.3 | 6.1 | -0.04 | 5.8 | 5.1 | 0.03 | 5.7 | 5.4 | 0.01 |
| Dementia | 0.7 | 1.3 | -0.05 | 0.9 | 1.0 | -0.01 | 0.8 | 1.1 | -0.03 |
| Gastroesophageal reflux disease | 2.1 | 2.1 | 0.00 | 2.3 | 1.9 | 0.03 | 2.3 | 2.0 | 0.02 |
| Hyperlipidemia | 22.2 | 24.7 | -0.06 | 22.6 | 22.9 | -0.01 | 23.1 | 23.2 | 0.00 |
| Hypertensive disorder | 42.1 | 42.9 | -0.02 | 41.7 | 40.8 | 0.02 | 42.5 | 41.6 | 0.02 |
| Obesity | 10.2 | 10.7 | -0.02 | 10.1 | 10.6 | -0.02 | 10.2 | 10.5 | -0.01 |
| Renal impairment | 4.6 | 5.7 | -0.05 | 5.4 | 4.7 | 0.03 | 5.6 | 4.8 | 0.04 |
| Urinary tract infectious disease | 4.1 | 4.2 | -0.01 | 4.1 | 3.9 | 0.01 | 4.2 | 4.1 | 0.01 |
| Primary osteoporosis | <0.1 | 0.1 | -0.02 | <0.1 | 0.2 | -0.02 | <0.1 | 0.1 | -0.02 |
| **Medical history: Cardiovascular disease** | | | | | | | | | |
| Atrial fibrillation | 2.6 | 3.6 | -0.06 | 3.1 | 2.9 | 0.01 | 3.3 | 3.0 | 0.02 |
| Cerebrovascular disease | 3.6 | 4.4 | -0.04 | 4.2 | 3.5 | 0.04 | 4.3 | 3.7 | 0.03 |
| Coronary arteriosclerosis | 5.3 | 9.5 | -0.16 | 7.0 | 5.8 | 0.05 | 7.4 | 6.8 | 0.02 |
| Heart disease | 22.4 | 34.1 | -0.26 | 28.2 | 25.1 | 0.07 | 29.0 | 26.7 | 0.05 |
| Heart failure | 6.7 | 9.9 | -0.11 | 8.7 | 7.4 | 0.05 | 8.7 | 7.9 | 0.03 |
| Ischemic heart disease | 9.6 | 17.1 | -0.22 | 12.7 | 10.6 | 0.06 | 13.9 | 12.3 | 0.05 |
| Peripheral vascular disease | 4.9 | 7.1 | -0.09 | 5.8 | 5.5 | 0.01 | 6.0 | 5.8 | 0.01 |
| Pulmonary embolism | 0.4 | 0.7 | -0.04 | 0.6 | 0.7 | -0.01 | 0.5 | 0.6 | -0.01 |
| Venous thrombosis | 1.4 | 1.3 | 0.00 | 1.4 | 1.2 | 0.02 | 1.5 | 1.3 | 0.02 |
| Medical history: Neoplasms | 3.6 | 4.1 | -0.03 | 3.7 | 3.8 | 0.00 | 4.0 | 3.9 | 0.01 |
| **Medication use** | | | | | | | | | |
| Agents acting on the renin-angiotensin system | 67.7 | 73.2 | -0.12 | 70.9 | 70.0 | 0.02 | 70.7 | 70.0 | 0.01 |
| Antibacterials for systemic use | 21.1 | 21.2 | 0.00 | 21.0 | 20.6 | 0.01 | 21.3 | 21.2 | 0.00 |
| Antithrombotic agents | 29.8 | 40.3 | -0.22 | 35.7 | 33.4 | 0.05 | 36.2 | 33.9 | 0.05 |
| Beta blocking agents | 43.1 | 53.2 | -0.20 | 50.0 | 47.5 | 0.05 | 48.8 | 46.9 | 0.04 |
| Calcium channel blockers | 33.1 | 36.1 | -0.06 | 34.6 | 34.5 | 0.00 | 34.9 | 34.4 | 0.01 |
| Diuretics | 44.1 | 50.4 | -0.13 | 48.4 | 46.7 | 0.03 | 48.1 | 46.6 | 0.03 |
| Immunosuppressants | 0.7 | 1.0 | -0.02 | 0.9 | 0.8 | 0.01 | 0.9 | 0.9 | 0.00 |
| Lipid modifying agents | 48.6 | 57.2 | -0.17 | 53.2 | 51.9 | 0.03 | 53.3 | 51.7 | 0.03 |

##### 5.6.3 OptumDOD

**Supplementary Table 71:** Baseline patient characteristics for dapagliflozin (T) and empagliflozin (C) new-users in the OptumDOD data source. We report proportion of initiators satisfying selected base-line characteristics and the standardized difference of population proportions (StdDiff) before and after propensity score adjustment. Less extreme StdDiffs through matching and stratification suggest improved balance between patient cohorts through adjustment. The ”Medication use” entries refer to second-level ATC medication classes.

| | Before adjustment | | | After matching | | | After stratification | | |
|---|---|---|---|---|---|---|---|---|---|
| Characteristic | T | C | StdDiff | T | C | StdDiff | T | C | StdDiff |
| Age: mean | 67.2 | 62.3 | 0.41 | 67.6 | 67.7 | -0.01 | 62.6 | 63.4 | -0.06 |
| Age: std | 11.4 | 11.1 | | 11.9 | 11.2 | | 11.9 | 12.0 | |
| Characteristic (in proportions) | T (%) | C (%) | StdDiff | T (%) | C (%) | StdDiff | T (%) | C (%) | StdDiff |
| Gender: female | 45.4 | 41.2 | 0.09 | 45.1 | 45.6 | -0.01 | 41.9 | 42.1 | 0.00 |
| **Medical history: General** | | | | | | | | | |
| Chronic liver disease | 2.3 | 1.3 | 0.09 | 2.1 | 2.1 | 0.00 | 1.4 | 1.5 | 0.00 |
| Chronic obstructive lung disease | 14.6 | 9.1 | 0.18 | 14.0 | 14.6 | -0.01 | 10.0 | 10.4 | -0.01 |
| Dementia | 2.6 | 1.3 | 0.11 | 2.5 | 2.5 | 0.00 | 1.5 | 1.5 | 0.00 |
| Gastroesophageal reflux disease | 23.6 | 20.3 | 0.08 | 23.5 | 23.6 | 0.00 | 20.5 | 21.0 | -0.01 |
| Hyperlipidemia | 84.7 | 81.8 | 0.08 | 84.5 | 84.9 | -0.01 | 80.8 | 82.5 | -0.04 |
| Hypertensive disorder | 85.5 | 79.8 | 0.15 | 85.4 | 85.8 | -0.01 | 80.7 | 81.1 | -0.01 |
| Obesity | 29.6 | 32.5 | -0.06 | 29.9 | 30.1 | 0.00 | 31.9 | 32.1 | 0.00 |
| Renal impairment | 30.3 | 14.2 | 0.42 | 28.1 | 29.6 | -0.03 | 16.2 | 17.8 | -0.04 |
| Urinary tract infectious disease | 10.0 | 6.9 | 0.12 | 9.4 | 9.3 | 0.00 | 7.4 | 7.5 | 0.00 |
| Acute urinary tract infection | 0.2 | 0.2 | -0.01 | 0.2 | 0.3 | -0.02 | 0.2 | 0.2 | 0.01 |
| Closed fracture of hip | 0.4 | 0.2 | 0.04 | 0.4 | 0.3 | 0.01 | 0.3 | 0.2 | 0.01 |
| Closed fracture of lower limb | 1.6 | 1.3 | 0.03 | 1.6 | 1.6 | 0.00 | 1.4 | 1.4 | 0.00 |
| **Medical history: Cardiovascular disease** | | | | | | | | | |
| Atrial fibrillation | 14.1 | 8.5 | 0.19 | 13.8 | 14.8 | -0.03 | 9.1 | 9.9 | -0.03 |
| Cerebrovascular disease | 9.5 | 6.6 | 0.11 | 9.4 | 9.9 | -0.02 | 6.8 | 7.3 | -0.02 |
| Coronary arteriosclerosis | 27.5 | 21.8 | 0.14 | 27.2 | 28.3 | -0.02 | 20.5 | 23.3 | -0.06 |
| Heart disease | 46.9 | 36.7 | 0.21 | 46.6 | 48.4 | -0.04 | 36.5 | 39.3 | -0.06 |
| Heart failure | 21.9 | 11.2 | 0.31 | 20.8 | 22.1 | -0.03 | 13.4 | 13.8 | -0.01 |
| Ischemic heart disease | 15.7 | 12.0 | 0.11 | 15.4 | 16.3 | -0.02 | 11.4 | 13.0 | -0.05 |
| Peripheral vascular disease | 19.1 | 13.4 | 0.16 | 19.1 | 19.5 | -0.01 | 14.4 | 14.8 | -0.01 |
| Pulmonary embolism | 1.3 | 1.0 | 0.03 | 1.3 | 1.4 | -0.01 | 1.0 | 1.1 | 0.00 |
| Venous thrombosis | 2.1 | 1.6 | 0.04 | 2.0 | 2.1 | -0.01 | 1.6 | 1.7 | -0.01 |
| Medical history: Neoplasms | 12.3 | 10.4 | 0.06 | 12.4 | 13.1 | -0.02 | 9.5 | 11.0 | -0.05 |
| **Medication use** | | | | | | | | | |
| Agents acting on the renin-angiotensin system | 78.7 | 71.6 | 0.16 | 78.3 | 78.9 | -0.01 | 72.5 | 73.2 | -0.02 |
| Antibacterials for systemic use | 55.9 | 53.1 | 0.06 | 54.8 | 55.7 | -0.02 | 52.9 | 53.7 | -0.02 |
| Antithrombotic agents | 27.0 | 21.1 | 0.14 | 26.8 | 27.8 | -0.02 | 20.9 | 22.6 | -0.04 |
| Beta blocking agents | 48.8 | 39.0 | 0.20 | 48.5 | 49.8 | -0.03 | 38.6 | 41.5 | -0.06 |
| Calcium channel blockers | 34.3 | 28.4 | 0.13 | 34.2 | 35.1 | -0.02 | 27.8 | 29.8 | -0.04 |
| Diuretics | 52.8 | 42.6 | 0.21 | 51.9 | 53.4 | -0.03 | 43.8 | 45.0 | -0.02 |
| Immunosuppressants | 3.3 | 3.0 | 0.02 | 3.4 | 3.3 | 0.01 | 3.5 | 3.1 | 0.03 |
| Lipid modifying agents | 83.8 | 80.3 | 0.09 | 84.3 | 84.7 | -0.01 | 79.2 | 81.2 | -0.05 |

##### 5.6.4 MDCR

**Supplementary Table 72:** Baseline patient characteristics for dapagliflozin (T) and empagliflozin (C) new-users in the MDCR data source. We report proportion of initiators satisfying selected base-line characteristics and the standardized difference of population proportions (StdDiff) before and after propensity score adjustment. Less extreme StdDiffs through matching and stratification suggest improved balance between patient cohorts through adjustment. The "Medication use" entries refer to second-level ATC medication classes.

| | Before adjustment | | | After matching | | | After stratification | | |
|---|---|---|---|---|---|---|---|---|---|
| Characteristic | T | C | StdDiff | T | C | StdDiff | T | C | StdDiff |
| Age: mean | 72.8 | 72.9 | -0.03 | 73.0 | 72.6 | 0.08 | 73.6 | 72.8 | 0.13 |
| Age: std | 5.9 | 6.0 | | 5.6 | 5.7 | | 5.6 | 5.7 | |
| Characteristic (in proportions) | T (%) | C (%) | StdDiff | T (%) | C (%) | StdDiff | T (%) | C (%) | StdDiff |
| Gender: female | 42.5 | 40.9 | 0.03 | 41.6 | 43.4 | -0.04 | 42.1 | 41.4 | 0.01 |
| **Medical history: General** | | | | | | | | | |
| Chronic liver disease | 1.8 | 1.3 | 0.04 | 1.6 | 1.4 | 0.01 | 2.1 | 1.4 | 0.06 |
| Chronic obstructive lung disease | 11.5 | 11.9 | -0.01 | 11.7 | 9.8 | 0.06 | 14.2 | 11.4 | 0.09 |
| Dementia | 1.9 | 1.6 | 0.02 | 2.1 | 1.5 | 0.05 | 3.2 | 1.6 | 0.12 |
| Gastroesophageal reflux disease | 17.0 | 19.4 | -0.06 | 18.3 | 18.7 | -0.01 | 20.4 | 19.2 | 0.03 |
| Hyperlipidemia | 82.5 | 84.5 | -0.06 | 83.6 | 83.1 | 0.01 | 85.2 | 84.1 | 0.03 |
| Hypertensive disorder | 85.8 | 86.4 | -0.02 | 86.4 | 85.1 | 0.04 | 88.2 | 86.1 | 0.06 |
| Obesity | 23.4 | 29.3 | -0.13 | 24.4 | 24.4 | 0.00 | 25.0 | 28.0 | -0.07 |
| Renal impairment | 22.6 | 20.8 | 0.04 | 23.8 | 21.7 | 0.05 | 21.8 | 21.2 | 0.01 |
| Urinary tract infectious disease | 8.1 | 7.4 | 0.03 | 7.7 | 8.9 | -0.04 | 6.7 | 7.8 | -0.04 |
| Acute urinary tract infection | <0.4 | 0.2 | 0.01 | <0.5 | 0.2 | -0.01 | <0.4 | 0.2 | -0.01 |
| Closed fracture of hip | <0.4 | 0.4 | -0.04 | <0.5 | 0.4 | -0.03 | <0.4 | 0.4 | -0.04 |
| Closed fracture of lower limb | 1.2 | 1.6 | -0.03 | 1.2 | 1.4 | -0.02 | 1.0 | 1.6 | -0.04 |
| **Medical history: Cardiovascular disease** | | | | | | | | | |
| Atrial fibrillation | 14.0 | 16.8 | -0.07 | 14.8 | 14.7 | 0.00 | 18.5 | 16.2 | 0.06 |
| Cerebrovascular disease | 13.0 | 10.3 | 0.08 | 12.6 | 10.7 | 0.06 | 12.5 | 10.5 | 0.06 |
| Coronary arteriosclerosis | 26.4 | 35.3 | -0.19 | 27.2 | 23.2 | 0.09 | 36.2 | 32.2 | 0.08 |
| Heart disease | 49.1 | 55.3 | -0.13 | 49.6 | 46.9 | 0.05 | 56.0 | 53.2 | 0.06 |
| Heart failure | 15.4 | 17.7 | -0.06 | 16.2 | 15.3 | 0.03 | 20.6 | 17.0 | 0.10 |
| Ischemic heart disease | 12.2 | 17.0 | -0.13 | 12.4 | 12.5 | 0.00 | 16.2 | 16.0 | 0.01 |
| Peripheral vascular disease | 15.9 | 16.9 | -0.03 | 15.9 | 15.2 | 0.02 | 17.0 | 16.4 | 0.02 |
| Pulmonary embolism | 1.1 | 1.1 | 0.00 | 1.0 | 0.9 | 0.00 | 1.4 | 1.1 | 0.04 |
| Venous thrombosis | 2.6 | 2.2 | 0.03 | 2.7 | 2.2 | 0.03 | 2.0 | 2.2 | -0.01 |
| Medical history: Neoplasms | 18.3 | 18.0 | 0.01 | 18.9 | 18.0 | 0.02 | 20.2 | 18.1 | 0.06 |
| **Medication use** | | | | | | | | | |
| Agents acting on the renin-angiotensin system | 76.8 | 77.8 | -0.02 | 77.5 | 78.5 | -0.03 | 75.2 | 78.0 | -0.07 |
| Antibacterials for systemic use | 60.2 | 57.4 | 0.06 | 58.5 | 60.2 | -0.03 | 57.7 | 58.3 | -0.01 |
| Antithrombotic agents | 26.8 | 31.8 | -0.11 | 28.0 | 28.1 | 0.00 | 33.1 | 30.9 | 0.05 |
| Beta blocking agents | 51.1 | 54.0 | -0.06 | 51.7 | 51.9 | 0.00 | 55.5 | 53.4 | 0.04 |
| Calcium channel blockers | 34.7 | 36.0 | -0.03 | 34.5 | 36.9 | -0.05 | 31.0 | 36.5 | -0.12 |
| Diuretics | 51.6 | 51.8 | 0.00 | 51.5 | 52.6 | -0.02 | 49.6 | 51.9 | -0.04 |
| Immunosuppressants | 4.2 | 3.9 | 0.02 | 3.9 | 4.3 | -0.02 | 4.3 | 4.0 | 0.02 |
| Lipid modifying agents | 85.3 | 86.5 | -0.03 | 85.4 | 87.1 | -0.05 | 84.8 | 86.7 | -0.05 |

##### 5.6.5 MDCD

**Supplementary Table 73:** Baseline patient characteristics for dapagliflozin (T) and empagliflozin (C) new-users in the MDCD data source. We report proportion of initiators satisfying selected base-line characteristics and the standardized difference of population proportions (StdDiff) before and after propensity score adjustment. Less extreme StdDiffs through matching and stratification suggest improved balance between patient cohorts through adjustment. The ”Medication use” entries refer to second-level ATC medication classes.

| | Before adjustment | | | After matching | | | After stratification | | |
|---|---|---|---|---|---|---|---|---|---|
| Characteristic | T | C | StdDiff | T | C | StdDiff | T | C | StdDiff |
| Age: mean | 51.0 | 51.0 | 0.01 | 51.0 | 51.2 | -0.02 | 50.8 | 51.1 | -0.03 |
| Age: std | 11.7 | 11.7 | | 11.3 | 11.4 | | 11.3 | 11.3 | |
| Characteristic (in proportions) | T (%) | C (%) | StdDiff | T (%) | C (%) | StdDiff | T (%) | C (%) | StdDiff |
| Gender: female | 56.3 | 56.0 | 0.01 | 55.8 | 57.4 | -0.03 | 55.2 | 56.6 | -0.03 |
| **Race** | | | | | | | | | |
| Black or African American | 25.2 | 23.4 | 0.04 | 24.1 | 26.2 | -0.05 | 22.0 | 24.4 | -0.06 |
| White | 50.9 | 57.0 | -0.12 | 52.2 | 50.7 | 0.03 | 56.7 | 54.5 | 0.04 |
| **Ethnicity** | | | | | | | | | |
| Hispanic or Latino | 7.9 | 5.2 | 0.12 | 7.9 | 7.0 | 0.04 | 6.8 | 6.0 | 0.03 |
| **Medical history: General** | | | | | | | | | |
| Chronic liver disease | 2.9 | 3.1 | -0.01 | 2.5 | 2.8 | -0.02 | 2.6 | 3.0 | -0.03 |
| Chronic obstructive lung disease | 20.0 | 18.1 | 0.05 | 19.3 | 19.3 | 0.00 | 18.2 | 18.5 | -0.01 |
| Dementia | 0.8 | 1.1 | -0.03 | 0.8 | 1.0 | -0.02 | 0.7 | 1.1 | -0.04 |
| Gastroesophageal reflux disease | 26.9 | 28.5 | -0.04 | 26.5 | 27.1 | -0.01 | 26.8 | 28.1 | -0.03 |
| Hyperlipidemia | 67.5 | 66.8 | 0.01 | 67.8 | 64.2 | 0.08 | 67.7 | 66.1 | 0.04 |
| Hypertensive disorder | 75.6 | 75.1 | 0.01 | 75.2 | 75.9 | -0.01 | 74.4 | 75.4 | -0.02 |
| Obesity | 43.6 | 42.2 | 0.03 | 43.0 | 42.7 | 0.01 | 42.6 | 42.6 | 0.00 |
| Renal impairment | 12.4 | 8.9 | 0.12 | 11.6 | 11.0 | 0.02 | 10.7 | 9.8 | 0.03 |
| Urinary tract infectious disease | 8.9 | 7.4 | 0.05 | 8.9 | 8.3 | 0.02 | 8.9 | 7.7 | 0.04 |
| Acute urinary tract infection | 0.5 | 0.4 | 0.02 | 0.5 | 0.4 | 0.01 | 0.5 | 0.4 | 0.02 |
| Closed fracture of hip | <0.3 | 0.2 | -0.01 | <0.3 | 0.3 | -0.03 | <0.3 | 0.2 | -0.02 |
| **Medical history: Cardiovascular disease** | | | | | | | | | |
| Atrial fibrillation | 5.7 | 4.5 | 0.06 | 5.4 | 5.3 | 0.00 | 4.8 | 4.9 | 0.00 |
| Cerebrovascular disease | 5.4 | 4.6 | 0.04 | 5.4 | 4.6 | 0.04 | 5.0 | 4.6 | 0.02 |
| Coronary arteriosclerosis | 14.8 | 15.7 | -0.03 | 14.7 | 16.3 | -0.04 | 13.8 | 16.0 | -0.06 |
| Heart disease | 32.9 | 32.5 | 0.01 | 31.2 | 35.7 | -0.10 | 30.1 | 33.9 | -0.08 |
| Heart failure | 17.7 | 13.0 | 0.13 | 15.7 | 18.1 | -0.06 | 14.2 | 15.2 | -0.03 |
| Ischemic heart disease | 12.5 | 11.1 | 0.04 | 12.1 | 12.0 | 0.00 | 11.7 | 11.5 | 0.00 |
| Peripheral vascular disease | 9.3 | 11.1 | -0.06 | 9.7 | 11.1 | -0.05 | 9.1 | 11.2 | -0.07 |
| Pulmonary embolism | 2.1 | 1.6 | 0.03 | 1.9 | 1.9 | 0.00 | 1.6 | 1.8 | -0.02 |
| Venous thrombosis | 1.4 | 1.8 | -0.03 | 1.4 | 1.9 | -0.04 | 1.3 | 1.8 | -0.04 |
| Medical history: Neoplasms | 6.4 | 5.7 | 0.03 | 6.3 | 5.5 | 0.03 | 6.5 | 5.6 | 0.04 |
| **Medication use** | | | | | | | | | |
| Agents acting on the renin-angiotensin system | 63.8 | 62.0 | 0.04 | 63.0 | 62.9 | 0.00 | 62.2 | 62.5 | -0.01 |
| Antibacterials for systemic use | 62.8 | 61.4 | 0.03 | 61.5 | 62.5 | -0.02 | 60.8 | 61.9 | -0.02 |
| Antithrombotic agents | 27.9 | 29.0 | -0.02 | 27.3 | 28.6 | -0.03 | 27.3 | 29.1 | -0.04 |
| Beta blocking agents | 39.8 | 36.1 | 0.07 | 38.6 | 39.3 | -0.01 | 36.8 | 37.6 | -0.02 |
| Calcium channel blockers | 26.9 | 24.6 | 0.05 | 26.4 | 26.5 | 0.00 | 25.3 | 25.3 | 0.00 |
| Diuretics | 48.9 | 43.7 | 0.10 | 47.6 | 48.6 | -0.02 | 43.6 | 45.7 | -0.04 |
| Immunosuppressants | 2.7 | 2.4 | 0.02 | 2.6 | 2.3 | 0.02 | 2.5 | 2.3 | 0.01 |
| Lipid modifying agents | 70.3 | 71.3 | -0.02 | 70.6 | 68.5 | 0.05 | 72.1 | 70.7 | 0.03 |

##### 5.6.6 Open Claims

**Supplementary Table 74:** Baseline patient characteristics for dapagliflozin (T) and empagliflozin (C) new-users in the Open Claims data source. We report proportion of initiators satisfying selected base-line characteristics and the standardized difference of population proportions (StdDiff) before and after propensity score adjustment. Less extreme StdDiffs through matching and stratification suggest improved balance between patient cohorts through adjustment. The ”Medication use” entries refer to second-level ATC medication classes.

| | Before adjustment | | | After matching | | | After stratification | | |
|---|---|---|---|---|---|---|---|---|---|
| Characteristic | T | C | StdDiff | T | C | StdDiff | T | C | StdDiff |
| Age: mean | 57.8 | 59.4 | -0.14 | 58.3 | 58.4 | -0.01 | 58.7 | 58.9 | -0.02 |
| Age: std | 11.8 | 12.0 | | 11.9 | 11.9 | | 11.9 | 11.9 | |
| Characteristic (in proportions) | T (%) | C (%) | StdDiff | T (%) | C (%) | StdDiff | T (%) | C (%) | StdDiff |
| Gender: female | 45.9 | 43.4 | 0.05 | 45.1 | 45.1 | 0.00 | 44.5 | 44.2 | 0.01 |
| **Medical history: General** | | | | | | | | | |
| Chronic liver disease | 1.0 | 0.7 | 0.03 | 0.8 | 0.8 | 0.00 | 0.8 | 0.8 | 0.00 |
| Chronic obstructive lung disease | 4.2 | 4.7 | -0.02 | 4.5 | 4.4 | 0.00 | 4.6 | 4.6 | 0.00 |
| Dementia | 0.4 | 0.4 | 0.00 | 0.4 | 0.4 | 0.00 | 0.4 | 0.4 | 0.00 |
| Gastroesophageal reflux disease | 8.0 | 8.3 | -0.01 | 8.0 | 8.1 | 0.00 | 8.2 | 8.3 | 0.00 |
| Hyperlipidemia | 44.4 | 43.9 | 0.01 | 43.9 | 43.9 | 0.00 | 44.1 | 44.0 | 0.00 |
| Hypertensive disorder | 49.3 | 48.7 | 0.01 | 49.0 | 48.7 | 0.00 | 49.1 | 48.8 | 0.01 |
| Obesity | 12.3 | 12.4 | 0.00 | 12.2 | 12.2 | 0.00 | 12.4 | 12.4 | 0.00 |
| Renal impairment | 6.0 | 5.4 | 0.03 | 6.3 | 6.5 | -0.01 | 5.6 | 5.8 | -0.01 |
| Urinary tract infectious disease | 4.0 | 3.5 | 0.02 | 3.9 | 3.7 | 0.01 | 3.8 | 3.7 | 0.01 |
| Acute urinary tract infection | 0.1 | 0.1 | 0.00 | 0.1 | 0.1 | 0.00 | 0.1 | 0.1 | 0.00 |
| Closed fracture of hip | 0.1 | 0.1 | 0.00 | 0.1 | 0.1 | 0.00 | 0.1 | 0.1 | 0.00 |
| Closed fracture of lower limb | 0.7 | 0.7 | 0.00 | 0.7 | 0.7 | 0.00 | 0.7 | 0.7 | 0.00 |
| **Medical history: Cardiovascular disease** | | | | | | | | | |
| Atrial fibrillation | 3.9 | 4.3 | -0.02 | 4.1 | 4.0 | 0.00 | 4.2 | 4.2 | 0.00 |
| Cerebrovascular disease | 2.4 | 2.7 | -0.02 | 2.5 | 2.5 | 0.00 | 2.6 | 2.7 | 0.00 |
| Coronary arteriosclerosis | 8.2 | 10.8 | -0.09 | 8.7 | 8.5 | 0.01 | 9.8 | 10.0 | 0.00 |
| Heart disease | 17.8 | 20.4 | -0.06 | 18.6 | 18.4 | 0.00 | 19.5 | 19.7 | 0.00 |
| Heart failure | 5.6 | 5.2 | 0.02 | 5.9 | 5.9 | 0.00 | 5.5 | 5.5 | 0.00 |
| Ischemic heart disease | 4.2 | 5.3 | -0.05 | 4.5 | 4.4 | 0.00 | 4.9 | 4.9 | 0.00 |
| Peripheral vascular disease | 4.1 | 4.9 | -0.04 | 4.4 | 4.3 | 0.00 | 4.6 | 4.6 | 0.00 |
| Pulmonary embolism | 0.4 | 0.5 | -0.01 | 0.5 | 0.4 | 0.00 | 0.5 | 0.5 | 0.00 |
| Venous thrombosis | 0.7 | 0.7 | -0.01 | 0.7 | 0.7 | 0.00 | 0.7 | 0.7 | 0.00 |
| Medical history: Neoplasms | 4.8 | 5.2 | -0.02 | 4.9 | 4.9 | 0.00 | 5.1 | 5.1 | 0.00 |
| **Medication use** | | | | | | | | | |
| Agents acting on the renin-angiotensin system | 69.2 | 68.5 | 0.01 | 69.1 | 69.3 | 0.00 | 68.4 | 68.7 | -0.01 |
| Antibacterials for systemic use | 54.9 | 52.5 | 0.05 | 53.9 | 54.3 | -0.01 | 53.4 | 53.4 | 0.00 |
| Antithrombotic agents | 17.4 | 21.1 | -0.09 | 18.5 | 18.3 | 0.00 | 19.9 | 19.9 | 0.00 |
| Beta blocking agents | 33.2 | 35.6 | -0.05 | 34.1 | 34.2 | 0.00 | 34.7 | 34.9 | -0.01 |
| Calcium channel blockers | 24.5 | 25.8 | -0.03 | 25.2 | 25.1 | 0.00 | 25.3 | 25.4 | 0.00 |
| Diuretics | 41.1 | 39.3 | 0.04 | 41.0 | 41.2 | 0.00 | 39.9 | 40.1 | 0.00 |
| Immunosuppressants | 2.6 | 2.8 | -0.01 | 2.7 | 2.7 | 0.00 | 2.7 | 2.7 | 0.00 |
| Lipid modifying agents | 71.0 | 75.0 | -0.09 | 72.2 | 72.0 | 0.00 | 73.6 | 73.6 | 0.00 |

###### 5.6.7 IMRD

##### 5.6.8 OptumEHR

**Supplementary Table 75:** Baseline patient characteristics for dapagliflozin (T) and empagliflozin (C) new-users in the OptumEHR data source. We report proportion of initiators satisfying selected base-line characteristics and the standardized difference of population proportions (StdDiff) before and after propensity score adjustment. Less extreme StdDiffs through matching and stratification suggest improved balance between patient cohorts through adjustment. The ”Medication use” entries refer to second-level ATC medication classes.

| | Before adjustment | | | After matching | | | After stratification | | |
|---|---|---|---|---|---|---|---|---|---|
| Characteristic | T | C | StdDiff | T | C | StdDiff | T | C | StdDiff |
| Age: mean | 55.7 | 57.8 | -0.18 | 56.4 | 56.4 | -0.01 | 57.2 | 57.2 | 0.00 |
| Age: std | 11.4 | 11.5 | | 11.6 | 11.5 | | 11.6 | 11.6 | |
| Characteristic (in proportions) | T (%) | C (%) | StdDiff | T (%) | C (%) | StdDiff | T (%) | C (%) | StdDiff |
| Gender: female | 46.0 | 42.4 | 0.07 | 44.7 | 44.9 | 0.00 | 43.0 | 43.6 | -0.01 |
| **Race** | | | | | | | | | |
| Asian | 2.1 | 3.0 | -0.05 | 2.3 | 2.4 | -0.01 | 2.5 | 2.7 | -0.02 |
| Black or African American | 12.3 | 10.8 | 0.05 | 12.3 | 12.3 | 0.00 | 11.0 | 11.4 | -0.01 |
| White | 78.4 | 78.3 | 0.00 | 77.9 | 77.9 | 0.00 | 78.8 | 78.2 | 0.01 |
| **Ethnicity** | | | | | | | | | |
| Hispanic or Latino | 6.1 | 6.0 | 0.00 | 6.4 | 6.0 | 0.02 | 6.2 | 6.0 | 0.01 |
| Not Hispanic or Latino | 85.9 | 83.5 | 0.07 | 85.0 | 85.2 | -0.01 | 84.6 | 84.2 | 0.01 |
| **Medical history: General** | | | | | | | | | |
| Chronic liver disease | 1.7 | 1.3 | 0.03 | 1.5 | 1.2 | 0.02 | 1.5 | 1.3 | 0.01 |
| Chronic obstructive lung disease | 4.5 | 5.1 | -0.03 | 4.9 | 4.5 | 0.02 | 5.1 | 4.8 | 0.01 |
| Dementia | 0.3 | 0.5 | -0.03 | 0.3 | 0.4 | -0.01 | 0.3 | 0.5 | -0.02 |
| Gastroesophageal reflux disease | 17.2 | 17.0 | 0.00 | 17.3 | 16.8 | 0.01 | 17.1 | 17.0 | 0.00 |
| Hyperlipidemia | 65.9 | 67.8 | -0.04 | 65.8 | 65.1 | 0.01 | 68.1 | 67.0 | 0.02 |
| Hypertensive disorder | 67.6 | 67.2 | 0.01 | 67.5 | 66.9 | 0.01 | 67.5 | 67.2 | 0.01 |
| Obesity | 32.6 | 33.0 | -0.01 | 32.3 | 32.6 | -0.01 | 32.8 | 32.9 | 0.00 |
| Renal impairment | 5.4 | 6.0 | -0.03 | 5.8 | 5.5 | 0.01 | 5.9 | 5.7 | 0.00 |
| Urinary tract infectious disease | 3.5 | 3.0 | 0.03 | 3.2 | 3.3 | -0.01 | 3.1 | 3.2 | 0.00 |
| Primary osteoporosis | 0.1 | 0.1 | 0.01 | 0.1 | 0.1 | 0.01 | 0.2 | 0.1 | 0.03 |
| Acute urinary tract infection | 0.4 | 0.4 | 0.00 | 0.4 | 0.4 | 0.00 | 0.4 | 0.4 | 0.00 |
| Closed fracture of hip | 0.0 | 0.1 | -0.01 | 0.0 | 0.0 | -0.01 | 0.0 | 0.0 | -0.01 |
| Closed fracture of lower limb | 0.7 | 0.6 | 0.01 | 0.7 | 0.6 | 0.02 | 0.7 | 0.6 | 0.02 |
| **Medical history: Cardiovascular disease** | | | | | | | | | |
| Atrial fibrillation | 3.8 | 4.9 | -0.05 | 4.1 | 4.2 | 0.00 | 4.8 | 4.6 | 0.01 |
| Cerebrovascular disease | 2.1 | 3.0 | -0.06 | 2.2 | 2.4 | -0.01 | 2.8 | 2.8 | 0.00 |
| Coronary arteriosclerosis | 10.9 | 15.2 | -0.12 | 11.8 | 10.9 | 0.03 | 14.9 | 13.6 | 0.04 |
| Heart disease | 20.2 | 25.6 | -0.13 | 21.6 | 21.0 | 0.01 | 25.0 | 23.8 | 0.03 |
| Heart failure | 4.9 | 4.9 | 0.00 | 5.4 | 5.3 | 0.00 | 5.3 | 5.0 | 0.02 |
| Ischemic heart disease | 5.3 | 6.8 | -0.06 | 5.7 | 5.2 | 0.03 | 6.8 | 6.2 | 0.03 |
| Peripheral vascular disease | 5.6 | 6.8 | -0.05 | 6.2 | 5.6 | 0.03 | 6.7 | 6.3 | 0.02 |
| Pulmonary embolism | 0.6 | 0.5 | 0.00 | 0.6 | 0.5 | 0.00 | 0.6 | 0.5 | 0.01 |
| Venous thrombosis | 0.9 | 0.9 | 0.00 | 0.9 | 0.9 | 0.00 | 1.0 | 0.9 | 0.00 |
| Chronic urinary tract infection | 0.1 | 0.1 | 0.00 | 0.1 | 0.1 | 0.00 | 0.1 | 0.1 | 0.00 |
| Medical history: Neoplasms | 4.9 | 5.9 | -0.04 | 5.2 | 5.0 | 0.01 | 5.8 | 5.5 | 0.01 |
| **Medication use** | | | | | | | | | |
| Agents acting on the renin-angiotensin system | 67.1 | 66.5 | 0.01 | 67.0 | 66.8 | 0.01 | 66.7 | 66.5 | 0.00 |
| Antibacterials for systemic use | 42.9 | 40.5 | 0.05 | 41.8 | 42.5 | -0.01 | 40.8 | 41.4 | -0.01 |
| Antithrombotic agents | 34.1 | 39.4 | -0.11 | 35.2 | 34.4 | 0.02 | 38.9 | 37.6 | 0.03 |
| Beta blocking agents | 31.9 | 34.7 | -0.06 | 32.9 | 32.4 | 0.01 | 35.0 | 33.9 | 0.02 |
| Calcium channel blockers | 22.4 | 24.3 | -0.04 | 23.3 | 23.1 | 0.00 | 23.9 | 23.7 | 0.00 |
| Diuretics | 39.3 | 38.0 | 0.03 | 39.1 | 39.5 | -0.01 | 38.5 | 38.6 | 0.00 |
| Immunosuppressants | 2.5 | 2.9 | -0.02 | 2.6 | 2.6 | 0.00 | 2.9 | 2.8 | 0.01 |
| Lipid modifying agents | 68.1 | 74.5 | -0.14 | 69.8 | 68.5 | 0.03 | 73.5 | 72.2 | 0.03 |

#### 5.7 semaglutide vs dapagliflozin: additional covariate balance checks

**Supplementary Table 76:** Additional patient baseline characteristics for semaglutide ("Sema") and dapagliflozin ("Dapa") new-users in CCAE, Open Claims and OptumEHR databases. We examined history of heart failure, use of sacubitril and occurrence of reduced ejection fraction (within OptumEHR) within one year prior to treatment initiation. We reported proportion of initiators satisfying selected base-line characteristics and the standardized difference of population proportions (StdDiff) before and after propensity score adjustment (matching and stratification). After PS adjustment, all these characteristics are balanced between the two arms with absolute StdDiff $< 0.1$.

| | Before adjustment | | | After matching | | | After stratification | | |
|---|---|---|---|---|---|---|---|---|---|
| Characteristic (in %) | Sema | Dapa | StdDiff | Sema | Dapa | StdDiff | Sema | Dapa | StdDiff |
| **CCAE:** | | | | | | | | | |
| Heart failure | 1.8 | 3.2 | -0.09 | 2.6 | 2.8 | -0.01 | 2.6 | 2.7 | -0.01 |
| Sacubitril | 0.2 | 1.0 | -0.11 | 0.4 | 0.7 | -0.04 | 0.4 | 0.8 | -0.05 |
| **Open Claims:** | | | | | | | | | |
| Heart failure | 2.5 | 5.6 | -0.16 | 3.5 | 3.6 | -0.01 | 3.9 | 4.8 | -0.04 |
| Sacubitril | 0.3 | 2.8 | -0.20 | 0.7 | 0.8 | -0.01 | 1.0 | 2.0 | -0.07 |
| **OptumEHR:** | | | | | | | | | |
| Heart failure | 2.5 | 4.9 | -0.13 | 3.9 | 3.9 | 0.00 | 4.5 | 3.7 | 0.04 |
| Sacubitril | 0.2 | 1.9 | -0.17 | 0.5 | 0.6 | -0.02 | 1.4 | 1.1 | 0.03 |
| Reduced ejection fraction | 0.1 | $<$0.1 | 0.02 | $<$0.1 | 0.1 | $<$ 0.01 | 0.1 | $<$ 0.1 | $<$ 0.01 |

### 6 Exposure cohort counts and follow-up time distributions

#### 6.1 Sample sizes and distribution of follow-up times for main exposure cohorts

**Supplementary Table 77:** Population size and on-treatment follow-up time for new-users of each drug within each data source. We report the total patient counts, and the minimum, 10th, 25th, 50th, 75th, and 90th percentiles, and maximum follow-up time (in days) for each exposure cohort.

| | | On-treatment time distribution (in days) | | | | | | |
|---|---|---|---|---|---|---|---|---|
| | Patients | Min | 10% | 25% | Median | 75% | 90% | Max |
| **GLP1RA:** | | | | | | | | |
| **dulaglutide** | | | | | | | | |
| CCAE | 15,697 | 0 | 27 | 65 | 167 | 569 | 729 | 2,782 |
| MDCR | 1,160 | 0 | 0 | 29 | 124 | 470 | 599 | 2,258 |
| MDCD | 2,632 | 0 | 27 | 51 | 122 | 405 | 518 | 1,779 |
| Clinformatic | 12,217 | 0 | 27 | 58 | 138 | 488 | 612 | 2,639 |
| Open Clair | 161,535 | 0 | 27 | 57 | 154 | 567 | 738 | 2,759 |
| OptumEHR | 14,107 | 1 | 29 | 83 | 101 | 314 | 399 | 2,333 |
| **Total:** | **207,348** | | | | | | | |
| **exenatide** | | | | | | | | |
| CCAE | 2,792 | 0 | 27 | 29 | 93 | 397 | 532 | 2,251 |
| MDCR | 263 | 0 | 0 | 27 | 81 | 245 | 401 | 1,744 |
| MDCD | 1,177 | 0 | 27 | 29 | 95 | 355 | 471 | 1,830 |
| Clinformatic | 2,246 | 0 | 27 | 43 | 104 | 418 | 572 | 2,606 |
| Open Clair | 35,632 | 0 | 27 | 29 | 86 | 380 | 519 | 2,866 |
| OptumEHR | 3,367 | 0 | 59 | 83 | 84 | 241 | 304 | 2,781 |
| **Total:** | **45,477** | | | | | | | |
| **semaglutide** | | | | | | | | |
| CCAE | 21,177 | 0 | 27 | 33 | 90 | 330 | 411 | 1,497 |
| MDCR | 1,290 | 0 | 25 | 29 | 83 | 261 | 326 | 1,386 |
| Clinformatic | 18,819 | 0 | 27 | 30 | 89 | 290 | 376 | 1,452 |
| Open Clair | 174,295 | 0 | 27 | 41 | 89 | 329 | 417 | 1,527 |
| CUIMC | 1,250 | 0 | 1 | 32 | 132 | 463 | 598 | 1,516 |
| OptumEHR | 14,064 | 4 | 29 | 60 | 61 | 187 | 237 | 1,328 |
| VA | 4,690 | 0 | 28 | 56 | 134 | 428 | 524 | 1,835 |
| **Total:** | **235,585** | | | | | | | |
| **SGLT2I:** | | | | | | | | |
| **canagliflozin** | | | | | | | | |
| CCAE | 9,352 | 0 | 29 | 58 | 154 | 531 | 712 | 2,832 |
| MDCR | 954 | 0 | 24 | 50 | 144 | 547 | 717 | 2,206 |
| MDCD | 1,879 | 0 | 29 | 29 | 113 | 458 | 609 | 2,053 |
| Clinformatic | 7,414 | 0 | 29 | 56 | 132 | 514 | 683 | 2,902 |
| Open Clair | 111,119 | 0 | 29 | 29 | 128 | 564 | 753 | 2,835 |
| OptumEHR | 10,242 | 0 | 29 | 29 | 51 | 150 | 206 | 2,730 |
| **Total:** | **140,960** | | | | | | | |
| **dapagliflozin** | | | | | | | | |
| CCAE | 17,305 | 0 | 29 | 63 | 160 | 566 | 732 | 2,837 |
| GermanyD/ | 4,729 | 9 | 27 | 97 | 167 | 497 | 649 | 2,751 |
| MDCR | 1,084 | 0 | 29 | 58 | 120 | 403 | 515 | 2,488 |
| MDCD | 1,579 | 0 | 29 | 34 | 94 | 303 | 375 | 1,759 |
| Clinformatic | 7,517 | 0 | 29 | 52 | 105 | 320 | 391 | 2,183 |
| Open Clair | 163,072 | 0 | 29 | 59 | 137 | 516 | 676 | 2,866 |
| IMRD | 1,746 | 0 | 27 | 74 | 224 | 739 | 886 | 1,486 |
| OptumEHR | 11,634 | 2 | 27 | 27 | 30 | 118 | 175 | 2,593 |
| **Total:** | **208,666** | | | | | | | |
| **empagliflozin** | | | | | | | | |
| CCAE | 24,019 | 0 | 29 | 66 | 157 | 528 | 666 | 2,839 |
| GermanyD/ | 4,152 | 9 | 29 | 99 | 186 | 566 | 739 | 2,779 |
| MDCR | 3,399 | 0 | 0 | 29 | 98 | 370 | 462 | 2,299 |
| MDCD | 3,249 | 0 | 29 | 54 | 115 | 379 | 466 | 1,752 |
| Clinformatic | 30,420 | 0 | 29 | 64 | 145 | 470 | 602 | 2,849 |
| Open Clair | 302,829 | 0 | 29 | 66 | 148 | 523 | 678 | 2,838 |
| IMRD | 1,420 | 0 | 27 | 58 | 168 | 549 | 654 | 1,486 |
| OptumEHR | 24,011 | 1 | 27 | 89 | 89 | 285 | 369 | 2,653 |
| **Total:** | **393,499** | | | | | | | |
| **ertugliflozin** | | | | | | | | |
| Open Clair | 13,676 | 1 | 29 | 57 | 122 | 414 | 524 | 1,537 |
| **Total:** | **13,676** | | | | | | | |

#### 6.2 Sample sizes and follow-up times for CVD subgroups

**Supplementary Table 78:** Population size and follow-up time for each new-user cohorts for individual GLP-1RA or SGLT2I within each data source, among patients with established cardiovascular diseases (CVD). We report the total patient counts, and median and interquartile (IQR) times. When executing comparative studies, we have excluded within-database populations with $<$ 1,000 subjects.

| | | | On-treatment time (in days) | | Total follow-up time (in days) | |
|---|---|---|---|---|---|---|
| | | Patients | median | IQR | median | IQR |
| **GLP1RA:** | | | | | | |
| | **dulaglutide** | | | | | |
| | CCAE | 3,372 | 165 | (67 - 563) | 473 | (198 - 1,223) |
| | Clinformatics | 4,234 | 126 | (58 - 456) | 441 | (184 - 1,186) |
| | OptumEHR | 3,017 | 105 | (83 - 328) | 618 | (316 - 1,371) |
| | **Total:** | **10,623** | | | | |
| | **exenatide** | | | | | |
| | CCAE | 605 | 99 | (47 - 480) | 752 | (310 - 1,853) |
| | Clinformatics | 702 | 103 | (46 - 411) | 900 | (424 - 1,702) |
| | **Total:** | **1,307** | | | | |
| | **semaglutide** | | | | | |
| | CCAE | 4,592 | 91 | (33 - 324) | 266 | (111 - 689) |
| | Clinformatics | 6,626 | 87 | (30 - 276) | 246 | (107 - 647) |
| | Open Claims | 47,373 | 89 | (41 - 314) | 355 | (187 - 858) |
| | OptumEHR | 3,255 | 65 | (60 - 196) | 343 | (181 - 778) |
| | VA | 1,781 | 135 | (56 - 452) | 497 | (258 - 896) |
| | **Total:** | **63,627** | | | | |
| **SGLT2I:** | | | | | | |
| | **canagliflozin** | | | | | |
| | CCAE | 2,085 | 149 | (57 - 522) | 835 | (357 - 2,025) |
| | Clinformatics | 2,230 | 120 | (55 - 459) | 962 | (492 - 2,075) |
| | Open Claims | 29,886 | 120 | (29 - 522) | 1,990 | (1,295 - 2,590) |
| | OptumEHR | 2,030 | 58 | (29 - 166) | 1,347 | (679 - 2,292) |
| | **Total:** | **36,231** | | | | |
| | **dapagliflozin** | | | | | |
| | CCAE | 3,940 | 157 | (64 - 529) | 473 | (194 - 1,330) |
| | GermanyDA | 2,049 | 160 | (97 - 475) | 530 | (265 - 1,525) |
| | Clinformatics | 4,217 | 105 | (52 - 304) | 234 | (113 - 533) |
| | Open Claims | 51,303 | 124 | (58 - 449) | 565 | (267 - 1,898) |
| | OptumEHR | 2,887 | 34 | (27 - 120) | 570 | (274 - 1,541) |
| | **Total:** | **64,396** | | | | |
| | **empagliflozin** | | | | | |
| | CCAE | 6,235 | 158 | (68 - 525) | 430 | (173 - 1,080) |
| | GermanyDA | 2,318 | 183 | (99 - 558) | 674 | (315 - 1,471) |
| | Clinformatics | 13,499 | 138 | (63 - 440) | 403 | (168 - 1,039) |
| | Open Claims | 107,208 | 143 | (67 - 501) | 622 | (290 - 1,400) |
| | OptumEHR | 7,351 | 89 | (89 - 292) | 545 | (265 - 1,210) |
| | **Total:** | **136,611** | | | | |
| | **ertugliflozin** | | | | | |
| | Open Claims | 3,447 | 120 | (57 - 413) | 661 | (343 - 1,141) |
| | **Total:** | **3,447** | | | | |

#### 6.3 Criteria for CVD subgroup

The “higher cardiovascular risk’ ’ subgroup included patients who, before the index date (treatment initiation date):

1. had at least 1 condition occurrence of ‘Conditions indicating established cardiovascular disease,’ starting anytime on or before cohort entry start date; allowing events outside observation period.
2. had at least 1 procedure occurrence of ‘Procedures indicating established cardiovascular disease,’ starting anytime on or before cohort entry start date; allowing events outside observation period.

### 7 Outcome incidence across pairwise comparisons and data sources

**Supplementary Table 79:** Exposures and outcome incidence for 3-point MACE in pairwise new-user cohorts under treatment of target (T) vs comparator (C) drugs across data sources. We report total patient counts, total exposure time in 1,000 patient-year (KYr) units, outcome incidence counts, and incidence rates per 1,000 patient-years (per KYr).

| | Patients | | Exposure (KYr) | | Outcomes | | Incidence (per KYr) | |
|---|---|---|---|---|---|---|---|---|
| | T | C | T | C | T | C | T | C |
| **dulaglutide vs semaglutide** | | | | | | | | |
| CCAE | 11,485 | 20,558 | 8,431 | 9,403 | 53 | 67 | 6.29 | 7.13 |
| OptumDOD | 10,087 | 18,093 | 6,494 | 7,567 | 77 | 91 | 11.9 | 12.0 |
| MDCR | 903 | 1,244 | 468 | 473 | 12 | 11 | 25.6 | 23.2 |
| OptumEHR | 12,658 | 13,784 | 6,306 | 4,299 | 37 | 35 | 5.87 | 8.14 |
| **dulaglutide vs dapagliflozin** | | | | | | | | |
| CCAE | 15,244 | 16,369 | 12,092 | 13,072 | 80 | 102 | 6.62 | 7.80 |
| OptumDOD | 11,770 | 6,664 | 7,995 | 3,080 | 94 | 88 | 11.8 | 28.6 |
| MDCR | 1,092 | 986 | 703 | 585 | 15 | 16 | 21.3 | 27.3 |
| MDCD | 2,462 | 1,380 | 1,417 | 640 | 25 | 20 | 17.6 | 31.2 |
| Open Claims | 154,680 | 150,366 | 121,082 | 112,155 | 677 | 740 | 5.59 | 6.60 |
| OptumEHR | 13,833 | 11,130 | 6,910 | 2,591 | 43 | 24 | 6.22 | 9.26 |
| **dulaglutide vs empagliflozin** | | | | | | | | |
| CCAE | 15,244 | 22,756 | 12,092 | 16,624 | 80 | 117 | 6.62 | 7.04 |
| OptumDOD | 11,770 | 28,217 | 7,995 | 18,969 | 94 | 298 | 11.8 | 15.7 |
| MDCR | 1,097 | 3,135 | 704 | 1,630 | 15 | 48 | 21.3 | 29.5 |
| MDCD | 2,464 | 2,906 | 1,417 | 1,527 | 25 | 36 | 17.6 | 23.6 |
| Open Claims | 154,680 | 280,159 | 121,082 | 204,845 | 677 | 1,524 | 5.59 | 7.44 |
| OptumEHR | 13,833 | 23,022 | 6,910 | 10,599 | 43 | 96 | 6.22 | 9.06 |
| **semaglutide vs dapagliflozin** | | | | | | | | |
| CCAE | 20,559 | 10,356 | 9,403 | 7,253 | 67 | 57 | 7.13 | 7.86 |
| OptumDOD | 18,093 | 5,664 | 7,567 | 2,509 | 91 | 83 | 12.0 | 33.1 |
| MDCR | 1,235 | 753 | 473 | 368 | 11 | 13 | 23.2 | 35.3 |
| Open Claims | 167,135 | 105,079 | 78,809 | 65,745 | 392 | 481 | 4.97 | 7.32 |
| OptumEHR | 13,784 | 8,772 | 4,299 | 1,999 | 35 | 19 | 8.14 | 9.50 |
| **semaglutide vs empagliflozin** | | | | | | | | |
| CCAE | 20,558 | 18,129 | 9,403 | 12,698 | 67 | 88 | 7.13 | 6.93 |
| OptumDOD | 18,093 | 24,787 | 7,567 | 15,675 | 91 | 257 | 12.0 | 16.4 |
| MDCR | 1,247 | 2,741 | 473 | 1,252 | 11 | 43 | 23.2 | 34.3 |
| Open Claims | 167,135 | 240,861 | 78,809 | 165,772 | 392 | 1,272 | 4.97 | 7.67 |
| CUIMC | 1,197 | 1,071 | 768 | 836 | 9 | 14 | 11.7 | 16.7 |
| OptumEHR | 13,784 | 21,803 | 4,299 | 10,020 | 35 | 93 | 8.14 | 9.28 |
| VA | 4,490 | 49,308 | 2,654 | 34,108 | 13 | 289 | 4.90 | 8.47 |
| **dapagliflozin vs empagliflozin** | | | | | | | | |
| CCAE | 16,723 | 22,901 | 13,353 | 16,740 | 103 | 117 | 7.71 | 6.99 |
| GermanyDA | 4,729 | 4,152 | 3,559 | 3,442 | 0 | 0 | | |
| OptumDOD | 6,757 | 28,219 | 3,112 | 18,969 | 88 | 298 | 28.3 | 15.7 |
| MDCR | 998 | 3,126 | 595 | 1,630 | 16 | 48 | 26.9 | 29.5 |
| MDCD | 1,387 | 2,909 | 643 | 1,528 | 20 | 36 | 31.1 | 23.6 |
| Open Claims | 153,348 | 281,053 | 115,205 | 205,764 | 757 | 1,527 | 6.57 | 7.42 |
| IMRD | 1,746 | 1,420 | 1,694 | 1,037 | 0 | 0 | | |
| OptumEHR | 11,328 | 23,092 | 2,634 | 10,646 | 24 | 96 | 9.11 | 9.02 |

**Supplementary Table 80:** Exposures and outcome incidence for 3-point MACE in pairwise new-user cohorts under treatment of target (T) vs comparator (C) drugs across data sources. We report total patient counts, total exposure time in 1,000 patient-year (KYr) units, outcome incidence counts, and incidence rates per 1,000 patient-years (per KYr).

| | Patients | | Exposure (KYr) | | Outcomes | | Incidence (per KYr) | |
|---|---|---|---|---|---|---|---|---|
| | T | C | T | C | T | C | T | C |
| **dulaglutide vs semaglutide** | | | | | | | | |
| CCAE | 11,485 | 20,558 | 8,431 | 9,403 | 53 | 67 | 6.29 | 7.13 |
| OptumDOD | 10,087 | 18,093 | 6,494 | 7,567 | 77 | 91 | 11.9 | 12.0 |
| MDCR | 903 | 1,244 | 468 | 473 | 12 | 11 | 25.6 | 23.2 |
| OptumEHR | 12,658 | 13,784 | 6,306 | 4,299 | 37 | 35 | 5.87 | 8.14 |
| **dulaglutide vs dapagliflozin** | | | | | | | | |
| CCAE | 15,244 | 16,369 | 12,092 | 13,072 | 80 | 102 | 6.62 | 7.80 |
| OptumDOD | 11,770 | 6,664 | 7,995 | 3,080 | 94 | 88 | 11.8 | 28.6 |
| MDCR | 1,092 | 986 | 703 | 585 | 15 | 16 | 21.3 | 27.3 |
| MDCD | 2,462 | 1,380 | 1,417 | 640 | 25 | 20 | 17.6 | 31.2 |
| Open Claims | 154,680 | 150,366 | 121,082 | 112,155 | 677 | 740 | 5.59 | 6.60 |
| OptumEHR | 13,833 | 11,130 | 6,910 | 2,591 | 43 | 24 | 6.22 | 9.26 |
| **dulaglutide vs empagliflozin** | | | | | | | | |
| CCAE | 15,244 | 22,756 | 12,092 | 16,624 | 80 | 117 | 6.62 | 7.04 |
| OptumDOD | 11,770 | 28,217 | 7,995 | 18,969 | 94 | 298 | 11.8 | 15.7 |
| MDCR | 1,097 | 3,135 | 704 | 1,630 | 15 | 48 | 21.3 | 29.5 |
| MDCD | 2,464 | 2,906 | 1,417 | 1,527 | 25 | 36 | 17.6 | 23.6 |
| Open Claims | 154,680 | 280,159 | 121,082 | 204,845 | 677 | 1,524 | 5.59 | 7.44 |
| OptumEHR | 13,833 | 23,022 | 6,910 | 10,599 | 43 | 96 | 6.22 | 9.06 |
| **semaglutide vs dapagliflozin** | | | | | | | | |
| CCAE | 20,559 | 10,356 | 9,403 | 7,253 | 67 | 57 | 7.13 | 7.86 |
| OptumDOD | 18,093 | 5,664 | 7,567 | 2,509 | 91 | 83 | 12.0 | 33.1 |
| MDCR | 1,235 | 753 | 473 | 368 | 11 | 13 | 23.2 | 35.3 |
| Open Claims | 167,135 | 105,079 | 78,809 | 65,745 | 392 | 481 | 4.97 | 7.32 |
| OptumEHR | 13,784 | 8,772 | 4,299 | 1,999 | 35 | 19 | 8.14 | 9.50 |
| **semaglutide vs empagliflozin** | | | | | | | | |
| CCAE | 20,558 | 18,129 | 9,403 | 12,698 | 67 | 88 | 7.13 | 6.93 |
| OptumDOD | 18,093 | 24,787 | 7,567 | 15,675 | 91 | 257 | 12.0 | 16.4 |
| MDCR | 1,247 | 2,741 | 473 | 1,252 | 11 | 43 | 23.2 | 34.3 |
| Open Claims | 167,135 | 240,861 | 78,809 | 165,772 | 392 | 1,272 | 4.97 | 7.67 |
| CUIMC | 1,197 | 1,071 | 768 | 836 | 9 | 14 | 11.7 | 16.7 |
| OptumEHR | 13,784 | 21,803 | 4,299 | 10,020 | 35 | 93 | 8.14 | 9.28 |
| VA | 4,490 | 49,308 | 2,654 | 34,108 | 13 | 289 | 4.90 | 8.47 |
| **dapagliflozin vs empagliflozin** | | | | | | | | |
| CCAE | 16,723 | 22,901 | 13,353 | 16,740 | 103 | 117 | 7.71 | 6.99 |
| GermanyDA | 4,729 | 4,152 | 3,559 | 3,442 | 0 | 0 | | |
| OptumDOD | 6,757 | 28,219 | 3,112 | 18,969 | 88 | 298 | 28.3 | 15.7 |
| MDCR | 998 | 3,126 | 595 | 1,630 | 16 | 48 | 26.9 | 29.5 |
| MDCD | 1,387 | 2,909 | 643 | 1,528 | 20 | 36 | 31.1 | 23.6 |
| Open Claims | 153,348 | 281,053 | 115,205 | 205,764 | 757 | 1,527 | 6.57 | 7.42 |
| IMRD | 1,746 | 1,420 | 1,694 | 1,037 | 0 | 0 | | |
| OptumEHR | 11,328 | 23,092 | 2,634 | 10,646 | 24 | 96 | 9.11 | 9.02 |

**Supplementary Table 81:** Exposures and outcome incidence for 3-point MACE in pairwise new-user cohorts under treatment of target (T) vs comparator (C) drugs across data sources. We report total patient counts, total exposure time in 1,000 patient-year (KYr) units, outcome incidence counts, and incidence rates per 1,000 patient-years (per KYr).

| | Patients | | Exposure (KYr) | | Outcomes | | Incidence (per KYr) | |
|---|---|---|---|---|---|---|---|---|
| | T | C | T | C | T | C | T | C |
| **dulaglutide vs semaglutide** | | | | | | | | |
| CCAE | 11,485 | 20,558 | 8,431 | 9,403 | 53 | 67 | 6.29 | 7.13 |
| OptumDOD | 10,087 | 18,093 | 6,494 | 7,567 | 77 | 91 | 11.9 | 12.0 |
| MDCR | 903 | 1,244 | 468 | 473 | 12 | 11 | 25.6 | 23.2 |
| OptumEHR | 12,658 | 13,784 | 6,306 | 4,299 | 37 | 35 | 5.87 | 8.14 |
| **dulaglutide vs dapagliflozin** | | | | | | | | |
| CCAE | 15,244 | 16,369 | 12,092 | 13,072 | 80 | 102 | 6.62 | 7.80 |
| OptumDOD | 11,770 | 6,664 | 7,995 | 3,080 | 94 | 88 | 11.8 | 28.6 |
| MDCR | 1,092 | 986 | 703 | 585 | 15 | 16 | 21.3 | 27.3 |
| MDCD | 2,462 | 1,380 | 1,417 | 640 | 25 | 20 | 17.6 | 31.2 |
| Open Claims | 154,680 | 150,366 | 121,082 | 112,155 | 677 | 740 | 5.59 | 6.60 |
| OptumEHR | 13,833 | 11,130 | 6,910 | 2,591 | 43 | 24 | 6.22 | 9.26 |
| **dulaglutide vs empagliflozin** | | | | | | | | |
| CCAE | 15,244 | 22,756 | 12,092 | 16,624 | 80 | 117 | 6.62 | 7.04 |
| OptumDOD | 11,770 | 28,217 | 7,995 | 18,969 | 94 | 298 | 11.8 | 15.7 |
| MDCR | 1,097 | 3,135 | 704 | 1,630 | 15 | 48 | 21.3 | 29.5 |
| MDCD | 2,464 | 2,906 | 1,417 | 1,527 | 25 | 36 | 17.6 | 23.6 |
| Open Claims | 154,680 | 280,159 | 121,082 | 204,845 | 677 | 1,524 | 5.59 | 7.44 |
| OptumEHR | 13,833 | 23,022 | 6,910 | 10,599 | 43 | 96 | 6.22 | 9.06 |
| **semaglutide vs dapagliflozin** | | | | | | | | |
| CCAE | 20,559 | 10,356 | 9,403 | 7,253 | 67 | 57 | 7.13 | 7.86 |
| OptumDOD | 18,093 | 5,664 | 7,567 | 2,509 | 91 | 83 | 12.0 | 33.1 |
| MDCR | 1,235 | 753 | 473 | 368 | 11 | 13 | 23.2 | 35.3 |
| Open Claims | 167,135 | 105,079 | 78,809 | 65,745 | 392 | 481 | 4.97 | 7.32 |
| OptumEHR | 13,784 | 8,772 | 4,299 | 1,999 | 35 | 19 | 8.14 | 9.50 |
| **semaglutide vs empagliflozin** | | | | | | | | |
| CCAE | 20,558 | 18,129 | 9,403 | 12,698 | 67 | 88 | 7.13 | 6.93 |
| OptumDOD | 18,093 | 24,787 | 7,567 | 15,675 | 91 | 257 | 12.0 | 16.4 |
| MDCR | 1,247 | 2,741 | 473 | 1,252 | 11 | 43 | 23.2 | 34.3 |
| Open Claims | 167,135 | 240,861 | 78,809 | 165,772 | 392 | 1,272 | 4.97 | 7.67 |
| CUIMC | 1,197 | 1,071 | 768 | 836 | 9 | 14 | 11.7 | 16.7 |
| OptumEHR | 13,784 | 21,803 | 4,299 | 10,020 | 35 | 93 | 8.14 | 9.28 |
| VA | 4,490 | 49,308 | 2,654 | 34,108 | 13 | 289 | 4.90 | 8.47 |
| **dapagliflozin vs empagliflozin** | | | | | | | | |
| CCAE | 16,723 | 22,901 | 13,353 | 16,740 | 103 | 117 | 7.71 | 6.99 |
| GermanyDA | 4,729 | 4,152 | 3,559 | 3,442 | 0 | 0 | | |
| OptumDOD | 6,757 | 28,219 | 3,112 | 18,969 | 88 | 298 | 28.3 | 15.7 |
| MDCR | 998 | 3,126 | 595 | 1,630 | 16 | 48 | 26.9 | 29.5 |
| MDCD | 1,387 | 2,909 | 643 | 1,528 | 20 | 36 | 31.1 | 23.6 |
| Open Claims | 153,348 | 281,053 | 115,205 | 205,764 | 757 | 1,527 | 6.57 | 7.42 |
| IMRD | 1,746 | 1,420 | 1,694 | 1,037 | 0 | 0 | | |
| OptumEHR | 11,328 | 23,092 | 2,634 | 10,646 | 24 | 96 | 9.11 | 9.02 |

**Supplementary Table 82:** Exposures and outcome incidence for 3-point MACE in pairwise new-user cohorts under treatment of target (T) vs comparator (C) drugs across data sources. We report total patient counts, total exposure time in 1,000 patient-year (KYr) units, outcome incidence counts, and incidence rates per 1,000 patient-years (per KYr).

| | Patients | | Exposure (KYr) | | Outcomes | | Incidence (per KYr) | |
|---|---|---|---|---|---|---|---|---|
| | T | C | T | C | T | C | T | C |
| **dulaglutide vs semaglutide** | | | | | | | | |
| CCAE | 11,485 | 20,558 | 8,431 | 9,403 | 53 | 67 | 6.29 | 7.13 |
| OptumDOD | 10,087 | 18,093 | 6,494 | 7,567 | 77 | 91 | 11.9 | 12.0 |
| MDCR | 903 | 1,244 | 468 | 473 | 12 | 11 | 25.6 | 23.2 |
| OptumEHR | 12,658 | 13,784 | 6,306 | 4,299 | 37 | 35 | 5.87 | 8.14 |
| **dulaglutide vs dapagliflozin** | | | | | | | | |
| CCAE | 15,244 | 16,369 | 12,092 | 13,072 | 80 | 102 | 6.62 | 7.80 |
| OptumDOD | 11,770 | 6,664 | 7,995 | 3,080 | 94 | 88 | 11.8 | 28.6 |
| MDCR | 1,092 | 986 | 703 | 585 | 15 | 16 | 21.3 | 27.3 |
| MDCD | 2,462 | 1,380 | 1,417 | 640 | 25 | 20 | 17.6 | 31.2 |
| Open Claims | 154,680 | 150,366 | 121,082 | 112,155 | 677 | 740 | 5.59 | 6.60 |
| OptumEHR | 13,833 | 11,130 | 6,910 | 2,591 | 43 | 24 | 6.22 | 9.26 |
| **dulaglutide vs empagliflozin** | | | | | | | | |
| CCAE | 15,244 | 22,756 | 12,092 | 16,624 | 80 | 117 | 6.62 | 7.04 |
| OptumDOD | 11,770 | 28,217 | 7,995 | 18,969 | 94 | 298 | 11.8 | 15.7 |
| MDCR | 1,097 | 3,135 | 704 | 1,630 | 15 | 48 | 21.3 | 29.5 |
| MDCD | 2,464 | 2,906 | 1,417 | 1,527 | 25 | 36 | 17.6 | 23.6 |
| Open Claims | 154,680 | 280,159 | 121,082 | 204,845 | 677 | 1,524 | 5.59 | 7.44 |
| OptumEHR | 13,833 | 23,022 | 6,910 | 10,599 | 43 | 96 | 6.22 | 9.06 |
| **semaglutide vs dapagliflozin** | | | | | | | | |
| CCAE | 20,559 | 10,356 | 9,403 | 7,253 | 67 | 57 | 7.13 | 7.86 |
| OptumDOD | 18,093 | 5,664 | 7,567 | 2,509 | 91 | 83 | 12.0 | 33.1 |
| MDCR | 1,235 | 753 | 473 | 368 | 11 | 13 | 23.2 | 35.3 |
| Open Claims | 167,135 | 105,079 | 78,809 | 65,745 | 392 | 481 | 4.97 | 7.32 |
| OptumEHR | 13,784 | 8,772 | 4,299 | 1,999 | 35 | 19 | 8.14 | 9.50 |
| **semaglutide vs empagliflozin** | | | | | | | | |
| CCAE | 20,558 | 18,129 | 9,403 | 12,698 | 67 | 88 | 7.13 | 6.93 |
| OptumDOD | 18,093 | 24,787 | 7,567 | 15,675 | 91 | 257 | 12.0 | 16.4 |
| MDCR | 1,247 | 2,741 | 473 | 1,252 | 11 | 43 | 23.2 | 34.3 |
| Open Claims | 167,135 | 240,861 | 78,809 | 165,772 | 392 | 1,272 | 4.97 | 7.67 |
| CUIMC | 1,197 | 1,071 | 768 | 836 | 9 | 14 | 11.7 | 16.7 |
| OptumEHR | 13,784 | 21,803 | 4,299 | 10,020 | 35 | 93 | 8.14 | 9.28 |
| VA | 4,490 | 49,308 | 2,654 | 34,108 | 13 | 289 | 4.90 | 8.47 |
| **dapagliflozin vs empagliflozin** | | | | | | | | |
| CCAE | 16,723 | 22,901 | 13,353 | 16,740 | 103 | 117 | 7.71 | 6.99 |
| GermanyDA | 4,729 | 4,152 | 3,559 | 3,442 | 0 | 0 | | |
| OptumDOD | 6,757 | 28,219 | 3,112 | 18,969 | 88 | 298 | 28.3 | 15.7 |
| MDCR | 998 | 3,126 | 595 | 1,630 | 16 | 48 | 26.9 | 29.5 |
| MDCD | 1,387 | 2,909 | 643 | 1,528 | 20 | 36 | 31.1 | 23.6 |
| Open Claims | 153,348 | 281,053 | 115,205 | 205,764 | 757 | 1,527 | 6.57 | 7.42 |
| IMRD | 1,746 | 1,420 | 1,694 | 1,037 | 0 | 0 | | |
| OptumEHR | 11,328 | 23,092 | 2,634 | 10,646 | 24 | 96 | 9.11 | 9.02 |

**Supplementary Table 83:** Exposures and outcome incidence for 3-point MACE in pairwise new-user cohorts under treatment of target (T) vs comparator (C) drugs across data sources. We report total patient counts, total exposure time in 1,000 patient-year (KYr) units, outcome incidence counts, and incidence rates per 1,000 patient-years (per KYr).

| | Patients | | Exposure (KYr) | | Outcomes | | Incidence (per KYr) | |
|---|---|---|---|---|---|---|---|---|
| | T | C | T | C | T | C | T | C |
| **dulaglutide vs semaglutide** | | | | | | | | |
| CCAE | 11,485 | 20,558 | 8,431 | 9,403 | 53 | 67 | 6.29 | 7.13 |
| OptumDOD | 10,087 | 18,093 | 6,494 | 7,567 | 77 | 91 | 11.9 | 12.0 |
| MDCR | 903 | 1,244 | 468 | 473 | 12 | 11 | 25.6 | 23.2 |
| OptumEHR | 12,658 | 13,784 | 6,306 | 4,299 | 37 | 35 | 5.87 | 8.14 |
| **dulaglutide vs dapagliflozin** | | | | | | | | |
| CCAE | 15,244 | 16,369 | 12,092 | 13,072 | 80 | 102 | 6.62 | 7.80 |
| OptumDOD | 11,770 | 6,664 | 7,995 | 3,080 | 94 | 88 | 11.8 | 28.6 |
| MDCR | 1,092 | 986 | 703 | 585 | 15 | 16 | 21.3 | 27.3 |
| MDCD | 2,462 | 1,380 | 1,417 | 640 | 25 | 20 | 17.6 | 31.2 |
| Open Claims | 154,680 | 150,366 | 121,082 | 112,155 | 677 | 740 | 5.59 | 6.60 |
| OptumEHR | 13,833 | 11,130 | 6,910 | 2,591 | 43 | 24 | 6.22 | 9.26 |
| **dulaglutide vs empagliflozin** | | | | | | | | |
| CCAE | 15,244 | 22,756 | 12,092 | 16,624 | 80 | 117 | 6.62 | 7.04 |
| OptumDOD | 11,770 | 28,217 | 7,995 | 18,969 | 94 | 298 | 11.8 | 15.7 |
| MDCR | 1,097 | 3,135 | 704 | 1,630 | 15 | 48 | 21.3 | 29.5 |
| MDCD | 2,464 | 2,906 | 1,417 | 1,527 | 25 | 36 | 17.6 | 23.6 |
| Open Claims | 154,680 | 280,159 | 121,082 | 204,845 | 677 | 1,524 | 5.59 | 7.44 |
| OptumEHR | 13,833 | 23,022 | 6,910 | 10,599 | 43 | 96 | 6.22 | 9.06 |
| **semaglutide vs dapagliflozin** | | | | | | | | |
| CCAE | 20,559 | 10,356 | 9,403 | 7,253 | 67 | 57 | 7.13 | 7.86 |
| OptumDOD | 18,093 | 5,664 | 7,567 | 2,509 | 91 | 83 | 12.0 | 33.1 |
| MDCR | 1,235 | 753 | 473 | 368 | 11 | 13 | 23.2 | 35.3 |
| Open Claims | 167,135 | 105,079 | 78,809 | 65,745 | 392 | 481 | 4.97 | 7.32 |
| OptumEHR | 13,784 | 8,772 | 4,299 | 1,999 | 35 | 19 | 8.14 | 9.50 |
| **semaglutide vs empagliflozin** | | | | | | | | |
| CCAE | 20,558 | 18,129 | 9,403 | 12,698 | 67 | 88 | 7.13 | 6.93 |
| OptumDOD | 18,093 | 24,787 | 7,567 | 15,675 | 91 | 257 | 12.0 | 16.4 |
| MDCR | 1,247 | 2,741 | 473 | 1,252 | 11 | 43 | 23.2 | 34.3 |
| Open Claims | 167,135 | 240,861 | 78,809 | 165,772 | 392 | 1,272 | 4.97 | 7.67 |
| CUIMC | 1,197 | 1,071 | 768 | 836 | 9 | 14 | 11.7 | 16.7 |
| OptumEHR | 13,784 | 21,803 | 4,299 | 10,020 | 35 | 93 | 8.14 | 9.28 |
| VA | 4,490 | 49,308 | 2,654 | 34,108 | 13 | 289 | 4.90 | 8.47 |
| **dapagliflozin vs empagliflozin** | | | | | | | | |
| CCAE | 16,723 | 22,901 | 13,353 | 16,740 | 103 | 117 | 7.71 | 6.99 |
| GermanyDA | 4,729 | 4,152 | 3,559 | 3,442 | 0 | 0 | | |
| OptumDOD | 6,757 | 28,219 | 3,112 | 18,969 | 88 | 298 | 28.3 | 15.7 |
| MDCR | 998 | 3,126 | 595 | 1,630 | 16 | 48 | 26.9 | 29.5 |
| MDCD | 1,387 | 2,909 | 643 | 1,528 | 20 | 36 | 31.1 | 23.6 |
| Open Claims | 153,348 | 281,053 | 115,205 | 205,764 | 757 | 1,527 | 6.57 | 7.42 |
| IMRD | 1,746 | 1,420 | 1,694 | 1,037 | 0 | 0 | | |
| OptumEHR | 11,328 | 23,092 | 2,634 | 10,646 | 24 | 96 | 9.11 | 9.02 |

**Supplementary Table 84:** Exposures and outcome incidence for 3-point MACE in pairwise new-user cohorts under treatment of target (T) vs comparator (C) drugs across data sources. We report total patient counts, total exposure time in 1,000 patient-year (KYr) units, outcome incidence counts, and incidence rates per 1,000 patient-years (per KYr).

| | Patients | | Exposure (KYr) | | Outcomes | | Incidence (per KYr) | |
|---|---|---|---|---|---|---|---|---|
| | T | C | T | C | T | C | T | C |
| **dulaglutide vs semaglutide** | | | | | | | | |
| CCAE | 11,485 | 20,558 | 8,431 | 9,403 | 53 | 67 | 6.29 | 7.13 |
| OptumDOD | 10,087 | 18,093 | 6,494 | 7,567 | 77 | 91 | 11.9 | 12.0 |
| MDCR | 903 | 1,244 | 468 | 473 | 12 | 11 | 25.6 | 23.2 |
| OptumEHR | 12,658 | 13,784 | 6,306 | 4,299 | 37 | 35 | 5.87 | 8.14 |
| **dulaglutide vs dapagliflozin** | | | | | | | | |
| CCAE | 15,244 | 16,369 | 12,092 | 13,072 | 80 | 102 | 6.62 | 7.80 |
| OptumDOD | 11,770 | 6,664 | 7,995 | 3,080 | 94 | 88 | 11.8 | 28.6 |
| MDCR | 1,092 | 986 | 703 | 585 | 15 | 16 | 21.3 | 27.3 |
| MDCD | 2,462 | 1,380 | 1,417 | 640 | 25 | 20 | 17.6 | 31.2 |
| Open Claims | 154,680 | 150,366 | 121,082 | 112,155 | 677 | 740 | 5.59 | 6.60 |
| OptumEHR | 13,833 | 11,130 | 6,910 | 2,591 | 43 | 24 | 6.22 | 9.26 |
| **dulaglutide vs empagliflozin** | | | | | | | | |
| CCAE | 15,244 | 22,756 | 12,092 | 16,624 | 80 | 117 | 6.62 | 7.04 |
| OptumDOD | 11,770 | 28,217 | 7,995 | 18,969 | 94 | 298 | 11.8 | 15.7 |
| MDCR | 1,097 | 3,135 | 704 | 1,630 | 15 | 48 | 21.3 | 29.5 |
| MDCD | 2,464 | 2,906 | 1,417 | 1,527 | 25 | 36 | 17.6 | 23.6 |
| Open Claims | 154,680 | 280,159 | 121,082 | 204,845 | 677 | 1,524 | 5.59 | 7.44 |
| OptumEHR | 13,833 | 23,022 | 6,910 | 10,599 | 43 | 96 | 6.22 | 9.06 |
| **semaglutide vs dapagliflozin** | | | | | | | | |
| CCAE | 20,559 | 10,356 | 9,403 | 7,253 | 67 | 57 | 7.13 | 7.86 |
| OptumDOD | 18,093 | 5,664 | 7,567 | 2,509 | 91 | 83 | 12.0 | 33.1 |
| MDCR | 1,235 | 753 | 473 | 368 | 11 | 13 | 23.2 | 35.3 |
| Open Claims | 167,135 | 105,079 | 78,809 | 65,745 | 392 | 481 | 4.97 | 7.32 |
| OptumEHR | 13,784 | 8,772 | 4,299 | 1,999 | 35 | 19 | 8.14 | 9.50 |
| **semaglutide vs empagliflozin** | | | | | | | | |
| CCAE | 20,558 | 18,129 | 9,403 | 12,698 | 67 | 88 | 7.13 | 6.93 |
| OptumDOD | 18,093 | 24,787 | 7,567 | 15,675 | 91 | 257 | 12.0 | 16.4 |
| MDCR | 1,247 | 2,741 | 473 | 1,252 | 11 | 43 | 23.2 | 34.3 |
| Open Claims | 167,135 | 240,861 | 78,809 | 165,772 | 392 | 1,272 | 4.97 | 7.67 |
| CUIMC | 1,197 | 1,071 | 768 | 836 | 9 | 14 | 11.7 | 16.7 |
| OptumEHR | 13,784 | 21,803 | 4,299 | 10,020 | 35 | 93 | 8.14 | 9.28 |
| VA | 4,490 | 49,308 | 2,654 | 34,108 | 13 | 289 | 4.90 | 8.47 |
| **dapagliflozin vs empagliflozin** | | | | | | | | |
| CCAE | 16,723 | 22,901 | 13,353 | 16,740 | 103 | 117 | 7.71 | 6.99 |
| GermanyDA | 4,729 | 4,152 | 3,559 | 3,442 | 0 | 0 | | |
| OptumDOD | 6,757 | 28,219 | 3,112 | 18,969 | 88 | 298 | 28.3 | 15.7 |
| MDCR | 998 | 3,126 | 595 | 1,630 | 16 | 48 | 26.9 | 29.5 |
| MDCD | 1,387 | 2,909 | 643 | 1,528 | 20 | 36 | 31.1 | 23.6 |
| Open Claims | 153,348 | 281,053 | 115,205 | 205,764 | 757 | 1,527 | 6.57 | 7.42 |
| IMRD | 1,746 | 1,420 | 1,694 | 1,037 | 0 | 0 | | |
| OptumEHR | 11,328 | 23,092 | 2,634 | 10,646 | 24 | 96 | 9.11 | 9.02 |

### 8 HR estimates across data sources and drug-level comparions: general population

#### 8.1 Meta-analytic HR estimates for all drug-level comparisons

**Supplementary Table 85:** Meta-analytic hazard ratio (HR) estimates and their 95% confidence intervals (CIs) for primary cardiovascular effectiveness outcomes between pairwise new-user cohorts of individual GLP-1RAs and SGLT2Is. We present calibrated estimates using the **on-treatment** time-at-risk definition and two PS adjustment approaches: stratification and matching.

| | 3-point MACE | | 4-point MACE | |
|---|---|---|---|---|
| | PS stratification | PS matching | PS stratification | PS matching |
| dulaglutide vs semaglutide | 0.87 (0.58 - 1.29) | 0.75 (0.54 - 1.04) | 1.01 (0.72 - 1.42) | 0.88 (0.59 - 1.31) |
| dulaglutide vs canagliflozin | 0.71 (0.37 - 1.35) | 0.76 (0.44 - 1.31) | 0.67 (0.42 - 1.09) | 0.76 (0.49 - 1.18) |
| dulaglutide vs dapagliflozin | 0.89 (0.80 - 1.00) | 0.90 (0.76 - 1.07) | 0.92 (0.84 - 1.01) | 0.93 (0.81 - 1.08) |
| dulaglutide vs empagliflozin | 0.91 (0.83 - 1.00) | 0.85 (0.75 - 0.96) | 0.94 (0.87 - 1.02) | 0.91 (0.78 - 1.06) |
| exenatide vs canagliflozin | 0.98 (0.82 - 1.16) | 0.98 (0.78 - 1.23) | 0.93 (0.71 - 1.22) | 0.95 (0.79 - 1.15) |
| exenatide vs ertugliflozin | 0.82 (0.52 - 1.29) | 1.00 (0.52 - 1.90) | 1.03 (0.71 - 1.51) | 1.17 (0.66 - 2.05) |
| semaglutide vs dapagliflozin | 0.95 (0.69 - 1.30) | 0.90 (0.73 - 1.10) | 0.83 (0.71 - 0.96) | 0.83 (0.70 - 0.99) |
| semaglutide vs empagliflozin | 1.05 (0.79 - 1.39) | 0.96 (0.76 - 1.22) | 0.95 (0.81 - 1.12) | 0.93 (0.82 - 1.05) |
| canagliflozin vs dapagliflozin | 0.98 (0.87 - 1.12) | 0.89 (0.53 - 1.51) | 1.01 (0.90 - 1.13) | 0.99 (0.86 - 1.15) |
| canagliflozin vs ertugliflozin | 0.96 (0.63 - 1.45) | 0.75 (0.46 - 1.25) | 1.13 (0.79 - 1.61) | 0.82 (0.53 - 1.30) |
| dapagliflozin vs empagliflozin | 1.02 (0.89 - 1.16) | 0.98 (0.88 - 1.10) | 1.06 (0.93 - 1.21) | 1.07 (0.95 - 1.21) |

**Supplementary Table 86:** Meta-analytic hazard ratio (HR) estimates and their 95% confidence intervals (CIs) for cardiovascular effectiveness outcomes between pairwise new-user cohorts of individual GLP-1RAs and SGLT2Is. We present calibrated HR estimates using the **on-treatment** time-at-risk definition with **PS stratification**. MI = myocardial infarction; HHF = hospitalization or ER visit with heart failure.

| | Acute MI | HHF | Stroke | Sudden cardiac death |
|---|---|---|---|---|
| dulaglutide vs semaglutide | 0.81 (0.46 - 1.42) | 1.13 (0.68 - 1.87) | 0.99 (0.56 - 1.76) | |
| dulaglutide vs canagliflozin | 1.63 (0.56 - 4.74) | 0.81 (0.43 - 1.50) | 0.35 (0.15 - 0.83) | |
| dulaglutide vs dapagliflozin | 0.86 (0.70 - 1.06) | 1.00 (0.89 - 1.13) | 0.91 (0.75 - 1.11) | 0.86 (0.63 - 1.17) |
| dulaglutide vs empagliflozin | 0.88 (0.76 - 1.01) | 1.00 (0.91 - 1.10) | 0.98 (0.87 - 1.11) | 1.08 (0.83 - 1.41) |
| exenatide vs canagliflozin | 0.89 (0.70 - 1.12) | 1.04 (0.77 - 1.40) | 1.15 (0.91 - 1.46) | 1.15 (0.66 - 2.02) |
| exenatide vs ertugliflozin | 1.26 (0.63 - 2.51) | 1.71 (0.96 - 3.03) | 0.72 (0.40 - 1.29) | |
| semaglutide vs dapagliflozin | 1.10 (0.55 - 2.19) | 0.79 (0.65 - 0.94) | 0.84 (0.67 - 1.06) | 0.67 (0.44 - 1.02) |
| semaglutide vs empagliflozin | 1.07 (0.70 - 1.64) | 0.96 (0.77 - 1.19) | 0.93 (0.79 - 1.11) | 1.16 (0.73 - 1.84) |
| canagliflozin vs dapagliflozin | 1.07 (0.91 - 1.27) | 1.06 (0.92 - 1.23) | 0.91 (0.70 - 1.17) | 0.97 (0.67 - 1.42) |
| canagliflozin vs ertugliflozin | 1.35 (0.74 - 2.46) | 1.75 (1.03 - 2.95) | 0.79 (0.45 - 1.40) | |
| dapagliflozin vs empagliflozin | 0.97 (0.84 - 1.12) | 1.09 (0.93 - 1.28) | 1.05 (0.89 - 1.24) | 1.23 (0.97 - 1.57) |

**Supplementary Table 87:** Meta-analytic hazard ratio (HR) estimates and their 95% confidence intervals (CIs) for cardiovascular effectiveness outcomes between pairwise new-user cohorts of individual GLP-1RAs and SGLT2Is. We present calibrated HR estimates using the **on-treatment** time-at-risk definition with **PS matching**. MI = myocardial infarction; HHF = hospitalization or ER visit with heart failure.

| | Acute MI | HHF | Stroke | Sudden cardiac death |
|---|---|---|---|---|
| dulaglutide vs exenatide | 0.77 (0.33 - 1.77) | 1.23 (0.74 - 2.02) | 0.56 (0.26 - 1.19) | |
| dulaglutide vs semaglutide | 0.69 (0.44 - 1.09) | 1.08 (0.77 - 1.52) | 0.86 (0.54 - 1.38) | 1.40 (0.46 - 4.25) |
| dulaglutide vs canagliflozin | 1.41 (0.70 - 2.85) | 1.00 (0.62 - 1.60) | 0.68 (0.21 - 2.15) | |
| dulaglutide vs dapagliflozin | 0.92 (0.74 - 1.16) | 0.97 (0.81 - 1.17) | 0.92 (0.74 - 1.16) | 0.76 (0.46 - 1.27) |
| dulaglutide vs empagliflozin | 0.86 (0.73 - 1.01) | 0.97 (0.76 - 1.23) | 0.87 (0.74 - 1.03) | 1.05 (0.74 - 1.49) |
| dulaglutide vs ertugliflozin | 1.07 (0.59 - 1.93) | 1.50 (0.87 - 2.57) | 0.74 (0.45 - 1.21) | 2.24 (0.45 - 11.2) |
| exenatide vs semaglutide | 1.44 (0.80 - 2.57) | 1.20 (0.77 - 1.87) | 1.11 (0.64 - 1.91) | |
| exenatide vs canagliflozin | 0.87 (0.63 - 1.22) | 1.01 (0.77 - 1.31) | 1.08 (0.72 - 1.62) | 0.84 (0.37 - 1.88) |
| exenatide vs dapagliflozin | 1.53 (0.55 - 4.24) | 0.98 (0.51 - 1.90) | 0.96 (0.67 - 1.37) | 0.56 (0.22 - 1.45) |
| exenatide vs empagliflozin | 1.01 (0.39 - 2.64) | 0.91 (0.57 - 1.44) | 1.36 (0.70 - 2.63) | |
| exenatide vs ertugliflozin | 1.37 (0.50 - 3.78) | 2.44 (0.99 - 5.97) | 1.06 (0.45 - 2.47) | |
| semaglutide vs canagliflozin | 0.80 (0.50 - 1.27) | 0.69 (0.48 - 0.98) | 0.92 (0.57 - 1.48) | 0.61 (0.27 - 1.38) |
| semaglutide vs dapagliflozin | 0.89 (0.68 - 1.16) | 0.75 (0.60 - 0.93) | 0.83 (0.64 - 1.10) | 0.76 (0.42 - 1.37) |
| semaglutide vs empagliflozin | 0.93 (0.72 - 1.22) | 0.93 (0.67 - 1.28) | 0.88 (0.72 - 1.08) | 1.21 (0.79 - 1.85) |
| semaglutide vs ertugliflozin | 0.90 (0.48 - 1.71) | 1.35 (0.76 - 2.39) | 0.60 (0.35 - 1.02) | |
| canagliflozin vs dapagliflozin | 1.10 (0.86 - 1.41) | 1.05 (0.85 - 1.29) | 1.00 (0.25 - 3.91) | 0.90 (0.49 - 1.63) |
| canagliflozin vs empagliflozin | 0.40 (0.18 - 0.88) | 0.76 (0.48 - 1.20) | 1.42 (0.19 - 10.8) | 3.83 (1.10 - 13.3) |
| canagliflozin vs ertugliflozin | 0.89 (0.43 - 1.82) | 1.41 (0.67 - 3.00) | 0.68 (0.35 - 1.31) | |
| dapagliflozin vs empagliflozin | 0.98 (0.79 - 1.21) | 1.23 (1.00 - 1.52) | 1.06 (0.91 - 1.24) | 0.99 (0.73 - 1.32) |
| dapagliflozin vs ertugliflozin | 1.04 (0.57 - 1.91) | 1.49 (0.86 - 2.57) | 0.76 (0.46 - 1.25) | 2.07 (0.55 - 7.82) |
| empagliflozin vs ertugliflozin | 1.17 (0.71 - 1.91) | 1.30 (0.82 - 2.07) | 0.64 (0.42 - 0.97) | 1.87 (0.51 - 6.84) |

**Supplementary Table 88:** Meta-analytic hazard ratio (HR) estimates and their 95% confidence intervals (CIs) for primary cardiovascular effectiveness outcomes between pairwise new-user cohorts of individual GLP-1RAs and SGLT2Is. We present calibrated estimates using the **intent-to-treat** time-at-risk definition and two PS adjustment approaches: stratification and matching.

| | 3-point MACE | | 4-point MACE | |
|---|---|---|---|---|
| | PS stratification | PS matching | PS stratification | PS matching |
| dulaglutide vs semaglutide | 1.01 (0.76 - 1.34) | 0.95 (0.79 - 1.13) | 1.11 (0.88 - 1.41) | 1.01 (0.87 - 1.17) |
| dulaglutide vs canagliflozin | 0.94 (0.72 - 1.22) | 0.99 (0.79 - 1.25) | 0.97 (0.79 - 1.20) | 1.00 (0.81 - 1.23) |
| dulaglutide vs dapagliflozin | 1.01 (0.89 - 1.15) | 1.01 (0.90 - 1.12) | 1.04 (0.91 - 1.19) | 1.04 (0.93 - 1.15) |
| dulaglutide vs empagliflozin | 1.00 (0.91 - 1.09) | 0.98 (0.90 - 1.06) | 1.01 (0.93 - 1.10) | 0.97 (0.86 - 1.09) |
| exenatide vs canagliflozin | 0.93 (0.82 - 1.06) | 0.95 (0.87 - 1.03) | 0.95 (0.84 - 1.08) | 0.96 (0.87 - 1.06) |
| exenatide vs ertugliflozin | 1.00 (0.79 - 1.26) | 1.04 (0.80 - 1.36) | 1.11 (0.91 - 1.35) | 1.18 (0.95 - 1.48) |
| semaglutide vs dapagliflozin | 0.92 (0.83 - 1.03) | 1.04 (0.91 - 1.18) | 0.94 (0.85 - 1.04) | 0.99 (0.89 - 1.11) |
| semaglutide vs empagliflozin | 1.00 (0.83 - 1.21) | 0.97 (0.84 - 1.13) | 0.98 (0.83 - 1.17) | 0.98 (0.85 - 1.14) |
| canagliflozin vs dapagliflozin | 1.05 (1.00 - 1.11) | 1.06 (1.00 - 1.12) | 1.07 (1.02 - 1.12) | 1.07 (1.01 - 1.12) |
| canagliflozin vs ertugliflozin | 1.08 (0.87 - 1.34) | 1.03 (0.80 - 1.32) | 1.12 (0.93 - 1.35) | 1.09 (0.87 - 1.37) |
| dapagliflozin vs empagliflozin | 0.99 (0.93 - 1.05) | 0.99 (0.93 - 1.05) | 0.97 (0.92 - 1.03) | 0.97 (0.92 - 1.03) |

**Supplementary Table 89:** Meta-analytic hazard ratio (HR) estimates and their 95% confidence intervals (CIs) for cardiovascular effectiveness outcomes between pairwise new-user cohorts of individual GLP-1RAs and SGLT2Is. We present calibrated HR estimates using the **intent-to-treat** time-at-risk definition with **PS stratification**. MI = myocardial infarction; HHF = hospitalization or ER visit with heart failure.

| | Acute MI | HHF | Stroke | Sudden cardiac death |
|---|---|---|---|---|
| dulaglutide vs semaglutide | 1.08 (0.73 - 1.61) | 1.22 (0.88 - 1.68) | 0.93 (0.63 - 1.36) | 0.53 (0.21 - 1.36) |
| dulaglutide vs canagliflozin | 1.02 (0.73 - 1.45) | 1.02 (0.79 - 1.32) | 0.86 (0.59 - 1.25) | 0.86 (0.41 - 1.81) |
| dulaglutide vs dapagliflozin | 1.01 (0.88 - 1.15) | 1.10 (0.94 - 1.28) | 1.01 (0.88 - 1.16) | 0.98 (0.78 - 1.24) |
| dulaglutide vs empagliflozin | 0.97 (0.88 - 1.07) | 1.02 (0.90 - 1.16) | 1.02 (0.93 - 1.13) | 1.05 (0.91 - 1.21) |
| exenatide vs canagliflozin | 0.89 (0.75 - 1.05) | 1.06 (0.98 - 1.15) | 0.97 (0.88 - 1.06) | 1.09 (0.94 - 1.25) |
| exenatide vs ertugliflozin | 1.28 (0.91 - 1.79) | 1.33 (1.02 - 1.72) | 0.85 (0.63 - 1.15) | 1.48 (0.79 - 2.76) |
| semaglutide vs dapagliflozin | 0.90 (0.74 - 1.10) | 0.95 (0.84 - 1.07) | 0.95 (0.83 - 1.09) | 1.23 (0.38 - 4.00) |
| semaglutide vs empagliflozin | 0.98 (0.74 - 1.30) | 1.01 (0.84 - 1.22) | 1.03 (0.84 - 1.26) | 1.05 (0.86 - 1.28) |
| canagliflozin vs dapagliflozin | 1.05 (0.98 - 1.12) | 1.10 (1.04 - 1.16) | 1.07 (1.00 - 1.14) | 1.11 (0.99 - 1.25) |
| canagliflozin vs ertugliflozin | 1.35 (1.00 - 1.83) | 1.19 (0.93 - 1.53) | 0.93 (0.70 - 1.24) | 1.13 (0.63 - 2.05) |
| dapagliflozin vs empagliflozin | 0.94 (0.87 - 1.01) | 0.96 (0.90 - 1.02) | 1.02 (0.95 - 1.10) | 1.01 (0.74 - 1.36) |

**Supplementary Table 90:** Meta-analytic hazard ratio (HR) estimates and their 95% confidence intervals (CIs) for cardiovascular effectiveness outcomes between pairwise new-user cohorts of individual GLP-1RAs and SGLT2Is. We present calibrated HR estimates using the **intent-to-treat** time-at-risk definition with **PS matching**. MI = myocardial infarction; HHF = hospitalization or ER visit with heart failure.

| | Acute MI | HHF | Stroke | Sudden cardiac death |
|---|---|---|---|---|
| dulaglutide vs exenatide | 0.92 (0.69 - 1.23) | 1.01 (0.82 - 1.24) | 1.05 (0.78 - 1.40) | 1.51 (0.63 - 3.64) |
| dulaglutide vs semaglutide | 0.90 (0.69 - 1.17) | 1.01 (0.84 - 1.22) | 1.03 (0.80 - 1.32) | 0.88 (0.55 - 1.41) |
| dulaglutide vs canagliflozin | 0.95 (0.71 - 1.27) | 0.99 (0.78 - 1.26) | 1.07 (0.80 - 1.43) | 0.93 (0.57 - 1.54) |
| dulaglutide vs dapagliflozin | 0.97 (0.85 - 1.11) | 1.10 (0.98 - 1.23) | 1.04 (0.92 - 1.19) | 0.82 (0.66 - 1.01) |
| dulaglutide vs empagliflozin | 0.97 (0.87 - 1.07) | 1.06 (0.91 - 1.23) | 0.97 (0.87 - 1.07) | 1.00 (0.85 - 1.19) |
| dulaglutide vs ertugliflozin | 1.14 (0.85 - 1.52) | 1.13 (0.88 - 1.44) | 0.83 (0.63 - 1.08) | 1.14 (0.65 - 1.98) |
| exenatide vs semaglutide | 1.29 (1.01 - 1.65) | 1.32 (1.11 - 1.58) | 1.09 (0.87 - 1.37) | 1.04 (0.67 - 1.59) |
| exenatide vs canagliflozin | 0.90 (0.78 - 1.06) | 1.06 (0.97 - 1.15) | 0.96 (0.87 - 1.05) | 1.00 (0.86 - 1.18) |
| exenatide vs dapagliflozin | 1.04 (0.63 - 1.71) | 1.18 (1.06 - 1.31) | 1.01 (0.90 - 1.14) | 1.14 (0.93 - 1.38) |
| exenatide vs empagliflozin | 0.88 (0.67 - 1.16) | 0.97 (0.80 - 1.18) | 0.92 (0.69 - 1.21) | 0.90 (0.49 - 1.66) |
| exenatide vs ertugliflozin | 1.54 (1.02 - 2.33) | 1.44 (1.06 - 1.96) | 0.92 (0.65 - 1.31) | 1.62 (0.78 - 3.36) |
| semaglutide vs canagliflozin | 0.77 (0.63 - 0.94) | 0.81 (0.69 - 0.95) | 0.94 (0.73 - 1.20) | 1.13 (0.77 - 1.67) |
| semaglutide vs dapagliflozin | 1.01 (0.86 - 1.20) | 0.96 (0.84 - 1.10) | 0.96 (0.82 - 1.13) | 0.85 (0.65 - 1.12) |
| semaglutide vs empagliflozin | 0.99 (0.77 - 1.27) | 1.05 (0.87 - 1.27) | 0.96 (0.83 - 1.11) | 1.16 (0.93 - 1.44) |
| semaglutide vs ertugliflozin | 0.89 (0.62 - 1.26) | 1.09 (0.81 - 1.46) | 0.76 (0.55 - 1.04) | 1.47 (0.79 - 2.75) |
| canagliflozin vs dapagliflozin | 1.05 (0.97 - 1.13) | 1.06 (1.00 - 1.13) | 1.07 (1.00 - 1.16) | 1.09 (0.95 - 1.25) |
| canagliflozin vs empagliflozin | 0.78 (0.62 - 0.97) | 0.90 (0.76 - 1.07) | 1.04 (0.68 - 1.59) | 0.96 (0.62 - 1.50) |
| canagliflozin vs ertugliflozin | 1.32 (0.91 - 1.90) | 1.23 (0.91 - 1.65) | 0.84 (0.61 - 1.16) | 1.16 (0.59 - 2.25) |
| dapagliflozin vs empagliflozin | 0.94 (0.87 - 1.02) | 1.00 (0.90 - 1.11) | 1.03 (0.95 - 1.12) | 1.15 (1.00 - 1.31) |
| dapagliflozin vs ertugliflozin | 1.05 (0.78 - 1.42) | 1.05 (0.83 - 1.33) | 0.88 (0.68 - 1.15) | 1.65 (0.94 - 2.88) |
| empagliflozin vs ertugliflozin | 1.15 (0.90 - 1.48) | 1.04 (0.84 - 1.28) | 0.75 (0.60 - 0.94) | 1.23 (0.74 - 2.05) |

**Supplementary Table 91:** Comparing Meta-analysis 1 and Meta-analysis 2 HR estimates for select drug-level comparisons and 3-pt MACE, with calibrated HR estimates, 95% CIs, and data sources included for each meta-analysis. The two meta-analyses used different study diagnostics thresholds for including data sources in the meta-analysis. Meta-analysis 1 included data sources with equipoise $> 0.25$, max SMD $< 0.15$, MDRR $< 4$; Meta-analysis 2 included data sources with equipoise $> 0.5$, max SMD $< 0.1$, MDRR $< 10$.

| | Meta-analysis 1 | | Meta-analysis 2 | |
|---|---|---|---|---|
| Comparison | HR (95% CI) | Data sources | HR (95% CI) | Data sources |
| semaglutide vs dapagliflozin | 0.95 (0.69, 1.30) | CCAE, OPENCLAIMS | 0.95 (0.69, 1.30) | CCAE, OPENCLAIMS |
| semaglutide vs empagliflozin | 1.05 (0.79, 1.39) | CCAE, OPENCLAIMS, Clinformatics | 1.07 (0.66, 1.74) | CCAE, OPENCLAIMS |
| dapagliflozin vs empagliflozin | 1.02 (0.89, 1.16) | CCAE, OPENCLAIMS, Clinformatics, OptumEHR | 1.00 (0.90, 1.11) | CCAE, OPENCLAIMS, OptumEHR |

**Supplementary Table 92:** Meta-analytic HR estimates for 3-pt and 4-pt MACE under two time-at-risk designs: on-treatment (shown in main paper) and on-treatment censored (follow-up was censored at treatment escalation or switching). Results are presented for select pairwise comparisons generated with PS stratification.

| | 3-pt MACE | | 4-pt MACE | |
|---|---|---|---|---|
| Comparison | On-treatment (main) | On-treatment censored | On-treatment (main) | On-treatment censored |
| semaglutide vs dapagliflozin | 0.95 (0.69, 1.30) | 0.58 (0.40, 0.83) | 0.83 (0.71, 0.96) | 0.65 (0.48, 0.87) |
| semaglutide vs empagliflozin | 1.05 (0.79, 1.39) | 0.67 (0.51, 0.88) | 0.95 (0.81, 1.12) | 0.77 (0.62, 0.95) |
| dapagliflozin vs empagliflozin | 1.02 (0.89, 1.16) | 1.31 (0.97, 1.76) | 1.06 (0.93, 1.21) | 1.21 (0.92, 1.58) |

#### 8.2 Under the on-treatment (OT) time-at-risk definition

##### 8.2.1 HR estimates using propensity score (PS) stratification

In the plot below, we show relative risk estimates of cardiovascular effectiveness outcomes between new-user cohorts of GLP-1RA and SGLT2I individual drugs. Each column shows one outcome, and drug-vs-drug comparisons are shown in rows. We plot calibrated hazard ratios (circles) and their 95 % confidence intervals (whiskers) using the on-treatment (OT) time-at-risk (TAR) definition with PS stratification. Estimates are presented across data sources as well as meta-analytic estimates (when $> 1$ data sources produced estimates). Grayed out entries did not pass study diagnostics.

**Supplementary Figure 36: HR estimates using PS stratification.** Each column shows one outcome, and drug-vs-drug comparisons are shown in rows. Grayed out entries did not pass study diagnostics.

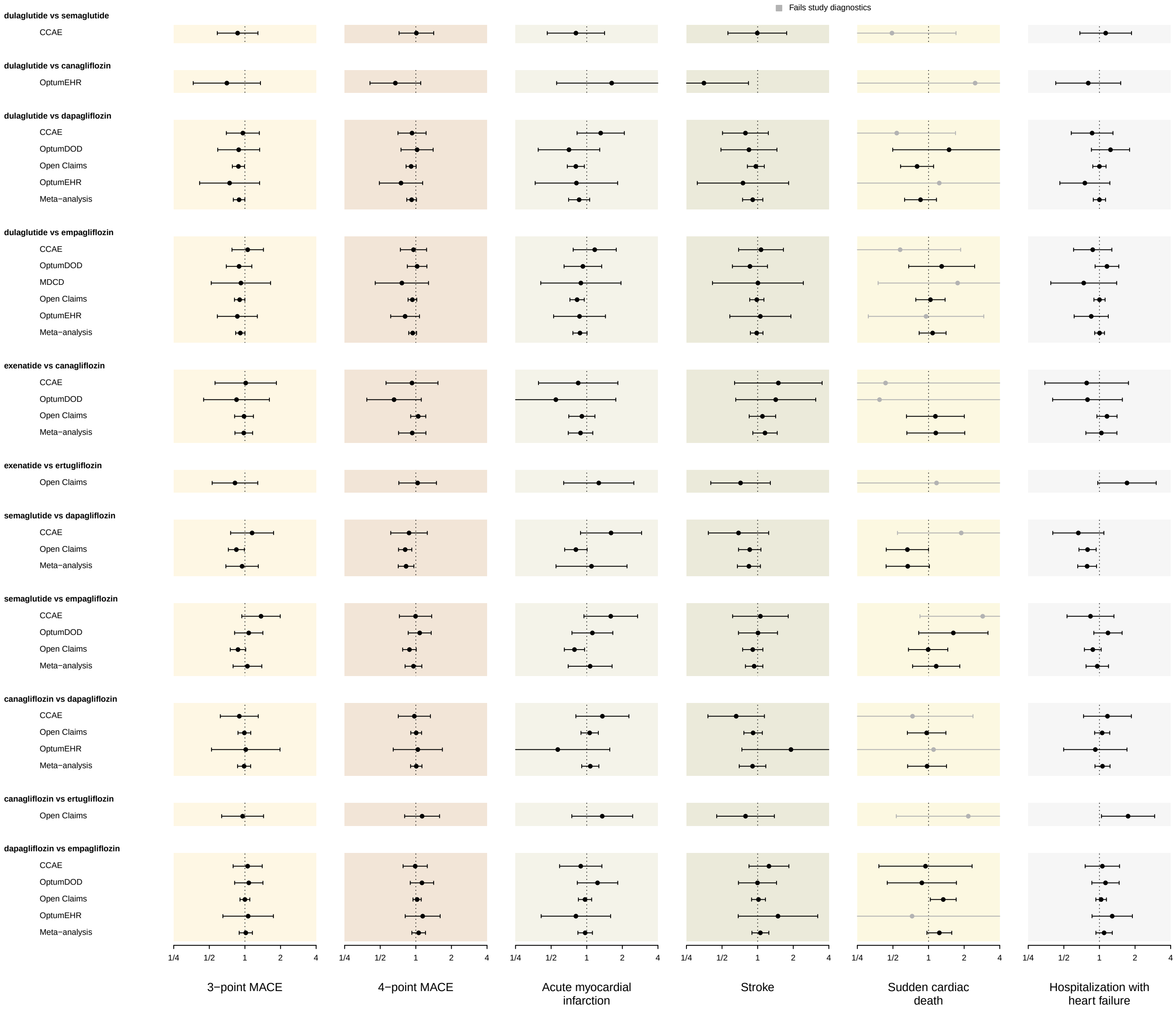

##### 8.2.2 HR estimates using propensity score (PS) matching

In the plots below, we show relative risk estimates of cardiovascular effectiveness outcomes between new-user cohorts of GLP-1RA and SGLT2I individual drugs. Each column shows one outcome, and drug-vs-drug comparisons are shown in rows. We plot calibrated hazard ratios (circles) and their 95 % confidence intervals (whiskers) using the on-treatment (OT) time-at-risk (TAR) definition with PS matching. Estimates are presented across data sources as well as meta-analytic estimates (when $> 1$ data sources produced estimates). Grayed out entries did not pass study diagnostics.

**Supplementary Figure 37: HR estimates using PS matching, part 1.** Each column shows one outcome, and drug-vs-drug comparisons are shown in rows. Grayed out entries did not pass study diagnostics.

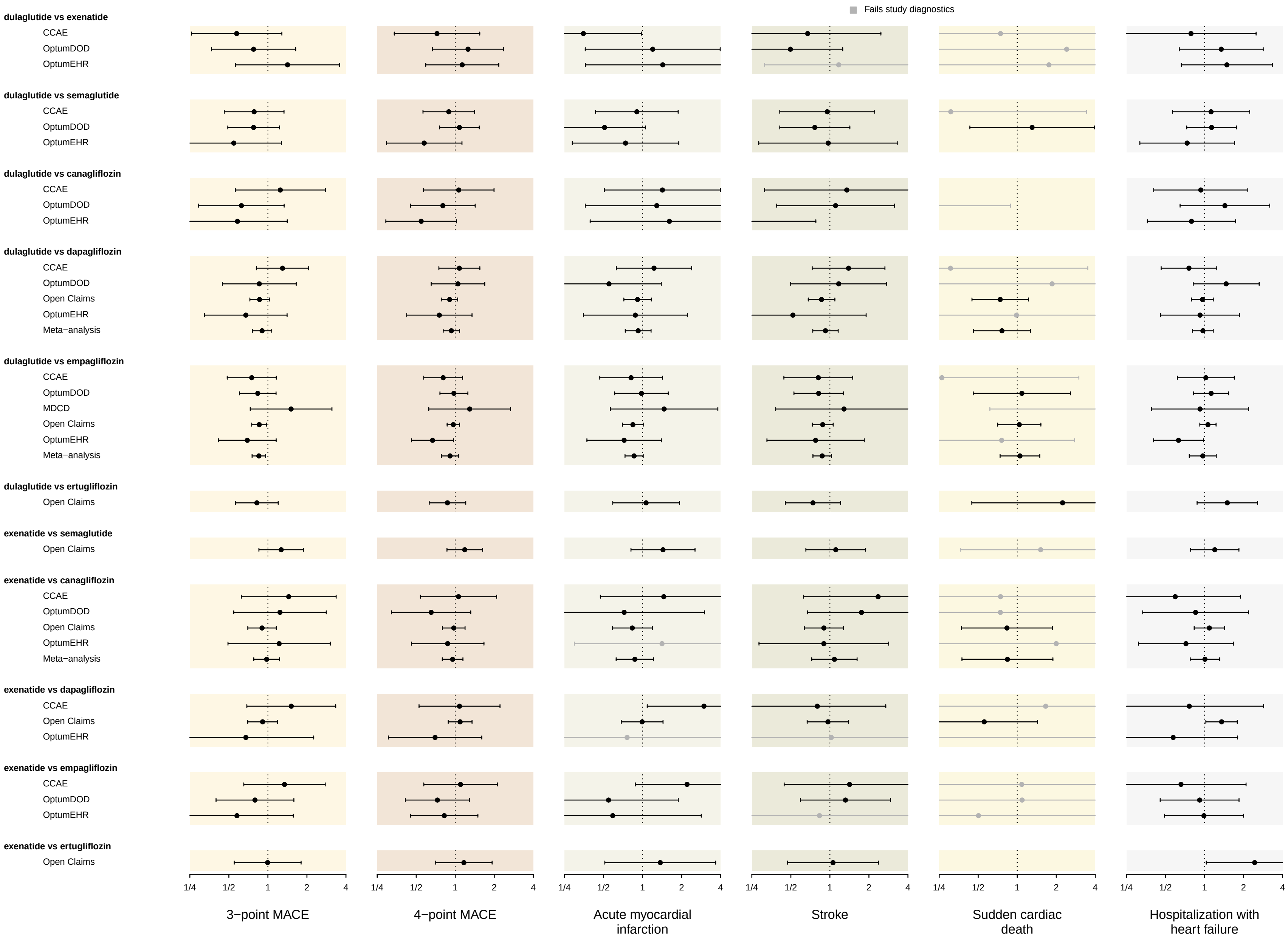

**Supplementary Figure 38: HR estimates using PS matching, part 2.** Each column shows one outcome, and drug-vs-drug comparisons are shown in rows. Grayed out entries did not pass study diagnostics.

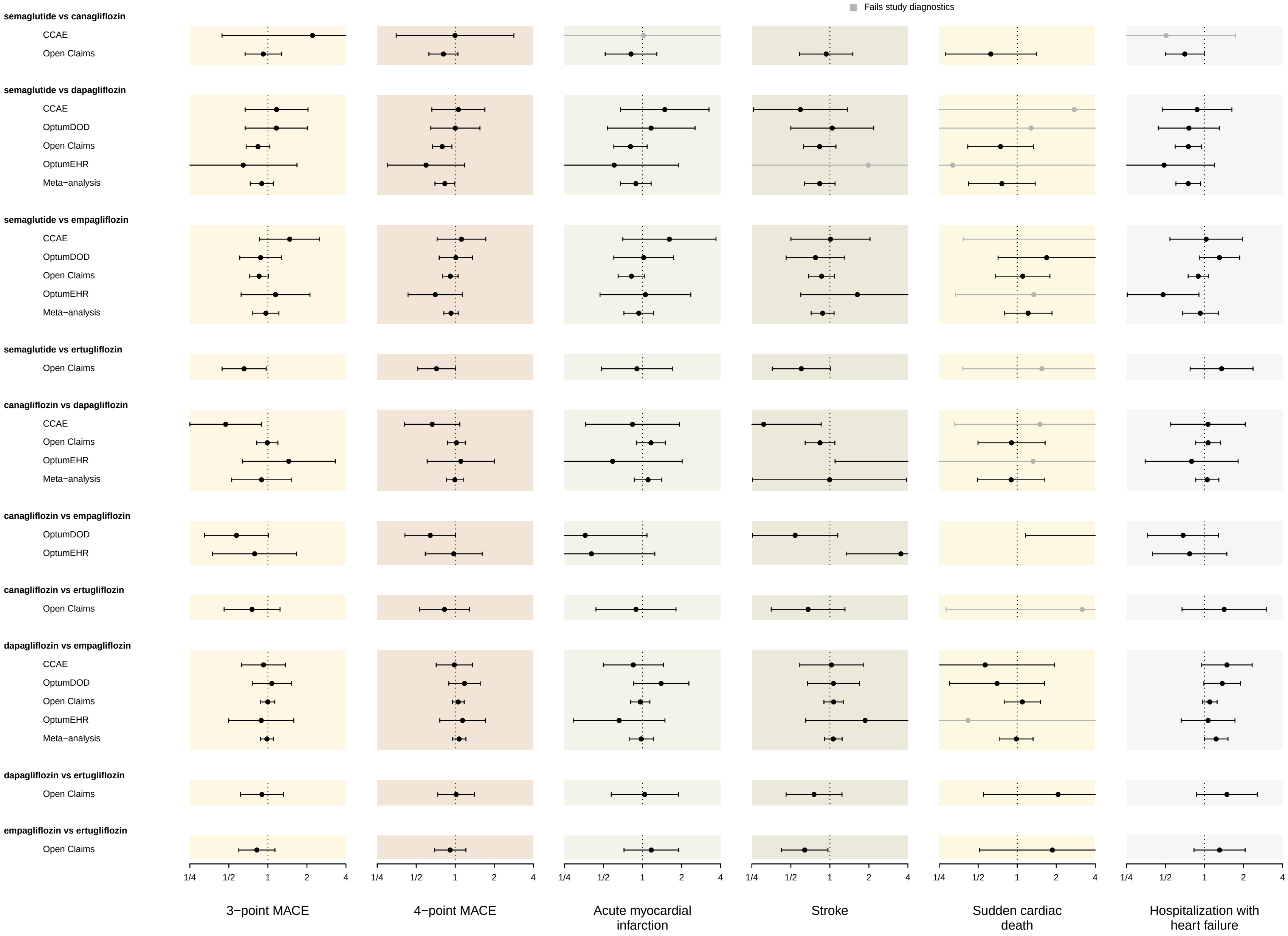

#### 8.3 Under the intent-to-treat (ITT) time-at-risk definitions

##### 8.3.1 HR estimates using PS stratification

In the plot below, we show relative risk estimates of cardiovascular effectiveness outcomes between new-user cohorts of GLP-1RA and SGLT2I individual drugs. Each column shows one outcome, and drug-vs-drug comparisons are shown in rows. We plot calibrated hazard ratios (circles) and their 95 % confidence intervals (whiskers) using the intent-to-treat (ITT) time-at-risk (TAR) definition with PS stratification. Estimates are presented across data sources as well as meta-analytic estimates (when $> 1$ data sources produced estimates). Grayed out entries did not pass study diagnostics.

**Supplementary Figure 39: HR estimates using PS stratification.** Each column shows one outcome, and drug-vs-drug comparisons are shown in rows. Grayed out entries did not pass study diagnostics.

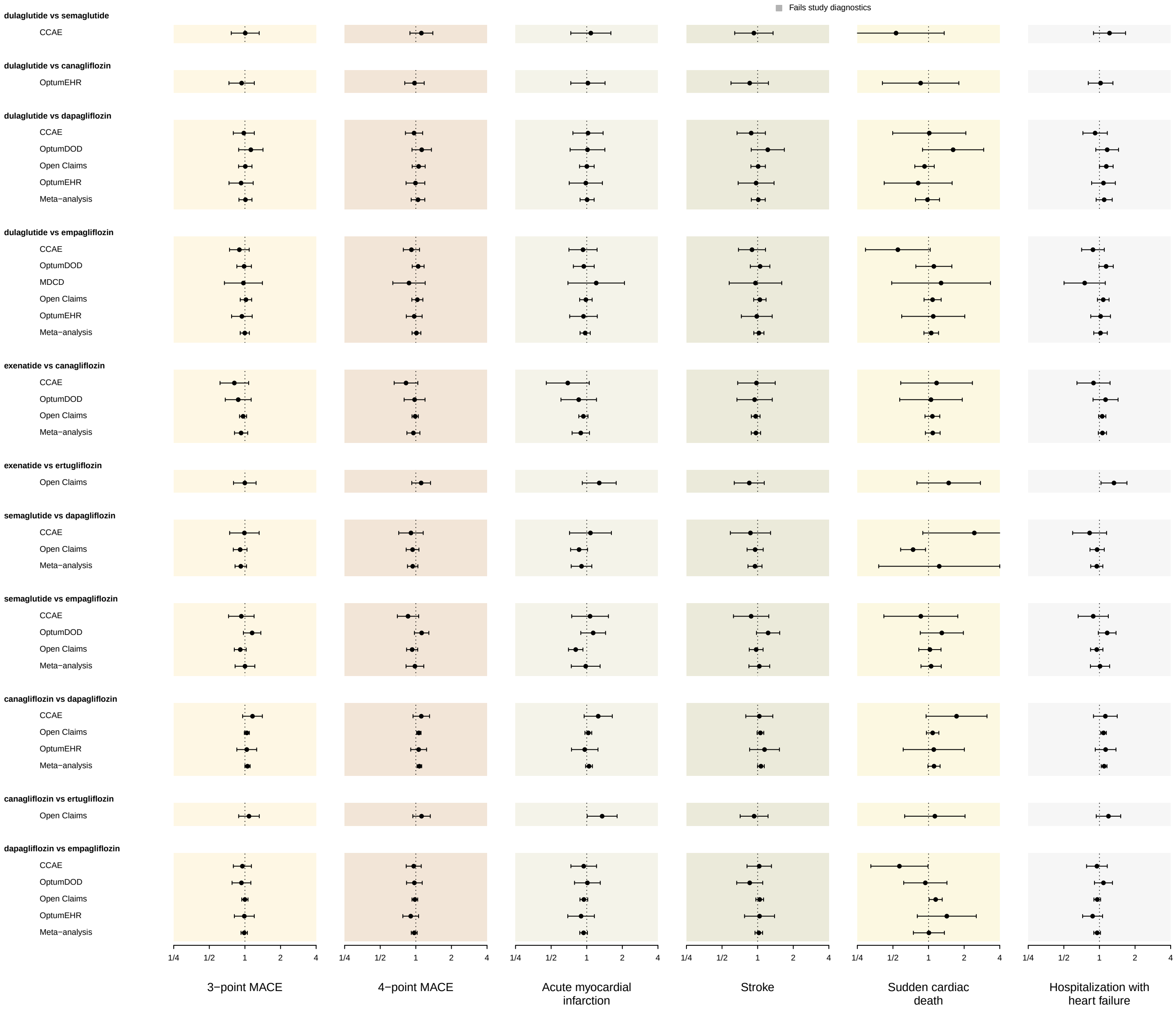

##### 8.3.2 HR estimates using PS matching

In the plot below, we show relative risk estimates of cardiovascular effectiveness outcomes between new-user cohorts of GLP-1RA and SGLT2I individual drugs. Each column shows one outcome, and drug-vs-drug comparisons are shown in rows. We plot calibrated hazard ratios (circles) and their 95 % confidence intervals (whiskers) using the intent-to-treat (ITT) time-at-risk (TAR) definition with PS matching. Estimates are presented across data sources as well as meta-analytic estimates (when $> 1$ data sources produced estimates). Grayed out entries did not pass study diagnostics.

**Supplementary Figure 40: HR estimates using PS matching, part 1.** Each column shows one outcome, and drug-vs-drug comparisons are shown in rows. Grayed out entries did not pass study diagnostics.

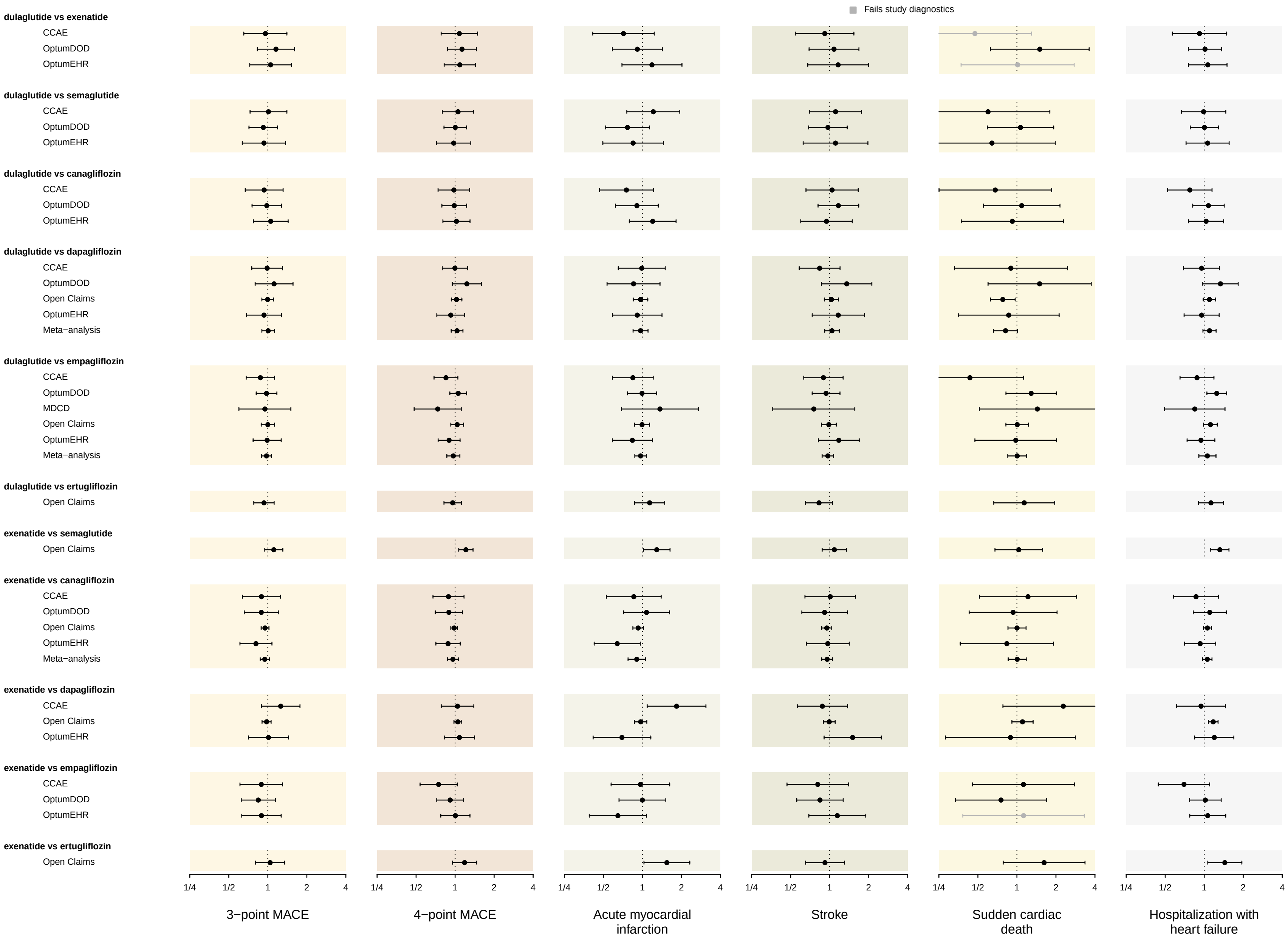

**Supplementary Figure 41: HR estimates using PS matching, part 2.** Each column shows one outcome, and drug-vs-drug comparisons are shown in rows. Grayed out entries did not pass study diagnostics.

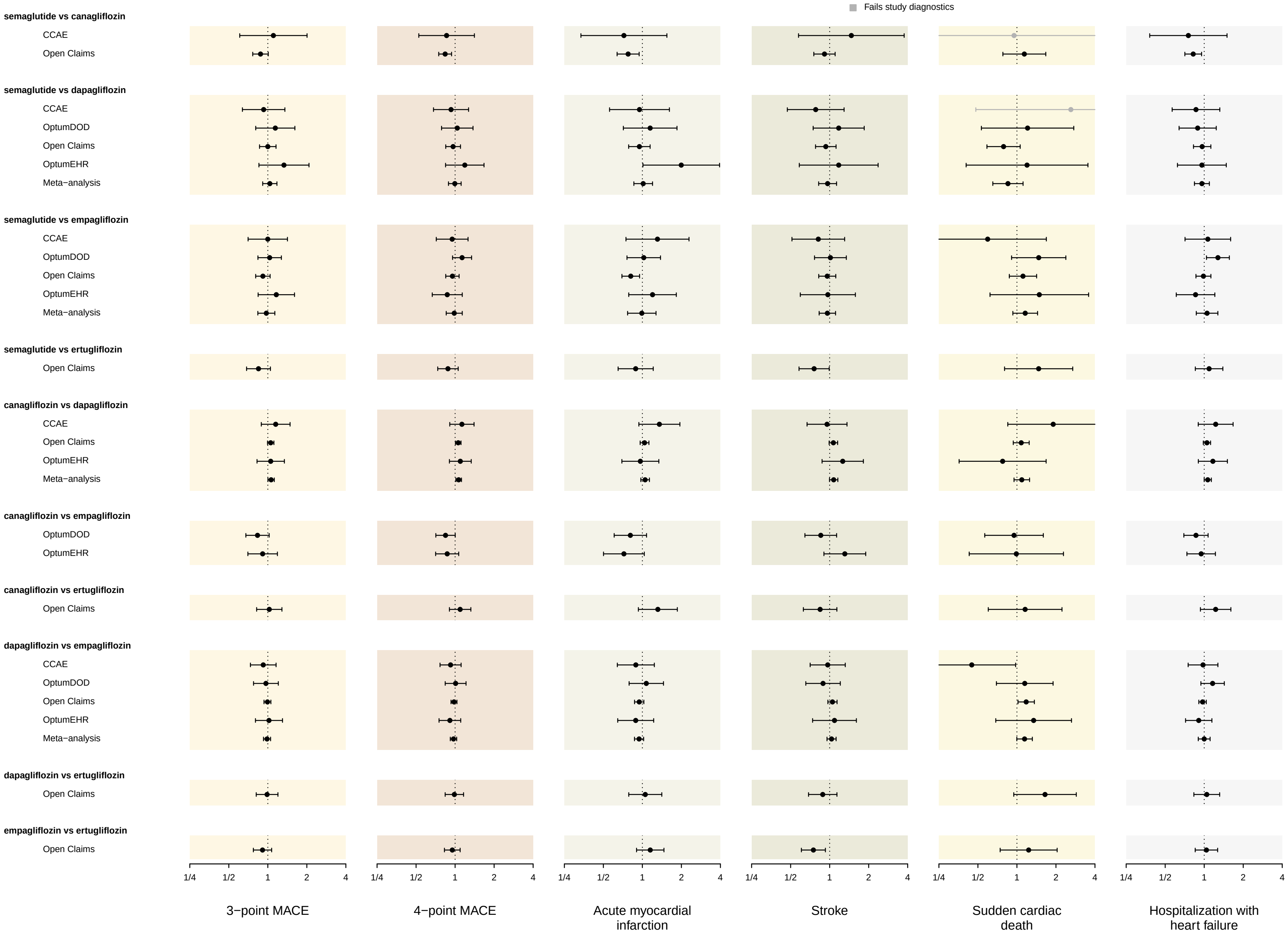

### 9 HR estimates across data sources and drug-level comparions: CVD subgroup

#### 9.1 Meta-analytic HR estimates for all drug-level comparisons

**Supplementary Table 93:** Meta-analytic hazard ratio (HR) estimates and their 95% confidence intervals (CIs) for primary cardiovascular effectiveness outcomes between pairwise new-user cohorts of individual GLP-1RAs and SGLT2Is, within the CVD subgroup. We present calibrated estimates using the **on-treatment** time-at-risk definition and two PS adjustment approaches: stratification and matching.

| | 3-point MACE | | 4-point MACE | |
|---|---|---|---|---|
| | PS stratification | PS matching | PS stratification | PS matching |
| dulaglutide vs semaglutide | 0.88 (0.44 - 1.76) | 0.69 (0.42 - 1.15) | 1.30 (0.75 - 2.26) | 0.96 (0.66 - 1.39) |
| dulaglutide vs dapagliflozin | 1.03 (0.59 - 1.79) | 1.03 (0.61 - 1.75) | 0.99 (0.63 - 1.55) | 0.99 (0.66 - 1.47) |
| dulaglutide vs empagliflozin | 0.86 (0.65 - 1.15) | 0.76 (0.54 - 1.06) | 1.04 (0.83 - 1.29) | 0.76 (0.49 - 1.20) |
| semaglutide vs dapagliflozin | 0.78 (0.60 - 1.02) | 0.94 (0.65 - 1.35) | 0.72 (0.57 - 0.91) | 0.77 (0.61 - 0.96) |
| semaglutide vs empagliflozin | 0.88 (0.73 - 1.06) | 0.83 (0.68 - 1.03) | 0.85 (0.73 - 0.99) | 0.88 (0.68 - 1.12) |
| canagliflozin vs dapagliflozin | 1.04 (0.86 - 1.26) | 0.82 (0.62 - 1.08) | 1.04 (0.89 - 1.22) | 0.90 (0.71 - 1.14) |
| dapagliflozin vs empagliflozin | 1.05 (0.92 - 1.20) | 1.11 (0.95 - 1.29) | 1.06 (0.95 - 1.19) | 1.16 (1.02 - 1.31) |

**Supplementary Table 94:** Meta-analytic hazard ratio (HR) estimates and their 95% confidence intervals (CIs) for cardiovascular effectiveness outcomes between pairwise new-user cohorts of individual GLP-1RAs and SGLT2Is, within the CVD subgroup. We present calibrated HR estimates using the **on-treatment** time-at-risk definition with **PS stratification**. MI = myocardial infarction; HHF = hospitalization or ER visit with heart failure.

| | Acute MI | HHF | Stroke | Sudden cardiac death |
|---|---|---|---|---|
| dulaglutide vs semaglutide | 0.68 (0.25 - 1.83) | 1.31 (0.64 - 2.71) | 1.06 (0.42 - 2.70) | |
| dulaglutide vs dapagliflozin | 1.12 (0.53 - 2.36) | 0.71 (0.39 - 1.26) | 0.96 (0.46 - 2.00) | |
| dulaglutide vs empagliflozin | 0.84 (0.56 - 1.24) | 1.09 (0.79 - 1.49) | 1.04 (0.71 - 1.54) | 0.93 (0.33 - 2.63) |
| semaglutide vs dapagliflozin | 0.76 (0.54 - 1.05) | 0.70 (0.54 - 0.90) | 0.80 (0.57 - 1.12) | 0.60 (0.33 - 1.09) |
| semaglutide vs empagliflozin | 0.83 (0.65 - 1.06) | 0.84 (0.70 - 1.01) | 0.94 (0.73 - 1.20) | 0.96 (0.57 - 1.63) |
| canagliflozin vs dapagliflozin | 1.04 (0.81 - 1.33) | 1.03 (0.84 - 1.26) | 0.97 (0.75 - 1.27) | 1.06 (0.61 - 1.85) |
| dapagliflozin vs empagliflozin | 1.01 (0.85 - 1.19) | 1.16 (0.90 - 1.48) | 1.05 (0.88 - 1.25) | 1.35 (0.97 - 1.88) |

**Supplementary Table 95:** Meta-analytic hazard ratio (HR) estimates and their 95% confidence intervals (CIs) for cardiovascular effectiveness outcomes between pairwise new-user cohorts of individual GLP-1RAs and SGLT2Is, within the CVD subgroup. We present calibrated HR estimates using the **on-treatment** time-at-risk definition with **PS matching**. MI = myocardial infarction; HHF = hospitalization or ER visit with heart failure.

| | Acute MI | HHF | Stroke | Sudden cardiac death |
|---|---|---|---|---|
| dulaglutide vs semaglutide | 0.70 (0.29 - 1.70) | 1.07 (0.62 - 1.84) | 0.51 (0.22 - 1.19) | |
| dulaglutide vs dapagliflozin | 1.08 (0.54 - 2.15) | 1.13 (0.68 - 1.88) | 1.10 (0.50 - 2.40) | |
| dulaglutide vs empagliflozin | 0.85 (0.56 - 1.29) | 0.78 (0.40 - 1.50) | 0.77 (0.47 - 1.27) | 1.39 (0.40 - 4.81) |
| semaglutide vs canagliflozin | 0.82 (0.43 - 1.54) | 0.89 (0.55 - 1.45) | 1.11 (0.50 - 2.43) | |
| semaglutide vs dapagliflozin | 0.77 (0.52 - 1.14) | 0.73 (0.56 - 0.96) | 1.04 (0.58 - 1.89) | 0.39 (0.15 - 1.03) |
| semaglutide vs empagliflozin | 0.84 (0.63 - 1.13) | 0.95 (0.61 - 1.49) | 0.89 (0.68 - 1.18) | 1.01 (0.42 - 2.43) |
| semaglutide vs ertugliflozin | 0.98 (0.35 - 2.73) | 1.36 (0.56 - 3.32) | 0.68 (0.30 - 1.52) | |
| canagliflozin vs dapagliflozin | 1.08 (0.77 - 1.53) | 0.85 (0.63 - 1.15) | 0.69 (0.46 - 1.03) | 1.14 (0.48 - 2.70) |
| canagliflozin vs empagliflozin | 0.22 (0.07 - 0.67) | 0.57 (0.31 - 1.03) | 0.43 (0.16 - 1.20) | |
| canagliflozin vs ertugliflozin | 1.39 (0.41 - 4.74) | 1.48 (0.50 - 4.37) | 0.87 (0.31 - 2.41) | |
| dapagliflozin vs empagliflozin | 1.05 (0.86 - 1.29) | 1.27 (1.01 - 1.60) | 1.25 (1.01 - 1.55) | 1.25 (0.84 - 1.86) |
| dapagliflozin vs ertugliflozin | 1.31 (0.49 - 3.48) | 1.53 (0.76 - 3.07) | 0.76 (0.36 - 1.58) | |
| empagliflozin vs ertugliflozin | 1.08 (0.48 - 2.43) | 1.09 (0.60 - 2.01) | 0.55 (0.30 - 0.99) | |

**Supplementary Table 96:** Meta-analytic hazard ratio (HR) estimates and their 95% confidence intervals (CIs) for primary cardiovascular effectiveness outcomes between pairwise new-user cohorts of individual GLP-1RAs and SGLT2Is, within the CVD subgroup. We present calibrated estimates using the **intent-to-treat** time-at-risk definition and two PS adjustment approaches: stratification and matching.

| | 3-point MACE | | 4-point MACE | |
|---|---|---|---|---|
| | PS stratification | PS matching | PS stratification | PS matching |
| dulaglutide vs semaglutide | 0.88 (0.53 - 1.47) | 0.79 (0.61 - 1.03) | 1.28 (0.87 - 1.90) | 0.92 (0.74 - 1.14) |
| dulaglutide vs dapagliflozin | 0.89 (0.61 - 1.29) | 1.20 (0.90 - 1.60) | 1.00 (0.74 - 1.34) | 1.15 (0.92 - 1.45) |
| dulaglutide vs empagliflozin | 0.91 (0.77 - 1.07) | 0.92 (0.76 - 1.11) | 0.96 (0.84 - 1.10) | 0.93 (0.80 - 1.09) |
| semaglutide vs dapagliflozin | 0.87 (0.76 - 1.00) | 1.00 (0.79 - 1.26) | 0.86 (0.76 - 0.96) | 0.89 (0.77 - 1.03) |
| semaglutide vs empagliflozin | 0.94 (0.84 - 1.05) | 1.03 (0.90 - 1.19) | 0.92 (0.84 - 1.01) | 0.96 (0.87 - 1.07) |
| canagliflozin vs dapagliflozin | 1.05 (0.97 - 1.14) | 1.07 (0.98 - 1.17) | 1.09 (1.02 - 1.17) | 1.06 (0.88 - 1.27) |
| dapagliflozin vs empagliflozin | 1.02 (0.95 - 1.09) | 1.02 (0.94 - 1.11) | 0.99 (0.94 - 1.06) | 1.03 (0.96 - 1.11) |

**Supplementary Table 97:** Meta-analytic hazard ratio (HR) estimates and their 95% confidence intervals (CIs) for cardiovascular effectiveness outcomes between pairwise new-user cohorts of individual GLP-1RAs and SGLT2Is, within the CVD subgroup. We present calibrated HR estimates using the **intent-to-treat** time-at-risk definition with **PS stratification**. MI = myocardial infarction; HHF = hospitalization or ER visit with heart failure.

| | Acute MI | HHF | Stroke | Sudden cardiac death |
|---|---|---|---|---|
| dulaglutide vs semaglutide | 0.87 (0.43 - 1.76) | 1.42 (0.89 - 2.26) | 0.93 (0.49 - 1.78) | |
| dulaglutide vs dapagliflozin | 1.05 (0.63 - 1.75) | 1.01 (0.70 - 1.45) | 0.71 (0.44 - 1.14) | |
| dulaglutide vs empagliflozin | 0.91 (0.73 - 1.14) | 1.04 (0.87 - 1.26) | 0.99 (0.80 - 1.24) | 0.85 (0.42 - 1.71) |
| semaglutide vs dapagliflozin | 0.87 (0.72 - 1.05) | 0.84 (0.73 - 0.96) | 0.94 (0.79 - 1.12) | 0.61 (0.45 - 0.83) |
| semaglutide vs empagliflozin | 0.85 (0.73 - 0.98) | 0.90 (0.81 - 1.01) | 1.06 (0.92 - 1.23) | 0.98 (0.67 - 1.43) |
| canagliflozin vs dapagliflozin | 1.05 (0.95 - 1.16) | 1.09 (1.00 - 1.18) | 1.04 (0.95 - 1.15) | 1.10 (0.93 - 1.31) |
| dapagliflozin vs empagliflozin | 0.93 (0.85 - 1.02) | 0.97 (0.91 - 1.04) | 1.08 (0.99 - 1.19) | 0.93 (0.26 - 3.34) |

**Supplementary Table 98:** Meta-analytic hazard ratio (HR) estimates and their 95% confidence intervals (CIs) for cardiovascular effectiveness outcomes between pairwise new-user cohorts of individual GLP-1RAs and SGLT2Is, within the CVD subgroup. We present calibrated HR estimates using the **intent-to-treat** time-at-risk definition with **PS matching**. MI = myocardial infarction; HHF = hospitalization or ER visit with heart failure.

| | Acute MI | HHF | Stroke | Sudden cardiac death |
|---|---|---|---|---|
| dulaglutide vs semaglutide | 0.71 (0.50 - 1.00) | 1.06 (0.83 - 1.36) | 0.80 (0.56 - 1.14) | 0.57 (0.23 - 1.46) |
| dulaglutide vs dapagliflozin | 1.23 (0.82 - 1.82) | 1.08 (0.82 - 1.43) | 1.27 (0.85 - 1.91) | 1.98 (0.50 - 7.79) |
| dulaglutide vs empagliflozin | 0.86 (0.67 - 1.10) | 0.94 (0.78 - 1.12) | 1.15 (0.88 - 1.49) | 0.67 (0.28 - 1.58) |
| semaglutide vs canagliflozin | 0.71 (0.54 - 0.94) | 0.80 (0.65 - 0.98) | 1.11 (0.84 - 1.46) | 0.88 (0.54 - 1.43) |
| semaglutide vs dapagliflozin | 1.14 (0.83 - 1.57) | 0.90 (0.76 - 1.05) | 0.97 (0.78 - 1.21) | 1.16 (0.30 - 4.54) |
| semaglutide vs empagliflozin | 0.96 (0.78 - 1.18) | 1.02 (0.83 - 1.26) | 1.08 (0.93 - 1.26) | 1.01 (0.76 - 1.33) |
| semaglutide vs ertugliflozin | 0.84 (0.54 - 1.31) | 1.10 (0.77 - 1.56) | 0.88 (0.59 - 1.30) | 1.30 (0.56 - 3.03) |
| canagliflozin vs dapagliflozin | 1.12 (0.99 - 1.26) | 0.91 (0.53 - 1.57) | 1.03 (0.92 - 1.15) | 1.11 (0.90 - 1.37) |
| canagliflozin vs empagliflozin | 0.83 (0.62 - 1.11) | 0.80 (0.64 - 0.99) | 1.01 (0.74 - 1.37) | 1.11 (0.58 - 2.14) |
| canagliflozin vs ertugliflozin | 1.40 (0.84 - 2.33) | 0.90 (0.62 - 1.31) | 0.86 (0.54 - 1.37) | 1.00 (0.42 - 2.38) |
| dapagliflozin vs empagliflozin | 0.92 (0.83 - 1.02) | 1.12 (0.92 - 1.36) | 1.02 (0.84 - 1.25) | 1.27 (1.06 - 1.52) |
| dapagliflozin vs ertugliflozin | 0.77 (0.51 - 1.18) | 1.01 (0.73 - 1.40) | 0.84 (0.57 - 1.23) | 1.62 (0.79 - 3.32) |
| empagliflozin vs ertugliflozin | 0.89 (0.63 - 1.26) | 0.95 (0.72 - 1.27) | 0.68 (0.49 - 0.93) | 1.12 (0.58 - 2.17) |

#### 9.2 Under the on-treatment (OT) time-at-risk definition

##### 9.2.1 HR estimates using PS stratification

In the plot below, we show relative risk estimates of cardiovascular effectiveness outcomes between new-user cohorts of GLP-1RA and SGLT2I individual drugs, among patients with established CVD. Each column shows one outcome, and drug-vs-drug comparisons are shown in rows. We plot calibrated hazard ratios (circles) and their 95 % confidence intervals (whiskers) using the on-treatment (OT) time-at-risk (TAR) definition with PS stratification. Estimates are presented across data sources as well as meta-analytic estimates (when $> 1$ data sources produced estimates). Grayed out entries did not pass study diagnostics.

**Supplementary Figure 42: HR estimates using PS stratification, CVD subgroup.** Each column shows one outcome, and drug-vs-drug comparisons are shown in rows. Grayed out entries did not pass study diagnostics.

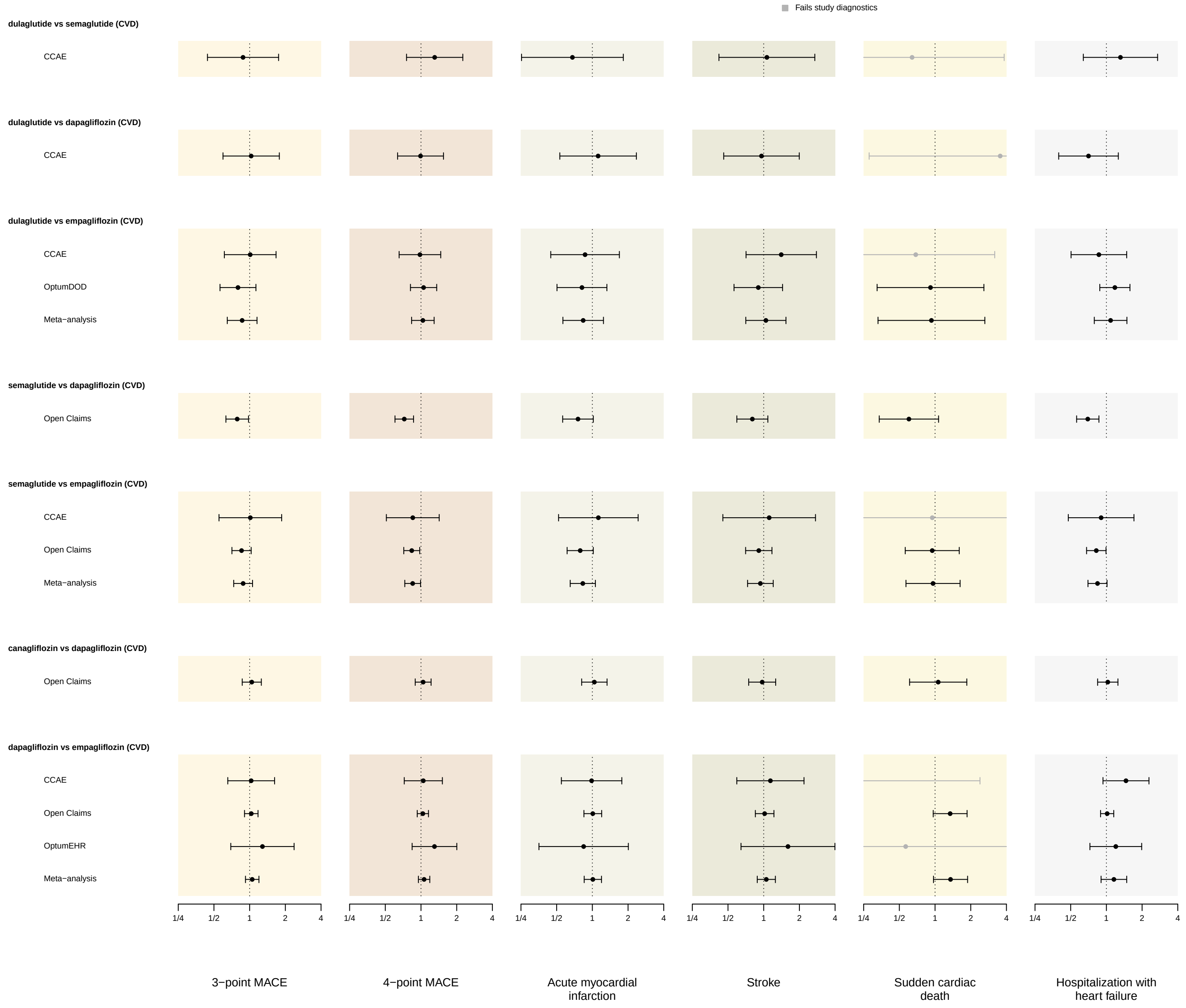

##### 9.2.2 HR estimates using PS matching

In the plots below, we show relative risk estimates of cardiovascular effectiveness outcomes between new-user cohorts of GLP-1RA and SGLT2I individual drugs, among patients with established CVD. Each column shows one outcome, and drug-vs-drug comparisons are shown in rows. We plot calibrated hazard ratios (circles) and their 95 % confidence intervals (whiskers) using the on-treatment (OT) time-at-risk (TAR) definition with PS matching. Estimates are presented across data sources as well as meta-analytic estimates (when $> 1$ data sources produced estimates). Grayed out entries did not pass study diagnostics.

**Supplementary Figure 43: HR estimates using PS matching for CVD subgroup, part 1.** Each column shows one outcome, and drug-vs-drug comparisons are shown in rows. Grayed out entries did not pass study diagnostics.

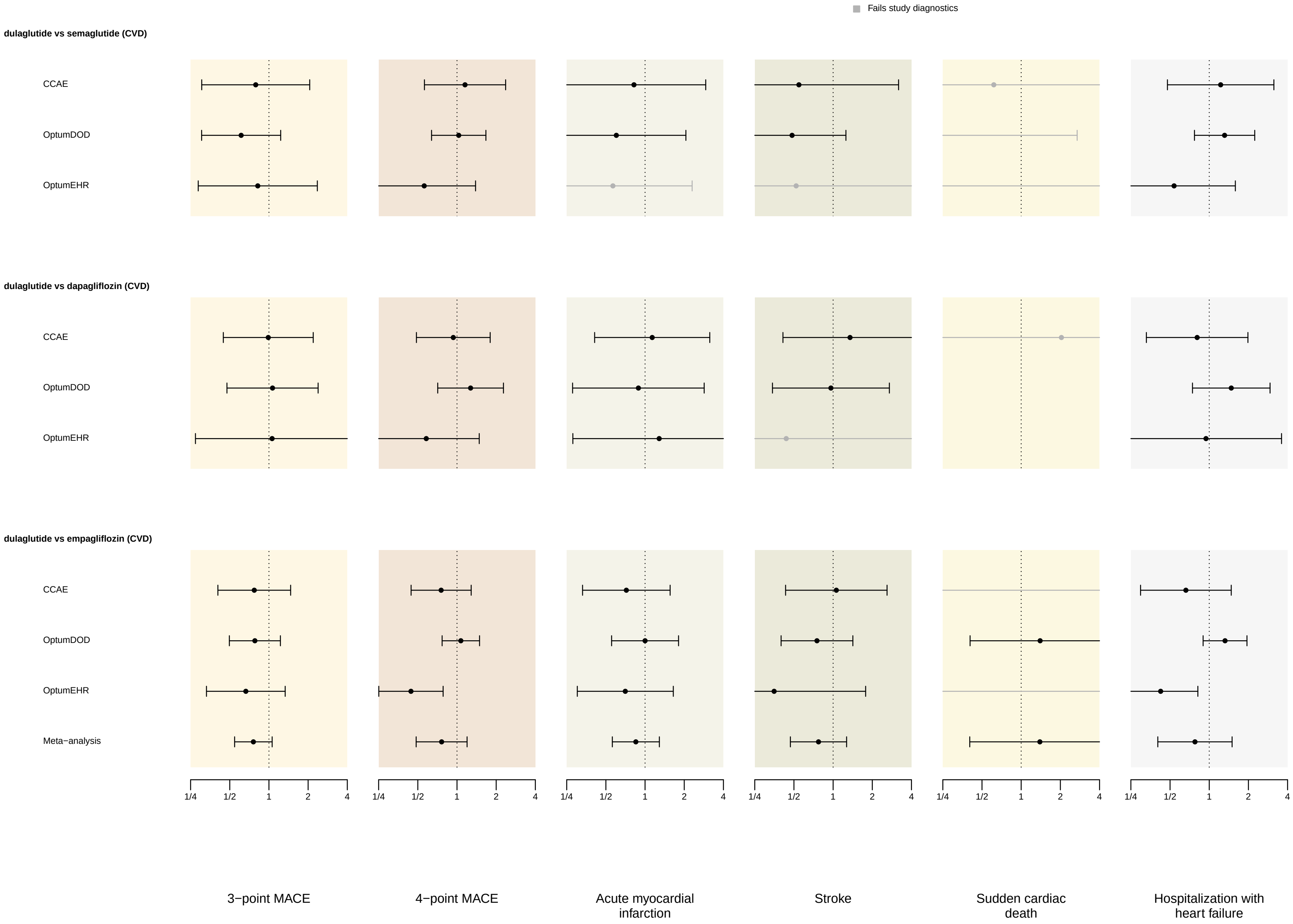

**Supplementary Figure 44: HR estimates using PS matching for CVD subgroup, part 2.** Each column shows one outcome, and drug-vs-drug comparisons are shown in rows. Grayed out entries did not pass study diagnostics.

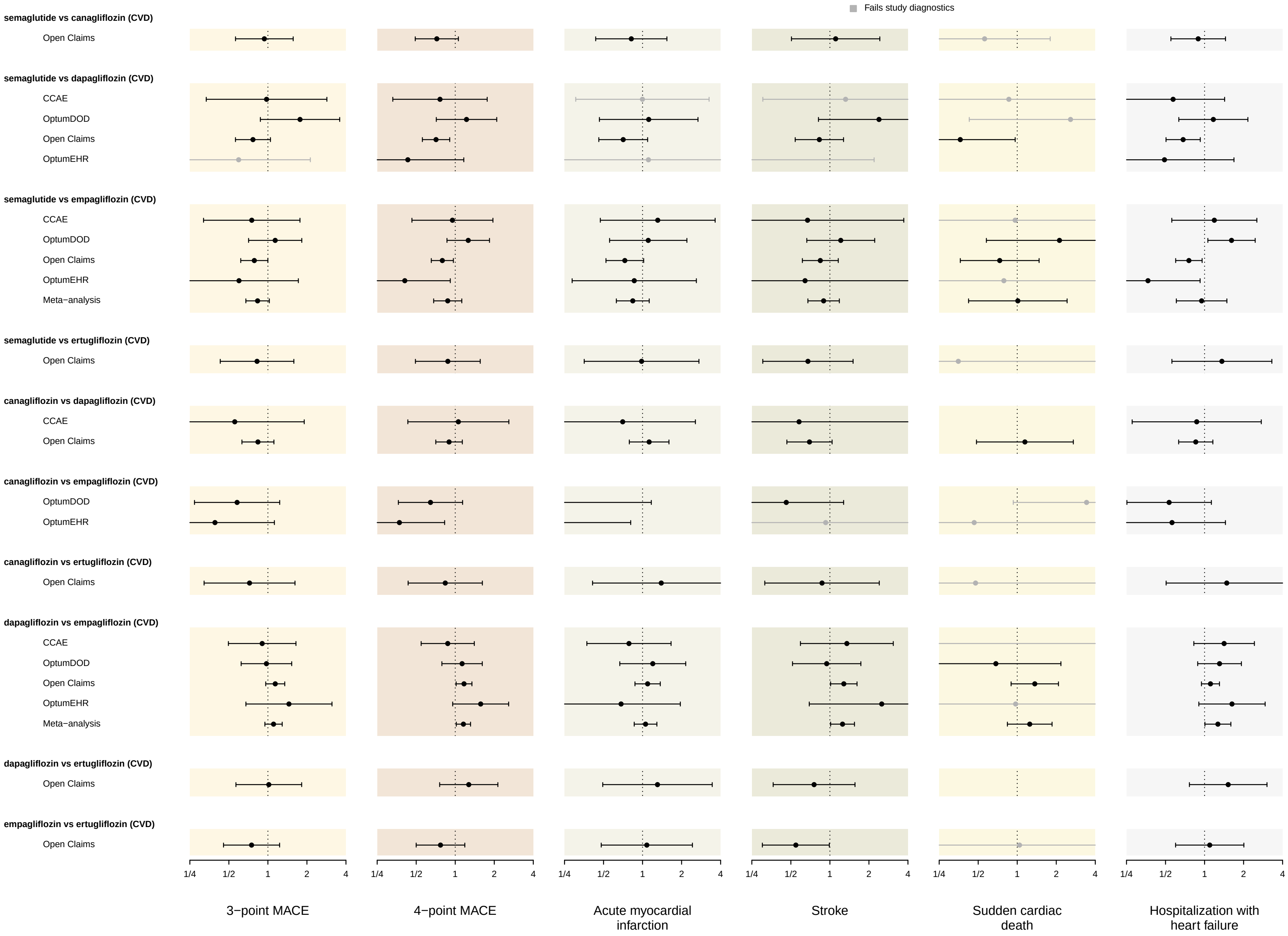

#### 9.3 Under the intent-to-treat (ITT) time-at-risk definitions

##### 9.3.1 HR estimates using PS stratification

In the plot below, we show relative risk estimates of cardiovascular effectiveness outcomes between new-user cohorts of GLP-1RA and SGLT2I individual drugs, among patients with established CVD. Each column shows one outcome, and drug-vs-drug comparisons are shown in rows. We plot calibrated hazard ratios (circles) and their 95 % confidence intervals (whiskers) using the intent-to-treat (ITT) time-at-risk (TAR) definition with PS stratification. Estimates are presented across data sources as well as meta-analytic estimates (when $> 1$ data sources produced estimates). Grayed out entries did not pass study diagnostics.

**Supplementary Figure 45: HR estimates using PS stratification, CVD subgroup.** Each column shows one outcome, and drug-vs-drug comparisons are shown in rows. Grayed out entries did not pass study diagnostics.

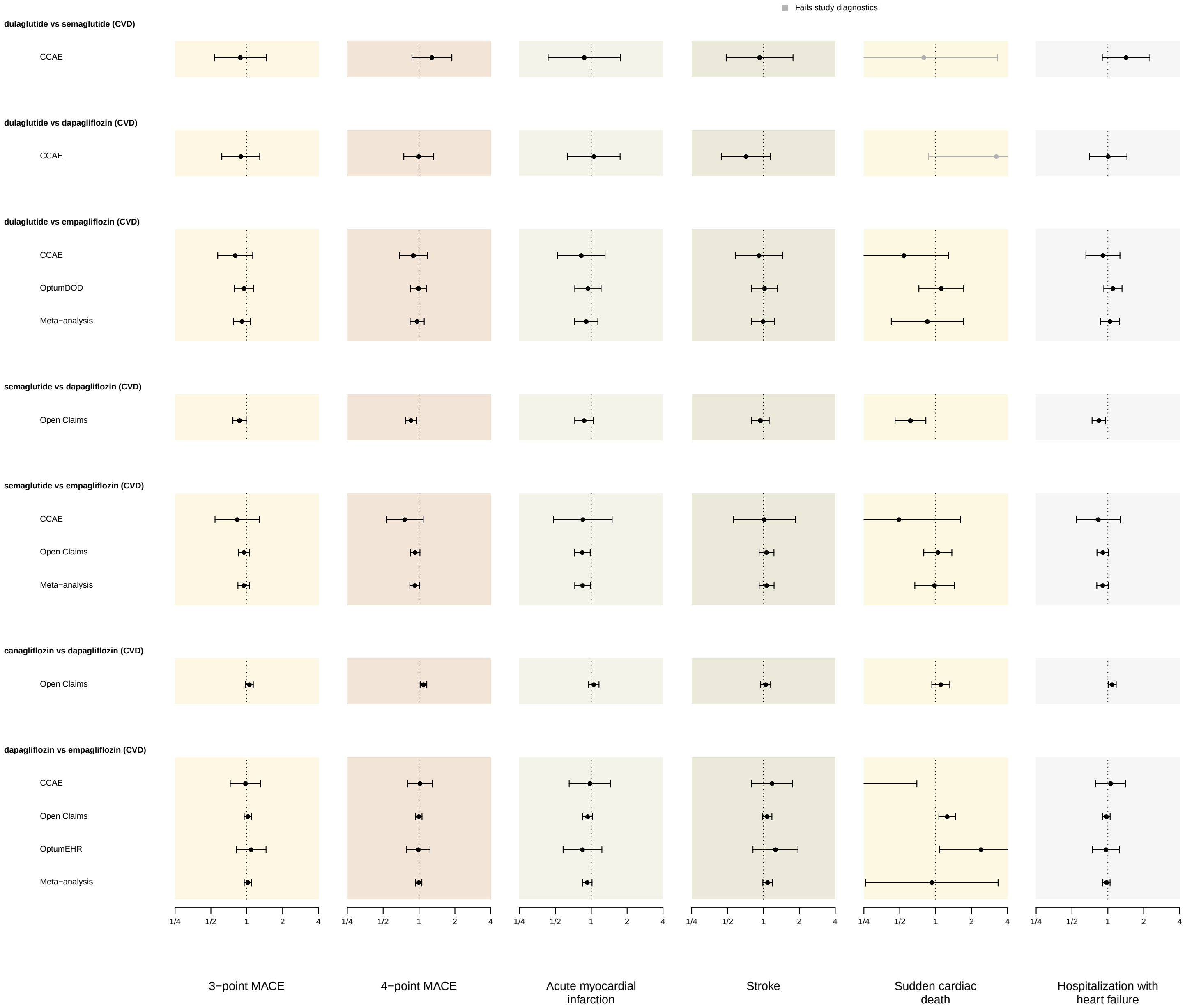

##### 9.3.2 HR estimates using PS matching

In the plots below, we show relative risk estimates of cardiovascular effectiveness outcomes between new-user cohorts of GLP-1RA and SGLT2I individual drugs, among patients with established CVD. Each column shows one outcome, and drug-vs-drug comparisons are shown in rows. We plot calibrated hazard ratios (circles) and their 95 % confidence intervals (whiskers) using the intent-to-treat (ITT) time-at-risk (TAR) definition with PS matching. Estimates are presented across data sources as well as meta-analytic estimates (when $> 1$ data sources produced estimates). Grayed out entries did not pass study diagnostics.

**Supplementary Figure 46: HR estimates using PS matching for CVD subgroup, part 1.** Each column shows one outcome, and drug-vs-drug comparisons are shown in rows. Grayed out entries did not pass study diagnostics.

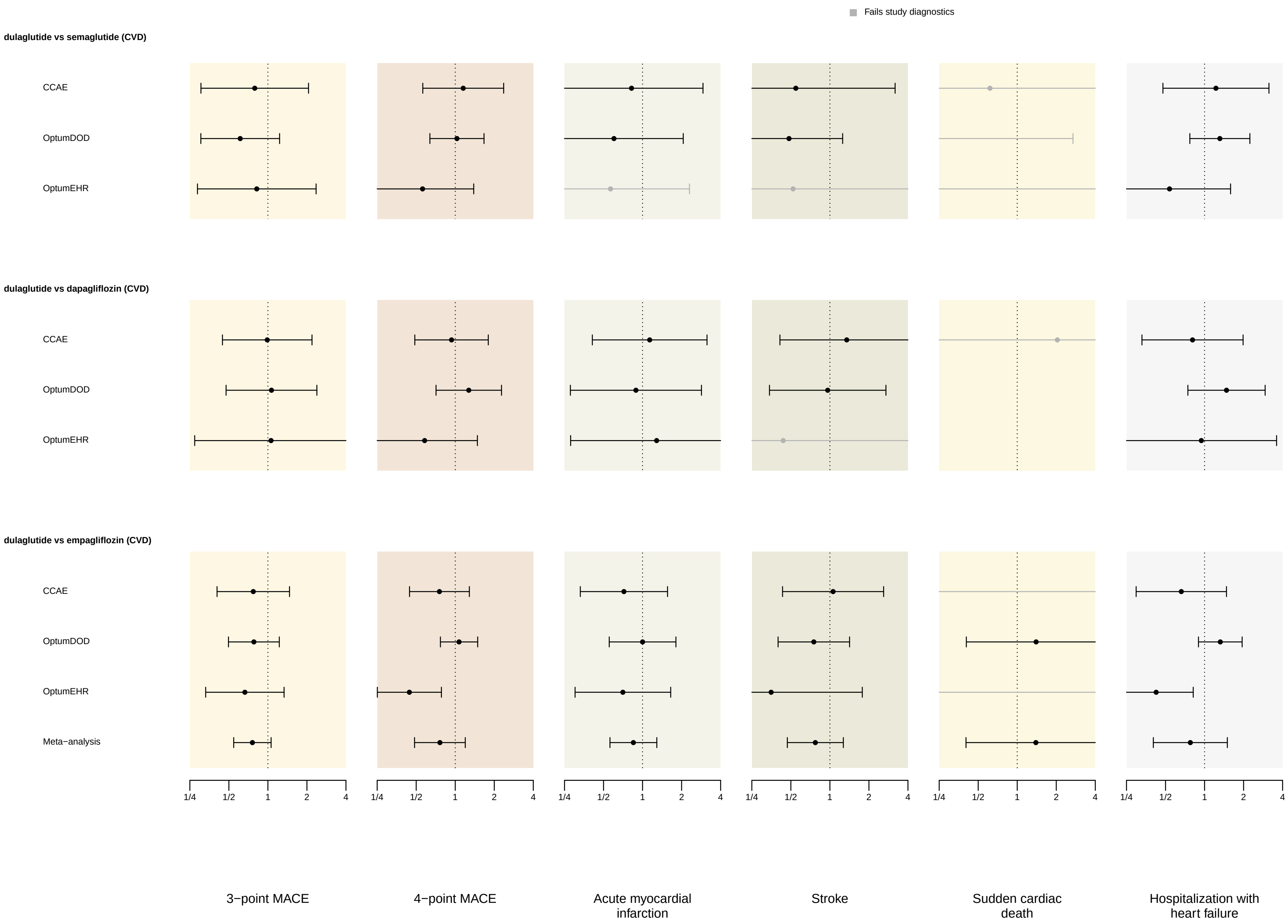

**Supplementary Figure 47: HR estimates using PS matching for CVD subgroup, part 2.** Each column shows one outcome, and drug-vs-drug comparisons are shown in rows. Grayed out entries did not pass study diagnostics.

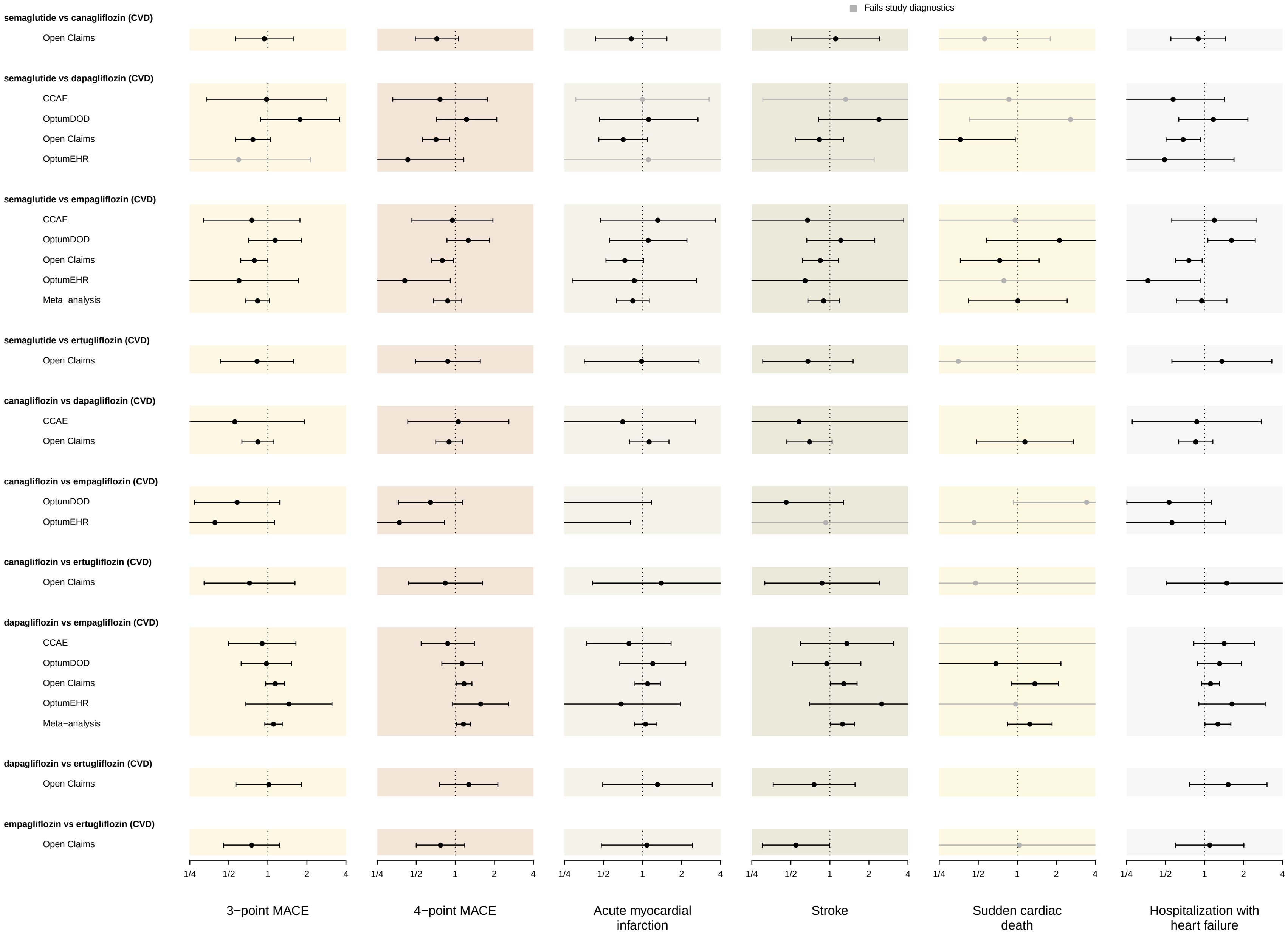

### 10 Study diagnostics across comparisons and data sources

#### 10.1 dulaglutide vs semaglutide

**Supplementary Figure 48:** Study diagnostics for comparing new-users of dulaglutide (target) and semaglutide (comparator) on 3-pt MACE, under the on-treatment TAR definition with PS stratification. We show diagnostics on empirical equipoise, covariate balance, minimally detectable relative risk (MDRR), and empirical average systematic error (EASE) evaluated via negative control analysis, for each data source that produced an estimate.

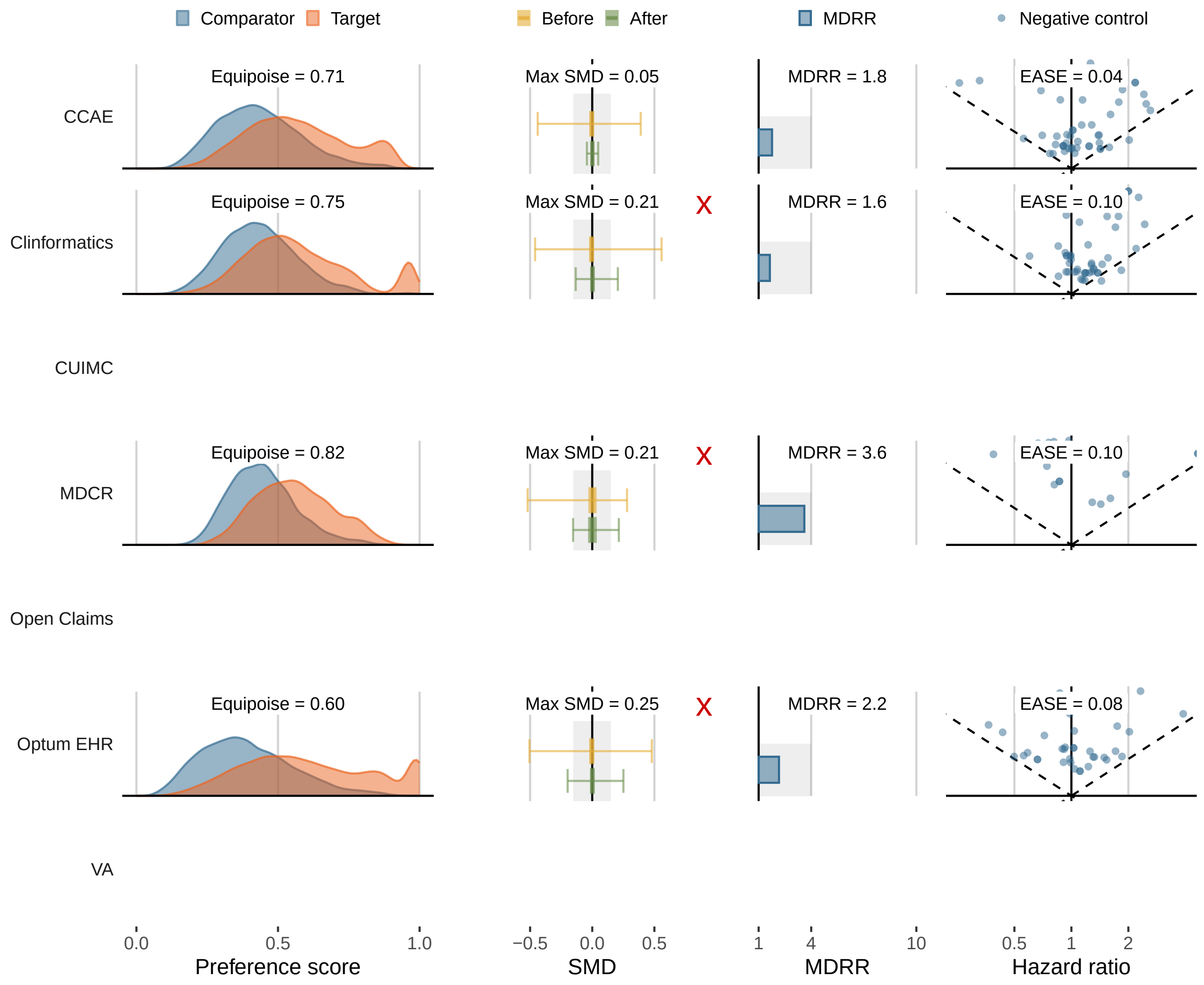

#### 10.2 dulaglutide vs dapagliflozin

**Supplementary Figure 49:** Study diagnostics for comparing new-users of dulaglutide (target) and dapagliflozin (comparator) on 3-pt MACE, under the on-treatment TAR definition with PS stratification. We show diagnostics on empirical equipoise, covariate balance, minimally detectable relative risk (MDRR), and empirical average systematic error (EASE) evaluated via negative control analysis, for each data source that produced an estimate.

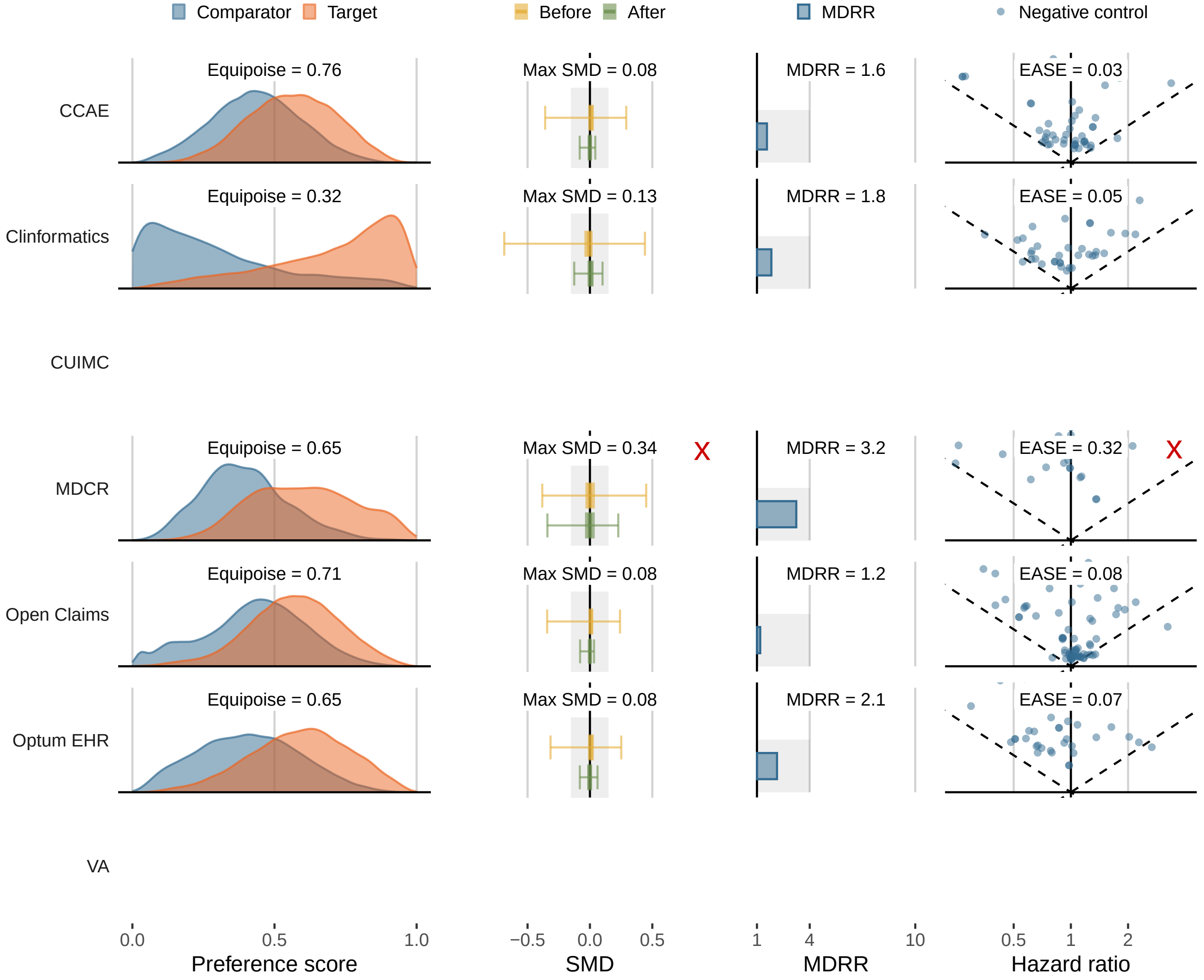

#### 10.3 dulaglutide vs empagliflozin

**Supplementary Figure 50:** Study diagnostics for comparing new-users of dulaglutide (target) and empagliflozin (comparator) on 3-pt MACE, under the on-treatment TAR definition with PS stratification. We show diagnostics on empirical equipoise, covariate balance, minimally detectable relative risk (MDRR), and empirical average systematic error (EASE) evaluated via negative control analysis, for each data source that produced an estimate.

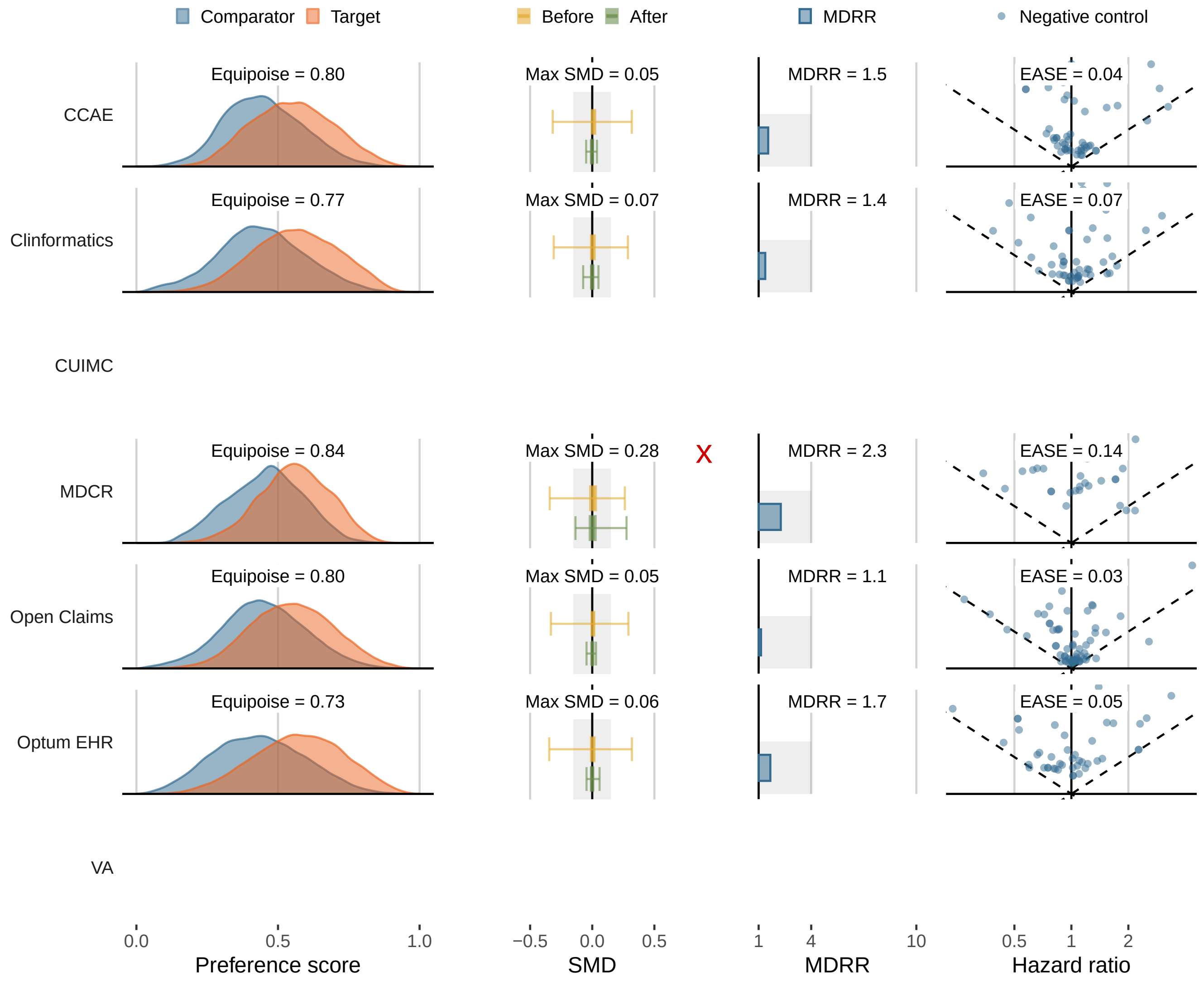

#### 10.4 semaglutide vs dapagliflozin

**Supplementary Figure 51:** Study diagnostics for comparing new-users of semaglutide (target) and dapagliflozin (comparator) on 3-pt MACE, under the on-treatment TAR definition with PS stratification. We show diagnostics on empirical equipoise, covariate balance, minimally detectable relative risk (MDRR), and empirical average systematic error (EASE) evaluated via negative control analysis, for each data source that produced an estimate.

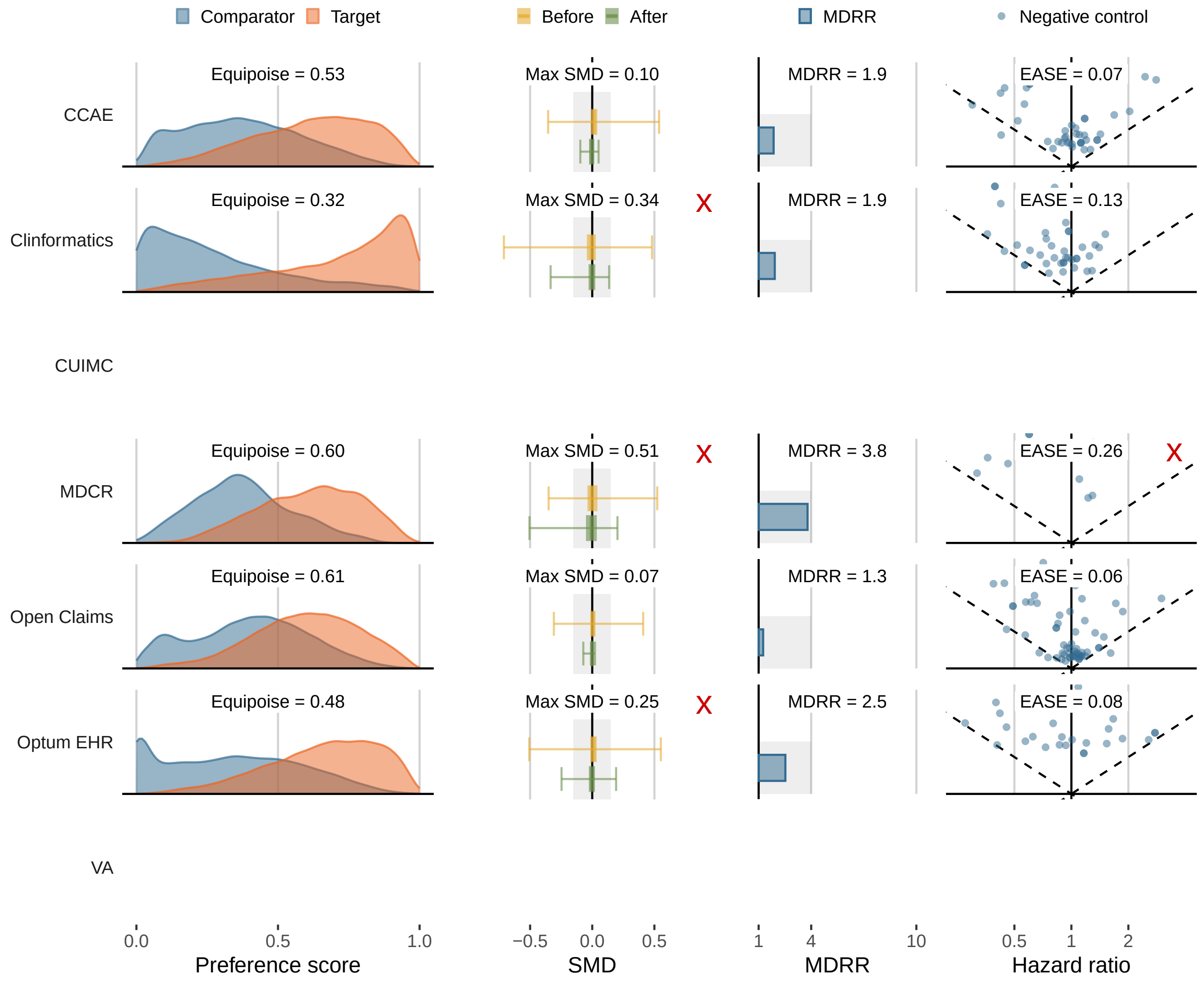

#### 10.5 semaglutide vs empagliflozin

**Supplementary Figure 52:** Study diagnostics for comparing new-users of semaglutide (target) and empagliflozin (comparator) on 3-pt MACE, under the on-treatment TAR definition with PS stratification. We show diagnostics on empirical equipoise, covariate balance, minimally detectable relative risk (MDRR), and empirical average systematic error (EASE) evaluated via negative control analysis, for each data source that produced an estimate.

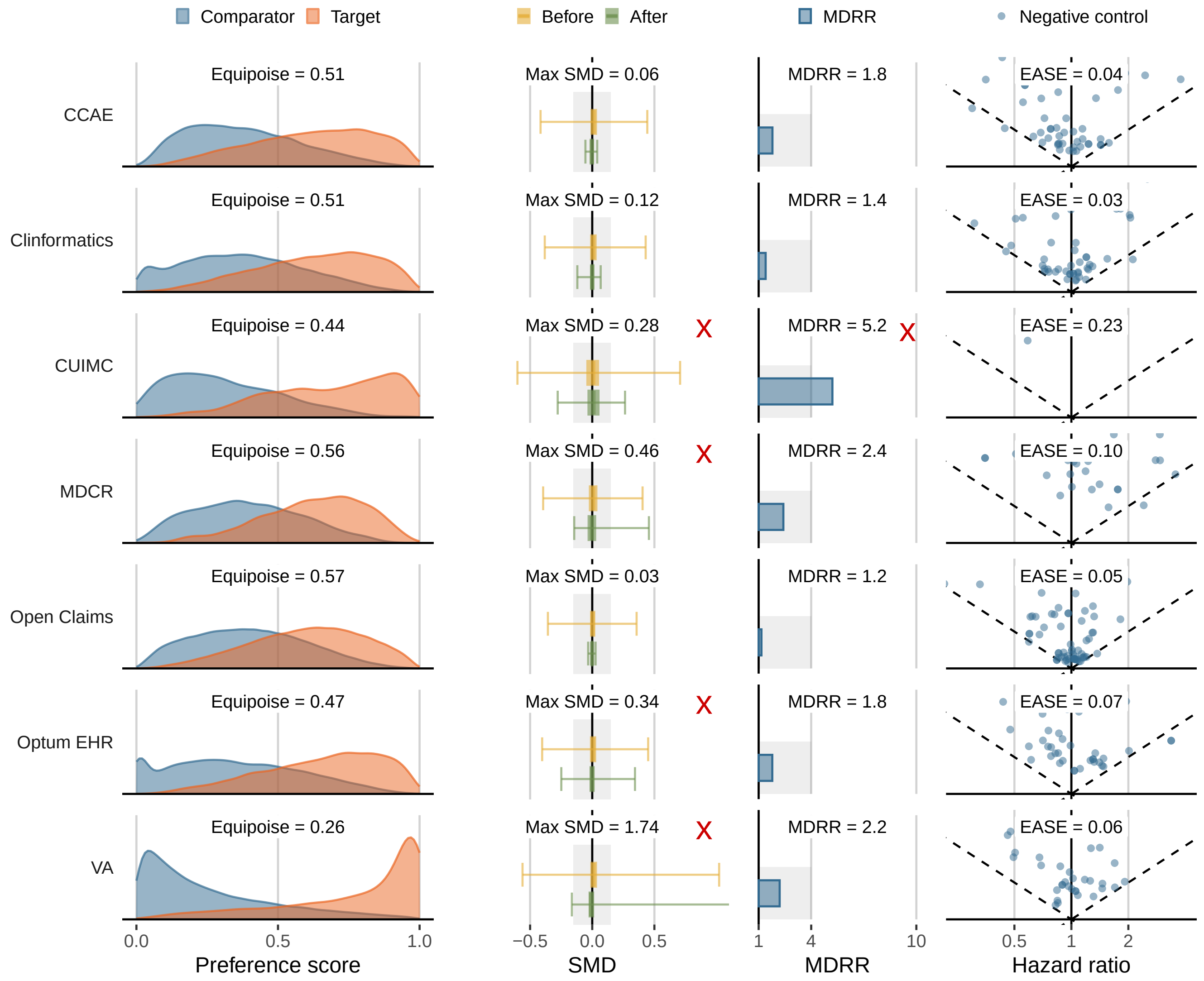

#### 10.6 dapagliflozin vs empagliflozin

**Supplementary Figure 53:** Study diagnostics for comparing new-users of dapagliflozin (target) and empagliflozin (comparator) on 3-pt MACE, under the on-treatment TAR definition with PS stratification. We show diagnostics on empirical equipoise, covariate balance, minimally detectable relative risk (MDRR), and empirical average systematic error (EASE) evaluated via negative control analysis, for each data source that produced an estimate.

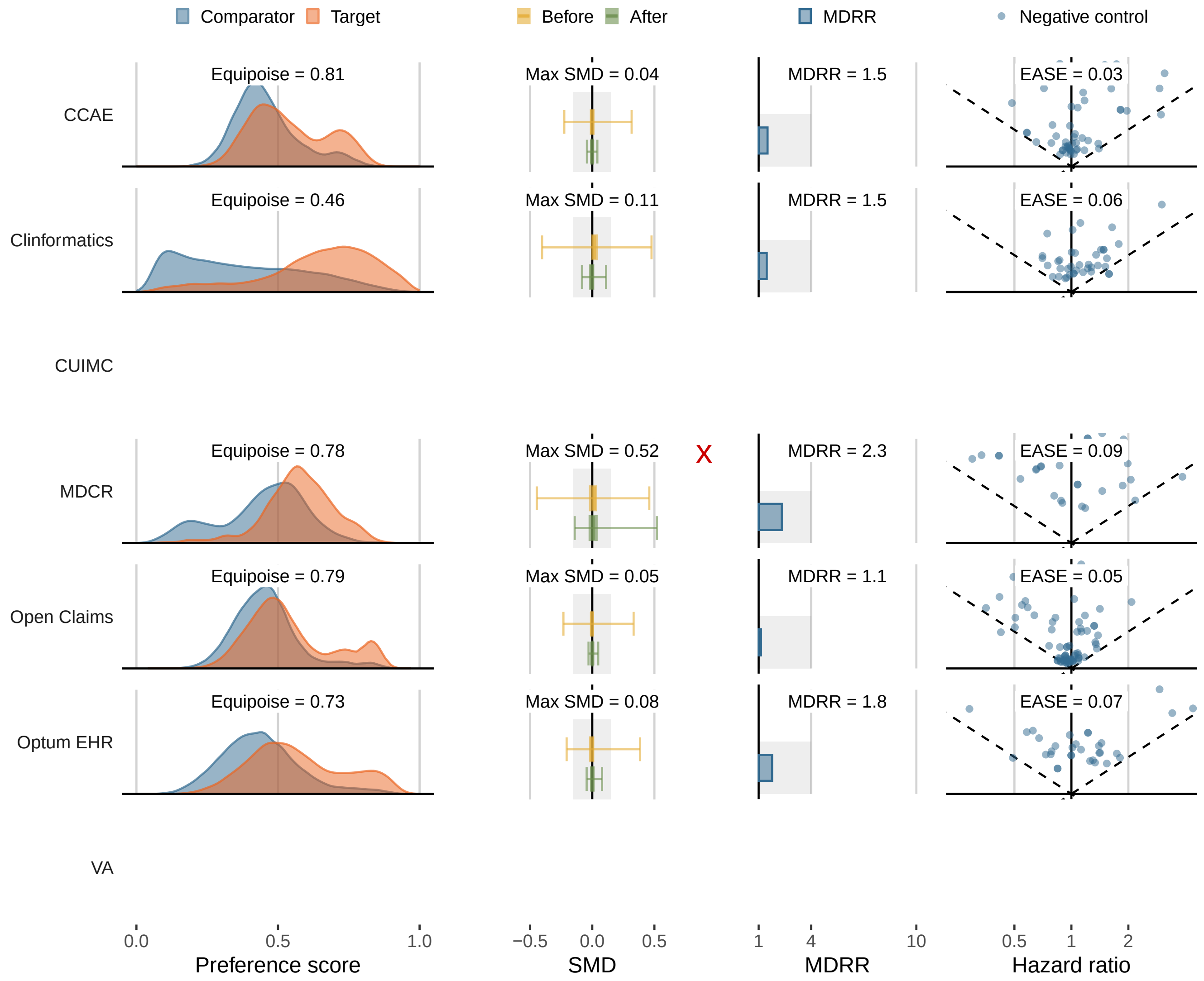

#### 11 Additional Analyses with IPCW

To explore the potential impact of informative censoring on our main analyses, we conducted additional analyses using inverse probability of censoring weights (IPCW) to adjust for potential informative censoring. Below we present hazard ratio estimates and 95% confidence intervals for 3-point MACE and 4-point MACE between new-users of semaglutide and empagliflozin, as well as between new-users of semaglutide and dapagliflozin, across three databases, CCAE, Clinformatics and OptumEHR, using Cox proportional hazards models with IPCW and/or inverse probability treatment weighting (IPTW) adjustments.

We noticed that, using IPCW does not substantially change the HR estimates, both with the unadjusted Cox model and with IPTW (for confounding) adjustments. In comparison, IPTW adjustment has a much larger impact on the HR estimates.

**Supplementary Table 99:** Comparing analytic results from four variant methods with IPCW (for censoring) and/or IPTW (for confounding). We summarize hazard ratio estimates and 95% confidence intervals for 3-point MACE and 4-point MACE between new-users of semaglutide and empagliflozin, across three databases, CCAE, Clinformatics and OptumEHR. Cox = Cox proportional hazards model; IPCW = inverse probability censoring weights; IPTW = inverse probability treatment weights.

| | CCAE | Clinformatics | OptumEHR |
|---|---|---|---|
| **3-point MACE** | | | |
| Unadjusted Cox | 0.87 (0.67, 1.14) | 0.69 (0.56, 0.84) | 0.67 (0.48, 0.93) |
| Cox with IPCW | 0.84 (0.63, 1.11) | 0.65 (0.53, 0.81) | 0.66 (0.47, 0.92) |
| Cox with IPTW | 0.98 (0.68, 1.40) | 1.05 (0.82, 1.34) | 1.06 (0.73, 1.55) |
| Cox with IPCW + IPTW | 0.91 (0.62, 1.34) | 0.99 (0.75, 1.29) | 1.07 (0.72, 1.57) |
| **4-point MACE** | | | |
| Unadjusted Cox | 0.76 (0.62, 0.94) | 0.74 (0.64, 0.84) | 0.60 (0.48, 0.76) |
| Cox with IPCW | 0.75 (0.61, 0.93) | 0.70 (0.61, 0.81) | 0.60 (0.47, 0.76) |
| Cox with IPTW | 0.88 (0.67, 1.15) | 1.06 (0.89, 1.25) | 0.90 (0.68, 1.19) |
| Cox with IPCW + IPTW | 0.87 (0.65, 1.17) | 1.01 (0.84, 1.22) | 0.88 (0.66, 1.18) |

**Supplementary Table 100:** Comparing analytic results from four variant methods with IPCW (for censoring) and/or IPTW (for confounding). We summarize hazard ratio estimates and 95% confidence intervals for 3-point MACE and 4-point MACE between new-users of semaglutide and dapagliflozin, across three databases, CCAE, Clinformatics and OptumEHR. Cox = Cox proportional hazards model; IPCW = inverse probability censoring weights; IPTW = inverse probability treatment weights.

| | CCAE | Clinformatics | OptumEHR |
|---|---|---|---|
| **3-point MACE** | | | |
| Unadjusted Cox | 0.92 (0.70, 1.22) | 0.39 (0.31, 0.50) | 0.77 (0.51, 1.17) |
| Cox with IPCW | 0.91 (0.67, 1.22) | 0.37 (0.29, 0.48) | 0.76 (0.49, 1.17) |
| Cox with IPTW | 0.97 (0.66, 1.44) | 0.68 (0.45, 1.01) | 0.98 (0.50, 1.93) |
| Cox with IPCW + IPTW | 0.97 (0.63, 1.49) | 0.60 (0.40, 0.89) | 0.98 (0.50, 1.91) |
| **4-point MACE** | | | |
| Unadjusted Cox | 0.84 (0.67, 1.04) | 0.39 (0.33, 0.46) | 0.60 (0.45, 0.79) |
| Cox with IPCW | 0.85 (0.67, 1.07) | 0.37 (0.31, 0.45) | 0.60 (0.45, 0.81) |
| Cox with IPTW | 0.89 (0.66, 1.19) | 0.64 (0.50, 0.82) | 0.84 (0.56, 1.27) |
| Cox with IPCW + IPTW | 0.93 (0.67, 1.29) | 0.58 (0.45, 0.75) | 0.86 (0.56, 1.31) |

[15] Kaspar M, Fette G, Guder G, Seidlmayer L, Ertl M, Dietrich G, et al. Underestimated prevalence of heart failure in hospital inpatients: a comparison of ICD codes and discharge letter information. Clinical research in cardiology:

official journal of the German Cardiac Society. 2018 Sep;107(9):778-87. doi: 10.1007/s00392-0181245z. Epub 2018 Apr 17.
